# Estimating the incidence and key risk factors of cardiovascular disease in patients at high risk of imminent fracture using routinely collected real-world data from the UK

**DOI:** 10.1101/2022.02.04.22270275

**Authors:** Marta Pineda-Moncusí, Leena El-Hussein, Antonella Delmestri, Cyrus Cooper, Moayyeri Alireza, Cesar Libanati, Emese Toth, Daniel Prieto-Alhambra, Sara Khalid

**Author notes:** Joint first authors (Marta Pineda-Moncusí, Leena El-Hussein).

## Abstract

The objective of this work was to estimate the incidence rate of cardiovascular disease (CVD) events (myocardial infarction, stroke or CVD-death) at 1 year among 3 cohorts of patients at high risk of fracture (osteoporosis; previous fracture; and anti-osteoporosis medication), and to identify the key risk factors of CVD events in these three cohorts. To do so, this prospective cohort study used data from the Clinical Practice Research Datalink, a primary care database from United Kingdom. MACE events (a composite outcome for the occurrence of either myocardial infarction (MI), stroke or CVD death) were identified in patients aged fifty or over at high or imminent fracture risk identified in three different cohorts (not mutually exclusive): recently diagnosed with osteoporosis (OST, n=65,295), incident fragility fracture (IFX, n=80,587), and starting oral bisphosphonates (OBP, n=145,959). MACE incidence rates (cases/1000 person-years) were: 19.63 (18.54;20.73) in OST, 51.09 (49.39;52.80) in IFX, and 26.26 (25.41;27.12) in OBP cohorts. Candidate risk factors from QRISK tool, and candidates from literature selected by lasso regression, were fitted into prediction models to estimate 1-year risk of MACE for each cohort. OST and OBP models had a discrimination of ≥ 70% and IFX cohort ≥ 60%. Calibration plots showed good internal validity of the models. Generally, models from lasso regression were better than QRISK. Main risk factors common in all MACE models were sex, alcohol, atrial fibrillation, anti-hypertensive medication, age, prior MI/stroke, established CVD, glomerular filtration rate, systolic blood pressure, and number of GP visits and concomitant medicines. Incidence of MACE in the studied populations was 4.35% or lower, with IFX as the cohort with the higher risk. Identified key risk factors highlight the differences of patients at high risk of fracture versus general population. Proposed models could improve prediction of CVD events in patients with osteoporosis in primary care settings.

## Introduction

Cardiovascular disease (CVD) and osteoporosis are both worldwide leading causes of morbidity and mortality^1 2^ and their prevalence increases with age.^3 4^ Several prediction tools have been developed for cardiovascular events (heart attacks or stroke), including the Framingham Heart Study,^5-7^ CHADS2 tool,^8-10^ and QRISK tool.^11 12^ These tools have been developed in general, usually younger, populations and have not been validated in patients with osteoporosis.

Identification of CVD risk factors is particularly challenging for patients with osteoporosis since the association between fracture risk or anti-osteoporosis treatment and cardiovascular events remains unclear.^13^ Some established risk factors for osteoporosis and fractures^14^ such as female gender and low weight have been found to be protective against CVD. Conversely, some other risk factors including age, low bone mineral density,^15^ prior fracture, obesity^16-18^ or type 2 diabetes^19^ are associated with an increased risk of CVD. Patients with a history of CVD have been shown to be at increased risk of osteoporotic fractures,^20^ while higher risk of stroke and coronary artery disease is observed among patients with osteoporotic fracture or low bone mineral density.^21^

The effects of anti-osteoporosis medications on CVD risk is inconclusive: despite no evidence from clinical trials that oral bisphosphonates (BP) have an impact on cardiovascular risk,^22 23^ some publications suggest a protective effect.^24^ Meanwhile, the European Medicines Agency (EMA) has advised contraindications to patients with a history of prior myocardial infarction (MI) or stroke regarding romosozumab, the most recent medication option for osteoporosis.^25^ This variable impact of anti-osteoporotic medication on CVD risk highlights the clinical utility of identifying patients who are being considered for osteoporosis treatment and might be at elevated risk of CVD.

To address this issue, our overarching aim was to assess the absolute risk of CVD experienced by elderly patients at higher fracture risk in the UK, as well as to identify key CVD risk factors (both generic and specific ones) for these patients. We estimate incidence rates of major adverse cardiovascular events (MACE) among cohorts newly diagnosed with osteoporosis, first recorded fracture, and oral BP therapy initiators obtained from the UK general population; and developed and internally validated models that predict 1-year MACE in these cohorts of high-risk patients. Additionally, a secondary analysis of 2-year MACE and a sensitivity analysis of MI/stroke prediction is reported.

## Methods

### Data Source

Data for this study were obtained from the Clinical Practice Research Datalink (CPRD) GOLD, which contains anonymised electronic primary care records for the UK (www.cprd.com/primarycare). In addition to demographic information, the data included medication prescriptions, clinical events, tests, referrals, and hospital admissions along with their major outcomes in a sample of >16 million patients (including deceased and transferred out; 2.3 million are current patients, covering approximately 3.6% of UK population).^26 27^ For this study, an extract from January 1, 1995 to January 31, 2017 was used. We used CPRD GOLD data linked to the Hospital Episode Statistics (HES) Admitted Patient Care, which contains clinical diagnoses during hospital admissions in England, to the Office for National Statistics (ONS) mortality records, and to the Index of Multiple Deprivation (IMD) dataset. We reconciled CPRD GOLD and ONS mortality dates of death following published guidelines.^28^

### Participants

The study population included patients at high or imminent fracture risk as identified from literature, divided into three cohorts:

1. Patients with an incident diagnosis of osteoporosis (read or ICD-10 codes). We refer to this group as the osteoporosis cohort (OST).
2. Patients with a first incident fracture at an osteoporotic site (all except face, skull and digits), diagnosed either through read codes or ICD-10 codes. This cohort is referred to as the imminent fracture risk cohort (IFX).
3. Incident users of oral bisphosphonates (BP) without BP use in the prior year, referred to as the oral BP treatment cohort (OBP).

Index date was defined as time of recorded incident diagnosis, first incident fracture, and incident use of BP, for the OST, IFX, and OBP cohorts, respectively. Participants were followed from index date up to a maximum of two years. We censored participants at the earliest of first: study outcome, death, transfer out of practice, or the end of follow-up period. Included patients were at least 50 years old and had at least one year of data available prior to index date. Participants could potentially be present in more than one of the cohorts above, with different index dates. For OBP cohort, subjects with use of any anti-osteoporotic drug (except calcium and vitamin D supplements) in the previous year were excluded.

### Candidate Risk Factors

The overall set of variables considered for inclusion in the prediction model contained risk factors from the QRISK model,^12^ as well as additional risk factors identified in the literature as being potentially associated with CVD.^15-17 20 21 29-37^ These included socio-demographic and lifestyle factors, laboratory measurements, medications, and co-morbidities (Table 1).

**Table 1.** Baseline characteristics of patients in the development set and the validation set.

| Variable | OST<br>n=65295 | IFX<br>n=80587 | OBP<br>n=145959 |
| --- | --- | --- | --- |
| Sex = Male (%) | 8616 (13.2) | 18794 (23.3) | 29547 (20.2) |
| SES (%) |  |  |  |
| 1 | 15953 (24.4) | 18155 (22.5) | 35837 (24.6) |
| 2 | 15643 (24.0) | 19194 (23.8) | 35738 (24.5) |
| 3 | 13794 (21.1) | 17177 (21.3) | 30877 (21.2) |
| 4 | 11921 (18.3) | 15381 (19.1) | 26566 (18.2) |
| 5 | 7918 (12.1) | 10613 (13.2) | 16828 (11.5) |
| Smoking** (%) |  |  |  |
| Ex | 20487 (31.4) | 25847 (32.1) | 50162 (34.4) |
| No | 34049 (52.1) | 41531 (51.5) | 75838 (52.0) |
| Yes | 10759 (16.5) | 13209 (16.4) | 19959 (13.7) |
| Drinking** (%) |  |  |  |
| Ex | 3564 (5.5) | 5676 (7.0) | 7226 (5.0) |
| No | 17062 (26.1) | 24325 (30.2) | 41791 (28.6) |
| Yes | 44669 (68.4) | 50586 (62.8) | 96942 (66.4) |
| Diabetes type I* (%) | 152 (0.2) | 223 (0.3) | 297 (0.2) |
| Diabetes type II* (%) | 3311 (5.1) | 5245 (6.5) | 8340 (5.7) |
| Chronic obstructive pulmonary disease* (%) | 4939 (7.6) | 4733 (5.9) | 11251 (7.7) |
| Chronic kidney disease* (%) | 5790 (8.9) | 10232 (12.7) | 13396 (9.2) |
| Rheumatoid arthritis* (%) | 5045 (7.7) | 3744 (4.6) | 19746 (13.5) |
| Lupus* (%) | 144 (0.2) | 75 (0.1) | 328 (0.2) |
| Systemic heart disease** (%) | 2341 (3.6) | 725 (0.9) | 5222 (3.6) |
| Anti-osteoporosis use** (%) | 11752 (18.0) | 7726 (9.6) |  |
| Heparin use** (%) | 372 (0.6) | 368 (0.5) | 1035 (0.7) |
| Beta-blocker use** (%) | 10564 (16.2) | 12698 (15.8) | 22753 (15.6) |
| Hypertension** (%) | 5091 (7.8) | 4048 (5.0) | 9514 (6.5) |
| Deep vein thrombosis or pulmonary embolism** (%) | 522 (0.8) | 545 (0.7) | 1334 (0.9) |
| Anticoagulant use** (%) | 3285 (5.0) | 4274 (5.3) | 7546 (5.2) |
| Antidepressants TCA** (%) | 7039 (10.8) | 6699 (8.3) | 14451 (9.9) |
| Antidepressants SSRI** (%) | 5990 (9.2) | 9209 (11.4) | 12651 (8.7) |
| Hypercholesterolemia** (%) | 1561 (2.4) | 843 (1.0) | 2543 (1.7) |
| Statin use** (%) | 15571 (23.8) | 17409 (21.6) | 33988 (23.3) |
| Osteoporosis history* |  | 8299 (10.3) | 46215 (31.7) |
| Family history of cardiovascular disease (%) | 6755 (10.3) | 5473 (6.8) | 13204 (9.0) |
| Family history of cardiovascular disease before age 60 (%) | 99 (0.2) | 58 (0.1) | 170 (0.1) |
| Heart failure* (%) | 2323 (3.6) | 4223 (5.2) | 5040 (3.5) |
| Migraine* (%) | 10103 (15.5) | 8672 (10.8) | 19961 (13.7) |
| Severe mental illness* (%) | 10203 (15.6) | 11911 (14.8) | 19256 (13.2) |
| Vascular Disease* (%) | 839 (1.3) | 1351 (1.7) | 1765 (1.2) |
| Atrial fibrillation* (%) | 3664 (5.6) | 5956 (7.4) | 8022 (5.5) |

| Variable | OST | IFX | OBP |
| --- | --- | --- | --- |
|  | n=65295 | n=80587 | n=145959 |
| On anti-hypertensive drug (%) | 37053 (56.7) | 47247 (58.6) | 79044 (54.2) |
| Antipsychotic use** (%) | 371 (0.6) | 1035 (1.3) | 773 (0.5) |
| Steroid use** (%) | 9367 (14.3) | 6423 (8.0) | 37201 (25.5) |
| Erectile dysfunction** (%) | 871 (1.3) | 1349 (1.7) | 2797 (1.9) |
| Age>75 | 31105 (47.6) | 57177 (71.0) | 77199 (52.9) |
| Age Group (%) |  |  |  |
| 50-59 | 8315 (12.7) | 6242 (7.7) | 16388 (11.2) |
| 60-69 | 15949 (24.4) | 9875 (12.3) | 31665 (21.7) |
| 70-79 | 21165 (32.4) | 18686 (23.2) | 45679 (31.3) |
| 80-89 | 16586 (25.4) | 32248 (40.0) | 42053 (28.8) |
| >89 | 3280 (5.0) | 13536 (16.8) | 10174 (7.0) |
| Charlson score (%) |  |  |  |
| 0 | 37782 (57.9) | 46485 (57.7) | 80717 (55.3) |
| 1 | 13274 (20.3) | 14012 (17.4) | 31132 (21.3) |
| 2 | 7960 (12.2) | 9380 (11.6) | 18141 (12.4) |
| ≥3 | 6279 (9.6) | 10710 (13.3) | 15969 (10.9) |
| Cardiovascular disease (%) |  |  |  |
| No | 58034 (88.9) | 69893 (86.7) | 128516 (88.0) |
| Ever >1 year before index date | 5616 (8.6) | 8702 (10.8) | 13474 (9.2) |
| 1 year before index | 719 (1.1) | 872 (1.1) | 1551 (1.1) |
| 6 months before index | 723 (1.1) | 909 (1.1) | 1763 (1.2) |
| 1 month before index | 203 (0.3) | 211 (0.3) | 655 (0.4) |
| MI or Stroke (%) |  |  |  |
| No | 61350 (94.0) | 72143 (89.5) | 135335 (92.7) |
| Ever >1 year before index date | 2850 (4.4) | 6172 (7.7) | 7481 (5.1) |
| 1 year before index | 1095 (1.7) | 2272 (2.8) | 3143 (2.2) |
| Established CVD *= Ever (%) | 7428 (11.4) | 13942 (17.3) | 18774 (12.9) |
| Any fracture history (%) |  |  |  |
| No | 49542 (75.9) | 67065 (83.2) | 114386 (78.4) |
| Ever >1 year before index date | 5813 (8.9) | 7298 (9.1) | 10598 (7.3) |
| 1 year before index | 9940 (15.2) | 6224 (7.7) | 20975 (14.4) |
| Hip fracture history (%) |  |  |  |
| No | 62040 (95.0) | 80569 (100.0) | 136610 (93.6) |
| Ever >1 year before index date | 1118 (1.7) | 5 (0.0) | 1720 (1.2) |
| 1 year before index | 2137 (3.3) | 13 (0.0) | 7629 (5.2) |
| Shoulder fracture history (%) |  |  |  |
| No | 64836 (99.3) | 80258 (99.6) | 145199 (99.5) |
| Ever >1 year before index date | 232 (0.4) | 159 (0.2) | 333 (0.2) |
| 1 year before index | 227 (0.3) | 170 (0.2) | 427 (0.3) |
| Spine fracture history (%) |  |  |  |
| No | 63880 (97.8) | 79625 (98.8) | 143554 (98.4) |
| Ever >1 year before index date | 326 (0.5) | 630 (0.8) | 356 (0.2) |
| 1 year before index | 1089 (1.7) | 332 (0.4) | 2049 (1.4) |
| Wrist fracture history (%) |  |  |  |
| No | 60237 (92.3) | 75102 (93.2) | 137118 (93.9) |
| Ever >1 year before index date | 2363 (3.6) | 2652 (3.3) | 4040 (2.8) |
| 1 year before index | 2695 (4.1) | 2833 (3.5) | 4801 (3.3) |
| BMI** (%) |  |  |  |
| <18.5 | 5615 (8.6) | 2194 (2.7) | 10877 (7.5) |
| 18.6 - 24.9 | 32564 (49.9) | 21678 (26.9) | 66542 (45.6) |
|  | n=65295 | n=80587 | n=145959 |
| 25 - 29.9 | 18343 (28.1) | 29565 (36.7) | 44109 (30.2) |
| 30 - 39.9 | 8164 (12.5) | 26563 (33.0) | 22452 (15.4) |
| >=40 | 609 (0.9) | 587 (0.7) | 1979 (1.4) |
| No. of GP visits** (%) |  |  |  |
| 0 | 3219 (4.9) | 16717 (20.7) | 15177 (10.4) |
| 1-5 | 16176 (24.8) | 18735 (23.2) | 36213 (24.8) |
| 6-10 | 17452 (26.7) | 16762 (20.8) | 32344 (22.2) |
| 11-15 | 11938 (18.3) | 11249 (14.0) | 24466 (16.8) |
| >=16 | 16510 (25.3) | 17124 (21.2) | 37759 (25.9) |
| No. of GP emergency visits** (%) |  |  |  |
| 0 | 53095 (81.3) | 63284 (78.5) | 119698 (82.0) |
| 1 | 6570 (10.1) | 8042 (10.0) | 14083 (9.6) |
| 2 | 2439 (3.7) | 3524 (4.4) | 5282 (3.6) |
| 3-5 | 2282 (3.5) | 3926 (4.9) | 4993 (3.4) |
| >=6 | 909 (1.4) | 1811 (2.2) | 1903 (1.3) |
| eGFR** (%) |  |  |  |
| <=29 | 708 (1.1) | 2232 (2.8) | 1777 (1.2) |
| 30 - 44 | 3470 (5.3) | 8228 (10.2) | 9749 (6.7) |
| 45 - 59 | 11950 (18.3) | 17901 (22.2) | 30290 (20.8) |
| 60 - 89 | 45497 (69.7) | 48714 (60.4) | 96838 (66.3) |
| >=90 | 3670 (5.6) | 3512 (4.4) | 7305 (5.0) |
| SBP** (%) |  |  |  |
| <120 | 9076 (13.9) | 11081 (13.8) | 18549 (12.7) |
| 120 - 139 | 26605 (40.7) | 31513 (39.1) | 57685 (39.5) |
| 140 - 159 | 22588 (34.6) | 27679 (34.3) | 51589 (35.3) |
| >=160 | 7026 (10.8) | 10314 (12.8) | 18136 (12.4) |
| DBP** (%) |  |  |  |
| <80 | 33729 (51.7) | 43144 (53.5) | 75132 (51.5) |
| 80 - 89 | 24505 (37.5) | 28496 (35.4) | 53798 (36.9) |
| 90 - 99 | 5764 (8.8) | 7034 (8.7) | 13678 (9.4) |
| >=100 | 1297 (2.0) | 1913 (2.4) | 3351 (2.3) |
| No. of concomitant medicines** (%) |  |  |  |
| 0 | 5171 (7.9) | 18117 (22.5) | 20662 (14.2) |
| 1 - 3 | 12858 (19.7) | 12417 (15.4) | 26686 (18.3) |
| 4 - 6 | 14446 (22.1) | 15115 (18.8) | 29125 (20.0) |
| 7 - 9 | 12375 (19.0) | 13314 (16.5) | 26049 (17.8) |
| 10 - 12 | 8788 (13.5) | 9389 (11.7) | 18868 (12.9) |
| >=13 | 11657 (17.9) | 12235 (15.2) | 24569 (16.8) |
| Cholesterol measurement**<br>(HDL/LDL) (%) |  |  |  |
| <=3.5 | 39143 (59.9) | 52781 (65.5) | 86247 (59.1) |
| 3.6 - 5 | 20792 (31.8) | 22077 (27.4) | 46654 (32.0) |
| >5 | 5360 (8.2) | 5729 (7.1) | 13058 (8.9) |
| No. of previous fractures* (%) |  |  |  |
| 0 | 46551 (71.3) | 42442 (52.7) | 112120 (76.8) |
| 1 | 9763 (15.0) | 26101 (32.4) | 18129 (12.4) |
| >=2 | 8981 (13.8) | 12044 (14.9) | 15710 (10.8) |

**Abbreviations:** OST, patients with incident diagnosis of osteoporosis; IFX, patients with incident fragility fracture; OBP, incident users of oral bisphosphonates; \* ever; \*\* in the year prior to start; SES, socio-economic status; MI, myocardial infarction; BMI, body mass index;

| Variable | OST | IFX | OBP |
| --- | --- | --- | --- |
|  | n=65295 | n=80587 | n=145959 |
| eGFR, estimated Glomerular Filtration Rate; SBP, cholesterol, systolic blood pressure; DBP, diastolic blood pressure. |  |  |  |

### Outcomes

The main outcome of the study was one-year occurrence of MACE:

MACE: a composite outcome of the first occurrence of either stroke, MI or death due to CVD (recorded as the primary cause of death in ONS).

Additionally, secondary analysis of two-year occurrence of MACE and sensitivity analysis excluding death (MI/stroke) at one- and two-year was reported in supplementary data:

MI/stroke: a composite outcome of the first occurrence of either stroke or MI.

### Statistical analysis

Baseline characteristics of all three cohorts were described. Incidence rates (IR) and their 95% confidence interval of each outcome at one and two years after index date were calculated (cases/1000 person-years) through ERIC Notebook person-time methodology.^38^

Then, performance of QRISK variables to estimate the one-year risk of MACE was assessed (QRISK variables are listed in Table S1). Finally, all the available candidate risk factors described in the above were combined into a prediction model (henceforth referred to as “ALL”). Lasso regression selected the key risk factors which were then entered into a final logistic regression equation. Model performance was evaluated for this final equation and model coefficients and intercept terms reported.

Missing data was handled using multiple imputation and combined using Rubin’s rules as required.^39^ Figure 1 describes the steps used to build the final model. Further explanations of the prediction model development are described in *supplementary file A*.

**Figure 1.**
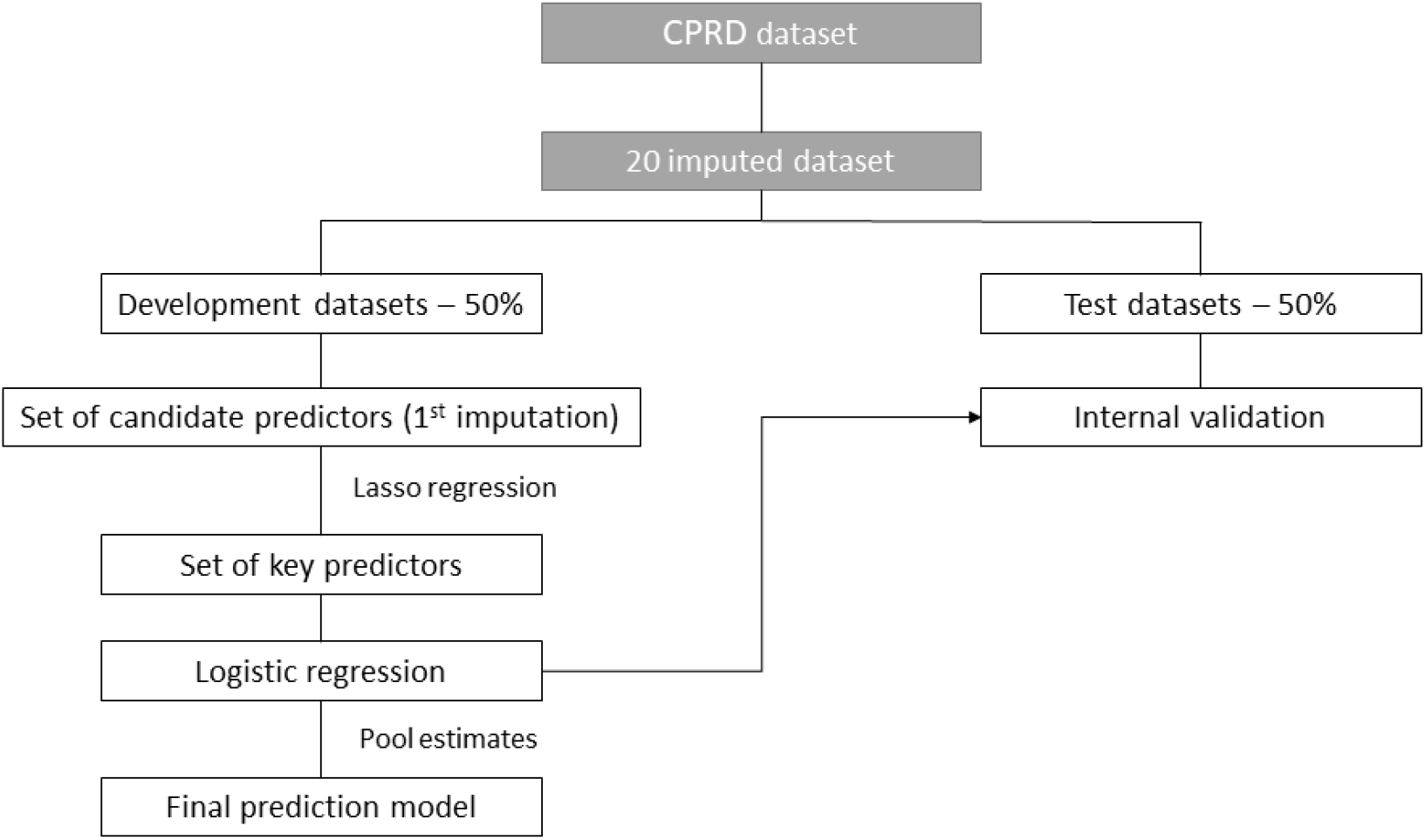
Steps in the development and validation of the prediction model.

The same steps were repeated for two-year MACE (secondary analysis), for one- and two-year MI/stroke outcomes (sensitivity analysis), and for gender-based models, included in the *supplementary file A*.

### Models Performance

We assessed the models internally using the validation datasets. Discrimination was evaluated by calculating the area under the curve (AUC). The AUC was produced for all 20 validation datasets then pooled using Rubin’s rules. Calibration was assessed by producing calibration plots of observed versus predicted probabilities, in tenths of predicted risk. Calibration plots were also produced for 10-year age groups and gender.

All statistical analyses took place in R version 3.6.0, including MICE, glmnet, rpart, gbm, caret, flextable, pROC and officer packages.

### Patient and public involvement

Used data was previously collected and all participant records were linked-anonymised. Hence, no patients or members of the public were directly implicated in the design or analysis of the reported data.

## Results

A total of 65,295, 80,587 and 145,959 participants were included in the OST, IFX and OBP cohorts, respectively (Fig 2). Baseline characteristics of each cohort are shown in Table 1. Mean age (years [standard deviation]) of each population was: 73.05 [10.67] in OST, 79.11 [11.22] in IFX and 74.35 [10.88] in OBP. Baseline characteristics stratified by outcome for the development and validation datasets are provided in the supplements (Table S2a-c).

**Figure 2.**
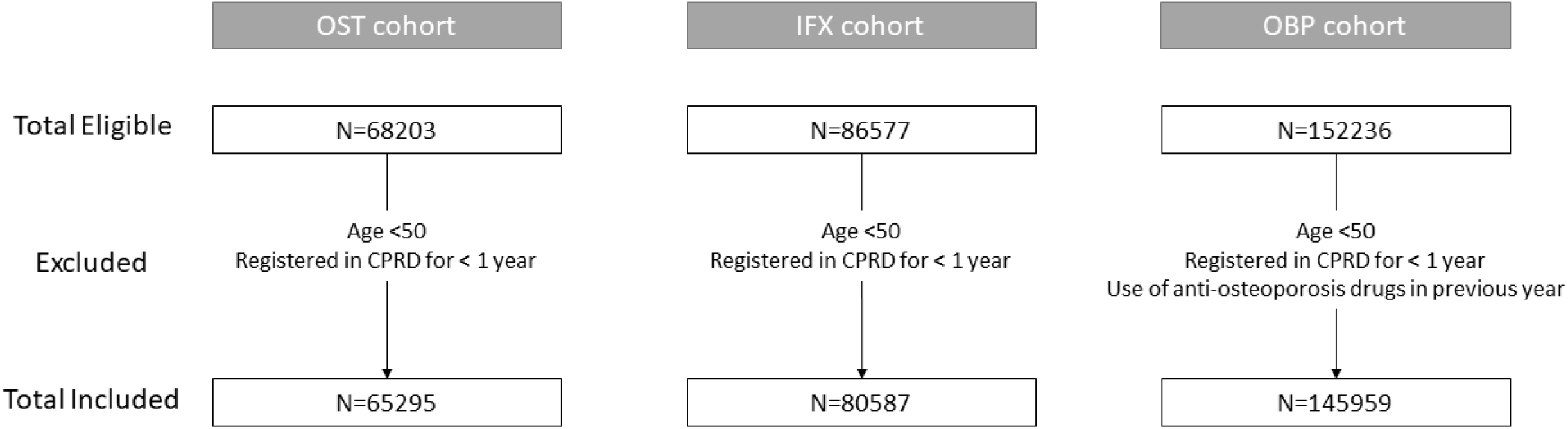
Study flow chart. Abbreviations: OST, patients with incident diagnosis of osteoporosis; IFX, patients with incident fragility fracture; OBP, incident users of oral bisphosphonates; CPRD, Clinical Practice Research Datalink.

During follow-up, 1,238 patients experienced MACE at one-year in the OST cohort, and the IR (95% CI) in units of cases/1000 person-years was 19.6 (18.5, 20.7). For the IFX cohort, one-year MACE IR was 51.1 (49.4, 52.8), while in the OBP cohort, IR was 26.3 (25.4, 27.1). Those who experienced MACE were older in general (higher proportion aged >75) with higher comorbidity (Charlson score), and higher prevalence of drug use (e.g., anti-hypertensives and beta-blockers).

Figure 3 summarises the IRs along with the number of occurrences and total follow-up for each outcome. Figure S1a-d in the supplements describes the IR of 1- and 2-years MACE and stroke/MI outcomes stratified by age groups, showing that IFX cohort had the highest incidences. Figure S2a-d display the IRs of two-year MACE, one- and two-year stroke/MI, and MACE and stroke/MI outcomes stratified by gender. Overall, despite the considerably lower proportion of males in this study, they suffered higher incidence rates of both outcomes in the one and two-year follow-up periods.

**Figure 3.**
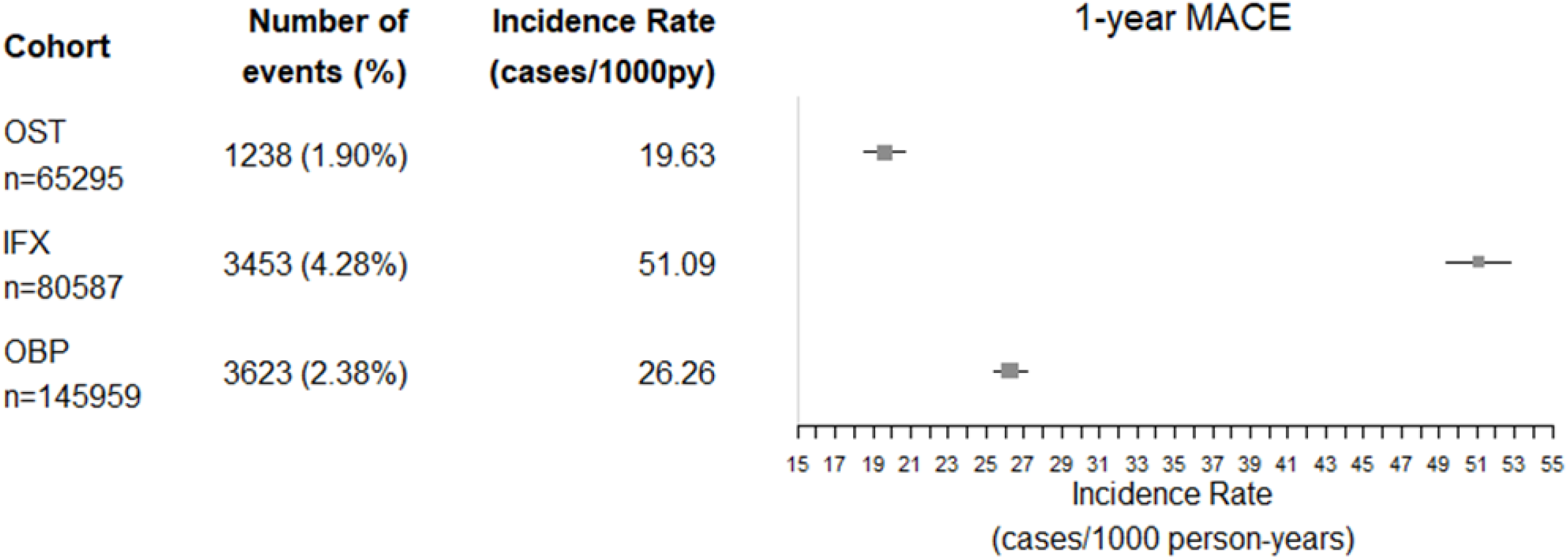
Incidence rates of MACE after one-of follow up. Incidence rate is reported with 95% confidence intervals. Abbreviations: OST, patients with incident diagnosis of osteoporosis; IFX, patients with incident fragility fracture; OBP, incident users of oral bisphosphonates; MACE, composite outcome for the occurrence of either myocardial infarction, stroke or cardiovascular disease death.

### Development and performance of the prediction model

Figure 4 summarises AUC values for the MACE models using QRISK and lasso regression selection of risk factors. The derived prediction models showed good discrimination (AUC was above 70%) for both OST and OBP models, while AUC values in IFX model were 66% and 69% in QRISK and lasso selection, respectively. Figure S3a-b report AUC values for two-year MACE and MI/stroke, respectively. The difference was minimal between one and two-year prediction models with no apparent pattern. MACE models had higher AUC values compared to the stroke/MI models. Predictive models using risk factors identified from lasso equated or outperformed the models using QRISK factors.

**Figure 4.**
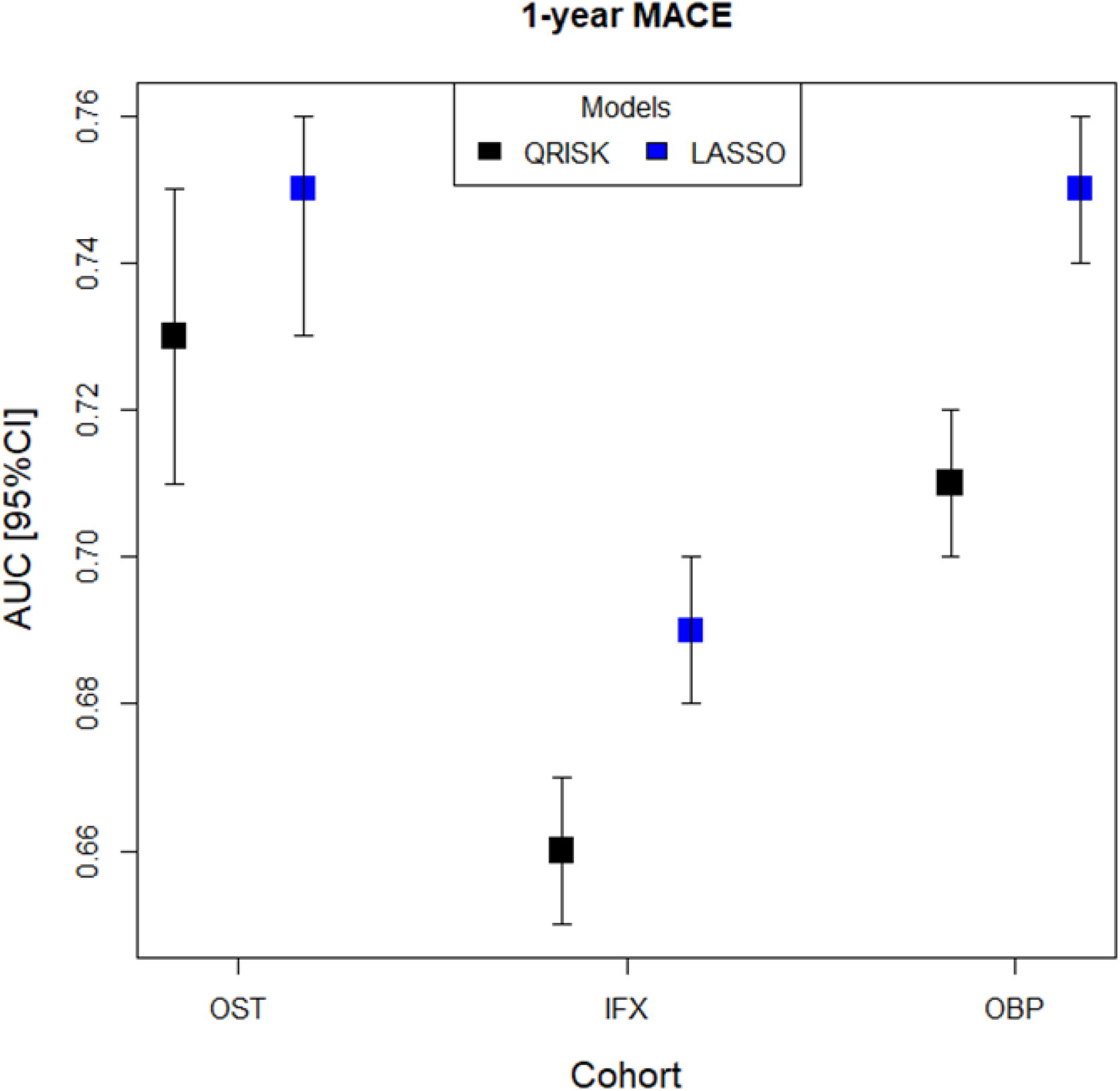
Area under ROC curve for internal validation of one-year MACE outcome using risk factors from QRISK and LASSO models. Abbreviations: OST, patients with incident diagnosis of osteoporosis; IFX, patients with incident fragility fracture; OBP, incident users of oral bisphosphonates; AUC, area under the curve; MACE, composite outcome for the occurrence of either myocardial infarction, stroke or cardiovascular disease death; MI, myocardial infarction.

Figure 5 shows the calibration plots of the one-year MACE models stratified by age and figure 6 shows the calibration of these models stratified by age and gender. Figure S4a-b presents the calibration plots of two-year MACE by age, and by age and gender, respectively. Figures S5 and S6 displays the calibration plots of the MI/stroke models stratified by age, and by age and gender, respectively. Generally, models were well-calibrated, with an over-predicting risk for the population <60 years old and under-predicting for those >80, probably caused by the lower proportion of participants belonging to either category. Figure S3c-d reports the AUC of gender-based models. The lower AUC values for men-only models could be related to the considerably lower sample size of those cohorts.

**Figure 5.**
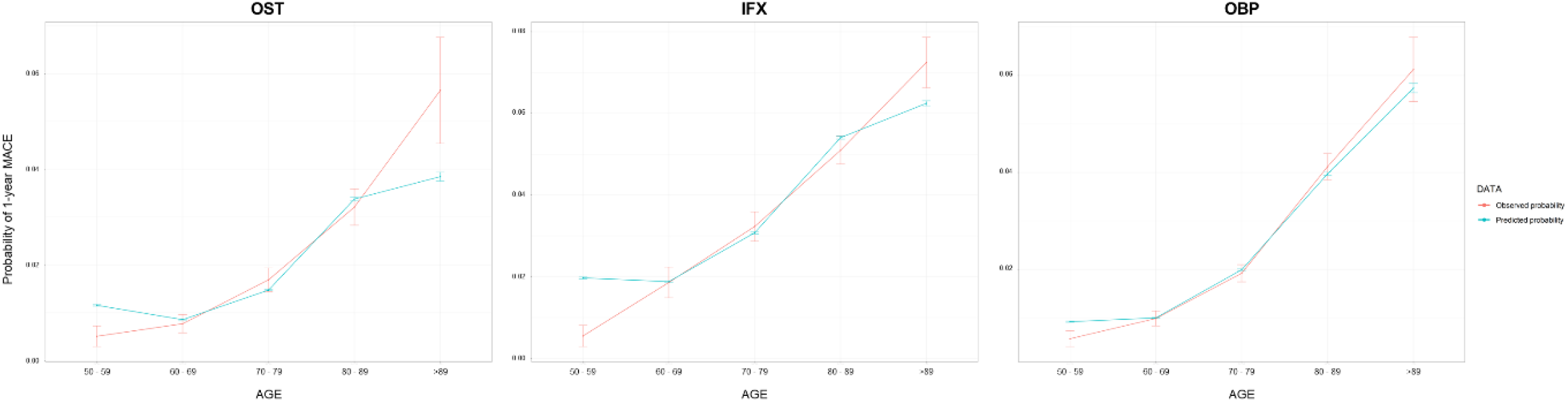
Calibration curves for internal validation of one-year MACE prediction by age deciles. Models using risk factors selected by lasso regression. From left to right: OST, IFX and OBP cohorts. Abbreviations: OST, patients with incident diagnosis of osteoporosis; IFX, patients with incident fragility fracture; OBP, incident users of oral bisphosphonates; MACE, composite outcome for the occurrence of either myocardial infarction, stroke or cardiovascular disease death.

**Figure 6.**
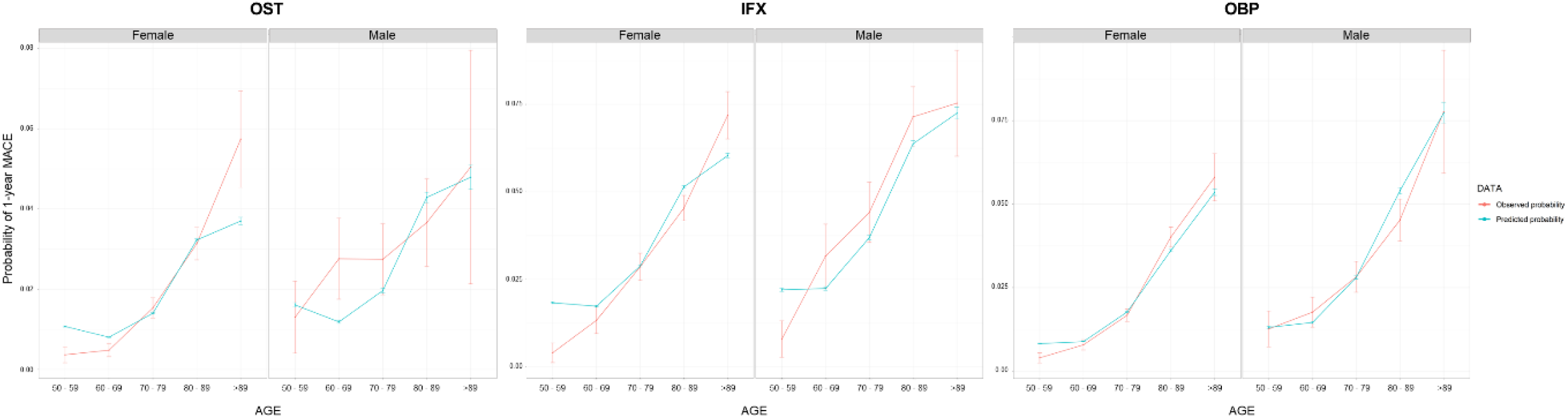
Calibration curves of one-year MACE prediction stratified by age and gender. Models using risk factors selected by lasso regression. From left to right: OST, IFX and OBP cohorts. Abbreviations: OST, patients with incident diagnosis of osteoporosis; IFX, patients with incident fragility fracture; OBP, incident users of oral bisphosphonates; MACE, composite outcome for the occurrence of either myocardial infarction, stroke or cardiovascular disease death.

Table 2 displays the risk factors selected from lasso for the overall models of one-year MACE along with their odd ratios (OR) and confidence intervals, and table S3 lists its beta coefficients. Gender, drinking, atrial fibrillation in the prior year, use of anti-hypertensive medication, age, prior MI or stroke, CVD history, number of GP visits and concomitant medicines in the prior year, and eGFR and SBP measurements in the prior year, appeared in all models. Predictors common in two of the three cohort models were smoking, heart failure, number of previous fractures, Charlson score, BMI; and number of GP emergency visits, DBP and Cholesterol measurements in the year prior to start.

**Table 2.** Predictors of one-year MACE overall models (risk factors selected by lasso regression)

| Predictor | OST<br>OR (95%CI) | IFX<br>OR (95%CI) | OBP<br>OR (95%CI) |
| --- | --- | --- | --- |
| Sex = Male (%) | 1.61 (1.3, 2) | 1.45 (1.29, 1.63) | 1.42 (1.25, 1.61) |
| SES (%) | x | x |  |
| 1 | x | x | ref |
| 2 | x | x | 1.17 (1.02, 1.36) |
| 3 | x | x | 1.28 (1.11, 1.48) |
| 4 | x | x | 1.34 (1.15, 1.56) |
| 5 | x | x | 1.29 (1.08, 1.54) |
| Smoking** |  | x |  |
| Ex | ref | x | ref |
| No | 0.93 (0.73, 1.18) | x | 0.98 (0.79, 1.23) |
| Yes | 1.42 (1.02, 1.98) | x | 1.14 (0.89, 1.47) |
| Drinking** |  |  |  |
| Ex | ref | ref | ref |
| No | 1.19 (0.7, 2) | 1.04 (0.78, 1.39) | 1.03 (0.71, 1.49) |
| Yes | 0.84 (0.51, 1.39) | 0.95 (0.74, 1.22) | 0.92 (0.65, 1.31) |
| Diabetes type I* | x | x | x |
| Diabetes type II* | x | x | 1.11 (0.9, 1.36) |
| Chronic obstructive pulmonary disease* | x | x | 1.05 (0.87, 1.27) |
| Chronic kidney disease* | x | x | 0.81 (0.61, 1.06) |
| Rheumatoid arthritis* | x | x | 0.88 (0.74, 1.04) |
| Lupus* | x | x | x |
| Systemic heart disease** | x | x | 0.62 (0.37, 1.03) |
| Anti-osteoporosis use** | x | x | x |
| Heparin use** | x | x | x |
| Beta-blocker use** | x | x | 1.16 (1.02, 1.33) |
| Hypertension** | x | x | x |
| Deep vein thrombosis or pulmonary embolism** | x | x | x |
| Anticoagulant use** | x | x | 0.93 (0.75, 1.14) |
| Antidepressants TCA** | x | x | x |
| Antidepressants SSRI** | x | x | 1.28 (1.09, 1.49) |
| Hypercholesterolemia** | x | x | x |
| Statin use** | x | x | x |
| Osteoporosis history* | x | x | 0.74 (0.66, 0.83) |
| Family history of cardiovascular disease | x | x | 0.94 (0.78, 1.14) |
| Family history of cardiovascular disease before age 60 | x | x | x |
| Heart failure* | x | 1.17 (0.97, 1.4) | 1.03 (0.84, 1.27) |
| Migraine* | x | x | x |
| Severe mental illness* | x | x | x |
| Vascular Disease* | x | x | x |
| Atrial fibrillation* | 1.61 (1.27, 2.05) | 1.12 (0.96, 1.31) | 1.29 (1.08, 1.55) |
| On anti-hypertensive drug | 1.23 (0.97, 1.55) | 1.3 (1.12, 1.51) | 1.05 (0.9, 1.22) |
| Antipsychotic use** | X | X | x |
| Steroid use** |  | X | x |
| Erectile dysfunction** | X | X | x |
| Age Group (%) |  |  |  |
| 50-59 | ref | ref | ref |
| 60-69 | 1.36 (0.79, 2.35) | 3.3 (1.87, 5.84) | 1.58 (1.14, 2.19) |
| 70-79 | 3.07 (1.85, 5.11) | 5.38 (3.13, 9.26) | 2.46 (1.79, 3.36) |
| 80-89 | 4.9 (2.9, 8.28) | 8.55 (4.99, 14.65) | 4.26 (3.09, 5.87) |
| >89 | 7.88 (4.44, 13.99) | 10.88 (6.3, 18.78) | 5.79 (4.06, 8.24) |
| Charlson score | x |  |  |
| 0 | x | ref | ref |
| 1 | x | 1.01 (0.87, 1.16) | 1.01 (0.87, 1.17) |
| 2 | x | 0.92 (0.77, 1.08) | 0.94 (0.79, 1.13) |
| ≥3 | x | 0.99 (0.83, 1.18) | 1.06 (0.85, 1.31) |
| Cardiovascular disease | x | x |  |
| No | x | x | ref |
| Ever >1 year before index date | x | x | 1.16 (1, 1.36) |
| 1 year before index | x | x | 1.18 (0.84, 1.66) |
| 6 months before index | x | x | 1.52 (1.14, 2.03) |
| 1 month before index | x | x | 2.18 (1.49, 3.21) |
| MI or Stroke |  |  |  |
| No | ref | ref | ref |
| Ever >1 year before index date | 0.99 (0.7, 1.4) | 1.42 (1.16, 1.74) | 1.34 (1.09, 1.64) |
| 1 year before index | 2.03 (1.38, 2.99) | 2.1 (1.65, 2.67) | 2.52 (2, 3.18) |
| Established CVD * | 1.9 (1.46, 2.48) | 1.35 (1.14, 1.61) | 1.49 (1.25, 1.79) |
| Any fracture history | x | x | x |
| No | x | x | x |
| Ever >1 year before index date | x | x | x |
| 1 year before index | x | x | x |
| Hip fracture history | x | x | x |
| No | x | x | x |
| Ever >1 year before index date | x | x | x |
| 1 year before index | x | x | x |
| Shoulder fracture history | x | x | x |
| No | x | x | x |
| Ever >1 year before index date | x | x | x |
| 1 year before index | x | x | x |
| Spine fracture history | x | x | x |
| No | x | x | x |
| Ever >1 year before index date | x | x | x |
| 1 year before index | x | x | x |
| Wrist fracture history | x | x | x |
| No | x | x | x |
| Ever >1 year before index date | x | x | x |
| 1 year before index | x | x | x |
| BMI** |  | x |  |
| <18.5 | ref | x | ref |
| 18.6 - 24.9 | 0.72 (0.49, 1.05) | x | 0.74 (0.57, 0.96) |
| 25 - 29.9 | 0.56 (0.35, 0.9) | x | 0.58 (0.41, 0.81) |
| 30 - 39.9 | 0.42 (0.2, 0.85) | x | 0.55 (0.36, 0.84) |
| ≥40 | 0.66 (0.22, 1.93) | x | 0.39 (0.17, 0.91) |
| No. of GP visits** |  |  |  |
| 0 | ref | ref | ref |
| 1-5 | 1.01 (0.61, 1.66) | 0.89 (0.67, 1.19) | 0.9 (0.73, 1.1) |
| 6-10 | 0.94 (0.56, 1.59) | 0.81 (0.6, 1.11) | 0.78 (0.61, 1.01) |
| 11-15 | 0.95 (0.55, 1.62) | 0.9 (0.65, 1.23) | 0.78 (0.6, 1.01) |
| ≥16 | 1.07 (0.63, 1.81) | 0.85 (0.62, 1.17) | 0.86 (0.67, 1.12) |
| No. of GP emergency visits** | x |  |  |
| 0 | x | ref | ref |
| 1 | x | 1.17 (1, 1.37) | 1.15 (0.99, 1.35) |
| 2 | x | 1.36 (1.11, 1.66) | 1.4 (1.13, 1.74) |
| 3-5 | x | 1.12 (0.91, 1.37) | 1.47 (1.2, 1.8) |
| ≥6 | x | 1.31 (1.01, 1.7) | 2.53 (1.96, 3.26) |
| eGFR** |  |  |  |
| ≤29 | ref | ref | ref |
| 30 – 44 | 1.11 (0.44, 2.81) | 0.98 (0.67, 1.42) | 0.91 (0.56, 1.5) |
| 45 – 59 | 0.89 (0.37, 2.15) | 0.79 (0.48, 1.3) | 0.83 (0.49, 1.4) |
| 60 – 89 | 0.76 (0.31, 1.86) | 0.71 (0.42, 1.19) | 0.58 (0.27, 1.27) |
| ≥90 | 0.79 (0.3, 2.07) | 0.65 (0.34, 1.22) | 0.5 (0.21, 1.22) |
| SBP** |  |  |  |
| <120 | ref | ref | ref |
| 120 - 139 | 1.19 (0.88, 1.61) | 1.06 (0.89, 1.26) | 1.09 (0.92, 1.31) |
| 140 - 159 | 1.32 (0.98, 1.77) | 1.08 (0.88, 1.33) | 1.24 (1.02, 1.52) |
| ≥160 | 1.32 (0.91, 1.89) | 1.36 (1.05, 1.77) | 1.39 (1.09, 1.77) |
| DBP** | x |  |  |
| <80 | x | ref | ref |
| 80 - 89 | x | 1.05 (0.86, 1.27) | 0.94 (0.82, 1.08) |
| 90 - 99 | x | 1.25 (0.94, 1.65) | 1.05 (0.84, 1.31) |
| ≥100 | x | 1.11 (0.64, 1.9) | 1.31 (0.89, 1.93) |
| No. of concomitant medicines** |  |  |  |
| 0 | ref | ref | ref |
| 1 – 3 | 0.69 (0.42, 1.12) | 1.07 (0.79, 1.46) | 0.87 (0.7, 1.07) |
| 4 – 6 | 0.83 (0.51, 1.36) | 1.15 (0.84, 1.56) | 0.75 (0.59, 0.96) |
| 7 – 9 | 1.01 (0.61, 1.66) | 1.31 (0.95, 1.79) | 0.88 (0.68, 1.14) |
| 10 – 12 | 1.12 (0.67, 1.87) | 1.37 (0.98, 1.9) | 1 (0.76, 1.31) |
| >=13 | 1.11 (0.66, 1.88) | 1.33 (0.96, 1.86) | 0.98 (0.74, 1.31) |
| Cholesterol measurement** (HDL/LDL) |  | x |  |
| <=3.5 | ref | x | ref |
| 3.6 – 5 | 1.34 (0.92, 1.95) | x | 1.2 (0.98, 1.46) |
| >5 | 1.74 (0.94, 3.2) | x | 1.35 (0.89, 2.03) |
| No. of previous fractures* |  |  | x |
| 0 | ref | ref | x |
| 1 | 1.24 (1.01, 1.53) | 0.91 (0.82, 1.02) | x |
| >=2 | 0.99 (0.79, 1.25) | 0.89 (0.76, 1.03) | x |

**Abbreviations:** OST, patients with incident diagnosis of osteoporosis; IFX, patients with incident fragility fracture; OBP, incident users of oral bisphosphonates; OR, odds ratio; CI, confidence intervals; MACE, composite outcome for the occurrence of either myocardial infarction, stroke or cardiovascular disease death; \* ever; \*\* in the year prior to start; SES, socio-economic status; MI, myocardial infarction; BMI, body mass index; eGFR, estimated Glomerular Filtration Rate; SBP, cholesterol, systolic blood pressure; DBP, diastolic blood pressure.

Table S4a-b summarise risk factors selected from lasso and ORs for two-year MACE and for MI/stroke models, respectively, and Table S5a-b list its beta coefficients. Table S6a-c reports all gender-based models and Table S7a-c lists its beta coefficients.

Detailed explanation and an example of how to obtain an estimate for an individual is reported in *supplementary file A*.

## Discussion

In this study, we evaluated the incidence of major adverse cardiovascular events though a composite outcome, MACE,^40^ and assessed risk factors of CVD to predict this outcome at one-year in three different cohorts. The IFX cohort can be used for a secondary fracture prevention program, the OBP cohort has the potential to be used in primary prevention since it approximates patients newly diagnosed and treated for osteoporosis, while the OST cohort can be used as a general screening in primary care.

We observed that patients’ incidence of MACE was slightly higher at one year than at two years, especially for IFX cohort. When stratifying by gender, men had higher incidence rates than women, which goes along with the results published by the British Health Foundation, where male incidences at UK in 2017 were higher than female’s (age-standardised incidence rate [per 100.000 inhabitants] for cardiovascular conditions: 1093.6 for men and 702.4 for women; for myocardial infarction: 192.0 for men and 82.9 for women; and for stroke/transient ischaemic attacks: 319.0 for men and 261.6 for women).^41^ In this line, IFX cohort had the highest incidence reported for MACE (51.1 /1000 person-years) which could be explained by this cohort having an older age (71% were older than 75 years old) and the biggest proportion of men (23.3%) among the three cohorts, followed by OBP (26.3, 20.2%) and OST (19.6, 13.2%) cohorts. The observation of higher incidences in IFX cohort was consistent when IR of each study cohort was stratified by age groups.

Fitting the list of risk factors from QRISK into a prediction model for MACE events, we obtained an AUC of 0.73, 0.66 and 0.71 in OST, IFX and OBP cohorts, respectively. However, starting from ALL risk factors list of CVD available in CPRD and selecting the most important through lasso regression, we obtained model equations that exceed QRISK (AUC in selected risk factors from ALL set: 0.75 in OST, 0.69 in IFX, and 0.75 in OBP). This list included generic features and those specific to the study population, and all of them can be found readily in primary care data. Among them, age had the largest statistically significant effect size.

Comparing our models to the existing cardiovascular prediction tools in general population, performance of Framingham and QRISK studies were: AUC >0.76 and >0.86, respectively.^5 11^ Framingham equations had been validated and recalibrated multiple times using different populations,^42^ while QRISK has a higher accuracy for UK population than the Framingham tool,^43^ However, both tools only permit risk calculation over long periods and there are no studies extrapolating them to shorter risk intervals. Additionally, these tools were not developed for the osteoporotic/fracture risk population, and they do not include specific risk factors for these particular patients (e.g., prior fractures and alcohol consumption), in whom short-term cardiovascular risk might be over- or underestimated. This can be observed in the performance of our sensitivity analysis of MI/Stroke outcome using QRISK list: the AUC values of QRISK in general population drops into a range of 0.62 to 0.70 when applying to osteoporotic/fracture risk population.

The proposed predictive models have good predictive power and internal validity (discrimination and calibration) in OBP and OST cohorts for one-year MACE events (the obtained equations are included in this article). However, IFX models did not exceed the 70% AUC threshold, considered as the minimum acceptable discrimination.^44^ Nonetheless, use of 1-year MACE model for IFX women is valuable since its AUC was 0.70%, there is a high incidence of this outcome in IFX cohort, and there are no existing models for this specific population. Secondary and sensitivity analysis show no differences using 2-years models and better performance of MACE than or MI/Stroke models.

### Strengths and limitations of this study

The proposed study is observational in nature, and hence cannot address causality but rather describe associations. There is no guarantee that all possible risk factors are included, but for all those factors that are, multivariable regression ensures that they are adjusted for (and hence reducing the risk of confounding). The three presented cohorts are not mutually exclusive but encompass the diversity of the population at high risk of fracture, and the different criteria used to evaluate them. Another limitation is the lack of external validity, which can be assessed in future studies to ensure the validity of the models across different populations. The enhanced performance observed in female population were expected due to the higher representation of them in our cohorts. The main strengths of this study are the large sample size and the application of machine learning for risk prediction^.45^

## Conclusions

To summarise, the incidence of MACE in the populations studied was 4.3% or lower, with IFX as the cohort with the higher risk. Efforts in predicting the study events states the differences between general and the osteoporotic/fracture risk population. The resulting algorithms include risk factors specific to the study population as well as more generic features that can be found easily in primary care data. Further work will focus on validating these models in external cohorts.

## Data Availability

Data that supports the findings of this study was provided by UK CPRD database. Availability of data is subject to protocol approval by CPRD's Research Data Governance Process.

## Acknowledgments

The authors acknowledge Mahkameh Mafi for project management; CPRD for their support; and Amgen and UCB Pharma for funding this study. A subsection of an earlier version of information reported in this manuscript was presented as a poster abstract (ID: P1129) by LE, SK, MA, ET, CL and, DPA at the WCO-IOF-ESCEO 2020 congress; available at https://doi.org/10.1007/s00198-020-05696-3.

## Authors’ roles

Study design: DPA, SK, MA, CL, ET. CPRD data management and advanced curation: AD. Data analysis: LE, SK. Data interpretation: MPM, LE, DPA, SK. Drafting manuscript: MPM, LE, CC, MA, CL, SK. Revising manuscript content: all authors. Approving final version of manuscript: all authors. SK takes responsibility for the integrity of the data analysis.

## Ethical approval

The Independent Scientific Advisory Committee of the Clinical Practice Research Datalink gave ethical approval for this work with protocol number 18_116R.

## Transparency Statement

The lead authors (MPM, LE and SK) affirm that the manuscript is an honest, accurate, and transparent account of the study being reported; that no important aspects of the study have been omitted; and that any discrepancies from the study as planned have been explained.

