## supplementary data for "Estimating the incidence and key risk factors of cardiovascular disease in patients at high risk of imminent fracture using routinely collected real-world data from the UK"

***Supplementary file A***

#### Details on the analytical steps

Multiple imputation with chain equations was applied to handle missing values for smoking, drinking, estimated Glomerular Filtration Rate (eGFR), body mass index (BMI), cholesterol, systolic blood pressure (SBP) and diastolic blood pressure (DBP) resulting in 20 imputed datasets. Each imputed dataset was then randomly split 50/50 into a development and a validation set. For managing the large amount of risk factors from the set of ALL variables, a lasso regression model was fitted to one imputed development set. The model selected those risk factors as features that resulted in the best area under the curve (AUC) using cross-validation within the development set. The final model was then estimated by applying logistic regression to all 20 imputed development sets using the lasso-selected features and adjusting the estimates and standard errors for the variability between the imputed datasets with Rubin’s rules

#### How to use the prediction models

To obtain the estimate risk of an event for an individual patient, the intercept term of the model will be sum to the value of the predictor variables multiplied by their respective coefficients (β):

$$“Y \approx \beta 0 + \beta 1X1 + \ldots + \beta nXn + \varepsilon”$$

*Where **Y** is the individual risk, **β0** is the intercept, **β1X1 +… + βnXn** are the coefficients (β) of the predictive variables (X), and **ε** the error that cannot be captured by the model.*

To transform the estimate into a probability, will need to use the formula of the logistic transformation:

**1/(1+exp(-(*estimate*))**

Example of how to use the proposed models:

We’ll going to predict the risk of 1-year MACE for a 72-year-old men who was newly diagnosed with osteoporosis. This patient is a current smoker, do not drink, has a prior CVD history and are using anti-hypertensive drugs. He has a BMI of 27.3, an eGFR value of 65, a SBP value of 141, and had 10 GP visits the prior year. He is using 5 concomitant medicines and had 1 prior fracture.

In this case, we are going to use 1-year MACE prediction for OST population, and the first step is to obtain the values of the coefficients for this patient.

Following table displays the coefficients from 1-year MACE model in OST population, and the specific patient coefficients:

| Predictor | OST<br>*Beta coefficients* | Patient characteristics<br>*(NO = 0)*<br>*(Yes = 1)* | Patient coefficients<br>*β0 and βn*Xn* |
|---|---|---|---|
| **Intercept (β0)** | -5.214 | | -5.214 |
| **Sex = Male (%)** | 0.479 | 1 | 0.479 |
| **Smoking**** | | | |
| **Ex** | ref | 0 | 0.000 |
| **No** | -0.070 | 0 | 0.000 |
| **Yes** | 0.353 | 1 | 0.353 |
| **Drinking**** | | | |
| **Ex** | ref | 0 | 0.000 |
| **No** | 0.171 | 1 | 0.171 |
| **Yes** | -0.170 | 0 | 0.000 |
| **Atrial fibrillation*** | 0.478 | 0 | 0.000 |
| **On anti-hypertensive drug** | 0.203 | 1 | 0.203 |
| **Age Group (%)** | | | |
| **50-59** | ref | 0 | 0.000 |
| **60-69** | 0.311 | 0 | 0.000 |
| **70-79** | 1.122 | 1 | 1.122 |
| **80-89** | 1.589 | 0 | 0.000 |
| **>89** | 2.065 | 0 | 0.000 |
| **MI or Stroke** | | | |
| **No** | ref | 0 | 0.000 |
| **Ever** | -0.009 | 0 | 0.000 |
| **1 year before index** | 0.709 | 0 | 0.000 |
| **Established CVD *** | 0.643 | 1 | 0.643 |
| **BMI**** | | | |
| **<18.5** | ref | 0 | 0.000 |
| **18.6 - 24.9** | -0.334 | 0 | 0.000 |
| **25 - 29.9** | -0.581 | 1 | -0.581 |
| **30 - 39.9** | -0.879 | 0 | 0.000 |
| **>=40** | -0.421 | 0 | 0.000 |
| **No. of GP visits**** | | | |
| **0** | ref | 0 | 0.000 |
| **1-5** | 0.009 | 0 | 0.000 |
| **6-10** | -0.063 | 1 | -0.063 |
| **11-15** | -0.056 | 0 | 0.000 |
| **>=16** | 0.064 | 0 | 0.000 |
| **eGFR**** | | | |
| **<=29** | ref | 0 | 0.000 |
| **30 – 44** | 0.101 | 0 | 0.000 |
| **45 – 59** | -0.112 | 0 | 0.000 |
| **60 – 89** | -0.280 | 1 | -0.280 |
| **>=90** | -0.236 | 0 | 0.000 |
| **SBP**** | | | |
| **<120** | ref | 0 | 0.000 |
| **120 - 139** | 0.175 | 0 | 0.000 |
| **140 - 159** | 0.278 | 1 | 0.278 |
| **>=160** | 0.274 | 0 | 0.000 |
| **No. of concomitant medicines**** | | | |
| **0** | ref | 0 | 0.000 |
| **1 – 3** | -0.377 | 0 | 0.000 |
| **4 – 6** | -0.183 | 1 | -0.183 |
| **7 – 9** | 0.008 | 0 | 0.000 |
| **10 – 12** | 0.111 | 0 | 0.000 |
| **>=13** | 0.105 | 0 | 0.000 |
| **No. of previous fractures*** | | | |
| **0** | ref | 0 | 0.000 |

| | | | |
|---|---|---|---|
| **1** | 0.217 | 1 | 0.217 |
| **>=2** | -0.008 | 0 | 0.000 |
| **Abbreviations**: OST, patients with incident diagnosis of osteoporosis; IFX, patients with incident fragility fracture; OBP, incident users of oral bisphosphonates; OR, odds ratio; CI, confidence intervals; MACE, composite outcome for the occurrence of either myocardial infarction, stroke or cardiovascular disease death; * ever; ** in the year prior to start; SES, socio-economic status; MI, myocardial infarction; BMI, body mass index; eGFR, estimated Glomerular Filtration Rate; SBP, cholesterol, systolic blood pressure; DBP, diastolic blood pressure. | | | |

Once knowing the specific patient coefficients, we are going to sum them (i.e., all values from "Patient coefficients *β0 and βn*Xn"* column) in order to obtain the patient's estimate. The result is -2.855.

Then, the estimate is going to be transformed into a probability:

**1/(1+exp(-(*estimate*)) =** 1/(1+exp(-(-2.855))) = 0.054423

Thus, the proposed patient has a 5.4% risk of having a MACE event in the following year after his risk assessment.

***Supplementary tables***

#### Table S1 List of available variables included in QRISK tool

| Socio-demographic | Lab measurements | Comorbidities | Cardiovascular disease history | Drug use |
|---|---|---|---|---|
| • Sex<br>• Ethnicity<br>• Age<br>• BMI**<br>• Smoking**<br>• **Deprivation (Town score)** | • SBP**<br>• Standardised SBP**<br>• Cholesterol measurement** | • Rheumatoid arthritis*<br>• Lupus*<br>• Severe mental illness*<br>• Diabetes type I*<br>• Diabetes type II*<br>• Chronic kidney disease*<br>• Migraine*<br>• Erectile dysfunction** | • Atrial fibrillation*<br>• Family history of cardiovascular disease before age 60 | • On bisphosphonate use<br>• Steroid use**<br>• Antipsychotic use** |

* Ever. ** In the year prior to start. In **bolt**, unavailable in CPRD. **Abbreviations**: BMI, body mass index; SPB, systolic blood pressure.

#### Table S2a Dataset split into train and test sets, stratified by outcome (OST cohort)

| OST cohort | One year | | | | | | | | Two years | | | | | | | |
|---|---|---|---|---|---|---|---|---|---|---|---|---|---|---|---|---|
| | MACE | | | | Stroke/MI | | | | MACE | | | | Stroke/MI | | | |
| | Development set (n=32648) | | Internal validation set (n=32647) | | Development set (n=32648) | | Internal validation set (n=32647) | | Development set (n=32648) | | Internal validation set (n=32647) | | Development set (n=32648) | | Internal validation set (n=32647) | |
| | Outcome=NO | Outcome=YES | Outcome=NO | Outcome=YES | Outcome=NO | Outcome=YES | Outcome=NO | Outcome=YES | Outcome=NO | Outcome=YES | Outcome=NO | Outcome=YES | Outcome=NO | Outcome=YES | Outcome=NO | Outcome=YES |
| n | 32029 | 619 | 32028 | 619 | 32135 | 513 | 32134 | 513 | 31492 | 1156 | 31492 | 1155 | 31701 | 947 | 31700 | 947 |
| Sex = Male (%) | 4186 (13.1) | 126 (20.4) | 4179 (13.0) | 125 (20.2) | 4283 (13.3) | 105 (20.5) | 4132 (12.9) | 96 (18.7) | 4051 (12.9) | 211 (18.3) | 4116 (13.1) | 238 (20.6) | 4235 (13.4) | 173 (18.3) | 4037 (12.7) | 171 (18.1) |
| SES (%) | | | | | | | | | | | | | | | | |
| 1 | 7904 (24.7) | 137 (22.1) | 7787 (24.3) | 125 (20.2) | 7833 (24.4) | 108 (21.1) | 7914 (24.6) | 98 (19.1) | 7743 (24.6) | 250 (21.6) | 7717 (24.5) | 243 (21.0) | 7783 (24.6) | 199 (21.0) | 7773 (24.5) | 198 (20.9) |
| 2 | 7655 (23.9) | 142 (22.9) | 7691 (24.0) | 155 (25.0) | 7645 (23.8) | 130 (25.3) | 7745 (24.1) | 123 (24.0) | 7590 (24.1) | 288 (24.9) | 7515 (23.9) | 250 (21.6) | 7662 (24.2) | 221 (23.3) | 7539 (23.8) | 221 (23.3) |
| 3 | 6749 (21.1) | 133 (21.5) | 6775 (21.2) | 137 (22.1) | 6814 (21.2) | 104 (20.3) | 6765 (21.1) | 111 (21.6) | 6615 (21.0) | 260 (22.5) | 6658 (21.1) | 261 (22.6) | 6628 (20.9) | 215 (22.7) | 6761 (21.3) | 190 (20.1) |
| 4 | 5855 (18.3) | 118 (19.1) | 5848 (18.3) | 100 (16.2) | 5889 (18.3) | 89 (17.3) | 5843 (18.2) | 100 (19.5) | 5766 (18.3) | 197 (17.0) | 5745 (18.2) | 213 (18.4) | 5802 (18.3) | 172 (18.2) | 5765 (18.2) | 182 (19.2) |
| 5 | 3827 (11.9) | 89 (14.4) | 3902 (12.2) | 100 (16.2) | 3929 (12.2) | 82 (16.0) | 3827 (11.9) | 80 (15.6) | 3738 (11.9) | 160 (13.8) | 3834 (12.2) | 186 (16.1) | 3797 (12.0) | 139 (14.7) | 3827 (12.1) | 155 (16.4) |
| Smoking** (%) | | | | | | | | | | | | | | | | |
| Ex | 10037 (31.3) | 199 (32.1) | 10062 (31.4) | 189 (30.5) | 10081 (31.4) | 169 (32.9) | 10075 (31.4) | 162 (31.6) | 9889 (31.4) | 385 (33.3) | 9839 (31.2) | 374 (32.4) | 9941 (31.4) | 290 (30.6) | 9929 (31.3) | 327 (34.5) |
| No | 16758 (52.3) | 314 (50.7) | 16661 (52.0) | 316 (51.1) | 16750 (52.1) | 259 (50.5) | 16771 (52.2) | 269 (52.4) | 16418 (52.1) | 597 (51.6) | 16447 (52.2) | 587 (50.8) | 16531 (52.1) | 515 (54.4) | 16538 (52.2) | 465 (49.1) |
| Yes | 5234 (16.3) | 106 (17.1) | 5305 (16.6) | 114 (18.4) | 5304 (16.5) | 85 (16.6) | 5288 (16.5) | 82 (16.0) | 5185 (16.5) | 174 (15.1) | 5206 (16.5) | 194 (16.8) | 5229 (16.5) | 142 (15.0) | 5233 (16.5) | 155 (16.4) |
| Drinking** (%) | | | | | | | | | | | | | | | | |
| Ex | 1702 (5.3) | 53 (8.6) | 1756 (5.5) | 53 (8.6) | 1741 (5.4) | 47 (9.2) | 1741 (5.4) | 35 (6.8) | 1708 (5.4) | 92 (8.0) | 1677 (5.3) | 87 (7.5) | 1699 (5.4) | 71 (7.5) | 1729 (5.5) | 65 (6.9) |
| No | 8244 (25.7) | 225 (36.3) | 8388 (26.2) | 205 (33.1) | 8364 (26.0) | 178 (34.7) | 8340 (26.0) | 180 (35.1) | 8175 (26.0) | 384 (33.2) | 8121 (25.8) | 382 (33.1) | 8262 (26.1) | 317 (33.5) | 8170 (25.8) | 313 (33.1) |
| Yes | 22083 (68.9) | 341 (55.1) | 21884 (68.3) | 361 (58.3) | 22030 (68.6) | 288 (56.1) | 22053 (68.6) | 298 (58.1) | 21609 (68.6) | 680 (58.8) | 21694 (68.9) | 686 (59.4) | 21740 (68.6) | 559 (59.0) | 21801 (68.8) | 569 (60.1) |
| Diabetes type I*= 1 (%) | 62 (0.2) | 2 (0.3) | 82 (0.3) | 6 (1.0) | 81 (0.3) | 2 (0.4) | 64 (0.2) | 5 (1.0) | 80 (0.3) | 3 (0.3) | 61 (0.2) | 8 (0.7) | 80 (0.3) | 4 (0.4) | 63 (0.2) | 5 (0.5) |

| OST cohort | One year | | | | | | | | Two years | | | | | | | |
|---|---|---|---|---|---|---|---|---|---|---|---|---|---|---|---|---|
| | MACE | | | | Stroke/MI | | | | MACE | | | | Stroke/MI | | | |
| | Development set (n=32648) | | Internal validation set (n=32647) | | Development set (n=32648) | | Internal validation set (n=32647) | | Development set (n=32648) | | Internal validation set (n=32647) | | Development set (n=32648) | | Internal validation set (n=32647) | |
| | Outcome=NO | Outcome=YES | Outcome=NO | Outcome=YES | Outcome=NO | Outcome=YES | Outcome=NO | Outcome=YES | Outcome=NO | Outcome=YES | Outcome=NO | Outcome=YES | Outcome=NO | Outcome=YES | Outcome=NO | Outcome=YES |
| Diabetes type II*= 1 (%) | 1632 (5.1) | 50 (8.1) | 1580 (4.9) | 49 (7.9) | 1634 (5.1) | 37 (7.2) | 1598 (5.0) | 42 (8.2) | 1554 (4.9) | 95 (8.2) | 1575 (5.0) | 87 (7.5) | 1583 (5.0) | 73 (7.7) | 1571 (5.0) | 84 (8.9) |
| Chronic obstructive pulmonary disease*= 1 (%) | 2332 (7.3) | 54 (8.7) | 2488 (7.8) | 65 (10.5) | 2477 (7.7) | 52 (10.1) | 2364 (7.4) | 46 (9.0) | 2368 (7.5) | 106 (9.2) | 2338 (7.4) | 127 (11.0) | 2338 (7.4) | 83 (8.8) | 2406 (7.6) | 112 (11.8) |
| Chronic kidney disease*= 1 (%) | 2781 (8.7) | 106 (17.1) | 2823 (8.8) | 80 (12.9) | 2810 (8.7) | 72 (14.0) | 2822 (8.8) | 86 (16.8) | 2791 (8.9) | 171 (14.8) | 2651 (8.4) | 177 (15.3) | 2738 (8.6) | 147 (15.5) | 2761 (8.7) | 144 (15.2) |
| Rheumatoid arthritis*= 1 (%) | 2424 (7.6) | 59 (9.5) | 2495 (7.8) | 67 (10.8) | 2452 (7.6) | 53 (10.3) | 2481 (7.7) | 59 (11.5) | 2452 (7.8) | 112 (9.7) | 2371 (7.5) | 110 (9.5) | 2412 (7.6) | 95 (10.0) | 2442 (7.7) | 96 (10.1) |
| Lupus*= 1 (%) | 69 (0.2) | 2 (0.3) | 73 (0.2) | 0 (0.0) | 65 (0.2) | 1 (0.2) | 78 (0.2) | 0 (0.0) | 77 (0.2) | 1 (0.1) | 65 (0.2) | 1 (0.1) | 73 (0.2) | 1 (0.1) | 68 (0.2) | 2 (0.2) |
| Systemic heart disease**= 1 (%) | 1154 (3.6) | 10 (1.6) | 1168 (3.6) | 9 (1.5) | 1146 (3.6) | 9 (1.8) | 1178 (3.7) | 8 (1.6) | 1192 (3.8) | 18 (1.6) | 1115 (3.5) | 16 (1.4) | 1110 (3.5) | 16 (1.7) | 1198 (3.8) | 17 (1.8) |
| Anti-osteoporosis use**= 1 (%) | 5671 (17.7) | 125 (20.2) | 5825 (18.2) | 131 (21.2) | 5766 (17.9) | 95 (18.5) | 5781 (18.0) | 110 (21.4) | 5633 (17.9) | 225 (19.5) | 5645 (17.9) | 249 (21.6) | 5715 (18.0) | 202 (21.3) | 5658 (17.8) | 177 (18.7) |
| Heparin use**= 1 (%) | 163 (0.5) | 3 (0.5) | 198 (0.6) | 8 (1.3) | 195 (0.6) | 3 (0.6) | 166 (0.5) | 8 (1.6) | 168 (0.5) | 11 (1.0) | 181 (0.6) | 12 (1.0) | 178 (0.6) | 13 (1.4) | 173 (0.5) | 8 (0.8) |
| Beta-blocker use**= 1 (%) | 5256 (16.4) | 154 (24.9) | 5016 (15.7) | 138 (22.3) | 5119 (15.9) | 123 (24.0) | 5180 (16.1) | 142 (27.7) | 5045 (16.0) | 286 (24.7) | 4966 (15.8) | 267 (23.1) | 5021 (15.8) | 262 (27.7) | 5049 (15.9) | 232 (24.5) |
| Hypertension**= 1 (%) | 2504 (7.8) | 69 (11.1) | 2464 (7.7) | 54 (8.7) | 2493 (7.8) | 60 (11.7) | 2485 (7.7) | 53 (10.3) | 2415 (7.7) | 105 (9.1) | 2443 (7.8) | 128 (11.1) | 2466 (7.8) | 97 (10.2) | 2428 (7.7) | 100 (10.6) |
| Deep vein thrombosis or pulmonary embolism**= 1 (%) | 257 (0.8) | 6 (1.0) | 244 (0.8) | 15 (2.4) | 267 (0.8) | 10 (1.9) | 238 (0.7) | 7 (1.4) | 249 (0.8) | 12 (1.0) | 245 (0.8) | 16 (1.4) | 244 (0.8) | 12 (1.3) | 254 (0.8) | 12 (1.3) |
| Anticoagulant use**= 1 (%) | 1578 (4.9) | 52 (8.4) | 1602 (5.0) | 53 (8.6) | 1609 (5.0) | 31 (6.0) | 1597 (5.0) | 48 (9.4) | 1576 (5.0) | 95 (8.2) | 1520 (4.8) | 94 (8.1) | 1554 (4.9) | 73 (7.7) | 1581 (5.0) | 77 (8.1) |
| Antidepressants TCA**= 1 (%) | 3418 (10.7) | 82 (13.2) | 3468 (10.8) | 71 (11.5) | 3419 (10.6) | 69 (13.5) | 3482 (10.8) | 69 (13.5) | 3361 (10.7) | 135 (11.7) | 3406 (10.8) | 137 (11.9) | 3400 (10.7) | 109 (11.5) | 3402 (10.7) | 128 (13.5) |
| Antidepressants SSRI**= 1 (%) | 2900 (9.1) | 84 (13.6) | 2928 (9.1) | 78 (12.6) | 2920 (9.1) | 68 (13.3) | 2942 (9.2) | 60 (11.7) | 2886 (9.2) | 146 (12.6) | 2808 (8.9) | 150 (13.0) | 2855 (9.0) | 116 (12.2) | 2903 (9.2) | 116 (12.2) |
| Hypercholesterolemia**= 1 (%) | 793 (2.5) | 16 (2.6) | 740 (2.3) | 12 (1.9) | 740 (2.3) | 16 (3.1) | 797 (2.5) | 8 (1.6) | 733 (2.3) | 28 (2.4) | 776 (2.5) | 24 (2.1) | 767 (2.4) | 13 (1.4) | 750 (2.4) | 31 (3.3) |
| Statin use**= 1 (%) | 7643 (23.9) | 196 (31.7) | 7542 (23.5) | 190 (30.7) | 7634 (23.8) | 165 (32.2) | 7602 (23.7) | 170 (33.1) | 7419 (23.6) | 347 (30.0) | 7434 (23.6) | 371 (32.1) | 7557 (23.8) | 294 (31.0) | 7402 (23.4) | 318 (33.6) |

| OST cohort | One year | | | | | | | | Two years | | | | | | | |
|---|---|---|---|---|---|---|---|---|---|---|---|---|---|---|---|---|
| | MACE | | | | Stroke/MI | | | | MACE | | | | Stroke/MI | | | |
| | **Development set (n=32648)** | | **Internal validation set (n=32647)** | | **Development set (n=32648)** | | **Internal validation set (n=32647)** | | **Development set (n=32648)** | | **Internal validation set (n=32647)** | | **Development set (n=32648)** | | **Internal validation set (n=32647)** | |
| | **Outcome=NO** | **Outcome=YES** | **Outcome=NO** | **Outcome=YES** | **Outcome=NO** | **Outcome=YES** | **Outcome=NO** | **Outcome=YES** | **Outcome=NO** | **Outcome=YES** | **Outcome=NO** | **Outcome=YES** | **Outcome=NO** | **Outcome=YES** | **Outcome=NO** | **Outcome=YES** |
| Family history of cardiovascular disease = 1 (%) | 3310 (10.3) | 55 (8.9) | 3343 (10.4) | 47 (7.6) | 3316 (10.3) | 42 (8.2) | 3343 (10.4) | 54 (10.5) | 3329 (10.6) | 94 (8.1) | 3234 (10.3) | 98 (8.5) | 3358 (10.6) | 87 (9.2) | 3213 (10.1) | 97 (10.2) |
| Family history of cardiovascular disease before age 60= 1 (%) | 46 (0.1) | 1 (0.2) | 52 (0.2) | 0 (0.0) | 59 (0.2) | 1 (0.2) | 39 (0.1) | 0 (0.0) | 48 (0.2) | 0 (0.0) | 50 (0.2) | 1 (0.1) | 54 (0.2) | 0 (0.0) | 44 (0.1) | 1 (0.1) |
| Heart failure*= 1 (%) | 1089 (3.4) | 41 (6.6) | 1127 (3.5) | 66 (10.7) | 1116 (3.5) | 45 (8.8) | 1118 (3.5) | 44 (8.6) | 1107 (3.5) | 95 (8.2) | 1030 (3.3) | 91 (7.9) | 1051 (3.3) | 75 (7.9) | 1124 (3.5) | 73 (7.7) |
| Migraine*= 1 (%) | 5040 (15.7) | 91 (14.7) | 4878 (15.2) | 94 (15.2) | 4921 (15.3) | 80 (15.6) | 5011 (15.6) | 91 (17.7) | 4862 (15.4) | 164 (14.2) | 4882 (15.5) | 195 (16.9) | 4803 (15.2) | 166 (17.5) | 4979 (15.7) | 155 (16.4) |
| Severe mental illness*= 1 (%) | 4890 (15.3) | 104 (16.8) | 5110 (16.0) | 99 (16.0) | 4940 (15.4) | 79 (15.4) | 5093 (15.8) | 91 (17.7) | 4983 (15.8) | 187 (16.2) | 4827 (15.3) | 206 (17.8) | 4931 (15.6) | 165 (17.4) | 4945 (15.6) | 162 (17.1) |
| Vascular Disease*= 1 (%) | 363 (1.1) | 16 (2.6) | 437 (1.4) | 23 (3.7) | 413 (1.3) | 19 (3.7) | 389 (1.2) | 18 (3.5) | 393 (1.2) | 39 (3.4) | 376 (1.2) | 31 (2.7) | 390 (1.2) | 32 (3.4) | 380 (1.2) | 37 (3.9) |
| Atrial fibrillation*= 1 (%) | 1788 (5.6) | 85 (13.7) | 1708 (5.3) | 83 (13.4) | 1764 (5.5) | 64 (12.5) | 1766 (5.5) | 70 (13.6) | 1707 (5.4) | 134 (11.6) | 1676 (5.3) | 147 (12.7) | 1710 (5.4) | 119 (12.6) | 1730 (5.5) | 105 (11.1) |
| On anti-hypertensive drug= 1 (%) | 18216 (56.9) | 464 (75.0) | 17934 (56.0) | 439 (70.9) | 18154 (56.5) | 381 (74.3) | 18131 (56.4) | 387 (75.4) | 17812 (56.6) | 829 (71.7) | 17545 (55.7) | 867 (75.1) | 17790 (56.1) | 730 (77.1) | 17847 (56.3) | 686 (72.4) |
| Antipsychotic use**= 1 (%) | 167 (0.5) | 4 (0.6) | 197 (0.6) | 3 (0.5) | 183 (0.6) | 2 (0.4) | 184 (0.6) | 2 (0.4) | 174 (0.6) | 9 (0.8) | 181 (0.6) | 7 (0.6) | 174 (0.5) | 6 (0.6) | 187 (0.6) | 4 (0.4) |
| Steroid use**= 1 (%) | 4494 (14.0) | 111 (17.9) | 4652 (14.5) | 110 (17.8) | 4624 (14.4) | 86 (16.8) | 4564 (14.2) | 93 (18.1) | 4534 (14.4) | 190 (16.4) | 4417 (14.0) | 226 (19.6) | 4457 (14.1) | 160 (16.9) | 4568 (14.4) | 182 (19.2) |
| Erectile dysfunction**= 1 (%) | 413 (1.3) | 11 (1.8) | 434 (1.4) | 13 (2.1) | 407 (1.3) | 7 (1.4) | 442 (1.4) | 15 (2.9) | 428 (1.4) | 20 (1.7) | 397 (1.3) | 26 (2.3) | 426 (1.3) | 13 (1.4) | 406 (1.3) | 26 (2.7) |
| Age>75 | 15094 (47.1) | 502 (81.1) | 15030 (46.9) | 479 (77.4) | 15174 (47.2) | 396 (77.2) | 15136 (47.1) | 399 (77.8) | 14674 (46.6) | 896 (77.5) | 14632 (46.5) | 903 (78.2) | 14786 (46.6) | 740 (78.1) | 14879 (46.9) | 700 (73.9) |
| Age Group (%) | | | | | | | | | | | | | | | | |
| 50-59 | 4143 (12.9) | 18 (2.9) | 4133 (12.9) | 21 (3.4) | 4167 (13.0) | 22 (4.3) | 4112 (12.8) | 14 (2.7) | 4131 (13.1) | 34 (2.9) | 4120 (13.1) | 30 (2.6) | 4095 (12.9) | 19 (2.0) | 4161 (13.1) | 40 (4.2) |
| 60-69 | 7821 (24.4) | 51 (8.2) | 8015 (25.0) | 62 (10.0) | 7906 (24.6) | 43 (8.4) | 7939 (24.7) | 61 (11.9) | 7797 (24.8) | 109 (9.4) | 7926 (25.2) | 117 (10.1) | 7892 (24.9) | 97 (10.2) | 7851 (24.8) | 109 (11.5) |
| 70-79 | 10501 (32.8) | 172 (27.8) | 10315 (32.2) | 177 (28.6) | 10412 (32.4) | 150 (29.2) | 10455 (32.5) | 148 (28.8) | 10215 (32.4) | 351 (30.4) | 10265 (32.6) | 334 (28.9) | 10396 (32.8) | 297 (31.4) | 10181 (32.1) | 291 (30.7) |

| OST cohort | One year | | | | | | | | Two years | | | | | | | |
|---|---|---|---|---|---|---|---|---|---|---|---|---|---|---|---|---|
| | MACE | | | | Stroke/MI | | | | MACE | | | | Stroke/MI | | | |
| | Development set (n=32648) | | Internal validation set (n=32647) | | Development set (n=32648) | | Internal validation set (n=32647) | | Development set (n=32648) | | Internal validation set (n=32647) | | Development set (n=32648) | | Internal validation set (n=32647) | |
| | Outcome=NO | Outcome=YES | Outcome=NO | Outcome=YES | Outcome=NO | Outcome=YES | Outcome=NO | Outcome=YES | Outcome=NO | Outcome=YES | Outcome=NO | Outcome=YES | Outcome=NO | Outcome=YES | Outcome=NO | Outcome=YES |
| 80-89 | 8028 (25.1) | 298 (48.1) | 7995 (25.0) | 265 (42.8) | 8094 (25.2) | 223 (43.5) | 8037 (25.0) | 232 (45.2) | 7856 (24.9) | 501 (43.3) | 7706 (24.5) | 523 (45.3) | 7798 (24.6) | 418 (44.1) | 7961 (25.1) | 409 (43.2) |
| >89 | 1536 (4.8) | 80 (12.9) | 1570 (4.9) | 94 (15.2) | 1556 (4.8) | 75 (14.6) | 1591 (5.0) | 58 (11.3) | 1493 (4.7) | 161 (13.9) | 1475 (4.7) | 151 (13.1) | 1520 (4.8) | 116 (12.2) | 1546 (4.9) | 98 (10.3) |
| Charlson score (%) | | | | | | | | | | | | | | | | |
| 0 | 18763 (58.6) | 280 (45.2) | 18477 (57.7) | 262 (42.3) | 18732 (58.3) | 226 (44.1) | 18603 (57.9) | 221 (43.1) | 18315 (58.2) | 508 (43.9) | 18443 (58.6) | 516 (44.7) | 18495 (58.3) | 417 (44.0) | 18455 (58.2) | 415 (43.8) |
| 1 | 6472 (20.2) | 143 (23.1) | 6496 (20.3) | 163 (26.3) | 6454 (20.1) | 131 (25.5) | 6568 (20.4) | 121 (23.6) | 6349 (20.2) | 291 (25.2) | 6353 (20.2) | 281 (24.3) | 6356 (20.0) | 225 (23.8) | 6447 (20.3) | 246 (26.0) |
| 2 | 3794 (11.8) | 83 (13.4) | 3995 (12.5) | 88 (14.2) | 3871 (12.0) | 67 (13.1) | 3951 (12.3) | 71 (13.8) | 3830 (12.2) | 172 (14.9) | 3813 (12.1) | 145 (12.6) | 3865 (12.2) | 140 (14.8) | 3842 (12.1) | 113 (11.9) |
| ≥3 | 3000 (9.4) | 113 (18.3) | 3060 (9.6) | 106 (17.1) | 3078 (9.6) | 89 (17.3) | 3012 (9.4) | 100 (19.5) | 2998 (9.5) | 185 (16.0) | 2883 (9.2) | 213 (18.4) | 2985 (9.4) | 165 (17.4) | 2956 (9.3) | 173 (18.3) |
| Cardiovascular disease (%) | | | | | | | | | | | | | | | | |
| No | 28476 (88.9) | 488 (78.8) | 28594 (89.3) | 476 (76.9) | 28578 (88.9) | 389 (75.8) | 28675 (89.2) | 392 (76.4) | 28079 (89.2) | 911 (78.8) | 28138 (89.3) | 906 (78.4) | 28290 (89.2) | 735 (77.6) | 28283 (89.2) | 726 (76.7) |
| Ever >1 year before index date | 2743 (8.6) | 90 (14.5) | 2687 (8.4) | 96 (15.5) | 2755 (8.6) | 83 (16.2) | 2690 (8.4) | 88 (17.2) | 2638 (8.4) | 170 (14.7) | 2628 (8.3) | 180 (15.6) | 2639 (8.3) | 145 (15.3) | 2664 (8.4) | 168 (17.7) |
| 1 year before index | 358 (1.1) | 15 (2.4) | 326 (1.0) | 20 (3.2) | 328 (1.0) | 14 (2.7) | 363 (1.1) | 14 (2.7) | 358 (1.1) | 30 (2.6) | 306 (1.0) | 25 (2.2) | 341 (1.1) | 19 (2.0) | 334 (1.1) | 25 (2.6) |
| 6 months before index | 360 (1.1) | 17 (2.7) | 322 (1.0) | 24 (3.9) | 361 (1.1) | 22 (4.3) | 324 (1.0) | 16 (3.1) | 331 (1.1) | 30 (2.6) | 324 (1.0) | 38 (3.3) | 342 (1.1) | 37 (3.9) | 319 (1.0) | 25 (2.6) |
| 1 month before index | 92 (0.3) | 9 (1.5) | 99 (0.3) | 3 (0.5) | 113 (0.4) | 5 (1.0) | 82 (0.3) | 3 (0.6) | 86 (0.3) | 15 (1.3) | 96 (0.3) | 6 (0.5) | 89 (0.3) | 11 (1.2) | 100 (0.3) | 3 (0.3) |
| MI or Stroke (%) | | | | | | | | | | | | | | | | |
| No | 30118 (94.0) | 499 (80.6) | 30236 (94.4) | 497 (80.3) | 30230 (94.1) | 409 (79.7) | 30279 (94.2) | 432 (84.2) | 29774 (94.5) | 970 (83.9) | 29670 (94.2) | 936 (81.0) | 29899 (94.3) | 798 (84.3) | 29863 (94.2) | 790 (83.4) |
| Ever >1 year before index date | 1380 (4.3) | 69 (11.1) | 1325 (4.1) | 76 (12.3) | 1373 (4.3) | 65 (12.7) | 1362 (4.2) | 50 (9.7) | 1245 (4.0) | 111 (9.6) | 1350 (4.3) | 144 (12.5) | 1308 (4.1) | 97 (10.2) | 1341 (4.2) | 104 (11.0) |
| 1 year before index | 531 (1.7) | 51 (8.2) | 467 (1.5) | 46 (7.4) | 532 (1.7) | 39 (7.6) | 493 (1.5) | 31 (6.0) | 473 (1.5) | 75 (6.5) | 472 (1.5) | 75 (6.5) | 494 (1.6) | 52 (5.5) | 496 (1.6) | 53 (5.6) |
| Established CVD *= Ever (%) | 3557 (11.1) | 187 (30.2) | 3495 (10.9) | 189 (30.5) | 3573 (11.1) | 161 (31.4) | 3545 (11.0) | 149 (29.0) | 3373 (10.7) | 314 (27.2) | 3404 (10.8) | 337 (29.2) | 3474 (11.0) | 272 (28.7) | 3423 (10.8) | 259 (27.3) |

| OST cohort | One year | | | | | | | | Two years | | | | | | | |
|---|---|---|---|---|---|---|---|---|---|---|---|---|---|---|---|---|
| | MACE | | | | Stroke/MI | | | | MACE | | | | Stroke/MI | | | |
| | Development set (n=32648) | | Internal validation set (n=32647) | | Development set (n=32648) | | Internal validation set (n=32647) | | Development set (n=32648) | | Internal validation set (n=32647) | | Development set (n=32648) | | Internal validation set (n=32647) | |
| | Outcome=NO | Outcome=YES | Outcome=NO | Outcome=YES | Outcome=NO | Outcome=YES | Outcome=NO | Outcome=YES | Outcome=NO | Outcome=YES | Outcome=NO | Outcome=YES | Outcome=NO | Outcome=YES | Outcome=NO | Outcome=YES |
| Any fracture history (%) | | | | | | | | | | | | | | | | |
| No | 24236 (75.7) | 449 (72.5) | 24408 (76.2) | 449 (72.5) | 24318 (75.7) | 369 (71.9) | 24490 (76.2) | 365 (71.2) | 23910 (75.9) | 867 (75.0) | 23930 (76.0) | 835 (72.3) | 24073 (75.9) | 690 (72.9) | 24089 (76.0) | 690 (72.9) |
| Ever >1 year before index date | 2899 (9.1) | 71 (11.5) | 2788 (8.7) | 55 (8.9) | 2934 (9.1) | 58 (11.3) | 2762 (8.6) | 59 (11.5) | 2780 (8.8) | 102 (8.8) | 2802 (8.9) | 129 (11.2) | 2821 (8.9) | 101 (10.7) | 2788 (8.8) | 103 (10.9) |
| 1 year before index | 4894 (15.3) | 99 (16.0) | 4832 (15.1) | 115 (18.6) | 4883 (15.2) | 86 (16.8) | 4882 (15.2) | 89 (17.3) | 4802 (15.2) | 187 (16.2) | 4760 (15.1) | 191 (16.5) | 4807 (15.2) | 156 (16.5) | 4823 (15.2) | 154 (16.3) |
| Hip fracture history (%) | | | | | | | | | | | | | | | | |
| No | 30453 (95.1) | 575 (92.9) | 30443 (95.1) | 569 (91.9) | 30577 (95.2) | 473 (92.2) | 30512 (95.0) | 478 (93.2) | 29921 (95.0) | 1080 (93.4) | 29978 (95.2) | 1061 (91.9) | 30121 (95.0) | 891 (94.1) | 30161 (95.1) | 867 (91.6) |
| Ever >1 year before index date | 541 (1.7) | 15 (2.4) | 541 (1.7) | 21 (3.4) | 554 (1.7) | 18 (3.5) | 535 (1.7) | 11 (2.1) | 548 (1.7) | 30 (2.6) | 500 (1.6) | 40 (3.5) | 553 (1.7) | 23 (2.4) | 511 (1.6) | 31 (3.3) |
| 1 year before index | 1035 (3.2) | 29 (4.7) | 1044 (3.3) | 29 (4.7) | 1004 (3.1) | 22 (4.3) | 1087 (3.4) | 24 (4.7) | 1023 (3.2) | 46 (4.0) | 1014 (3.2) | 54 (4.7) | 1027 (3.2) | 33 (3.5) | 1028 (3.2) | 49 (5.2) |
| Shoulder fracture history (%) | | | | | | | | | | | | | | | | |
| No | 31803 (99.3) | 616 (99.5) | 31805 (99.3) | 612 (98.9) | 31909 (99.3) | 511 (99.6) | 31910 (99.3) | 506 (98.6) | 31265 (99.3) | 1149 (99.4) | 31276 (99.3) | 1146 (99.2) | 31478 (99.3) | 938 (99.0) | 31479 (99.3) | 941 (99.4) |
| Ever >1 year before index date | 124 (0.4) | 0 (0.0) | 107 (0.3) | 1 (0.2) | 115 (0.4) | 0 (0.0) | 117 (0.4) | 0 (0.0) | 120 (0.4) | 0 (0.0) | 109 (0.3) | 3 (0.3) | 115 (0.4) | 2 (0.2) | 115 (0.4) | 0 (0.0) |
| 1 year before index | 102 (0.3) | 3 (0.5) | 116 (0.4) | 6 (1.0) | 111 (0.3) | 2 (0.4) | 107 (0.3) | 7 (1.4) | 107 (0.3) | 7 (0.6) | 107 (0.3) | 6 (0.5) | 108 (0.3) | 7 (0.7) | 106 (0.3) | 6 (0.6) |
| Spine fracture history (%) | | | | | | | | | | | | | | | | |
| No | 31352 (97.9) | 601 (97.1) | 31328 (97.8) | 599 (96.8) | 31434 (97.8) | 498 (97.1) | 31454 (97.9) | 494 (96.3) | 30827 (97.9) | 1129 (97.7) | 30815 (97.9) | 1109 (96.0) | 31009 (97.8) | 908 (95.9) | 31039 (97.9) | 924 (97.6) |
| Ever >1 year before index date | 159 (0.5) | 7 (1.1) | 156 (0.5) | 4 (0.6) | 165 (0.5) | 5 (1.0) | 152 (0.5) | 4 (0.8) | 154 (0.5) | 7 (0.6) | 152 (0.5) | 13 (1.1) | 156 (0.5) | 10 (1.1) | 154 (0.5) | 6 (0.6) |
| 1 year before index | 518 (1.6) | 11 (1.8) | 544 (1.7) | 16 (2.6) | 536 (1.7) | 10 (1.9) | 528 (1.6) | 15 (2.9) | 511 (1.6) | 20 (1.7) | 525 (1.7) | 33 (2.9) | 536 (1.7) | 29 (3.1) | 507 (1.6) | 17 (1.8) |
| Wrist fracture history (%) | | | | | | | | | | | | | | | | |
| No | 29468 (92.0) | 579 (93.5) | 29620 (92.5) | 570 (92.1) | 29609 (92.1) | 471 (91.8) | 29680 (92.4) | 477 (93.0) | 29021 (92.2) | 1077 (93.2) | 29068 (92.3) | 1071 (92.7) | 29209 (92.1) | 870 (91.9) | 29277 (92.4) | 881 (93.0) |
| Ever >1 year before index date | 1183 (3.7) | 21 (3.4) | 1139 (3.6) | 20 (3.2) | 1216 (3.8) | 19 (3.7) | 1112 (3.5) | 16 (3.1) | 1132 (3.6) | 36 (3.1) | 1155 (3.7) | 40 (3.5) | 1183 (3.7) | 38 (4.0) | 1112 (3.5) | 30 (3.2) |

| OST cohort | One year | | | | | | | | Two years | | | | | | | |
|---|---|---|---|---|---|---|---|---|---|---|---|---|---|---|---|---|
| | MACE | | | | Stroke/MI | | | | MACE | | | | Stroke/MI | | | |
| | Development set (n=32648) | | Internal validation set (n=32647) | | Development set (n=32648) | | Internal validation set (n=32647) | | Development set (n=32648) | | Internal validation set (n=32647) | | Development set (n=32648) | | Internal validation set (n=32647) | |
| | Outcome=NO | Outcome=YES | Outcome=NO | Outcome=YES | Outcome=NO | Outcome=YES | Outcome=NO | Outcome=YES | Outcome=NO | Outcome=YES | Outcome=NO | Outcome=YES | Outcome=NO | Outcome=YES | Outcome=NO | Outcome=YES |
| 1 year before index | 1378 (4.3) | 19 (3.1) | 1269 (4.0) | 29 (4.7) | 1310 (4.1) | 23 (4.5) | 1342 (4.2) | 20 (3.9) | 1339 (4.3) | 43 (3.7) | 1269 (4.0) | 44 (3.8) | 1309 (4.1) | 39 (4.1) | 1311 (4.1) | 36 (3.8) |
| BMI** (%) | | | | | | | | | | | | | | | | |
| <18.5 | 2754 (8.6) | 93 (15.0) | 2691 (8.4) | 77 (12.4) | 2693 (8.4) | 63 (12.3) | 2796 (8.7) | 63 (12.3) | 2637 (8.4) | 156 (13.5) | 2672 (8.5) | 150 (13.0) | 2700 (8.5) | 121 (12.8) | 2681 (8.5) | 113 (11.9) |
| 18.6 - 24.9 | 16003 (50.0) | 310 (50.1) | 15903 (49.7) | 348 (56.2) | 15960 (49.7) | 277 (54.0) | 16069 (50.0) | 258 (50.3) | 15780 (50.1) | 608 (52.6) | 15575 (49.5) | 601 (52.0) | 15815 (49.9) | 482 (50.9) | 15786 (49.8) | 481 (50.8) |
| 25 - 29.9 | 8949 (27.9) | 153 (24.7) | 9110 (28.4) | 131 (21.2) | 9131 (28.4) | 128 (25.0) | 8963 (27.9) | 121 (23.6) | 8826 (28.0) | 269 (23.3) | 8964 (28.5) | 284 (24.6) | 8950 (28.2) | 252 (26.6) | 8917 (28.1) | 224 (23.7) |
| 30 - 39.9 | 4016 (12.5) | 57 (9.2) | 4033 (12.6) | 58 (9.4) | 4021 (12.5) | 40 (7.8) | 4038 (12.6) | 65 (12.7) | 3962 (12.6) | 114 (9.9) | 3982 (12.6) | 106 (9.2) | 3949 (12.5) | 84 (8.9) | 4015 (12.7) | 116 (12.2) |
| >=40 | 307 (1.0) | 6 (1.0) | 291 (0.9) | 5 (0.8) | 330 (1.0) | 5 (1.0) | 268 (0.8) | 6 (1.2) | 287 (0.9) | 9 (0.8) | 299 (0.9) | 14 (1.2) | 287 (0.9) | 8 (0.8) | 301 (0.9) | 13 (1.4) |
| No. of GP visits** (%) | | | | | | | | | | | | | | | | |
| 0 | 1555 (4.9) | 32 (5.2) | 1596 (5.0) | 36 (5.8) | 1593 (5.0) | 27 (5.3) | 1578 (4.9) | 21 (4.1) | 1509 (4.8) | 71 (6.1) | 1576 (5.0) | 63 (5.5) | 1520 (4.8) | 47 (5.0) | 1604 (5.1) | 48 (5.1) |
| 1-5 | 8001 (25.0) | 95 (15.3) | 7959 (24.9) | 121 (19.5) | 7916 (24.6) | 90 (17.5) | 8084 (25.2) | 86 (16.8) | 7797 (24.8) | 200 (17.3) | 7980 (25.3) | 199 (17.2) | 7964 (25.1) | 161 (17.0) | 7891 (24.9) | 160 (16.9) |
| 6-10 | 8588 (26.8) | 138 (22.3) | 8600 (26.9) | 126 (20.4) | 8694 (27.1) | 109 (21.2) | 8537 (26.6) | 112 (21.8) | 8507 (27.0) | 259 (22.4) | 8427 (26.8) | 259 (22.4) | 8570 (27.0) | 222 (23.4) | 8453 (26.7) | 207 (21.9) |
| 11-15 | 5798 (18.1) | 111 (17.9) | 5898 (18.4) | 131 (21.2) | 5865 (18.3) | 91 (17.7) | 5878 (18.3) | 104 (20.3) | 5792 (18.4) | 219 (18.9) | 5689 (18.1) | 238 (20.6) | 5782 (18.2) | 174 (18.4) | 5791 (18.3) | 191 (20.2) |
| >=16 | 8087 (25.2) | 243 (39.3) | 7975 (24.9) | 205 (33.1) | 8067 (25.1) | 196 (38.2) | 8057 (25.1) | 190 (37.0) | 7887 (25.0) | 407 (35.2) | 7820 (24.8) | 396 (34.3) | 7865 (24.8) | 343 (36.2) | 7961 (25.1) | 341 (36.0) |
| No. of GP emergency visits** (%) | | | | | | | | | | | | | | | | |
| 0 | 26075 (81.4) | 407 (65.8) | 26200 (81.8) | 413 (66.7) | 26216 (81.6) | 339 (66.1) | 26186 (81.5) | 354 (69.0) | 25664 (81.5) | 815 (70.5) | 25832 (82.0) | 784 (67.9) | 25837 (81.5) | 680 (71.8) | 25924 (81.8) | 654 (69.1) |
| 1 | 3237 (10.1) | 88 (14.2) | 3161 (9.9) | 84 (13.6) | 3217 (10.0) | 74 (14.4) | 3218 (10.0) | 61 (11.9) | 3188 (10.1) | 144 (12.5) | 3083 (9.8) | 155 (13.4) | 3198 (10.1) | 105 (11.1) | 3138 (9.9) | 129 (13.6) |
| 2 | 1174 (3.7) | 47 (7.6) | 1171 (3.7) | 47 (7.6) | 1159 (3.6) | 39 (7.6) | 1201 (3.7) | 40 (7.8) | 1162 (3.7) | 85 (7.4) | 1118 (3.6) | 74 (6.4) | 1151 (3.6) | 63 (6.7) | 1157 (3.6) | 68 (7.2) |
| 3-5 | 1107 (3.5) | 52 (8.4) | 1078 (3.4) | 45 (7.3) | 1095 (3.4) | 36 (7.0) | 1114 (3.5) | 37 (7.2) | 1081 (3.4) | 64 (5.5) | 1041 (3.3) | 96 (8.3) | 1121 (3.5) | 65 (6.9) | 1038 (3.3) | 58 (6.1) |

| OST cohort | One year | | | | | | | | Two years | | | | | | | |
|---|---|---|---|---|---|---|---|---|---|---|---|---|---|---|---|---|
| | MACE | | | | Stroke/MI | | | | MACE | | | | Stroke/MI | | | |
| | Development set (n=32648) | | Internal validation set (n=32647) | | Development set (n=32648) | | Internal validation set (n=32647) | | Development set (n=32648) | | Internal validation set (n=32647) | | Development set (n=32648) | | Internal validation set (n=32647) | |
| | Outcome=NO | Outcome=YES | Outcome=NO | Outcome=YES | Outcome=NO | Outcome=YES | Outcome=NO | Outcome=YES | Outcome=NO | Outcome=YES | Outcome=NO | Outcome=YES | Outcome=NO | Outcome=YES | Outcome=NO | Outcome=YES |
| >=6 | 436 (1.4) | 25 (4.0) | 418 (1.3) | 30 (4.8) | 448 (1.4) | 25 (4.9) | 415 (1.3) | 21 (4.1) | 397 (1.3) | 48 (4.2) | 418 (1.3) | 46 (4.0) | 394 (1.2) | 34 (3.6) | 443 (1.4) | 38 (4.0) |
| eGFR** (%) | | | | | | | | | | | | | | | | |
| <=29 | 309 (1.0) | 23 (3.7) | 359 (1.1) | 17 (2.7) | 339 (1.1) | 12 (2.3) | 331 (1.0) | 26 (5.1) | 342 (1.1) | 32 (2.8) | 308 (1.0) | 26 (2.3) | 344 (1.1) | 18 (1.9) | 313 (1.0) | 33 (3.5) |
| 30 - 44 | 1672 (5.2) | 71 (11.5) | 1664 (5.2) | 63 (10.2) | 1676 (5.2) | 66 (12.9) | 1662 (5.2) | 66 (12.9) | 1584 (5.0) | 121 (10.5) | 1624 (5.2) | 141 (12.2) | 1576 (5.0) | 122 (12.9) | 1670 (5.3) | 102 (10.8) |
| 45 - 59 | 5867 (18.3) | 170 (27.5) | 5760 (18.0) | 153 (24.7) | 5795 (18.0) | 141 (27.5) | 5876 (18.3) | 138 (26.9) | 5703 (18.1) | 281 (24.3) | 5659 (18.0) | 307 (26.6) | 5704 (18.0) | 239 (25.2) | 5762 (18.2) | 245 (25.9) |
| 60 - 89 | 22375 (69.9) | 325 (52.5) | 22430 (70.0) | 367 (59.3) | 22558 (70.2) | 273 (53.2) | 22400 (69.7) | 266 (51.9) | 22043 (70.0) | 680 (58.8) | 22155 (70.4) | 619 (53.6) | 22241 (70.2) | 523 (55.2) | 22211 (70.1) | 522 (55.1) |
| >=90 | 1806 (5.6) | 30 (4.8) | 1815 (5.7) | 19 (3.1) | 1767 (5.5) | 21 (4.1) | 1865 (5.8) | 17 (3.3) | 1820 (5.8) | 42 (3.6) | 1746 (5.5) | 62 (5.4) | 1836 (5.8) | 45 (4.8) | 1744 (5.5) | 45 (4.8) |
| SBP** (%) | | | | | | | | | | | | | | | | |
| <120 | 4464 (13.9) | 69 (11.1) | 4449 (13.9) | 94 (15.2) | 4476 (13.9) | 60 (11.7) | 4465 (13.9) | 75 (14.6) | 4440 (14.1) | 136 (11.8) | 4359 (13.8) | 141 (12.2) | 4434 (14.0) | 112 (11.8) | 4416 (13.9) | 114 (12.0) |
| 120 - 139 | 13104 (40.9) | 223 (36.0) | 13053 (40.8) | 225 (36.3) | 13108 (40.8) | 179 (34.9) | 13137 (40.9) | 181 (35.3) | 12877 (40.9) | 418 (36.2) | 12894 (40.9) | 416 (36.0) | 12927 (40.8) | 333 (35.2) | 12999 (41.0) | 346 (36.5) |
| 140 - 159 | 11063 (34.5) | 250 (40.4) | 11070 (34.6) | 205 (33.1) | 11106 (34.6) | 194 (37.8) | 11099 (34.5) | 189 (36.8) | 10810 (34.3) | 433 (37.5) | 10909 (34.6) | 436 (37.7) | 10937 (34.5) | 375 (39.6) | 10936 (34.5) | 340 (35.9) |
| >=160 | 3398 (10.6) | 77 (12.4) | 3456 (10.8) | 95 (15.3) | 3445 (10.7) | 80 (15.6) | 3433 (10.7) | 68 (13.3) | 3365 (10.7) | 169 (14.6) | 3330 (10.6) | 162 (14.0) | 3403 (10.7) | 127 (13.4) | 3349 (10.6) | 147 (15.5) |
| DBP** (%) | | | | | | | | | | | | | | | | |
| <80 | 16431 (51.3) | 350 (56.5) | 16616 (51.9) | 332 (53.6) | 16547 (51.5) | 281 (54.8) | 16617 (51.7) | 284 (55.4) | 16245 (51.6) | 620 (53.6) | 16251 (51.6) | 613 (53.1) | 16367 (51.6) | 505 (53.3) | 16352 (51.6) | 505 (53.3) |
| 80 - 89 | 12144 (37.9) | 208 (33.6) | 11934 (37.3) | 219 (35.4) | 12106 (37.7) | 175 (34.1) | 12049 (37.5) | 175 (34.1) | 11868 (37.7) | 424 (36.7) | 11795 (37.5) | 418 (36.2) | 11895 (37.5) | 349 (36.9) | 11919 (37.6) | 342 (36.1) |
| 90 - 99 | 2795 (8.7) | 50 (8.1) | 2868 (9.0) | 51 (8.2) | 2847 (8.9) | 45 (8.8) | 2829 (8.8) | 43 (8.4) | 2755 (8.7) | 90 (7.8) | 2822 (9.0) | 97 (8.4) | 2825 (8.9) | 77 (8.1) | 2786 (8.8) | 76 (8.0) |
| >=100 | 659 (2.1) | 11 (1.8) | 610 (1.9) | 17 (2.7) | 635 (2.0) | 12 (2.3) | 639 (2.0) | 11 (2.1) | 624 (2.0) | 22 (1.9) | 624 (2.0) | 27 (2.3) | 614 (1.9) | 16 (1.7) | 643 (2.0) | 24 (2.5) |
| No. of concomitant medicines** (%) | | | | | | | | | | | | | | | | |

| OST cohort | One year | | | | | | | | Two years | | | | | | | |
|---|---|---|---|---|---|---|---|---|---|---|---|---|---|---|---|---|
| | MACE | | | | Stroke/MI | | | | MACE | | | | Stroke/MI | | | |
| | Development set (n=32648) | | Internal validation set (n=32647) | | Development set (n=32648) | | Internal validation set (n=32647) | | Development set (n=32648) | | Internal validation set (n=32647) | | Development set (n=32648) | | Internal validation set (n=32647) | |
| | Outcome=NO | Outcome=YES | Outcome=NO | Outcome=YES | Outcome=NO | Outcome=YES | Outcome=NO | Outcome=YES | Outcome=NO | Outcome=YES | Outcome=NO | Outcome=YES | Outcome=NO | Outcome=YES | Outcome=NO | Outcome=YES |
| 0 | 2541 (7.9) | 38 (6.1) | 2548 (8.0) | 44 (7.1) | 2580 (8.0) | 35 (6.8) | 2532 (7.9) | 24 (4.7) | 2423 (7.7) | 81 (7.0) | 2601 (8.3) | 66 (5.7) | 2504 (7.9) | 50 (5.3) | 2563 (8.1) | 54 (5.7) |
| 1 - 3 | 6366 (19.9) | 51 (8.2) | 6380 (19.9) | 61 (9.9) | 6377 (19.8) | 49 (9.6) | 6382 (19.9) | 50 (9.7) | 6327 (20.1) | 115 (9.9) | 6301 (20.0) | 115 (10.0) | 6399 (20.2) | 109 (11.5) | 6259 (19.7) | 91 (9.6) |
| 4 - 6 | 7192 (22.5) | 103 (16.6) | 7060 (22.0) | 91 (14.7) | 7123 (22.2) | 69 (13.5) | 7168 (22.3) | 86 (16.8) | 7012 (22.3) | 200 (17.3) | 7047 (22.4) | 187 (16.2) | 7103 (22.4) | 147 (15.5) | 7032 (22.2) | 164 (17.3) |
| 7 - 9 | 6061 (18.9) | 121 (19.5) | 6052 (18.9) | 141 (22.8) | 5981 (18.6) | 116 (22.6) | 6177 (19.2) | 101 (19.7) | 5984 (19.0) | 239 (20.7) | 5919 (18.8) | 233 (20.2) | 5985 (18.9) | 194 (20.5) | 6007 (18.9) | 189 (20.0) |
| 10 - 12 | 4253 (13.3) | 119 (19.2) | 4326 (13.5) | 90 (14.5) | 4360 (13.6) | 86 (16.8) | 4260 (13.3) | 82 (16.0) | 4202 (13.3) | 201 (17.4) | 4179 (13.3) | 206 (17.8) | 4205 (13.3) | 178 (18.8) | 4252 (13.4) | 153 (16.2) |
| >=13 | 5616 (17.5) | 187 (30.2) | 5662 (17.7) | 192 (31.0) | 5714 (17.8) | 158 (30.8) | 5615 (17.5) | 170 (33.1) | 5544 (17.6) | 320 (27.7) | 5445 (17.3) | 348 (30.1) | 5505 (17.4) | 269 (28.4) | 5587 (17.6) | 296 (31.3) |
| Cholesterol measurement** (HDL/LDL) (%) | | | | | | | | | | | | | | | | |
| <=3.5 | 19256 (60.1) | 355 (57.4) | 19195 (59.9) | 337 (54.4) | 19260 (59.9) | 308 (60.0) | 19287 (60.0) | 288 (56.1) | 18799 (59.7) | 662 (57.3) | 18990 (60.3) | 692 (59.9) | 19074 (60.2) | 546 (57.7) | 18958 (59.8) | 565 (59.7) |
| 3.6 - 5 | 10175 (31.8) | 211 (34.1) | 10189 (31.8) | 217 (35.1) | 10227 (31.8) | 157 (30.6) | 10223 (31.8) | 185 (36.1) | 10164 (32.3) | 392 (33.9) | 9875 (31.4) | 361 (31.3) | 10081 (31.8) | 305 (32.2) | 10096 (31.8) | 310 (32.7) |
| >5 | 2598 (8.1) | 53 (8.6) | 2644 (8.3) | 65 (10.5) | 2648 (8.2) | 48 (9.4) | 2624 (8.2) | 40 (7.8) | 2529 (8.0) | 102 (8.8) | 2627 (8.3) | 102 (8.8) | 2546 (8.0) | 96 (10.1) | 2646 (8.3) | 72 (7.6) |
| No. of previous fractures* (%) | | | | | | | | | | | | | | | | |
| 0 | 22780 (71.1) | 404 (65.3) | 22957 (71.7) | 410 (66.2) | 22909 (71.3) | 339 (66.1) | 22979 (71.5) | 324 (63.2) | 22504 (71.5) | 799 (69.1) | 22495 (71.4) | 753 (65.2) | 22636 (71.4) | 616 (65.0) | 22660 (71.5) | 639 (67.5) |
| 1 | 4797 (15.0) | 129 (20.8) | 4730 (14.8) | 107 (17.3) | 4835 (15.0) | 99 (19.3) | 4724 (14.7) | 105 (20.5) | 4666 (14.8) | 198 (17.1) | 4693 (14.9) | 206 (17.8) | 4734 (14.9) | 179 (18.9) | 4686 (14.8) | 164 (17.3) |
| >=2 | 4452 (13.9) | 86 (13.9) | 4341 (13.6) | 102 (16.5) | 4391 (13.7) | 75 (14.6) | 4431 (13.8) | 84 (16.4) | 4322 (13.7) | 159 (13.8) | 4304 (13.7) | 196 (17.0) | 4331 (13.7) | 152 (16.1) | 4354 (13.7) | 144 (15.2) |

**Abbreviations**: OST, patients with incident diagnosis of osteoporosis; IFX, patients with incident fragility fracture; OBP, incident users of oral bisphosphonates; MACE, composite outcome for the occurrence of either myocardial infarction, stroke or cardiovascular disease death; MI, myocardial infarction; * ever; ** in the year prior to start; SES, socio-economic status; BMI, body mass index; eGFR, estimated Glomerular Filtration Rate; SBP, cholesterol, systolic blood pressure; DBP, diastolic blood pressure.

#### Table S2b Dataset split into train and test sets, stratified by outcome (IFX cohort)

| IFX cohort | One year | | | | | | | | Two years | | | | | | | |
|---|---|---|---|---|---|---|---|---|---|---|---|---|---|---|---|---|
| | MACE | | | | Stroke/MI | | | | MACE | | | | Stroke/MI | | | |
| | Development set (n=40294) | | Internal validation set (n=40293) | | Development set (n=40294) | | Internal validation set (n=40293) | | Development set (n=40294) | | Internal validation set (n=40293) | | Development set (n=40294) | | Internal validation set (n=40293) | |
| | Outcome=NO | Outcome=YES | Outcome=NO | Outcome=YES | Outcome=NO | Outcome=YES | Outcome=NO | Outcome=YES | Outcome=NO | Outcome=YES | Outcome=NO | Outcome=YES | Outcome=NO | Outcome=YES | Outcome=NO | Outcome=YES |
| n | 38567 | 1727 | 38567 | 1726 | 39244 | 1050 | 39244 | 1049 | 37680 | 2614 | 37680 | 2613 | 38616 | 1678 | 38616 | 1677 |
| Sex = Male (%) | 8945 (23.2) | 467 (27.0) | 8895 (23.1) | 487 (28.2) | 9106 (23.2) | 269 (25.6) | 9159 (23.3) | 260 (24.8) | 8680 (23.0) | 697 (26.7) | 8733 (23.2) | 684 (26.2) | 9020 (23.4) | 415 (24.7) | 8955 (23.2) | 404 (24.1) |
| SES (%) | | | | | | | | | | | | | | | | |
| 1 | 8743 (22.7) | 366 (21.2) | 8684 (22.5) | 362 (21.0) | 8912 (22.7) | 224 (21.3) | 8822 (22.5) | 197 (18.8) | 8520 (22.6) | 564 (21.6) | 8519 (22.6) | 552 (21.1) | 8719 (22.6) | 330 (19.7) | 8752 (22.7) | 354 (21.1) |
| 2 | 9193 (23.8) | 404 (23.4) | 9188 (23.8) | 409 (23.7) | 9303 (23.7) | 251 (23.9) | 9397 (23.9) | 243 (23.2) | 8897 (23.6) | 602 (23.0) | 9072 (24.1) | 623 (23.8) | 9104 (23.6) | 376 (22.4) | 9313 (24.1) | 401 (23.9) |
| 3 | 8162 (21.2) | 368 (21.3) | 8290 (21.5) | 357 (20.7) | 8351 (21.3) | 211 (20.1) | 8376 (21.3) | 239 (22.8) | 8176 (21.7) | 566 (21.7) | 7908 (21.0) | 527 (20.2) | 8335 (21.6) | 382 (22.8) | 8123 (21.0) | 337 (20.1) |
| 4 | 7372 (19.1) | 338 (19.6) | 7332 (19.0) | 339 (19.6) | 7523 (19.2) | 202 (19.2) | 7447 (19.0) | 209 (19.9) | 7063 (18.7) | 498 (19.1) | 7296 (19.4) | 524 (20.1) | 7417 (19.2) | 324 (19.3) | 7300 (18.9) | 340 (20.3) |
| 5 | 5067 (13.1) | 247 (14.3) | 5042 (13.1) | 257 (14.9) | 5124 (13.1) | 159 (15.1) | 5170 (13.2) | 160 (15.3) | 4994 (13.3) | 378 (14.5) | 4855 (12.9) | 386 (14.8) | 5020 (13.0) | 262 (15.6) | 5087 (13.2) | 244 (14.5) |
| Smoking** (%) | | | | | | | | | | | | | | | | |
| Ex | 12488 (32.4) | 590 (34.2) | 12205 (31.6) | 564 (32.7) | 12554 (32.0) | 338 (32.2) | 12586 (32.1) | 369 (35.2) | 12141 (32.2) | 843 (32.2) | 11981 (31.8) | 882 (33.8) | 12351 (32.0) | 577 (34.4) | 12370 (32.0) | 549 (32.7) |
| No | 19696 (51.1) | 921 (53.3) | 19973 (51.8) | 941 (54.5) | 20256 (51.6) | 573 (54.6) | 20147 (51.3) | 555 (52.9) | 19316 (51.3) | 1396 (53.4) | 19476 (51.7) | 1343 (51.4) | 19870 (51.5) | 857 (51.1) | 19930 (51.6) | 874 (52.1) |
| Yes | 6383 (16.6) | 216 (12.5) | 6389 (16.6) | 221 (12.8) | 6434 (16.4) | 139 (13.2) | 6511 (16.6) | 125 (11.9) | 6223 (16.5) | 375 (14.3) | 6223 (16.5) | 388 (14.8) | 6395 (16.6) | 244 (14.5) | 6316 (16.4) | 254 (15.1) |
| Drinking** (%) | | | | | | | | | | | | | | | | |
| Ex | 2713 (7.0) | 143 (8.3) | 2683 (7.0) | 137 (7.9) | 2680 (6.8) | 94 (9.0) | 2814 (7.2) | 88 (8.4) | 2655 (7.0) | 224 (8.6) | 2564 (6.8) | 233 (8.9) | 2660 (6.9) | 159 (9.5) | 2710 (7.0) | 147 (8.8) |
| No | 11437 (29.7) | 617 (35.7) | 11644 (30.2) | 627 (36.3) | 11780 (30.0) | 371 (35.3) | 11815 (30.1) | 359 (34.2) | 11175 (29.7) | 945 (36.2) | 11253 (29.9) | 952 (36.4) | 11549 (29.9) | 583 (34.7) | 11612 (30.1) | 581 (34.6) |
| Yes | 24417 (63.3) | 967 (56.0) | 24240 (62.9) | 962 (55.7) | 24784 (63.2) | 585 (55.7) | 24615 (62.7) | 602 (57.4) | 23850 (63.3) | 1445 (55.3) | 23863 (63.3) | 1428 (54.6) | 24407 (63.2) | 936 (55.8) | 24294 (62.9) | 949 (56.6) |
| Diabetes type I*= 1 (%) | 109 (0.3) | 6 (0.3) | 98 (0.3) | 10 (0.6) | 109 (0.3) | 5 (0.5) | 103 (0.3) | 6 (0.6) | 106 (0.3) | 9 (0.3) | 97 (0.3) | 11 (0.4) | 99 (0.3) | 6 (0.4) | 111 (0.3) | 7 (0.4) |

| IFX cohort | One year | | | | | | | | Two years | | | | | | | |
|---|---|---|---|---|---|---|---|---|---|---|---|---|---|---|---|---|
| | MACE | | | | Stroke/MI | | | | MACE | | | | Stroke/MI | | | |
| | Development set (n=40294) | | Internal validation set (n=40293) | | Development set (n=40294) | | Internal validation set (n=40293) | | Development set (n=40294) | | Internal validation set (n=40293) | | Development set (n=40294) | | Internal validation set (n=40293) | |
| | Outcome=NO | Outcome=YES | Outcome=NO | Outcome=YES | Outcome=NO | Outcome=YES | Outcome=NO | Outcome=YES | Outcome=NO | Outcome=YES | Outcome=NO | Outcome=YES | Outcome=NO | Outcome=YES | Outcome=NO | Outcome=YES |
| Diabetes type II*= 1 (%) | 2470 (6.4) | 134 (7.8) | 2487 (6.4) | 154 (8.9) | 2482 (6.3) | 99 (9.4) | 2552 (6.5) | 112 (10.7) | 2444 (6.5) | 204 (7.8) | 2385 (6.3) | 212 (8.1) | 2462 (6.4) | 164 (9.8) | 2464 (6.4) | 155 (9.2) |
| Chronic obstructive pulmonary disease*= 1 (%) | 2237 (5.8) | 106 (6.1) | 2263 (5.9) | 127 (7.4) | 2363 (6.0) | 69 (6.6) | 2231 (5.7) | 70 (6.7) | 2160 (5.7) | 171 (6.5) | 2247 (6.0) | 155 (5.9) | 2224 (5.8) | 110 (6.6) | 2298 (6.0) | 101 (6.0) |
| Chronic kidney disease*= 1 (%) | 4761 (12.3) | 327 (18.9) | 4842 (12.6) | 302 (17.5) | 4917 (12.5) | 200 (19.0) | 4938 (12.6) | 177 (16.9) | 4724 (12.5) | 462 (17.7) | 4606 (12.2) | 440 (16.8) | 4857 (12.6) | 300 (17.9) | 4806 (12.4) | 269 (16.0) |
| Rheumatoid arthritis*= 1 (%) | 1788 (4.6) | 98 (5.7) | 1786 (4.6) | 72 (4.2) | 1811 (4.6) | 57 (5.4) | 1816 (4.6) | 60 (5.7) | 1711 (4.5) | 143 (5.5) | 1766 (4.7) | 124 (4.7) | 1771 (4.6) | 92 (5.5) | 1788 (4.6) | 93 (5.5) |
| Lupus*= 1 (%) | 38 (0.1) | 3 (0.2) | 34 (0.1) | 0 (0.0) | 38 (0.1) | 0 (0.0) | 35 (0.1) | 2 (0.2) | 31 (0.1) | 2 (0.1) | 39 (0.1) | 3 (0.1) | 34 (0.1) | 3 (0.2) | 37 (0.1) | 1 (0.1) |
| Systemic heart disease**= 1 (%) | 374 (1.0) | 1 (0.1) | 347 (0.9) | 3 (0.2) | 358 (0.9) | 1 (0.1) | 364 (0.9) | 2 (0.2) | 345 (0.9) | 13 (0.5) | 361 (1.0) | 6 (0.2) | 373 (1.0) | 8 (0.5) | 335 (0.9) | 9 (0.5) |
| Anti-osteoporosis use**= 1 (%) | 3618 (9.4) | 186 (10.8) | 3750 (9.7) | 172 (10.0) | 3716 (9.5) | 108 (10.3) | 3770 (9.6) | 132 (12.6) | 3551 (9.4) | 253 (9.7) | 3643 (9.7) | 279 (10.7) | 3667 (9.5) | 186 (11.1) | 3698 (9.6) | 175 (10.4) |
| Heparin use**= 1 (%) | 167 (0.4) | 9 (0.5) | 186 (0.5) | 6 (0.3) | 184 (0.5) | 6 (0.6) | 172 (0.4) | 6 (0.6) | 180 (0.5) | 17 (0.7) | 162 (0.4) | 9 (0.3) | 175 (0.5) | 12 (0.7) | 174 (0.5) | 7 (0.4) |
| Beta-blocker use**= 1 (%) | 6063 (15.7) | 375 (21.7) | 5863 (15.2) | 397 (23.0) | 6078 (15.5) | 256 (24.4) | 6124 (15.6) | 240 (22.9) | 5798 (15.4) | 585 (22.4) | 5765 (15.3) | 550 (21.0) | 6041 (15.6) | 385 (22.9) | 5879 (15.2) | 393 (23.4) |
| Hypertension**= 1 (%) | 1900 (4.9) | 107 (6.2) | 1947 (5.0) | 94 (5.4) | 1970 (5.0) | 84 (8.0) | 1928 (4.9) | 66 (6.3) | 1893 (5.0) | 163 (6.2) | 1844 (4.9) | 148 (5.7) | 1934 (5.0) | 119 (7.1) | 1883 (4.9) | 112 (6.7) |
| Deep vein thrombosis or pulmonary embolism**= 1 (%) | 242 (0.6) | 17 (1.0) | 273 (0.7) | 13 (0.8) | 263 (0.7) | 10 (1.0) | 262 (0.7) | 10 (1.0) | 268 (0.7) | 20 (0.8) | 233 (0.6) | 24 (0.9) | 271 (0.7) | 19 (1.1) | 244 (0.6) | 11 (0.7) |
| Anticoagulant use**= 1 (%) | 2016 (5.2) | 151 (8.7) | 1977 (5.1) | 130 (7.5) | 2095 (5.3) | 85 (8.1) | 1999 (5.1) | 95 (9.1) | 1970 (5.2) | 195 (7.5) | 1922 (5.1) | 187 (7.2) | 2031 (5.3) | 138 (8.2) | 1984 (5.1) | 121 (7.2) |
| Antidepressants TCA**= 1 (%) | 3173 (8.2) | 159 (9.2) | 3205 (8.3) | 162 (9.4) | 3269 (8.3) | 95 (9.0) | 3231 (8.2) | 104 (9.9) | 3125 (8.3) | 223 (8.5) | 3088 (8.2) | 263 (10.1) | 3176 (8.2) | 159 (9.5) | 3205 (8.3) | 159 (9.5) |
| Antidepressants SSRI**= 1 (%) | 4364 (11.3) | 231 (13.4) | 4381 (11.4) | 233 (13.5) | 4553 (11.6) | 135 (12.9) | 4401 (11.2) | 120 (11.4) | 4230 (11.2) | 352 (13.5) | 4301 (11.4) | 326 (12.5) | 4357 (11.3) | 175 (10.4) | 4464 (11.6) | 213 (12.7) |
| Hypercholesterolemia**= 1 (%) | 417 (1.1) | 16 (0.9) | 389 (1.0) | 21 (1.2) | 415 (1.1) | 17 (1.6) | 402 (1.0) | 9 (0.9) | 381 (1.0) | 25 (1.0) | 407 (1.1) | 30 (1.1) | 407 (1.1) | 16 (1.0) | 396 (1.0) | 24 (1.4) |
| Statin use**= 1 (%) | 8271 (21.4) | 465 (26.9) | 8213 (21.3) | 460 (26.7) | 8356 (21.3) | 303 (28.9) | 8458 (21.6) | 292 (27.8) | 8051 (21.4) | 661 (25.3) | 8001 (21.2) | 696 (26.6) | 8347 (21.6) | 476 (28.4) | 8149 (21.1) | 437 (26.1) |

| IFX cohort | One year | | | | | | | | Two years | | | | | | | |
|---|---|---|---|---|---|---|---|---|---|---|---|---|---|---|---|---|
| | MACE | | | | Stroke/MI | | | | MACE | | | | Stroke/MI | | | |
| | Development set (n=40294) | | Internal validation set (n=40293) | | Development set (n=40294) | | Internal validation set (n=40293) | | Development set (n=40294) | | Internal validation set (n=40293) | | Development set (n=40294) | | Internal validation set (n=40293) | |
| | Outcome=NO | Outcome=YES | Outcome=NO | Outcome=YES | Outcome=NO | Outcome=YES | Outcome=NO | Outcome=YES | Outcome=NO | Outcome=YES | Outcome=NO | Outcome=YES | Outcome=NO | Outcome=YES | Outcome=NO | Outcome=YES |
| Osteoporosis history* = 1 | 3884 (10.1) | 193 (11.2) | 4047 (10.5) | 175 (10.1) | 4006 (10.2) | 115 (11.0) | 4057 (10.3) | 121 (11.5) | 3825 (10.2) | 278 (10.6) | 3909 (10.4) | 287 (11.0) | 3912 (10.1) | 190 (11.3) | 4008 (10.4) | 189 (11.3) |
| Family history of cardiovascular disease = 1 (%) | 2633 (6.8) | 124 (7.2) | 2624 (6.8) | 92 (5.3) | 2704 (6.9) | 60 (5.7) | 2628 (6.7) | 81 (7.7) | 2599 (6.9) | 157 (6.0) | 2566 (6.8) | 151 (5.8) | 2612 (6.8) | 117 (7.0) | 2650 (6.9) | 94 (5.6) |
| Family history of cardiovascular disease before age 60= 1 (%) | 31 (0.1) | 1 (0.1) | 26 (0.1) | 0 (0.0) | 25 (0.1) | 0 (0.0) | 33 (0.1) | 0 (0.0) | 34 (0.1) | 0 (0.0) | 23 (0.1) | 1 (0.0) | 24 (0.1) | 0 (0.0) | 34 (0.1) | 0 (0.0) |
| Heart failure*= 1 (%) | 1933 (5.0) | 170 (9.8) | 1957 (5.1) | 163 (9.4) | 2048 (5.2) | 90 (8.6) | 1991 (5.1) | 94 (9.0) | 1921 (5.1) | 212 (8.1) | 1848 (4.9) | 242 (9.3) | 1974 (5.1) | 135 (8.0) | 1978 (5.1) | 136 (8.1) |
| Migraine*= 1 (%) | 4123 (10.7) | 185 (10.7) | 4189 (10.9) | 175 (10.1) | 4210 (10.7) | 124 (11.8) | 4222 (10.8) | 116 (11.1) | 4106 (10.9) | 268 (10.3) | 4010 (10.6) | 288 (11.0) | 4163 (10.8) | 206 (12.3) | 4126 (10.7) | 177 (10.6) |
| Severe mental illness*= 1 (%) | 5663 (14.7) | 247 (14.3) | 5742 (14.9) | 259 (15.0) | 5875 (15.0) | 158 (15.0) | 5711 (14.6) | 167 (15.9) | 5611 (14.9) | 397 (15.2) | 5506 (14.6) | 397 (15.2) | 5666 (14.7) | 274 (16.3) | 5714 (14.8) | 257 (15.3) |
| Vascular Disease*= 1 (%) | 613 (1.6) | 58 (3.4) | 625 (1.6) | 55 (3.2) | 650 (1.7) | 36 (3.4) | 626 (1.6) | 39 (3.7) | 592 (1.6) | 82 (3.1) | 599 (1.6) | 78 (3.0) | 612 (1.6) | 56 (3.3) | 632 (1.6) | 51 (3.0) |
| Atrial fibrillation*= 1 (%) | 2765 (7.2) | 231 (13.4) | 2746 (7.1) | 214 (12.4) | 2886 (7.4) | 129 (12.3) | 2792 (7.1) | 149 (14.2) | 2689 (7.1) | 327 (12.5) | 2658 (7.1) | 282 (10.8) | 2791 (7.2) | 197 (11.7) | 2768 (7.2) | 200 (11.9) |
| On anti-hypertensive drug= 1 (%) | 22354 (58.0) | 1256 (72.7) | 22389 (58.1) | 1248 (72.3) | 22769 (58.0) | 758 (72.2) | 22954 (58.5) | 766 (73.0) | 21866 (58.0) | 1852 (70.8) | 21700 (57.6) | 1829 (70.0) | 22410 (58.0) | 1219 (72.6) | 22474 (58.2) | 1144 (68.2) |
| Antipsychotic use**= 1 (%) | 489 (1.3) | 23 (1.3) | 481 (1.2) | 42 (2.4) | 499 (1.3) | 17 (1.6) | 506 (1.3) | 13 (1.2) | 472 (1.3) | 48 (1.8) | 478 (1.3) | 37 (1.4) | 526 (1.4) | 21 (1.3) | 469 (1.2) | 19 (1.1) |
| Steroid use**= 1 (%) | 3044 (7.9) | 144 (8.3) | 3100 (8.0) | 135 (7.8) | 3172 (8.1) | 95 (9.0) | 3062 (7.8) | 94 (9.0) | 2976 (7.9) | 199 (7.6) | 3035 (8.1) | 213 (8.2) | 3052 (7.9) | 157 (9.4) | 3083 (8.0) | 131 (7.8) |
| Erectile dysfunction**= 1 (%) | 653 (1.7) | 27 (1.6) | 643 (1.7) | 26 (1.5) | 634 (1.6) | 13 (1.2) | 682 (1.7) | 20 (1.9) | 615 (1.6) | 39 (1.5) | 647 (1.7) | 48 (1.8) | 629 (1.6) | 38 (2.3) | 655 (1.7) | 27 (1.6) |
| Age>75 | 27037 (70.1) | 1560 (90.3) | 27058 (70.2) | 1522 (88.2) | 27678 (70.5) | 912 (86.9) | 27677 (70.5) | 910 (86.7) | 26303 (69.8) | 2301 (88.0) | 26267 (69.7) | 2306 (88.3) | 27190 (70.4) | 1432 (85.3) | 27098 (70.2) | 1457 (86.9) |
| Age Group (%) | | | | | | | | | | | | | | | | |
| 50-59 | 3131 (8.1) | 10 (0.6) | 3084 (8.0) | 17 (1.0) | 3089 (7.9) | 13 (1.2) | 3129 (8.0) | 11 (1.0) | 3034 (8.1) | 32 (1.2) | 3143 (8.3) | 33 (1.3) | 3090 (8.0) | 25 (1.5) | 3099 (8.0) | 28 (1.7) |
| 60-69 | 4862 (12.6) | 67 (3.9) | 4854 (12.6) | 92 (5.3) | 4911 (12.5) | 64 (6.1) | 4844 (12.3) | 56 (5.3) | 4820 (12.8) | 126 (4.8) | 4795 (12.7) | 134 (5.1) | 4811 (12.5) | 103 (6.1) | 4869 (12.6) | 92 (5.5) |

| IFX cohort | One year | | | | | | | | Two years | | | | | | | |
|---|---|---|---|---|---|---|---|---|---|---|---|---|---|---|---|---|
| | MACE | | | | Stroke/MI | | | | MACE | | | | Stroke/MI | | | |
| | Development set (n=40294) | | Internal validation set (n=40293) | | Development set (n=40294) | | Internal validation set (n=40293) | | Development set (n=40294) | | Internal validation set (n=40293) | | Development set (n=40294) | | Internal validation set (n=40293) | |
| | Outcome=NO | Outcome=YES | Outcome=NO | Outcome=YES | Outcome=NO | Outcome=YES | Outcome=NO | Outcome=YES | Outcome=NO | Outcome=YES | Outcome=NO | Outcome=YES | Outcome=NO | Outcome=YES | Outcome=NO | Outcome=YES |
| 70-79 | 9118 (23.6) | 315 (18.2) | 8954 (23.2) | 299 (17.3) | 9159 (23.3) | 222 (21.1) | 9091 (23.2) | 214 (20.4) | 8806 (23.4) | 495 (18.9) | 8901 (23.6) | 484 (18.5) | 9013 (23.3) | 356 (21.2) | 8955 (23.2) | 362 (21.6) |
| 80-89 | 15156 (39.3) | 887 (51.4) | 15379 (39.9) | 826 (47.9) | 15497 (39.5) | 521 (49.6) | 15699 (40.0) | 531 (50.6) | 14848 (39.4) | 1334 (51.0) | 14750 (39.1) | 1316 (50.4) | 15194 (39.3) | 880 (52.4) | 15336 (39.7) | 838 (50.0) |
| >89 | 6300 (16.3) | 448 (25.9) | 6296 (16.3) | 492 (28.5) | 6588 (16.8) | 230 (21.9) | 6481 (16.5) | 237 (22.6) | 6172 (16.4) | 627 (24.0) | 6091 (16.2) | 646 (24.7) | 6508 (16.9) | 314 (18.7) | 6357 (16.5) | 357 (21.3) |
| Charlson score (%) | | | | | | | | | | | | | | | | |
| 0 | 22568 (58.5) | 805 (46.6) | 22296 (57.8) | 816 (47.3) | 22785 (58.1) | 492 (46.9) | 22711 (57.9) | 497 (47.4) | 22001 (58.4) | 1293 (49.5) | 21925 (58.2) | 1266 (48.5) | 22447 (58.1) | 810 (48.3) | 22383 (58.0) | 845 (50.4) |
| 1 | 6584 (17.1) | 318 (18.4) | 6768 (17.5) | 342 (19.8) | 6802 (17.3) | 197 (18.8) | 6818 (17.4) | 195 (18.6) | 6436 (17.1) | 475 (18.2) | 6593 (17.5) | 508 (19.4) | 6669 (17.3) | 300 (17.9) | 6733 (17.4) | 310 (18.5) |
| 2 | 4503 (11.7) | 250 (14.5) | 4391 (11.4) | 236 (13.7) | 4501 (11.5) | 137 (13.0) | 4585 (11.7) | 157 (15.0) | 4311 (11.4) | 348 (13.3) | 4354 (11.6) | 367 (14.0) | 4409 (11.4) | 230 (13.7) | 4513 (11.7) | 228 (13.6) |
| ≥3 | 4912 (12.7) | 354 (20.5) | 5112 (13.3) | 332 (19.2) | 5156 (13.1) | 224 (21.3) | 5130 (13.1) | 200 (19.1) | 4932 (13.1) | 498 (19.1) | 4808 (12.8) | 472 (18.1) | 5091 (13.2) | 338 (20.1) | 4987 (12.9) | 294 (17.5) |
| Cardiovascular disease (%) | | | | | | | | | | | | | | | | |
| No | 33633 (87.2) | 1357 (78.6) | 33540 (87.0) | 1363 (79.0) | 34098 (86.9) | 812 (77.3) | 34169 (87.1) | 814 (77.6) | 32866 (87.2) | 2080 (79.6) | 32873 (87.2) | 2074 (79.4) | 33730 (87.3) | 1287 (76.7) | 33543 (86.9) | 1333 (79.5) |
| Ever >1 year before index date | 4030 (10.4) | 281 (16.3) | 4116 (10.7) | 275 (15.9) | 4191 (10.7) | 170 (16.2) | 4157 (10.6) | 184 (17.5) | 3926 (10.4) | 400 (15.3) | 3952 (10.5) | 424 (16.2) | 3971 (10.3) | 297 (17.7) | 4172 (10.8) | 262 (15.6) |
| 1 year before index | 399 (1.0) | 36 (2.1) | 401 (1.0) | 36 (2.1) | 402 (1.0) | 30 (2.9) | 422 (1.1) | 18 (1.7) | 394 (1.0) | 58 (2.2) | 379 (1.0) | 41 (1.6) | 403 (1.0) | 36 (2.1) | 404 (1.0) | 29 (1.7) |
| 6 months before index | 406 (1.1) | 37 (2.1) | 431 (1.1) | 35 (2.0) | 455 (1.2) | 26 (2.5) | 407 (1.0) | 21 (2.0) | 396 (1.1) | 55 (2.1) | 406 (1.1) | 52 (2.0) | 425 (1.1) | 36 (2.1) | 405 (1.0) | 43 (2.6) |
| 1 month before index | 99 (0.3) | 16 (0.9) | 79 (0.2) | 17 (1.0) | 98 (0.2) | 12 (1.1) | 89 (0.2) | 12 (1.1) | 98 (0.3) | 21 (0.8) | 70 (0.2) | 22 (0.8) | 87 (0.2) | 22 (1.3) | 92 (0.2) | 10 (0.6) |
| MI or Stroke (%) | | | | | | | | | | | | | | | | |
| No | 34791 (90.2) | 1320 (76.4) | 34674 (89.9) | 1358 (78.7) | 35255 (89.8) | 824 (78.5) | 35223 (89.8) | 841 (80.2) | 33994 (90.2) | 2052 (78.5) | 34052 (90.4) | 2045 (78.3) | 34678 (89.8) | 1333 (79.4) | 34774 (90.1) | 1358 (81.0) |
| Ever >1 year before index date | 2788 (7.2) | 277 (16.0) | 2857 (7.4) | 250 (14.5) | 2931 (7.5) | 148 (14.1) | 2952 (7.5) | 141 (13.4) | 2711 (7.2) | 382 (14.6) | 2697 (7.2) | 382 (14.6) | 2912 (7.5) | 234 (13.9) | 2808 (7.3) | 218 (13.0) |
| 1 year before index | 988 (2.6) | 130 (7.5) | 1036 (2.7) | 118 (6.8) | 1058 (2.7) | 78 (7.4) | 1069 (2.7) | 67 (6.4) | 975 (2.6) | 180 (6.9) | 931 (2.5) | 186 (7.1) | 1026 (2.7) | 111 (6.6) | 1034 (2.7) | 101 (6.0) |

| IFX cohort | One year | | | | | | | | Two years | | | | | | | |
|---|---|---|---|---|---|---|---|---|---|---|---|---|---|---|---|---|
| | MACE | | | | Stroke/MI | | | | MACE | | | | Stroke/MI | | | |
| | Development set (n=40294) | | Internal validation set (n=40293) | | Development set (n=40294) | | Internal validation set (n=40293) | | Development set (n=40294) | | Internal validation set (n=40293) | | Development set (n=40294) | | Internal validation set (n=40293) | |
| | Outcome=NO | Outcome=YES | Outcome=NO | Outcome=YES | Outcome=NO | Outcome=YES | Outcome=NO | Outcome=YES | Outcome=NO | Outcome=YES | Outcome=NO | Outcome=YES | Outcome=NO | Outcome=YES | Outcome=NO | Outcome=YES |
| Established CVD *= Ever (%) | 6291 (16.3) | 583 (33.8) | 6502 (16.9) | 566 (32.8) | 6630 (16.9) | 340 (32.4) | 6650 (16.9) | 322 (30.7) | 6200 (16.5) | 821 (31.4) | 6089 (16.2) | 832 (31.8) | 6474 (16.8) | 523 (31.2) | 6469 (16.8) | 476 (28.4) |
| Any fracture history (%) | | | | | | | | | | | | | | | | |
| No | 32016 (83.0) | 1464 (84.8) | 32108 (83.3) | 1477 (85.6) | 32731 (83.4) | 888 (84.6) | 32556 (83.0) | 890 (84.8) | 31274 (83.0) | 2219 (84.9) | 31370 (83.3) | 2202 (84.3) | 32068 (83.0) | 1413 (84.2) | 32173 (83.3) | 1411 (84.1) |
| Ever >1 year before index date | 3492 (9.1) | 160 (9.3) | 3481 (9.0) | 165 (9.6) | 3537 (9.0) | 101 (9.6) | 3556 (9.1) | 104 (9.9) | 3458 (9.2) | 253 (9.7) | 3334 (8.8) | 253 (9.7) | 3495 (9.1) | 175 (10.4) | 3468 (9.0) | 160 (9.5) |
| 1 year before index | 3059 (7.9) | 103 (6.0) | 2978 (7.7) | 84 (4.9) | 2976 (7.6) | 61 (5.8) | 3132 (8.0) | 55 (5.2) | 2948 (7.8) | 142 (5.4) | 2976 (7.9) | 158 (6.0) | 3053 (7.9) | 90 (5.4) | 2975 (7.7) | 106 (6.3) |
| Hip fracture history (%) | | | | | | | | | | | | | | | | |
| No | 38556 (100.0) | 1727 (100.0) | 38561 (100.0) | 1725 (99.9) | 39236 (100.0) | 1049 (99.9) | 39235 (100.0) | 1049 (100.0) | 37671 (100.0) | 2613 (100.0) | 37672 (100.0) | 2613 (100.0) | 38607 (100.0) | 1677 (99.9) | 38608 (100.0) | 1677 (100.0) |
| Ever >1 year before index date | 2 (0.0) | 0 (0.0) | 2 (0.0) | 1 (0.1) | 3 (0.0) | 1 (0.1) | 1 (0.0) | 0 (0.0) | 2 (0.0) | 1 (0.0) | 2 (0.0) | 0 (0.0) | 2 (0.0) | 1 (0.1) | 2 (0.0) | 0 (0.0) |
| 1 year before index | 9 (0.0) | 0 (0.0) | 4 (0.0) | 0 (0.0) | 5 (0.0) | 0 (0.0) | 8 (0.0) | 0 (0.0) | 7 (0.0) | 0 (0.0) | 6 (0.0) | 0 (0.0) | 7 (0.0) | 0 (0.0) | 6 (0.0) | 0 (0.0) |
| Shoulder fracture history (%) | | | | | | | | | | | | | | | | |
| No | 38408 (99.6) | 1721 (99.7) | 38411 (99.6) | 1718 (99.5) | 39090 (99.6) | 1042 (99.2) | 39079 (99.6) | 1047 (99.8) | 37535 (99.6) | 2605 (99.7) | 37516 (99.6) | 2602 (99.6) | 38466 (99.6) | 1671 (99.6) | 38450 (99.6) | 1671 (99.6) |
| Ever | 70 (0.2) | 2 (0.1) | 81 (0.2) | 6 (0.3) | 80 (0.2) | 5 (0.5) | 72 (0.2) | 2 (0.2) | 65 (0.2) | 5 (0.2) | 84 (0.2) | 5 (0.2) | 76 (0.2) | 5 (0.3) | 75 (0.2) | 3 (0.2) |
| 1 year before index | 89 (0.2) | 4 (0.2) | 75 (0.2) | 2 (0.1) | 74 (0.2) | 3 (0.3) | 93 (0.2) | 0 (0.0) | 80 (0.2) | 4 (0.2) | 80 (0.2) | 6 (0.2) | 74 (0.2) | 2 (0.1) | 91 (0.2) | 3 (0.2) |
| Spine fracture history (%) | | | | | | | | | | | | | | | | |
| No | 38099 (98.8) | 1700 (98.4) | 38134 (98.9) | 1692 (98.0) | 38789 (98.8) | 1032 (98.3) | 38778 (98.8) | 1026 (97.8) | 37259 (98.9) | 2570 (98.3) | 37226 (98.8) | 2570 (98.4) | 38158 (98.8) | 1649 (98.3) | 38169 (98.8) | 1649 (98.3) |
| Ever >1 year before index date | 304 (0.8) | 17 (1.0) | 282 (0.7) | 27 (1.6) | 300 (0.8) | 11 (1.0) | 301 (0.8) | 18 (1.7) | 278 (0.7) | 30 (1.1) | 288 (0.8) | 34 (1.3) | 295 (0.8) | 19 (1.1) | 294 (0.8) | 22 (1.3) |
| 1 year before index | 164 (0.4) | 10 (0.6) | 151 (0.4) | 7 (0.4) | 155 (0.4) | 7 (0.7) | 165 (0.4) | 5 (0.5) | 143 (0.4) | 14 (0.5) | 166 (0.4) | 9 (0.3) | 163 (0.4) | 10 (0.6) | 153 (0.4) | 6 (0.4) |
| Wrist fracture history (%) | | | | | | | | | | | | | | | | |
| No | 35865 (93.0) | 1637 (94.8) | 35951 (93.2) | 1649 (95.5) | 36603 (93.3) | 999 (95.1) | 36501 (93.0) | 999 (95.2) | 35019 (92.9) | 2480 (94.9) | 35131 (93.2) | 2472 (94.6) | 35913 (93.0) | 1585 (94.5) | 36018 (93.3) | 1586 (94.6) |
| Ever >1 year before index date | 1294 (3.4) | 56 (3.2) | 1243 (3.2) | 59 (3.4) | 1270 (3.2) | 34 (3.2) | 1316 (3.4) | 32 (3.1) | 1271 (3.4) | 82 (3.1) | 1203 (3.2) | 96 (3.7) | 1323 (3.4) | 56 (3.3) | 1216 (3.1) | 57 (3.4) |

| IFX cohort | One year | | | | | | | | Two years | | | | | | | |
|---|---|---|---|---|---|---|---|---|---|---|---|---|---|---|---|---|
| | MACE | | | | Stroke/MI | | | | MACE | | | | Stroke/MI | | | |
| | Development set (n=40294) | | Internal validation set (n=40293) | | Development set (n=40294) | | Internal validation set (n=40293) | | Development set (n=40294) | | Internal validation set (n=40293) | | Development set (n=40294) | | Internal validation set (n=40293) | |
| | Outcome=NO | Outcome=YES | Outcome=NO | Outcome=YES | Outcome=NO | Outcome=YES | Outcome=NO | Outcome=YES | Outcome=NO | Outcome=YES | Outcome=NO | Outcome=YES | Outcome=NO | Outcome=YES | Outcome=NO | Outcome=YES |
| 1 year before index | 1408 (3.7) | 34 (2.0) | 1373 (3.6) | 18 (1.0) | 1371 (3.5) | 17 (1.6) | 1427 (3.6) | 18 (1.7) | 1390 (3.7) | 52 (2.0) | 1346 (3.6) | 45 (1.7) | 1380 (3.6) | 37 (2.2) | 1382 (3.6) | 34 (2.0) |
| BMI** (%) | | | | | | | | | | | | | | | | |
| <18.5 | 1031 (2.7) | 56 (3.2) | 1043 (2.7) | 64 (3.7) | 1046 (2.7) | 35 (3.3) | 1082 (2.8) | 31 (3.0) | 1054 (2.8) | 83 (3.2) | 973 (2.6) | 84 (3.2) | 1034 (2.7) | 51 (3.0) | 1059 (2.7) | 50 (3.0) |
| 18.6 - 24.9 | 10390 (26.9) | 492 (28.5) | 10331 (26.8) | 465 (26.9) | 10570 (26.9) | 295 (28.1) | 10514 (26.8) | 299 (28.5) | 10146 (26.9) | 757 (29.0) | 10055 (26.7) | 720 (27.6) | 10427 (27.0) | 465 (27.7) | 10306 (26.7) | 480 (28.6) |
| 25 - 29.9 | 14180 (36.8) | 649 (37.6) | 14110 (36.6) | 626 (36.3) | 14408 (36.7) | 382 (36.4) | 14384 (36.7) | 391 (37.3) | 13750 (36.5) | 923 (35.3) | 13942 (37.0) | 950 (36.4) | 14240 (36.9) | 579 (34.5) | 14117 (36.6) | 629 (37.5) |
| 30 - 39.9 | 12697 (32.9) | 518 (30.0) | 12798 (33.2) | 550 (31.9) | 12930 (32.9) | 333 (31.7) | 12979 (33.1) | 321 (30.6) | 12456 (33.1) | 835 (31.9) | 12435 (33.0) | 837 (32.0) | 12643 (32.7) | 575 (34.3) | 12836 (33.2) | 509 (30.4) |
| >=40 | 269 (0.7) | 12 (0.7) | 285 (0.7) | 21 (1.2) | 290 (0.7) | 5 (0.5) | 285 (0.7) | 7 (0.7) | 274 (0.7) | 16 (0.6) | 275 (0.7) | 22 (0.8) | 272 (0.7) | 8 (0.5) | 298 (0.8) | 9 (0.5) |
| No. of GP visits** (%) | | | | | | | | | | | | | | | | |
| 0 | 8103 (21.0) | 242 (14.0) | 8074 (20.9) | 298 (17.3) | 8297 (21.1) | 181 (17.2) | 8071 (20.6) | 168 (16.0) | 7956 (21.1) | 466 (17.8) | 7857 (20.9) | 438 (16.8) | 8108 (21.0) | 284 (16.9) | 7989 (20.7) | 336 (20.0) |
| 1-5 | 8956 (23.2) | 364 (21.1) | 9069 (23.5) | 346 (20.0) | 9017 (23.0) | 189 (18.0) | 9305 (23.7) | 224 (21.4) | 8808 (23.4) | 522 (20.0) | 8851 (23.5) | 554 (21.2) | 8989 (23.3) | 329 (19.6) | 9082 (23.5) | 335 (20.0) |
| 6-10 | 8150 (21.1) | 386 (22.4) | 7902 (20.5) | 324 (18.8) | 8147 (20.8) | 230 (21.9) | 8182 (20.8) | 203 (19.4) | 7803 (20.7) | 525 (20.1) | 7883 (20.9) | 551 (21.1) | 7937 (20.6) | 353 (21.0) | 8144 (21.1) | 328 (19.6) |
| 11-15 | 5360 (13.9) | 247 (14.3) | 5352 (13.9) | 290 (16.8) | 5494 (14.0) | 156 (14.9) | 5436 (13.9) | 163 (15.5) | 5198 (13.8) | 399 (15.3) | 5240 (13.9) | 412 (15.8) | 5503 (14.3) | 252 (15.0) | 5236 (13.6) | 258 (15.4) |
| >=16 | 7998 (20.7) | 488 (28.3) | 8170 (21.2) | 468 (27.1) | 8289 (21.1) | 294 (28.0) | 8250 (21.0) | 291 (27.7) | 7915 (21.0) | 702 (26.9) | 7849 (20.8) | 658 (25.2) | 8079 (20.9) | 460 (27.4) | 8165 (21.1) | 420 (25.0) |
| No. of GP emergency visits** (%) | | | | | | | | | | | | | | | | |
| 0 | 30483 (79.0) | 1167 (67.6) | 30429 (78.9) | 1205 (69.8) | 30762 (78.4) | 753 (71.7) | 30996 (79.0) | 773 (73.7) | 29873 (79.3) | 1845 (70.6) | 29742 (78.9) | 1824 (69.8) | 30352 (78.6) | 1224 (72.9) | 30453 (78.9) | 1255 (74.8) |
| 1 | 3815 (9.9) | 230 (13.3) | 3783 (9.8) | 214 (12.4) | 3944 (10.0) | 138 (13.1) | 3835 (9.8) | 125 (11.9) | 3673 (9.7) | 320 (12.2) | 3717 (9.9) | 332 (12.7) | 3886 (10.1) | 198 (11.8) | 3754 (9.7) | 204 (12.2) |
| 2 | 1640 (4.3) | 123 (7.1) | 1659 (4.3) | 102 (5.9) | 1681 (4.3) | 61 (5.8) | 1719 (4.4) | 63 (6.0) | 1592 (4.2) | 150 (5.7) | 1604 (4.3) | 178 (6.8) | 1645 (4.3) | 89 (5.3) | 1695 (4.4) | 95 (5.7) |
| 3-5 | 1811 (4.7) | 131 (7.6) | 1842 (4.8) | 142 (8.2) | 1956 (5.0) | 55 (5.2) | 1849 (4.7) | 66 (6.3) | 1757 (4.7) | 193 (7.4) | 1789 (4.7) | 187 (7.2) | 1886 (4.9) | 109 (6.5) | 1847 (4.8) | 84 (5.0) |

| IFX cohort | One year | | | | | | | | Two years | | | | | | | |
|---|---|---|---|---|---|---|---|---|---|---|---|---|---|---|---|---|
| | MACE | | | | Stroke/MI | | | | MACE | | | | Stroke/MI | | | |
| | Development set (n=40294) | | Internal validation set (n=40293) | | Development set (n=40294) | | Internal validation set (n=40293) | | Development set (n=40294) | | Internal validation set (n=40293) | | Development set (n=40294) | | Internal validation set (n=40293) | |
| | Outcome=NO | Outcome=YES | Outcome=NO | Outcome=YES | Outcome=NO | Outcome=YES | Outcome=NO | Outcome=YES | Outcome=NO | Outcome=YES | Outcome=NO | Outcome=YES | Outcome=NO | Outcome=YES | Outcome=NO | Outcome=YES |
| >=6 | 818 (2.1) | 76 (4.4) | 854 (2.2) | 63 (3.7) | 901 (2.3) | 43 (4.1) | 845 (2.2) | 22 (2.1) | 785 (2.1) | 106 (4.1) | 828 (2.2) | 92 (3.5) | 847 (2.2) | 58 (3.5) | 867 (2.2) | 39 (2.3) |
| eGFR** (%) | | | | | | | | | | | | | | | | |
| <=29 | 1011 (2.6) | 98 (5.7) | 1043 (2.7) | 80 (4.6) | 1039 (2.6) | 47 (4.5) | 1090 (2.8) | 56 (5.3) | 1028 (2.7) | 107 (4.1) | 977 (2.6) | 120 (4.6) | 1046 (2.7) | 75 (4.5) | 1050 (2.7) | 61 (3.6) |
| 30 - 44 | 3828 (9.9) | 276 (16.0) | 3855 (10.0) | 269 (15.6) | 3932 (10.0) | 167 (15.9) | 3958 (10.1) | 171 (16.3) | 3792 (10.1) | 394 (15.1) | 3673 (9.7) | 369 (14.1) | 3890 (10.1) | 241 (14.4) | 3855 (10.0) | 242 (14.4) |
| 45 - 59 | 8527 (22.1) | 486 (28.1) | 8406 (21.8) | 482 (27.9) | 8624 (22.0) | 272 (25.9) | 8729 (22.2) | 276 (26.3) | 8289 (22.0) | 693 (26.5) | 8229 (21.8) | 690 (26.4) | 8653 (22.4) | 428 (25.5) | 8419 (21.8) | 401 (23.9) |
| 60 - 89 | 23541 (61.0) | 820 (47.5) | 23508 (61.0) | 845 (49.0) | 23877 (60.8) | 539 (51.3) | 23783 (60.6) | 515 (49.1) | 22917 (60.8) | 1352 (51.7) | 23100 (61.3) | 1345 (51.5) | 23340 (60.4) | 880 (52.4) | 23568 (61.0) | 926 (55.2) |
| >=90 | 1660 (4.3) | 47 (2.7) | 1755 (4.6) | 50 (2.9) | 1772 (4.5) | 25 (2.4) | 1684 (4.3) | 31 (3.0) | 1654 (4.4) | 68 (2.6) | 1701 (4.5) | 89 (3.4) | 1687 (4.4) | 54 (3.2) | 1724 (4.5) | 47 (2.8) |
| SBP** (%) | | | | | | | | | | | | | | | | |
| <120 | 5303 (13.8) | 208 (12.0) | 5320 (13.8) | 250 (14.5) | 5372 (13.7) | 133 (12.7) | 5457 (13.9) | 119 (11.3) | 5170 (13.7) | 334 (12.8) | 5244 (13.9) | 333 (12.7) | 5398 (14.0) | 220 (13.1) | 5291 (13.7) | 172 (10.3) |
| 120 - 139 | 15152 (39.3) | 594 (34.4) | 15148 (39.3) | 619 (35.9) | 15327 (39.1) | 348 (33.1) | 15469 (39.4) | 369 (35.2) | 14794 (39.3) | 884 (33.8) | 14880 (39.5) | 955 (36.5) | 15142 (39.2) | 551 (32.8) | 15216 (39.4) | 604 (36.0) |
| 140 - 159 | 13291 (34.5) | 621 (36.0) | 13188 (34.2) | 579 (33.5) | 13573 (34.6) | 390 (37.1) | 13351 (34.0) | 365 (34.8) | 13004 (34.5) | 943 (36.1) | 12851 (34.1) | 881 (33.7) | 13251 (34.3) | 606 (36.1) | 13231 (34.3) | 591 (35.2) |
| >=160 | 4821 (12.5) | 304 (17.6) | 4911 (12.7) | 278 (16.1) | 4972 (12.7) | 179 (17.0) | 4967 (12.7) | 196 (18.7) | 4712 (12.5) | 453 (17.3) | 4705 (12.5) | 444 (17.0) | 4825 (12.5) | 301 (17.9) | 4878 (12.6) | 310 (18.5) |
| DBP** (%) | | | | | | | | | | | | | | | | |
| <80 | 20630 (53.5) | 897 (51.9) | 20626 (53.5) | 991 (57.4) | 20941 (53.4) | 555 (52.9) | 21068 (53.7) | 580 (55.3) | 19984 (53.0) | 1442 (55.2) | 20277 (53.8) | 1441 (55.1) | 20689 (53.6) | 918 (54.7) | 20609 (53.4) | 928 (55.3) |
| 80 - 89 | 13615 (35.3) | 603 (34.9) | 13727 (35.6) | 551 (31.9) | 13904 (35.4) | 365 (34.8) | 13892 (35.4) | 335 (31.9) | 13513 (35.9) | 853 (32.6) | 13262 (35.2) | 868 (33.2) | 13681 (35.4) | 543 (32.4) | 13730 (35.6) | 542 (32.3) |
| 90 - 99 | 3418 (8.9) | 170 (9.8) | 3300 (8.6) | 146 (8.5) | 3482 (8.9) | 102 (9.7) | 3351 (8.5) | 99 (9.4) | 3303 (8.8) | 243 (9.3) | 3249 (8.6) | 239 (9.1) | 3334 (8.6) | 165 (9.8) | 3373 (8.7) | 162 (9.7) |
| >=100 | 904 (2.3) | 57 (3.3) | 914 (2.4) | 38 (2.2) | 917 (2.3) | 28 (2.7) | 933 (2.4) | 35 (3.3) | 880 (2.3) | 76 (2.9) | 892 (2.4) | 65 (2.5) | 912 (2.4) | 52 (3.1) | 904 (2.3) | 45 (2.7) |
| No. of concomitant medicines** (%) | | | | | | | | | | | | | | | | |

| IFX cohort | One year | | | | | | | | Two years | | | | | | | |
|---|---|---|---|---|---|---|---|---|---|---|---|---|---|---|---|---|
| | MACE | | | | Stroke/MI | | | | MACE | | | | Stroke/MI | | | |
| | Development set (n=40294) | | Internal validation set (n=40293) | | Development set (n=40294) | | Internal validation set (n=40293) | | Development set (n=40294) | | Internal validation set (n=40293) | | Development set (n=40294) | | Internal validation set (n=40293) | |
| | Outcome=NO | Outcome=YES | Outcome=NO | Outcome=YES | Outcome=NO | Outcome=YES | Outcome=NO | Outcome=YES | Outcome=NO | Outcome=YES | Outcome=NO | Outcome=YES | Outcome=NO | Outcome=YES | Outcome=NO | Outcome=YES |
| 0 | 8790 (22.8) | 261 (15.1) | 8784 (22.8) | 282 (16.3) | 8942 (22.8) | 178 (17.0) | 8823 (22.5) | 174 (16.6) | 8619 (22.9) | 478 (18.3) | 8580 (22.8) | 440 (16.8) | 8720 (22.6) | 291 (17.3) | 8766 (22.7) | 340 (20.3) |
| 1 - 3 | 6065 (15.7) | 179 (10.4) | 5998 (15.6) | 175 (10.1) | 6139 (15.6) | 109 (10.4) | 6074 (15.5) | 95 (9.1) | 5952 (15.8) | 262 (10.0) | 5904 (15.7) | 299 (11.4) | 6062 (15.7) | 164 (9.8) | 6006 (15.6) | 185 (11.0) |
| 4 - 6 | 7307 (18.9) | 323 (18.7) | 7170 (18.6) | 315 (18.3) | 7305 (18.6) | 174 (16.6) | 7439 (19.0) | 197 (18.8) | 7067 (18.8) | 463 (17.7) | 7087 (18.8) | 498 (19.1) | 7139 (18.5) | 315 (18.8) | 7380 (19.1) | 281 (16.8) |
| 7 - 9 | 6289 (16.3) | 309 (17.9) | 6376 (16.5) | 340 (19.7) | 6426 (16.4) | 201 (19.1) | 6505 (16.6) | 182 (17.3) | 6110 (16.2) | 475 (18.2) | 6231 (16.5) | 498 (19.1) | 6394 (16.6) | 299 (17.8) | 6316 (16.4) | 305 (18.2) |
| 10 - 12 | 4437 (11.5) | 281 (16.3) | 4428 (11.5) | 243 (14.1) | 4554 (11.6) | 158 (15.0) | 4523 (11.5) | 154 (14.7) | 4305 (11.4) | 408 (15.6) | 4319 (11.5) | 357 (13.7) | 4541 (11.8) | 259 (15.4) | 4369 (11.3) | 220 (13.1) |
| >=13 | 5679 (14.7) | 374 (21.7) | 5811 (15.1) | 371 (21.5) | 5878 (15.0) | 230 (21.9) | 5880 (15.0) | 247 (23.5) | 5627 (14.9) | 528 (20.2) | 5559 (14.8) | 521 (19.9) | 5760 (14.9) | 350 (20.9) | 5779 (15.0) | 346 (20.6) |
| Cholesterol measurement** (HDL/LDL) (%) | | | | | | | | | | | | | | | | |
| <=3.5 | 25209 (65.4) | 1134 (65.7) | 25332 (65.7) | 1106 (64.1) | 25886 (66.0) | 699 (66.6) | 25508 (65.0) | 688 (65.6) | 24772 (65.7) | 1651 (63.2) | 24662 (65.5) | 1696 (64.9) | 25339 (65.6) | 1077 (64.2) | 25292 (65.5) | 1073 (64.0) |
| 3.6 - 5 | 10597 (27.5) | 457 (26.5) | 10533 (27.3) | 490 (28.4) | 10628 (27.1) | 281 (26.8) | 10893 (27.8) | 275 (26.2) | 10263 (27.2) | 723 (27.7) | 10349 (27.5) | 742 (28.4) | 10586 (27.4) | 459 (27.4) | 10564 (27.4) | 468 (27.9) |
| >5 | 2761 (7.2) | 136 (7.9) | 2702 (7.0) | 130 (7.5) | 2730 (7.0) | 70 (6.7) | 2843 (7.2) | 86 (8.2) | 2645 (7.0) | 240 (9.2) | 2669 (7.1) | 175 (6.7) | 2691 (7.0) | 142 (8.5) | 2760 (7.1) | 136 (8.1) |
| No. of previous fractures* (%) | | | | | | | | | | | | | | | | |
| 0 | 20251 (52.5) | 943 (54.6) | 20300 (52.6) | 948 (54.9) | 20793 (53.0) | 547 (52.1) | 20568 (52.4) | 534 (50.9) | 19816 (52.6) | 1421 (54.4) | 19808 (52.6) | 1397 (53.5) | 20251 (52.4) | 857 (51.1) | 20441 (52.9) | 893 (53.2) |
| 1 | 12567 (32.6) | 537 (31.1) | 12442 (32.3) | 555 (32.2) | 12613 (32.1) | 347 (33.0) | 12792 (32.6) | 349 (33.3) | 12206 (32.4) | 820 (31.4) | 12233 (32.5) | 842 (32.2) | 12534 (32.5) | 554 (33.0) | 12489 (32.3) | 524 (31.2) |
| >=2 | 5749 (14.9) | 247 (14.3) | 5825 (15.1) | 223 (12.9) | 5838 (14.9) | 156 (14.9) | 5884 (15.0) | 166 (15.8) | 5658 (15.0) | 373 (14.3) | 5639 (15.0) | 374 (14.3) | 5831 (15.1) | 267 (15.9) | 5686 (14.7) | 260 (15.5) |

**Abbreviations**: OST, patients with incident diagnosis of osteoporosis; IFX, patients with incident fragility fracture; OBP, incident users of oral bisphosphonates; MACE, composite outcome for the occurrence of either myocardial infarction, stroke or cardiovascular disease death; MI, myocardial infarction; * ever; ** in the year prior to start; SES, socio-economic status; BMI, body mass index; eGFR, estimated Glomerular Filtration Rate; SBP, cholesterol, systolic blood pressure; DBP, diastolic blood pressure.

#### Table S2c Dataset split into train and test sets, stratified by outcome (OBP cohort)

| OBP cohort | One year | | | | | | | | Two years | | | | | | | |
|---|---|---|---|---|---|---|---|---|---|---|---|---|---|---|---|---|
| | MACE | | | | Stroke/MI | | | | MACE | | | | Stroke/MI | | | |
| | Development set (n=72980) | | Internal validation set (n=72979) | | Development set (n=72980) | | Internal validation set (n=72979) | | Development set (n=72980) | | Internal validation set (n=72979) | | Development set (n=72980) | | Internal validation set (n=72979) | |
| | Outcome=NO | Outcome=YES | Outcome=NO | Outcome=YES | Outcome=NO | Outcome=YES | Outcome=NO | Outcome=YES | Outcome=NO | Outcome=YES | Outcome=NO | Outcome=YES | Outcome=NO | Outcome=YES | Outcome=NO | Outcome=YES |
| n | 71168 | 1812 | 71168 | 1811 | 71547 | 1433 | 71546 | 1433 | 69745 | 3235 | 69744 | 3235 | 70427 | 2553 | 70427 | 2552 |
| Sex = Male (%) | 14170 (19.9) | 493 (27.2) | 14413 (20.3) | 471 (26.0) | 14336 (20.0) | 372 (26.0) | 14438 (20.2) | 401 (28.0) | 13869 (19.9) | 875 (27.0) | 13984 (20.1) | 819 (25.3) | 14206 (20.2) | 676 (26.5) | 13996 (19.9) | 669 (26.2) |
| SES (%) | | | | | | | | | | | | | | | | |
| 1 | 17629 (24.8) | 406 (22.4) | 17444 (24.5) | 358 (19.8) | 17665 (24.7) | 309 (21.6) | 17583 (24.6) | 280 (19.5) | 17238 (24.7) | 682 (21.1) | 17176 (24.6) | 741 (22.9) | 17254 (24.5) | 555 (21.7) | 17462 (24.8) | 566 (22.2) |
| 2 | 17462 (24.5) | 435 (24.0) | 17405 (24.5) | 436 (24.1) | 17541 (24.5) | 338 (23.6) | 17518 (24.5) | 341 (23.8) | 17125 (24.6) | 773 (23.9) | 17108 (24.5) | 732 (22.6) | 17302 (24.6) | 599 (23.5) | 17257 (24.5) | 580 (22.7) |
| 3 | 15052 (21.1) | 396 (21.9) | 15030 (21.1) | 399 (22.0) | 15098 (21.1) | 314 (21.9) | 15154 (21.2) | 311 (21.7) | 14656 (21.0) | 718 (22.2) | 14766 (21.2) | 737 (22.8) | 14845 (21.1) | 551 (21.6) | 14910 (21.2) | 571 (22.4) |
| 4 | 12771 (17.9) | 334 (18.4) | 13096 (18.4) | 365 (20.2) | 12935 (18.1) | 279 (19.5) | 13059 (18.3) | 293 (20.4) | 12702 (18.2) | 638 (19.7) | 12636 (18.1) | 590 (18.2) | 12870 (18.3) | 486 (19.0) | 12708 (18.0) | 502 (19.7) |
| 5 | 8194 (11.5) | 236 (13.0) | 8146 (11.4) | 252 (13.9) | 8251 (11.5) | 191 (13.3) | 8178 (11.4) | 208 (14.5) | 7966 (11.4) | 419 (13.0) | 8012 (11.5) | 431 (13.3) | 8097 (11.5) | 360 (14.1) | 8038 (11.4) | 333 (13.0) |
| Smoking** (%) | | | | | | | | | | | | | | | | |
| Ex | 24339 (34.2) | 626 (34.5) | 24602 (34.6) | 595 (32.9) | 24541 (34.3) | 486 (33.9) | 24624 (34.4) | 511 (35.7) | 24015 (34.4) | 1132 (35.0) | 23855 (34.2) | 1160 (35.9) | 24344 (34.6) | 922 (36.1) | 23984 (34.1) | 912 (35.7) |
| No | 37064 (52.1) | 984 (54.3) | 36785 (51.7) | 1005 (55.5) | 37102 (51.9) | 775 (54.1) | 37206 (52.0) | 755 (52.7) | 36192 (51.9) | 1688 (52.2) | 36262 (52.0) | 1696 (52.4) | 36489 (51.8) | 1306 (51.2) | 36744 (52.2) | 1299 (50.9) |
| Yes | 9765 (13.7) | 202 (11.1) | 9781 (13.7) | 211 (11.7) | 9904 (13.8) | 172 (12.0) | 9716 (13.6) | 167 (11.7) | 9538 (13.7) | 415 (12.8) | 9627 (13.8) | 379 (11.7) | 9594 (13.6) | 325 (12.7) | 9699 (13.8) | 341 (13.4) |
| Drinking** (%) | | | | | | | | | | | | | | | | |
| Ex | 3386 (4.8) | 115 (6.3) | 3613 (5.1) | 112 (6.2) | 3555 (5.0) | 80 (5.6) | 3506 (4.9) | 85 (5.9) | 3361 (4.8) | 243 (7.5) | 3400 (4.9) | 222 (6.9) | 3445 (4.9) | 178 (7.0) | 3440 (4.9) | 163 (6.4) |
| No | 20245 (28.4) | 642 (35.4) | 20254 (28.5) | 650 (35.9) | 20380 (28.5) | 513 (35.8) | 20406 (28.5) | 492 (34.3) | 19831 (28.4) | 1129 (34.9) | 19734 (28.3) | 1097 (33.9) | 20152 (28.6) | 850 (33.3) | 19923 (28.3) | 866 (33.9) |
| Yes | 47537 (66.8) | 1055 (58.2) | 47301 (66.5) | 1049 (57.9) | 47612 (66.5) | 840 (58.6) | 47634 (66.6) | 856 (59.7) | 46553 (66.7) | 1863 (57.6) | 46610 (66.8) | 1916 (59.2) | 46830 (66.5) | 1525 (59.7) | 47064 (66.8) | 1523 (59.7) |
| Diabetes type I*= 1 (%) | 154 (0.2) | 5 (0.3) | 134 (0.2) | 4 (0.2) | 152 (0.2) | 3 (0.2) | 137 (0.2) | 5 (0.3) | 133 (0.2) | 12 (0.4) | 147 (0.2) | 5 (0.2) | 135 (0.2) | 6 (0.2) | 151 (0.2) | 5 (0.2) |

| OBP cohort | One year | | | | | | | | Two years | | | | | | | |
|---|---|---|---|---|---|---|---|---|---|---|---|---|---|---|---|---|
| | MACE | | | | Stroke/MI | | | | MACE | | | | Stroke/MI | | | |
| | Development set (n=72980) | | Internal validation set (n=72979) | | Development set (n=72980) | | Internal validation set (n=72979) | | Development set (n=72980) | | Internal validation set (n=72979) | | Development set (n=72980) | | Internal validation set (n=72979) | |
| | Outcome=NO | Outcome=YES | Outcome=NO | Outcome=YES | Outcome=NO | Outcome=YES | Outcome=NO | Outcome=YES | Outcome=NO | Outcome=YES | Outcome=NO | Outcome=YES | Outcome=NO | Outcome=YES | Outcome=NO | Outcome=YES |
| Diabetes type II*= 1 (%) | 4024 (5.7) | 157 (8.7) | 4008 (5.6) | 151 (8.3) | 4006 (5.6) | 128 (8.9) | 4065 (5.7) | 141 (9.8) | 3971 (5.7) | 286 (8.8) | 3832 (5.5) | 251 (7.8) | 3970 (5.6) | 217 (8.5) | 3921 (5.6) | 232 (9.1) |
| Chronic obstructive pulmonary disease*= 1 (%) | 5443 (7.6) | 171 (9.4) | 5481 (7.7) | 156 (8.6) | 5590 (7.8) | 125 (8.7) | 5383 (7.5) | 153 (10.7) | 5332 (7.6) | 286 (8.8) | 5355 (7.7) | 278 (8.6) | 5447 (7.7) | 231 (9.0) | 5330 (7.6) | 243 (9.5) |
| Chronic kidney disease*= 1 (%) | 6490 (9.1) | 243 (13.4) | 6408 (9.0) | 255 (14.1) | 6490 (9.1) | 207 (14.4) | 6498 (9.1) | 201 (14.0) | 6325 (9.1) | 442 (13.7) | 6183 (8.9) | 446 (13.8) | 6371 (9.0) | 376 (14.7) | 6301 (8.9) | 348 (13.6) |
| Rheumatoid arthritis*= 1 (%) | 9616 (13.5) | 191 (10.5) | 9747 (13.7) | 192 (10.6) | 9658 (13.5) | 177 (12.4) | 9739 (13.6) | 172 (12.0) | 9623 (13.8) | 379 (11.7) | 9357 (13.4) | 387 (12.0) | 9440 (13.4) | 331 (13.0) | 9626 (13.7) | 349 (13.7) |
| Lupus*= 1 (%) | 182 (0.3) | 1 (0.1) | 143 (0.2) | 2 (0.1) | 157 (0.2) | 0 (0.0) | 168 (0.2) | 3 (0.2) | 166 (0.2) | 4 (0.1) | 155 (0.2) | 3 (0.1) | 154 (0.2) | 3 (0.1) | 167 (0.2) | 4 (0.2) |
| Systemic heart disease**= 1 (%) | 2578 (3.6) | 15 (0.8) | 2610 (3.7) | 19 (1.0) | 2600 (3.6) | 15 (1.0) | 2593 (3.6) | 14 (1.0) | 2488 (3.6) | 44 (1.4) | 2654 (3.8) | 36 (1.1) | 2571 (3.7) | 32 (1.3) | 2580 (3.7) | 39 (1.5) |
| Heparin use**= 1 (%) | 504 (0.7) | 9 (0.5) | 513 (0.7) | 9 (0.5) | 526 (0.7) | 7 (0.5) | 494 (0.7) | 8 (0.6) | 461 (0.7) | 18 (0.6) | 532 (0.8) | 24 (0.7) | 495 (0.7) | 20 (0.8) | 508 (0.7) | 12 (0.5) |
| Beta-blocker use**= 1 (%) | 11046 (15.5) | 401 (22.1) | 10904 (15.3) | 402 (22.2) | 10994 (15.4) | 357 (24.9) | 11057 (15.5) | 345 (24.1) | 10492 (15.0) | 738 (22.8) | 10836 (15.5) | 687 (21.2) | 10662 (15.1) | 612 (24.0) | 10863 (15.4) | 616 (24.1) |
| Hypertension**= 1 (%) | 4614 (6.5) | 121 (6.7) | 4651 (6.5) | 128 (7.1) | 4709 (6.6) | 105 (7.3) | 4588 (6.4) | 112 (7.8) | 4501 (6.5) | 228 (7.0) | 4536 (6.5) | 249 (7.7) | 4555 (6.5) | 210 (8.2) | 4552 (6.5) | 197 (7.7) |
| Deep vein thrombosis or pulmonary embolism**= 1 (%) | 672 (0.9) | 16 (0.9) | 624 (0.9) | 22 (1.2) | 666 (0.9) | 19 (1.3) | 638 (0.9) | 11 (0.8) | 648 (0.9) | 33 (1.0) | 619 (0.9) | 34 (1.1) | 672 (1.0) | 32 (1.3) | 612 (0.9) | 18 (0.7) |
| Anticoagulant use**= 1 (%) | 3719 (5.2) | 136 (7.5) | 3548 (5.0) | 143 (7.9) | 3658 (5.1) | 123 (8.6) | 3664 (5.1) | 101 (7.0) | 3490 (5.0) | 262 (8.1) | 3534 (5.1) | 260 (8.0) | 3612 (5.1) | 209 (8.2) | 3524 (5.0) | 201 (7.9) |
| Antidepressants TCA**= 1 (%) | 6980 (9.8) | 172 (9.5) | 7117 (10.0) | 182 (10.0) | 7027 (9.8) | 156 (10.9) | 7104 (9.9) | 164 (11.4) | 6938 (9.9) | 328 (10.1) | 6870 (9.9) | 315 (9.7) | 6961 (9.9) | 282 (11.0) | 6927 (9.8) | 281 (11.0) |
| Antidepressants SSRI**= 1 (%) | 6098 (8.6) | 197 (10.9) | 6149 (8.6) | 207 (11.4) | 6133 (8.6) | 151 (10.5) | 6215 (8.7) | 152 (10.6) | 5958 (8.5) | 341 (10.5) | 5989 (8.6) | 363 (11.2) | 6044 (8.6) | 273 (10.7) | 6083 (8.6) | 251 (9.8) |
| Hypercholesterolemia**= 1 (%) | 1292 (1.8) | 27 (1.5) | 1200 (1.7) | 24 (1.3) | 1257 (1.8) | 20 (1.4) | 1243 (1.7) | 23 (1.6) | 1230 (1.8) | 43 (1.3) | 1223 (1.8) | 47 (1.5) | 1232 (1.7) | 37 (1.4) | 1239 (1.8) | 35 (1.4) |
| Statin use**= 1 (%) | 16557 (23.3) | 532 (29.4) | 16373 (23.0) | 526 (29.0) | 16554 (23.1) | 424 (29.6) | 16535 (23.1) | 475 (33.1) | 16086 (23.1) | 962 (29.7) | 15997 (22.9) | 943 (29.1) | 16186 (23.0) | 788 (30.9) | 16207 (23.0) | 807 (31.6) |
| Osteoporosis history* = 1 | 22732 (31.9) | 417 (23.0) | 22661 (31.8) | 405 (22.4) | 22827 (31.9) | 351 (24.5) | 22681 (31.7) | 356 (24.8) | 22330 (32.0) | 738 (22.8) | 22364 (32.1) | 783 (24.2) | 22511 (32.0) | 637 (25.0) | 22424 (31.8) | 643 (25.2) |

| OBP cohort | One year | | | | | | | | Two years | | | | | | | |
|---|---|---|---|---|---|---|---|---|---|---|---|---|---|---|---|---|
| | MACE | | | | Stroke/MI | | | | MACE | | | | Stroke/MI | | | |
| | Development set (n=72980) | | Internal validation set (n=72979) | | Development set (n=72980) | | Internal validation set (n=72979) | | Development set (n=72980) | | Internal validation set (n=72979) | | Development set (n=72980) | | Internal validation set (n=72979) | |
| | Outcome=NO | Outcome=YES | Outcome=NO | Outcome=YES | Outcome=NO | Outcome=YES | Outcome=NO | Outcome=YES | Outcome=NO | Outcome=YES | Outcome=NO | Outcome=YES | Outcome=NO | Outcome=YES | Outcome=NO | Outcome=YES |
| Family history of cardiovascular disease = 1 (%) | 6478 (9.1) | 119 (6.6) | 6476 (9.1) | 131 (7.2) | 6462 (9.0) | 104 (7.3) | 6516 (9.1) | 122 (8.5) | 6326 (9.1) | 236 (7.3) | 6403 (9.2) | 239 (7.4) | 6455 (9.2) | 207 (8.1) | 6331 (9.0) | 211 (8.3) |
| Family history of cardiovascular disease before age 60= 1 (%) | 89 (0.1) | 0 (0.0) | 77 (0.1) | 4 (0.2) | 98 (0.1) | 1 (0.1) | 69 (0.1) | 2 (0.1) | 77 (0.1) | 1 (0.0) | 89 (0.1) | 3 (0.1) | 94 (0.1) | 2 (0.1) | 73 (0.1) | 1 (0.0) |
| Heart failure*= 1 (%) | 2415 (3.4) | 130 (7.2) | 2369 (3.3) | 126 (7.0) | 2428 (3.4) | 105 (7.3) | 2393 (3.3) | 114 (8.0) | 2327 (3.3) | 218 (6.7) | 2274 (3.3) | 221 (6.8) | 2378 (3.4) | 172 (6.7) | 2312 (3.3) | 178 (7.0) |
| Migraine*= 1 (%) | 9678 (13.6) | 221 (12.2) | 9832 (13.8) | 230 (12.7) | 9690 (13.5) | 194 (13.5) | 9860 (13.8) | 217 (15.1) | 9627 (13.8) | 412 (12.7) | 9539 (13.7) | 383 (11.8) | 9648 (13.7) | 354 (13.9) | 9611 (13.6) | 348 (13.6) |
| Severe mental illness*= 1 (%) | 9365 (13.2) | 230 (12.7) | 9437 (13.3) | 224 (12.4) | 9537 (13.3) | 182 (12.7) | 9348 (13.1) | 189 (13.2) | 9219 (13.2) | 411 (12.7) | 9203 (13.2) | 423 (13.1) | 9418 (13.4) | 334 (13.1) | 9168 (13.0) | 336 (13.2) |
| Vascular Disease*= 1 (%) | 832 (1.2) | 56 (3.1) | 841 (1.2) | 36 (2.0) | 830 (1.2) | 40 (2.8) | 859 (1.2) | 36 (2.5) | 773 (1.1) | 80 (2.5) | 833 (1.2) | 79 (2.4) | 796 (1.1) | 67 (2.6) | 841 (1.2) | 61 (2.4) |
| Atrial fibrillation*= 1 (%) | 3855 (5.4) | 203 (11.2) | 3750 (5.3) | 214 (11.8) | 3773 (5.3) | 171 (11.9) | 3908 (5.5) | 170 (11.9) | 3684 (5.3) | 343 (10.6) | 3643 (5.2) | 352 (10.9) | 3796 (5.4) | 255 (10.0) | 3665 (5.2) | 306 (12.0) |
| On anti-hypertensive drug= 1 (%) | 38540 (54.2) | 1157 (63.9) | 38219 (53.7) | 1128 (62.3) | 38610 (54.0) | 948 (66.2) | 38516 (53.8) | 970 (67.7) | 37252 (53.4) | 2043 (63.2) | 37645 (54.0) | 2104 (65.0) | 37816 (53.7) | 1739 (68.1) | 37815 (53.7) | 1674 (65.6) |
| Antipsychotic use**= 1 (%) | 361 (0.5) | 22 (1.2) | 379 (0.5) | 11 (0.6) | 372 (0.5) | 7 (0.5) | 386 (0.5) | 8 (0.6) | 370 (0.5) | 23 (0.7) | 354 (0.5) | 26 (0.8) | 394 (0.6) | 10 (0.4) | 357 (0.5) | 12 (0.5) |
| Steroid use**= 1 (%) | 18111 (25.4) | 445 (24.6) | 18181 (25.5) | 464 (25.6) | 18267 (25.5) | 379 (26.4) | 18159 (25.4) | 396 (27.6) | 17760 (25.5) | 842 (26.0) | 17820 (25.6) | 779 (24.1) | 17947 (25.5) | 686 (26.9) | 17883 (25.4) | 685 (26.8) |
| Erectile dysfunction**= 1 (%) | 1376 (1.9) | 40 (2.2) | 1343 (1.9) | 38 (2.1) | 1383 (1.9) | 41 (2.9) | 1342 (1.9) | 31 (2.2) | 1332 (1.9) | 68 (2.1) | 1323 (1.9) | 74 (2.3) | 1331 (1.9) | 60 (2.4) | 1340 (1.9) | 66 (2.6) |
| Age>75 | 37245 (52.3) | 1465 (80.8) | 37009 (52.0) | 1480 (81.7) | 37455 (52.4) | 1132 (79.0) | 37487 (52.4) | 1125 (78.5) | 36108 (51.8) | 2566 (79.3) | 35902 (51.5) | 2623 (81.1) | 36707 (52.1) | 1992 (78.0) | 36519 (51.9) | 1981 (77.6) |
| Age Group (%) | | | | | | | | | | | | | | | | |
| 50-59 | 8098 (11.4) | 43 (2.4) | 8200 (11.5) | 47 (2.6) | 8142 (11.4) | 37 (2.6) | 8164 (11.4) | 45 (3.1) | 8116 (11.6) | 78 (2.4) | 8116 (11.6) | 78 (2.4) | 8100 (11.5) | 68 (2.7) | 8147 (11.6) | 73 (2.9) |
| 60-69 | 15607 (21.9) | 153 (8.4) | 15748 (22.1) | 157 (8.7) | 15719 (22.0) | 129 (9.0) | 15676 (21.9) | 141 (9.8) | 15461 (22.2) | 296 (9.1) | 15632 (22.4) | 276 (8.5) | 15527 (22.0) | 253 (9.9) | 15641 (22.2) | 244 (9.6) |
| 70-79 | 22289 (31.3) | 465 (25.7) | 22485 (31.6) | 440 (24.3) | 22402 (31.3) | 383 (26.7) | 22496 (31.4) | 398 (27.8) | 21985 (31.5) | 876 (27.1) | 21995 (31.5) | 823 (25.4) | 22035 (31.3) | 702 (27.5) | 22187 (31.5) | 755 (29.6) |

| OBP cohort | One year | | | | | | | | Two years | | | | | | | |
|---|---|---|---|---|---|---|---|---|---|---|---|---|---|---|---|---|
| | MACE | | | | Stroke/MI | | | | MACE | | | | Stroke/MI | | | |
| | Development set (n=72980) | | Internal validation set (n=72979) | | Development set (n=72980) | | Internal validation set (n=72979) | | Development set (n=72980) | | Internal validation set (n=72979) | | Development set (n=72980) | | Internal validation set (n=72979) | |
| | Outcome=NO | Outcome=YES | Outcome=NO | Outcome=YES | Outcome=NO | Outcome=YES | Outcome=NO | Outcome=YES | Outcome=NO | Outcome=YES | Outcome=NO | Outcome=YES | Outcome=NO | Outcome=YES | Outcome=NO | Outcome=YES |
| 80-89 | 20318 (28.5) | 847 (46.7) | 20028 (28.1) | 860 (47.5) | 20437 (28.6) | 687 (47.9) | 20284 (28.4) | 645 (45.0) | 19621 (28.1) | 1467 (45.3) | 19407 (27.8) | 1558 (48.2) | 19976 (28.4) | 1203 (47.1) | 19721 (28.0) | 1153 (45.2) |
| >89 | 4856 (6.8) | 304 (16.8) | 4707 (6.6) | 307 (17.0) | 4847 (6.8) | 197 (13.7) | 4926 (6.9) | 204 (14.2) | 4562 (6.5) | 518 (16.0) | 4594 (6.6) | 500 (15.5) | 4789 (6.8) | 327 (12.8) | 4731 (6.7) | 327 (12.8) |
| Charlson score (%) | | | | | | | | | | | | | | | | |
| 0 | 39525 (55.5) | 912 (50.3) | 39354 (55.3) | 926 (51.1) | 39531 (55.3) | 682 (47.6) | 39821 (55.7) | 683 (47.7) | 38612 (55.4) | 1640 (50.7) | 38847 (55.7) | 1618 (50.0) | 39020 (55.4) | 1227 (48.1) | 39251 (55.7) | 1219 (47.8) |
| 1 | 15190 (21.3) | 340 (18.8) | 15256 (21.4) | 346 (19.1) | 15345 (21.4) | 294 (20.5) | 15213 (21.3) | 280 (19.5) | 14961 (21.5) | 648 (20.0) | 14878 (21.3) | 645 (19.9) | 15033 (21.3) | 525 (20.6) | 15030 (21.3) | 544 (21.3) |
| 2 | 8803 (12.4) | 243 (13.4) | 8848 (12.4) | 247 (13.6) | 8938 (12.5) | 200 (14.0) | 8789 (12.3) | 214 (14.9) | 8709 (12.5) | 405 (12.5) | 8566 (12.3) | 461 (14.3) | 8798 (12.5) | 373 (14.6) | 8616 (12.2) | 354 (13.9) |
| ≥3 | 7650 (10.7) | 317 (17.5) | 7710 (10.8) | 292 (16.1) | 7733 (10.8) | 257 (17.9) | 7723 (10.8) | 256 (17.9) | 7463 (10.7) | 542 (16.8) | 7453 (10.7) | 511 (15.8) | 7576 (10.8) | 428 (16.8) | 7530 (10.7) | 435 (17.0) |
| Cardiovascular disease (%) | | | | | | | | | | | | | | | | |
| No | 62945 (88.4) | 1389 (76.7) | 62804 (88.2) | 1378 (76.1) | 63095 (88.2) | 1061 (74.0) | 63297 (88.5) | 1063 (74.2) | 61818 (88.6) | 2495 (77.1) | 61689 (88.5) | 2514 (77.7) | 62317 (88.5) | 1938 (75.9) | 62338 (88.5) | 1923 (75.4) |
| Ever >1 year before index date | 6397 (9.0) | 289 (15.9) | 6487 (9.1) | 301 (16.6) | 6545 (9.1) | 253 (17.7) | 6422 (9.0) | 254 (17.7) | 6176 (8.9) | 535 (16.5) | 6263 (9.0) | 500 (15.5) | 6272 (8.9) | 453 (17.7) | 6317 (9.0) | 432 (16.9) |
| 1 year before index | 728 (1.0) | 46 (2.5) | 745 (1.0) | 32 (1.8) | 792 (1.1) | 30 (2.1) | 692 (1.0) | 37 (2.6) | 685 (1.0) | 71 (2.2) | 729 (1.0) | 66 (2.0) | 745 (1.1) | 61 (2.4) | 695 (1.0) | 50 (2.0) |
| 6 months before index | 804 (1.1) | 57 (3.1) | 834 (1.2) | 68 (3.8) | 823 (1.2) | 61 (4.3) | 829 (1.2) | 50 (3.5) | 764 (1.1) | 98 (3.0) | 799 (1.1) | 102 (3.2) | 799 (1.1) | 68 (2.7) | 792 (1.1) | 104 (4.1) |
| 1 month before index | 294 (0.4) | 31 (1.7) | 298 (0.4) | 32 (1.8) | 292 (0.4) | 28 (2.0) | 306 (0.4) | 29 (2.0) | 302 (0.4) | 36 (1.1) | 264 (0.4) | 53 (1.6) | 294 (0.4) | 33 (1.3) | 285 (0.4) | 43 (1.7) |
| MI or Stroke (%) | | | | | | | | | | | | | | | | |
| No | 66305 (93.2) | 1407 (77.6) | 66192 (93.0) | 1431 (79.0) | 66458 (92.9) | 1161 (81.0) | 66585 (93.1) | 1131 (78.9) | 65070 (93.3) | 2589 (80.0) | 65106 (93.3) | 2570 (79.4) | 65613 (93.2) | 2083 (81.6) | 65565 (93.1) | 2074 (81.3) |
| Ever >1 year before index date | 3472 (4.9) | 235 (13.0) | 3550 (5.0) | 224 (12.4) | 3624 (5.1) | 162 (11.3) | 3518 (4.9) | 177 (12.4) | 3399 (4.9) | 399 (12.3) | 3271 (4.7) | 412 (12.7) | 3390 (4.8) | 304 (11.9) | 3497 (5.0) | 290 (11.4) |
| 1 year before index | 1391 (2.0) | 170 (9.4) | 1426 (2.0) | 156 (8.6) | 1465 (2.0) | 110 (7.7) | 1443 (2.0) | 125 (8.7) | 1276 (1.8) | 247 (7.6) | 1367 (2.0) | 253 (7.8) | 1424 (2.0) | 166 (6.5) | 1365 (1.9) | 188 (7.4) |
| Established CVD *= Ever (%) | 8724 (12.3) | 586 (32.3) | 8890 (12.5) | 574 (31.7) | 9049 (12.6) | 432 (30.1) | 8847 (12.4) | 446 (31.1) | 8383 (12.0) | 988 (30.5) | 8432 (12.1) | 971 (30.0) | 8570 (12.2) | 733 (28.7) | 8725 (12.4) | 746 (29.2) |

| OBP cohort | One year | | | | | | | | Two years | | | | | | | |
|---|---|---|---|---|---|---|---|---|---|---|---|---|---|---|---|---|
| | MACE | | | | Stroke/MI | | | | MACE | | | | Stroke/MI | | | |
| | Development set (n=72980) | | Internal validation set (n=72979) | | Development set (n=72980) | | Internal validation set (n=72979) | | Development set (n=72980) | | Internal validation set (n=72979) | | Development set (n=72980) | | Internal validation set (n=72979) | |
| | Outcome=NO | Outcome=YES | Outcome=NO | Outcome=YES | Outcome=NO | Outcome=YES | Outcome=NO | Outcome=YES | Outcome=NO | Outcome=YES | Outcome=NO | Outcome=YES | Outcome=NO | Outcome=YES | Outcome=NO | Outcome=YES |
| Any fracture history (%) | | | | | | | | | | | | | | | | |
| No | 55890 (78.5) | 1347 (74.3) | 55766 (78.4) | 1383 (76.4) | 56065 (78.4) | 1076 (75.1) | 56182 (78.5) | 1063 (74.2) | 54753 (78.5) | 2449 (75.7) | 54757 (78.5) | 2427 (75.0) | 55277 (78.5) | 1895 (74.2) | 55268 (78.5) | 1946 (76.3) |
| Ever >1 year before index date | 5128 (7.2) | 139 (7.7) | 5197 (7.3) | 134 (7.4) | 5167 (7.2) | 113 (7.9) | 5198 (7.3) | 120 (8.4) | 5022 (7.2) | 258 (8.0) | 5085 (7.3) | 233 (7.2) | 5082 (7.2) | 204 (8.0) | 5114 (7.3) | 198 (7.8) |
| 1 year before index | 10150 (14.3) | 326 (18.0) | 10205 (14.3) | 294 (16.2) | 10315 (14.4) | 244 (17.0) | 10166 (14.2) | 250 (17.4) | 9970 (14.3) | 528 (16.3) | 9902 (14.2) | 575 (17.8) | 10068 (14.3) | 454 (17.8) | 10045 (14.3) | 408 (16.0) |
| Hip fracture history (%) | | | | | | | | | | | | | | | | |
| No | 66623 (93.6) | 1637 (90.3) | 66709 (93.7) | 1641 (90.6) | 66951 (93.6) | 1297 (90.5) | 67050 (93.7) | 1312 (91.6) | 65416 (93.8) | 2927 (90.5) | 65351 (93.7) | 2916 (90.1) | 66012 (93.7) | 2307 (90.4) | 65959 (93.7) | 2332 (91.4) |
| Ever >1 year before index date | 808 (1.1) | 34 (1.9) | 844 (1.2) | 34 (1.9) | 843 (1.2) | 32 (2.2) | 822 (1.1) | 23 (1.6) | 794 (1.1) | 66 (2.0) | 804 (1.2) | 56 (1.7) | 799 (1.1) | 46 (1.8) | 833 (1.2) | 42 (1.6) |
| 1 year before index | 3737 (5.3) | 141 (7.8) | 3615 (5.1) | 136 (7.5) | 3753 (5.2) | 104 (7.3) | 3674 (5.1) | 98 (6.8) | 3535 (5.1) | 242 (7.5) | 3589 (5.1) | 263 (8.1) | 3616 (5.1) | 200 (7.8) | 3635 (5.2) | 178 (7.0) |
| Shoulder fracture history (%) | | | | | | | | | | | | | | | | |
| No | 70809 (99.5) | 1800 (99.3) | 70787 (99.5) | 1803 (99.6) | 71214 (99.5) | 1421 (99.2) | 71140 (99.4) | 1424 (99.4) | 69362 (99.5) | 3221 (99.6) | 69405 (99.5) | 3211 (99.3) | 70063 (99.5) | 2532 (99.2) | 70066 (99.5) | 2538 (99.5) |
| Ever >1 year before index date | 168 (0.2) | 3 (0.2) | 159 (0.2) | 3 (0.2) | 146 (0.2) | 2 (0.1) | 182 (0.3) | 3 (0.2) | 164 (0.2) | 5 (0.2) | 154 (0.2) | 10 (0.3) | 162 (0.2) | 8 (0.3) | 158 (0.2) | 5 (0.2) |
| 1 year before index | 191 (0.3) | 9 (0.5) | 222 (0.3) | 5 (0.3) | 187 (0.3) | 10 (0.7) | 224 (0.3) | 6 (0.4) | 219 (0.3) | 9 (0.3) | 185 (0.3) | 14 (0.4) | 202 (0.3) | 13 (0.5) | 203 (0.3) | 9 (0.4) |
| Spine fracture history (%) | | | | | | | | | | | | | | | | |
| No | 70030 (98.4) | 1776 (98.0) | 69977 (98.3) | 1771 (97.8) | 70385 (98.4) | 1400 (97.7) | 70368 (98.4) | 1401 (97.8) | 68591 (98.3) | 3180 (98.3) | 68622 (98.4) | 3161 (97.7) | 69274 (98.4) | 2496 (97.8) | 69280 (98.4) | 2504 (98.1) |
| Ever >1 year before index date | 173 (0.2) | 6 (0.3) | 169 (0.2) | 8 (0.4) | 164 (0.2) | 6 (0.4) | 180 (0.3) | 6 (0.4) | 161 (0.2) | 7 (0.2) | 176 (0.3) | 12 (0.4) | 175 (0.2) | 12 (0.5) | 164 (0.2) | 5 (0.2) |
| 1 year before index | 965 (1.4) | 30 (1.7) | 1022 (1.4) | 32 (1.8) | 998 (1.4) | 27 (1.9) | 998 (1.4) | 26 (1.8) | 993 (1.4) | 48 (1.5) | 946 (1.4) | 62 (1.9) | 978 (1.4) | 45 (1.8) | 983 (1.4) | 43 (1.7) |
| Wrist fracture history (%) | | | | | | | | | | | | | | | | |
| No | 66880 (94.0) | 1713 (94.5) | 66803 (93.9) | 1722 (95.1) | 67187 (93.9) | 1351 (94.3) | 67225 (94.0) | 1355 (94.6) | 65473 (93.9) | 3059 (94.6) | 65524 (93.9) | 3062 (94.7) | 66106 (93.9) | 2388 (93.5) | 66203 (94.0) | 2421 (94.9) |
| Ever >1 year before index date | 1963 (2.8) | 46 (2.5) | 1987 (2.8) | 44 (2.4) | 1949 (2.7) | 42 (2.9) | 2014 (2.8) | 35 (2.4) | 1985 (2.8) | 89 (2.8) | 1886 (2.7) | 80 (2.5) | 2000 (2.8) | 83 (3.3) | 1899 (2.7) | 58 (2.3) |

| OBP cohort | One year | | | | | | | | Two years | | | | | | | |
|---|---|---|---|---|---|---|---|---|---|---|---|---|---|---|---|---|
| | MACE | | | | Stroke/MI | | | | MACE | | | | Stroke/MI | | | |
| | Development set (n=72980) | | Internal validation set (n=72979) | | Development set (n=72980) | | Internal validation set (n=72979) | | Development set (n=72980) | | Internal validation set (n=72979) | | Development set (n=72980) | | Internal validation set (n=72979) | |
| | Outcome=NO | Outcome=YES | Outcome=NO | Outcome=YES | Outcome=NO | Outcome=YES | Outcome=NO | Outcome=YES | Outcome=NO | Outcome=YES | Outcome=NO | Outcome=YES | Outcome=NO | Outcome=YES | Outcome=NO | Outcome=YES |
| 1 year before index | 2325 (3.3) | 53 (2.9) | 2378 (3.3) | 45 (2.5) | 2411 (3.4) | 40 (2.8) | 2307 (3.2) | 43 (3.0) | 2287 (3.3) | 87 (2.7) | 2334 (3.3) | 93 (2.9) | 2321 (3.3) | 82 (3.2) | 2325 (3.3) | 73 (2.9) |
| BMI** (%) | | | | | | | | | | | | | | | | |
| <18.5 | 5194 (7.3) | 248 (13.7) | 5206 (7.3) | 229 (12.6) | 5269 (7.4) | 160 (11.2) | 5288 (7.4) | 160 (11.2) | 4995 (7.2) | 357 (11.0) | 5144 (7.4) | 381 (11.8) | 5135 (7.3) | 234 (9.2) | 5239 (7.4) | 269 (10.5) |
| 18.6 - 24.9 | 32370 (45.5) | 902 (49.8) | 32352 (45.5) | 918 (50.7) | 32519 (45.5) | 704 (49.1) | 32649 (45.6) | 670 (46.8) | 31650 (45.4) | 1616 (50.0) | 31653 (45.4) | 1623 (50.2) | 32017 (45.5) | 1194 (46.8) | 32076 (45.5) | 1255 (49.2) |
| 25 - 29.9 | 21582 (30.3) | 429 (23.7) | 21631 (30.4) | 467 (25.8) | 21755 (30.4) | 373 (26.0) | 21571 (30.1) | 410 (28.6) | 21257 (30.5) | 833 (25.7) | 21202 (30.4) | 817 (25.3) | 21354 (30.3) | 735 (28.8) | 21347 (30.3) | 673 (26.4) |
| 30 - 39.9 | 11060 (15.5) | 218 (12.0) | 10987 (15.4) | 187 (10.3) | 11024 (15.4) | 186 (13.0) | 11064 (15.5) | 178 (12.4) | 10872 (15.6) | 400 (12.4) | 10789 (15.5) | 391 (12.1) | 10971 (15.6) | 368 (14.4) | 10784 (15.3) | 329 (12.9) |
| >=40 | 962 (1.4) | 15 (0.8) | 992 (1.4) | 10 (0.6) | 980 (1.4) | 10 (0.7) | 974 (1.4) | 15 (1.0) | 971 (1.4) | 29 (0.9) | 956 (1.4) | 23 (0.7) | 950 (1.3) | 22 (0.9) | 981 (1.4) | 26 (1.0) |
| No. of GP visits** (%) | | | | | | | | | | | | | | | | |
| 0 | 7330 (10.3) | 259 (14.3) | 7350 (10.3) | 238 (13.1) | 7338 (10.3) | 167 (11.7) | 7512 (10.5) | 160 (11.2) | 7287 (10.4) | 441 (13.6) | 7054 (10.1) | 395 (12.2) | 7359 (10.4) | 256 (10.0) | 7255 (10.3) | 307 (12.0) |
| 1-5 | 17739 (24.9) | 385 (21.2) | 17686 (24.9) | 403 (22.3) | 17828 (24.9) | 281 (19.6) | 17815 (24.9) | 289 (20.2) | 17315 (24.8) | 701 (21.7) | 17484 (25.1) | 713 (22.0) | 17498 (24.8) | 540 (21.2) | 17666 (25.1) | 509 (19.9) |
| 6-10 | 15800 (22.2) | 303 (16.7) | 15913 (22.4) | 328 (18.1) | 16021 (22.4) | 260 (18.1) | 15792 (22.1) | 271 (18.9) | 15625 (22.4) | 585 (18.1) | 15539 (22.3) | 595 (18.4) | 15576 (22.1) | 468 (18.3) | 15798 (22.4) | 502 (19.7) |
| 11-15 | 12035 (16.9) | 282 (15.6) | 11878 (16.7) | 271 (15.0) | 12149 (17.0) | 229 (16.0) | 11870 (16.6) | 218 (15.2) | 11756 (16.9) | 487 (15.1) | 11700 (16.8) | 523 (16.2) | 11690 (16.6) | 436 (17.1) | 11951 (17.0) | 389 (15.2) |
| >=16 | 18264 (25.7) | 583 (32.2) | 18341 (25.8) | 571 (31.5) | 18211 (25.5) | 496 (34.6) | 18557 (25.9) | 495 (34.5) | 17762 (25.5) | 1021 (31.6) | 17967 (25.8) | 1009 (31.2) | 18304 (26.0) | 853 (33.4) | 17757 (25.2) | 845 (33.1) |
| No. of GP emergency visits** (%) | | | | | | | | | | | | | | | | |
| 0 | 58545 (82.3) | 1296 (71.5) | 58536 (82.3) | 1321 (72.9) | 58771 (82.1) | 1033 (72.1) | 58859 (82.3) | 1035 (72.2) | 57400 (82.3) | 2384 (73.7) | 57541 (82.5) | 2373 (73.4) | 57934 (82.3) | 1877 (73.5) | 58001 (82.4) | 1886 (73.9) |
| 1 | 6791 (9.5) | 232 (12.8) | 6853 (9.6) | 207 (11.4) | 6870 (9.6) | 186 (13.0) | 6855 (9.6) | 172 (12.0) | 6713 (9.6) | 373 (11.5) | 6596 (9.5) | 401 (12.4) | 6807 (9.7) | 308 (12.1) | 6657 (9.5) | 311 (12.2) |
| 2 | 2581 (3.6) | 98 (5.4) | 2500 (3.5) | 103 (5.7) | 2588 (3.6) | 89 (6.2) | 2523 (3.5) | 82 (5.7) | 2435 (3.5) | 183 (5.7) | 2491 (3.6) | 173 (5.3) | 2466 (3.5) | 150 (5.9) | 2523 (3.6) | 143 (5.6) |
| 3-5 | 2401 (3.4) | 108 (6.0) | 2373 (3.3) | 111 (6.1) | 2427 (3.4) | 66 (4.6) | 2405 (3.4) | 95 (6.6) | 2312 (3.3) | 190 (5.9) | 2306 (3.3) | 185 (5.7) | 2347 (3.3) | 142 (5.6) | 2362 (3.4) | 142 (5.6) |

| OBP cohort | One year | | | | | | | | Two years | | | | | | | |
|---|---|---|---|---|---|---|---|---|---|---|---|---|---|---|---|---|
| | MACE | | | | Stroke/MI | | | | MACE | | | | Stroke/MI | | | |
| | Development set (n=72980) | | Internal validation set (n=72979) | | Development set (n=72980) | | Internal validation set (n=72979) | | Development set (n=72980) | | Internal validation set (n=72979) | | Development set (n=72980) | | Internal validation set (n=72979) | |
| | Outcome=NO | Outcome=YES | Outcome=NO | Outcome=YES | Outcome=NO | Outcome=YES | Outcome=NO | Outcome=YES | Outcome=NO | Outcome=YES | Outcome=NO | Outcome=YES | Outcome=NO | Outcome=YES | Outcome=NO | Outcome=YES |
| >=6 | 850 (1.2) | 78 (4.3) | 906 (1.3) | 69 (3.8) | 891 (1.2) | 59 (4.1) | 904 (1.3) | 49 (3.4) | 885 (1.3) | 105 (3.2) | 810 (1.2) | 103 (3.2) | 873 (1.2) | 76 (3.0) | 884 (1.3) | 70 (2.7) |
| eGFR** (%) | | | | | | | | | | | | | | | | |
| <=29 | 871 (1.2) | 39 (2.2) | 825 (1.2) | 42 (2.3) | 841 (1.2) | 31 (2.2) | 869 (1.2) | 36 (2.5) | 801 (1.1) | 69 (2.1) | 826 (1.2) | 81 (2.5) | 823 (1.2) | 55 (2.2) | 829 (1.2) | 70 (2.7) |
| 30 - 44 | 4700 (6.6) | 206 (11.4) | 4631 (6.5) | 212 (11.7) | 4755 (6.6) | 178 (12.4) | 4648 (6.5) | 168 (11.7) | 4540 (6.5) | 389 (12.0) | 4417 (6.3) | 403 (12.5) | 4527 (6.4) | 333 (13.0) | 4601 (6.5) | 288 (11.3) |
| 45 - 59 | 14678 (20.6) | 513 (28.3) | 14590 (20.5) | 509 (28.1) | 14826 (20.7) | 421 (29.4) | 14640 (20.5) | 403 (28.1) | 14359 (20.6) | 883 (27.3) | 14170 (20.3) | 878 (27.1) | 14481 (20.6) | 730 (28.6) | 14384 (20.4) | 695 (27.2) |
| 60 - 89 | 47272 (66.4) | 999 (55.1) | 47572 (66.8) | 995 (54.9) | 47447 (66.3) | 763 (53.2) | 47845 (66.9) | 783 (54.6) | 46485 (66.6) | 1795 (55.5) | 46787 (67.1) | 1771 (54.7) | 47021 (66.8) | 1361 (53.3) | 47038 (66.8) | 1418 (55.6) |
| >=90 | 3647 (5.1) | 55 (3.0) | 3550 (5.0) | 53 (2.9) | 3678 (5.1) | 40 (2.8) | 3544 (5.0) | 43 (3.0) | 3560 (5.1) | 99 (3.1) | 3544 (5.1) | 102 (3.2) | 3575 (5.1) | 74 (2.9) | 3575 (5.1) | 81 (3.2) |
| SBP** (%) | | | | | | | | | | | | | | | | |
| <120 | 9053 (12.7) | 222 (12.3) | 9071 (12.7) | 203 (11.2) | 9057 (12.7) | 162 (11.3) | 9157 (12.8) | 173 (12.1) | 8988 (12.9) | 358 (11.1) | 8839 (12.7) | 364 (11.3) | 8842 (12.6) | 256 (10.0) | 9168 (13.0) | 283 (11.1) |
| 120 - 139 | 28165 (39.6) | 642 (35.4) | 28219 (39.7) | 659 (36.4) | 28382 (39.7) | 500 (34.9) | 28294 (39.5) | 509 (35.5) | 27539 (39.5) | 1200 (37.1) | 27821 (39.9) | 1125 (34.8) | 28015 (39.8) | 922 (36.1) | 27831 (39.5) | 917 (35.9) |
| 140 - 159 | 25143 (35.3) | 676 (37.3) | 25126 (35.3) | 644 (35.6) | 25230 (35.3) | 520 (36.3) | 25312 (35.4) | 527 (36.8) | 24602 (35.3) | 1161 (35.9) | 24611 (35.3) | 1215 (37.6) | 25088 (35.6) | 974 (38.2) | 24614 (34.9) | 913 (35.8) |
| >=160 | 8807 (12.4) | 272 (15.0) | 8752 (12.3) | 305 (16.8) | 8878 (12.4) | 251 (17.5) | 8783 (12.3) | 224 (15.6) | 8616 (12.4) | 516 (16.0) | 8473 (12.1) | 531 (16.4) | 8482 (12.0) | 401 (15.7) | 8814 (12.5) | 439 (17.2) |
| DBP** (%) | | | | | | | | | | | | | | | | |
| <80 | 36582 (51.4) | 993 (54.8) | 36535 (51.3) | 1022 (56.4) | 36689 (51.3) | 798 (55.7) | 36858 (51.5) | 787 (54.9) | 35762 (51.3) | 1766 (54.6) | 35797 (51.3) | 1807 (55.9) | 36117 (51.3) | 1400 (54.8) | 36222 (51.4) | 1393 (54.6) |
| 80 - 89 | 26337 (37.0) | 605 (33.4) | 26284 (36.9) | 572 (31.6) | 26469 (37.0) | 468 (32.7) | 26398 (36.9) | 463 (32.3) | 25697 (36.8) | 1099 (34.0) | 25943 (37.2) | 1059 (32.7) | 26145 (37.1) | 877 (34.4) | 25931 (36.8) | 845 (33.1) |
| 90 - 99 | 6592 (9.3) | 157 (8.7) | 6758 (9.5) | 171 (9.4) | 6718 (9.4) | 124 (8.7) | 6695 (9.4) | 141 (9.8) | 6663 (9.6) | 290 (9.0) | 6447 (9.2) | 278 (8.6) | 6607 (9.4) | 204 (8.0) | 6617 (9.4) | 250 (9.8) |
| >=100 | 1657 (2.3) | 57 (3.1) | 1591 (2.2) | 46 (2.5) | 1671 (2.3) | 43 (3.0) | 1595 (2.2) | 42 (2.9) | 1623 (2.3) | 80 (2.5) | 1557 (2.2) | 91 (2.8) | 1558 (2.2) | 72 (2.8) | 1657 (2.4) | 64 (2.5) |
| No. of concomitant medicines** (%) | | | | | | | | | | | | | | | | |

| OBP cohort | One year | | | | | | | | Two years | | | | | | | |
|---|---|---|---|---|---|---|---|---|---|---|---|---|---|---|---|---|
| | MACE | | | | Stroke/MI | | | | MACE | | | | Stroke/MI | | | |
| | Development set (n=72980) | | Internal validation set (n=72979) | | Development set (n=72980) | | Internal validation set (n=72979) | | Development set (n=72980) | | Internal validation set (n=72979) | | Development set (n=72980) | | Internal validation set (n=72979) | |
| | Outcome=NO | Outcome=YES | Outcome=NO | Outcome=YES | Outcome=NO | Outcome=YES | Outcome=NO | Outcome=YES | Outcome=NO | Outcome=YES | Outcome=NO | Outcome=YES | Outcome=NO | Outcome=YES | Outcome=NO | Outcome=YES |
| 0 | 10025 (14.1) | 317 (17.5) | 10026 (14.1) | 294 (16.2) | 9996 (14.0) | 205 (14.3) | 10260 (14.3) | 201 (14.0) | 9911 (14.2) | 553 (17.1) | 9708 (13.9) | 490 (15.1) | 9962 (14.1) | 336 (13.2) | 9986 (14.2) | 378 (14.8) |
| 1 - 3 | 13032 (18.3) | 197 (10.9) | 13232 (18.6) | 225 (12.4) | 13213 (18.5) | 156 (10.9) | 13157 (18.4) | 160 (11.2) | 13013 (18.7) | 358 (11.1) | 12918 (18.5) | 397 (12.3) | 13035 (18.5) | 289 (11.3) | 13074 (18.6) | 288 (11.3) |
| 4 - 6 | 14290 (20.1) | 254 (14.0) | 14329 (20.1) | 252 (13.9) | 14363 (20.1) | 211 (14.7) | 14363 (20.1) | 188 (13.1) | 14041 (20.1) | 452 (14.0) | 14126 (20.3) | 506 (15.6) | 14092 (20.0) | 398 (15.6) | 14250 (20.2) | 385 (15.1) |
| 7 - 9 | 12823 (18.0) | 332 (18.3) | 12597 (17.7) | 297 (16.4) | 12869 (18.0) | 252 (17.6) | 12670 (17.7) | 258 (18.0) | 12454 (17.9) | 564 (17.4) | 12443 (17.8) | 588 (18.2) | 12628 (17.9) | 464 (18.2) | 12507 (17.8) | 450 (17.6) |
| 10 - 12 | 9184 (12.9) | 276 (15.2) | 9106 (12.8) | 302 (16.7) | 9140 (12.8) | 241 (16.8) | 9251 (12.9) | 236 (16.5) | 8878 (12.7) | 531 (16.4) | 8966 (12.9) | 493 (15.2) | 9017 (12.8) | 420 (16.5) | 9016 (12.8) | 415 (16.3) |
| >=13 | 11814 (16.6) | 436 (24.1) | 11878 (16.7) | 441 (24.4) | 11966 (16.7) | 368 (25.7) | 11845 (16.6) | 390 (27.2) | 11448 (16.4) | 777 (24.0) | 11583 (16.6) | 761 (23.5) | 11693 (16.6) | 646 (25.3) | 11594 (16.5) | 636 (24.9) |
| Cholesterol measurement** (HDL/LDL) (%) | | | | | | | | | | | | | | | | |
| <=3.5 | 42161 (59.2) | 1040 (57.4) | 41958 (59.0) | 1088 (60.1) | 42183 (59.0) | 850 (59.3) | 42367 (59.2) | 847 (59.1) | 41201 (59.1) | 1949 (60.2) | 41217 (59.1) | 1880 (58.1) | 41788 (59.3) | 1525 (59.7) | 41410 (58.8) | 1524 (59.7) |
| 3.6 - 5 | 22710 (31.9) | 602 (33.2) | 22794 (32.0) | 548 (30.3) | 22896 (32.0) | 446 (31.1) | 22867 (32.0) | 445 (31.1) | 22336 (32.0) | 986 (30.5) | 22311 (32.0) | 1021 (31.6) | 22364 (31.8) | 757 (29.7) | 22748 (32.3) | 785 (30.8) |
| >5 | 6297 (8.8) | 170 (9.4) | 6416 (9.0) | 175 (9.7) | 6468 (9.0) | 137 (9.6) | 6312 (8.8) | 141 (9.8) | 6208 (8.9) | 300 (9.3) | 6216 (8.9) | 334 (10.3) | 6275 (8.9) | 271 (10.6) | 6269 (8.9) | 243 (9.5) |
| No. of previous fractures* (%) | | | | | | | | | | | | | | | | |
| 0 | 54809 (77.0) | 1323 (73.0) | 54649 (76.8) | 1339 (73.9) | 55028 (76.9) | 1044 (72.9) | 55008 (76.9) | 1040 (72.6) | 53668 (76.9) | 2392 (73.9) | 53681 (77.0) | 2379 (73.5) | 54207 (77.0) | 1849 (72.4) | 54160 (76.9) | 1904 (74.6) |
| 1 | 8772 (12.3) | 256 (14.1) | 8835 (12.4) | 266 (14.7) | 8802 (12.3) | 215 (15.0) | 8899 (12.4) | 213 (14.9) | 8597 (12.3) | 453 (14.0) | 8622 (12.4) | 457 (14.1) | 8704 (12.4) | 364 (14.3) | 8693 (12.3) | 368 (14.4) |
| >=2 | 7587 (10.7) | 233 (12.9) | 7684 (10.8) | 206 (11.4) | 7717 (10.8) | 174 (12.1) | 7639 (10.7) | 180 (12.6) | 7480 (10.7) | 390 (12.1) | 7441 (10.7) | 399 (12.3) | 7516 (10.7) | 340 (13.3) | 7574 (10.8) | 280 (11.0) |

**Abbreviations:** OST, patients with incident diagnosis of osteoporosis; IFX, patients with incident fragility fracture; OBP, incident users of oral bisphosphonates; MACE, composite outcome for the occurrence of either myocardial infarction, stroke or cardiovascular disease death; MI, myocardial infarction; * ever; ** in the year prior to start; SES, socio-economic status; BMI, body mass index; eGFR, estimated Glomerular Filtration Rate; SBP, cholesterol, systolic blood pressure; DBP, diastolic blood pressure.

**Table S3 Model Equations of one-year MACE (risk factors selected by lasso regression)**

| Predictor | OST Beta coefficients | IFX Beta coefficients | OBP Beta coefficients |
|---|---|---|---|
| **Intercept** | -5.214 | -5.396 | -4.416 |
| **Sex = Male (%)** | 0.479 | 0.369 | 0.349 |
| **SES (%)** | x | | |
| **1** | x | x | ref |
| **2** | x | x | 0.160 |
| **3** | x | x | 0.248 |
| **4** | x | x | 0.293 |
| **5** | x | x | 0.256 |
| **Smoking**** | | x | |
| **Ex** | ref | x | ref |
| **No** | -0.070 | x | -0.016 |
| **Yes** | 0.353 | x | 0.134 |
| **Drinking**** | | x | |
| **Ex** | ref | ref | ref |
| **No** | 0.171 | 0.044 | 0.025 |
| **Yes** | -0.170 | -0.053 | -0.084 |
| **Diabetes type I*** | x | x | x |
| **Diabetes type II*** | x | x | 0.101 |
| **Chronic obstructive pulmonary disease*** | x | x | 0.048 |
| **Chronic kidney disease*** | x | x | -0.216 |
| **Rheumatoid arthritis*** | x | x | -0.127 |
| **Lupus*** | x | x | x |
| **Systemic heart disease**** | x | x | -0.479 |
| **Anti-osteoporosis use**** | x | x | x |
| **Heparin use**** | x | x | x |
| **Beta-blocker use**** | x | x | 0.149 |
| **Hypertension**** | x | x | x |
| **Deep vein thrombosis or pulmonary embolism**** | x | x | x |
| **Anticoagulant use**** | x | x | -0.078 |

| Predictor | OST<br>Beta coefficients | IFX<br>Beta coefficients | OBP<br>Beta coefficients |
|---|---|---|---|
| **Antidepressants TCA**** | x | x | x |
| **Antidepressants SSRI**** | x | | 0.243 |
| **Hypercholesterolemia**** | x | x | x |
| **Statin use**** | x | | x |
| **Osteoporosis history*** | x | x | -0.301 |
| **Family history of cardiovascular disease (%)** | x | x | -0.058 |
| **Family history of cardiovascular disease before age 60** | x | x | x |
| **Heart failure*** | x | 0.157 | 0.031 |
| **Migraine*** | x | x | x |
| **Severe mental illness*** | x | x | x |
| **Vascular Disease*** | X | x | x |
| **Atrial fibrillation*** | 0.478 | 0.113 | 0.258 |
| **On anti-hypertensive drug** | 0.203 | 0.263 | 0.047 |
| **Antipsychotic use**** | x | x | x |
| **Steroid use**** | x | x | x |
| **Erectile dysfunction**** | x | x | x |
| **Age Group (%)** | | | |
| **50-59** | ref | ref | ref |
| **60-69** | 0.311 | 1.195 | 0.457 |
| **70-79** | 1.122 | 1.683 | 0.899 |
| **80-89** | 1.589 | 2.146 | 1.449 |
| **>89** | 2.065 | 2.387 | 1.756 |
| **Charlson score** | x | | |
| **0** | x | ref | ref |
| **1** | x | 0.008 | 0.012 |
| **2** | x | -0.087 | -0.059 |
| **≥3** | x | -0.009 | 0.055 |
| **Cardiovascular disease** | x | x | |
| **No** | x | x | ref |
| **Ever >1 year before index date** | x | x | 0.152 |
| **1 year before index** | x | x | 0.166 |

| Predictor | OST<br>Beta coefficients | IFX<br>Beta coefficients | OBP<br>Beta coefficients |
|---|---|---|---|
| **6 months before index** | x | x | 0.420 |
| **1 month before index** | x | x | 0.781 |
| **MI or Stroke** | | | |
| **No** | ref | ref | ref |
| **Ever >1 year before index date** | -0.009 | 0.350 | 0.290 |
| **1 year before index** | 0.709 | 0.741 | 0.925 |
| **Established CVD *** | 0.643 | 0.303 | 0.401 |
| **Any fracture history** | x | x | x |
| **No** | x | x | x |
| **Ever >1 year before index date** | x | x | x |
| **1 year before index** | x | x | x |
| **Hip fracture history** | x | x | x |
| **No** | x | x | x |
| **Ever >1 year before index date** | x | x | x |
| **1 year before index** | x | x | x |
| **Shoulder fracture history** | x | x | x |
| **No** | x | x | x |
| **Ever >1 year before index date** | x | x | x |
| **1 year before index** | x | x | x |
| **Spine fracture history** | x | x | x |
| **No** | x | x | x |
| **Ever >1 year before index date** | x | x | x |
| **1 year before index** | x | x | x |
| **Wrist fracture history** | x | x | x |
| **No** | x | x | x |
| **Ever >1 year before index date** | x | x | x |
| **1 year before index** | x | x | x |
| **BMI**** | | x | |

| Predictor | OST<br>Beta coefficients | IFX<br>Beta coefficients | OBP<br>Beta coefficients |
|---|---|---|---|
| **<18.5** | ref | x | ref |
| **18.6 - 24.9** | -0.334 | x | -0.302 |
| **25 - 29.9** | -0.581 | x | -0.546 |
| **30 - 39.9** | -0.879 | x | -0.601 |
| **>=40** | -0.421 | x | -0.946 |
| **No. of GP visits**** | | | |
| **0** | ref | ref | ref |
| **1-5** | 0.009 | -0.118 | -0.109 |
| **6-10** | -0.063 | -0.207 | -0.243 |
| **11-15** | -0.056 | -0.110 | -0.252 |
| **>=16** | 0.064 | -0.157 | -0.146 |
| **No. of GP emergency visits**** | x | | |
| **0** | x | ref | ref |
| **1** | x | 0.158 | 0.143 |
| **2** | x | 0.307 | 0.339 |
| **3-5** | x | 0.114 | 0.384 |
| **>=6** | x | 0.271 | 0.928 |
| **eGFR**** | | | |
| **<=29** | ref | ref | ref |
| **30 – 44** | 0.101 | -0.023 | -0.089 |
| **45 – 59** | -0.112 | -0.233 | -0.190 |
| **60 – 89** | -0.280 | -0.348 | -0.541 |
| **>=90** | -0.236 | -0.438 | -0.687 |
| **SBP**** | | | |
| **<120** | ref | ref | ref |
| **120 - 139** | 0.175 | 0.054 | 0.090 |
| **140 - 159** | 0.278 | 0.080 | 0.216 |
| **>=160** | 0.274 | 0.308 | 0.329 |
| **DBP**** | x | | |
| **<80** | x | ref | ref |
| **80 - 89** | x | 0.047 | -0.058 |
| **90 - 99** | x | 0.222 | 0.050 |
| **>=100** | x | 0.101 | 0.271 |
| **No. of concomitant medicines**** | | | |

| Predictor | OST<br>Beta coefficients | IFX<br>Beta coefficients | OBP<br>Beta coefficients |
|---|---|---|---|
| **0** | ref | ref | ref |
| **1 – 3** | -0.377 | 0.070 | -0.142 |
| **4 – 6** | -0.183 | 0.137 | -0.287 |
| **7 – 9** | 0.008 | 0.268 | -0.128 |
| **10 – 12** | 0.111 | 0.313 | -0.001 |
| **>=13** | 0.105 | 0.288 | -0.020 |
| **Cholesterol measurement** (HDL/LDL)** | x | x | |
| **<=3.5** | x | x | ref |
| **3.6 – 5** | x | x | 0.180 |
| **>5** | x | x | 0.299 |
| **No. of previous fractures*** | | | x |
| **0** | ref | ref | x |
| **1** | 0.217 | -0.090 | x |
| **>=2** | -0.008 | -0.119 | x |
| **Abbreviations**: OST, patients with incident diagnosis of osteoporosis; IFX, patients with incident fragility fracture; OBP, incident users of oral bisphosphonates; OR, odds ratio; CI, confidence intervals; MACE, composite outcome for the occurrence of either myocardial infarction, stroke or cardiovascular disease death; * ever; ** in the year prior to start; SES, socio-economic status; MI, myocardial infarction; BMI, body mass index; eGFR, estimated Glomerular Filtration Rate; SBP, cholesterol, systolic blood pressure; DBP, diastolic blood pressure. | | | |

#### Table S4a. Predictors of 2-year MACE models (risk factors selected by lasso regression)

| Predictor | OST<br>OR (95%CI) | IFX<br>OR (95%CI) | OBP<br>OR (95%CI) |
|---|---|---|---|
| **Sex = Male (%)** | 1.53 (1.3, 1.81) | 1.36 (1.22, 1.51) | 1.41 (1.28, 1.55) |
| **SES (%)** | | x | x |
| **1** | ref | x | x |
| **2** | 0.96 (0.8, 1.14) | x | x |
| **3** | 0.99 (0.83, 1.19) | x | x |
| **4** | 0.89 (0.73, 1.08) | x | x |
| **5** | 0.97 (0.78, 1.2) | x | x |
| **Smoking**** | | x | |
| **Ex** | ref | x | ref |

| Predictor | OST<br>OR (95%CI) | IFX<br>OR (95%CI) | OBP<br>OR (95%CI) |
|---|---|---|---|
| **No** | 1 (0.83, 1.2) | x | 0.94 (0.82, 1.09) |
| **Yes** | 1.25 (1, 1.56) | x | 1.15 (0.94, 1.4) |
| **Drinking**** | | | |
| **Ex** | ref | ref | ref |
| **No** | 1.03 (0.73, 1.45) | 0.89 (0.7, 1.12) | 0.88 (0.67, 1.15) |
| **Yes** | 0.89 (0.6, 1.31) | 0.75 (0.6, 0.95) | 0.76 (0.58, 0.98) |
| **Diabetes type I*** | x | x | x |
| **Diabetes type II*** | x | x | 1.32 (1.14, 1.52) |
| **Chronic obstructive pulmonary disease*** | x | x | x |
| **Chronic kidney disease*** | x | x | x |
| **Rheumatoid arthritis*** | x | x | 0.86 (0.76, 0.97) |
| **Lupus*** | x | x | x |
| **Systemic heart disease**** | x | x | x |
| **Anti-osteoporosis use**** | x | x | x |
| **Heparin use**** | x | x | x |
| **Beta-blocker use**** | 1.04 (0.89, 1.21) | 1.17 (1.04, 1.32) | 1.15 (1.04, 1.26) |
| **Hypertension**** | x | x | x |
| **Deep vein thrombosis or pulmonary embolism**** | x | x | x |
| **Anticoagulant use**** | x | x | x |
| **Antidepressants TCA**** | x | x | x |
| **Antidepressants SSRI**** | 1.38 (1.15, 1.66) | x | x |
| **Hypercholesterolemia**** | x | x | x |
| **Statin use**** | x | 0.91 (0.76, 1.09) | x |
| **Osteoporosis history*** | x | x | 0.81 (0.74, 0.88) |
| **Family history of cardiovascular disease** | x | x | x |
| **Family history of cardiovascular disease before age 60** | x | x | x |
| **Heart failure*** | x | x | x |
| **Migraine*** | x | x | x |
| **Severe mental illness*** | x | x | x |
| **Vascular Disease*** | x | x | x |
| **Atrial fibrillation*** | 1.15 (0.94, 1.4) | 1.07 (0.93, 1.23) | 1.35 (1.19, 1.53) |
| **On anti-hypertensive drug** | 1.3 (1.08, 1.55) | 1.22 (1.08, 1.38) | x |
| **Antipsychotic use**** | x | x | x |
| **Steroid use**** | x | | x |
| **Erectile dysfunction**** | x | x | x |
| **Age Group (%)** | | | |
| **50-59** | ref | ref | ref |

| Predictor | OST<br>OR (95%CI) | IFX<br>OR (95%CI) | OBP<br>OR (95%CI) |
|---|---|---|---|
| 60-69 | 1.36 (0.94, 1.98) | 2.72 (1.82, 4.06) | 1.88 (1.45, 2.44) |
| 70-79 | 2.63 (1.85, 3.74) | 4.46 (3.02, 6.6) | 3.5 (2.73, 4.5) |
| 80-89 | 4.45 (3.1, 6.38) | 7.37 (4.92, 11.05) | 5.81 (4.49, 7.51) |
| >89 | 6.54 (4.36, 9.82) | 7.72 (5.01, 11.89) | 7.97 (6.02, 10.56) |
| Charlson score | | x | |
| 0 | ref | x | ref |
| 1 | 1.21 (1.03, 1.42) | x | 0.98 (0.88, 1.1) |
| 2 | 0.91 (0.75, 1.11) | x | 0.87 (0.77, 1) |
| ≥3 | 1.07 (0.87, 1.31) | x | 0.89 (0.77, 1.04) |
| Cardiovascular disease | x | x | |
| No | x | x | ref |
| Ever >1 year before index date | x | x | 1.05 (0.94, 1.19) |
| 1 year before index | x | x | 1.11 (0.84, 1.45) |
| 6 months before index | x | x | 1.31 (1.03, 1.66) |
| 1 month before index | x | x | 1.86 (1.32, 2.62) |
| MI or Stroke | | | |
| No | ref | ref | ref |
| Ever >1 year before index date | 1.16 (0.89, 1.51) | 1.26 (1.06, 1.49) | 1.3 (1.11, 1.52) |
| 1 year before index | 2.04 (1.49, 2.79) | 1.66 (1.35, 2.04) | 1.87 (1.55, 2.26) |
| Established CVD * | 1.65 (1.34, 2.04) | 1.53 (1.33, 1.76) | 1.68 (1.47, 1.92) |
| Any fracture history | x | x | x |
| No | x | x | x |
| Ever >1 year before index date | x | x | x |
| 1 year before index | x | x | x |
| Hip fracture history | x | x | x |
| No | x | x | x |
| Ever >1 year before index date | x | x | x |
| 1 year before index | x | x | x |
| Shoulder fracture history | x | x | x |
| No | x | x | x |
| Ever >1 year before index date | x | x | x |
| 1 year before index | x | x | x |
| Spine fracture history | x | x | x |
| No | x | x | x |
| Ever >1 year before index date | x | x | x |
| 1 year before index | x | x | x |
| Wrist fracture history | x | x | x |
| No | x | x | x |
| Ever >1 year before index date | x | x | x |

| Predictor | OST<br>OR (95%CI) | IFX<br>OR (95%CI) | OBP<br>OR (95%CI) |
|---|---|---|---|
| **1 year before index** | x | x | x |
| **BMI**** | | x | |
| **<18.5** | ref | x | ref |
| **18.6 - 24.9** | 0.76 (0.56, 1.03) | x | 0.79 (0.66, 0.96) |
| **25 - 29.9** | 0.65 (0.44, 0.94) | x | 0.66 (0.51, 0.85) |
| **30 - 39.9** | 0.59 (0.36, 0.97) | x | 0.61 (0.45, 0.81) |
| **>=40** | 0.76 (0.32, 1.79) | x | 0.62 (0.37, 1.04) |
| **No. of GP visits**** | | | |
| **0** | ref | ref | ref |
| **1-5** | 0.49 (0.35, 0.69) | 0.92 (0.73, 1.16) | 1 (0.86, 1.17) |
| **6-10** | 0.53 (0.37, 0.75) | 0.79 (0.61, 1.02) | 0.89 (0.74, 1.08) |
| **11-15** | 0.49 (0.34, 0.7) | 0.82 (0.63, 1.07) | 0.86 (0.71, 1.04) |
| **>=16** | 0.55 (0.38, 0.78) | 0.81 (0.62, 1.05) | 0.93 (0.77, 1.13) |
| **No. of GP emergency visits**** | | | x |
| **0** | ref | ref | x |
| **1** | 1.19 (0.99, 1.43) | 1.16 (1.02, 1.32) | x |
| **2** | 1.32 (1.03, 1.7) | 1.4 (1.18, 1.66) | x |
| **3-5** | 1.17 (0.9, 1.52) | 1.2 (1.01, 1.42) | x |
| **>=6** | 1.4 (0.98, 2.01) | 1.35 (1.08, 1.69) | x |
| **eGFR**** | | | x |
| **<=29** | ref | ref | ref |
| **30 – 44** | 1.02 (0.57, 1.84) | 1 (0.73, 1.36) | 0.92 (0.66, 1.29) |
| **45 – 59** | 0.83 (0.47, 1.47) | 0.88 (0.6, 1.3) | 0.78 (0.51, 1.18) |
| **60 – 89** | 0.65 (0.37, 1.15) | 0.86 (0.59, 1.25) | 0.63 (0.39, 1.03) |
| **>=90** | 0.67 (0.33, 1.36) | 0.83 (0.47, 1.45) | 0.61 (0.33, 1.12) |
| **SBP**** | | | |
| **<120** | ref | ref | ref |
| **120 - 139** | 1.04 (0.84, 1.29) | 0.99 (0.86, 1.15) | 1.03 (0.9, 1.17) |
| **140 - 159** | 1.25 (1.01, 1.55) | 1.08 (0.91, 1.29) | 1.14 (0.99, 1.31) |
| **>=160** | 1.39 (1.08, 1.8) | 1.35 (1.08, 1.69) | 1.36 (1.14, 1.64) |
| **DBP**** | x | | x |
| **<80** | x | ref | x |
| **80 - 89** | x | 0.99 (0.86, 1.14) | x |
| **90 - 99** | x | 1.22 (0.97, 1.52) | x |
| **>=100** | x | 1.11 (0.7, 1.76) | x |
| **No. of concomitant medicines**** | | | |
| **0** | ref | ref | ref |
| **1 – 3** | 0.72 (0.5, 1.03) | 1.04 (0.82, 1.33) | 0.77 (0.66, 0.91) |
| **4 – 6** | 0.8 (0.56, 1.15) | 1.06 (0.83, 1.36) | 0.87 (0.73, 1.03) |

| Predictor | OST<br>OR (95%CI) | IFX<br>OR (95%CI) | OBP<br>OR (95%CI) |
|---|---|---|---|
| **7 – 9** | 1.05 (0.73, 1.52) | 1.22 (0.95, 1.58) | 1.03 (0.86, 1.23) |
| **10 – 12** | 1.1 (0.75, 1.6) | 1.24 (0.95, 1.62) | 1.2 (0.99, 1.45) |
| **>=13** | 1.11 (0.75, 1.64) | 1.3 (0.99, 1.71) | 1.22 (1, 1.48) |
| **Cholesterol measurement** (HDL/LDL)** | | | |
| **<=3.5** | ref | ref | ref |
| **3.6 – 5** | 1.24 (0.95, 1.62) | 1.18 (0.73, 1.91) | 1.16 (1, 1.35) |
| **>5** | 1.53 (1.07, 2.19) | 1.34 (0.45, 3.99) | 1.36 (1.03, 1.81) |
| **No. of previous fractures*** | | x | x |
| **0** | ref | x | x |
| **1** | 1.06 (0.9, 1.25) | x | x |
| **>=2** | 0.81 (0.68, 0.98) | x | x |

**Abbreviations**: OST, patients with incident diagnosis of osteoporosis; IFX, patients with incident fragility fracture; OBP, incident users of oral bisphosphonates; OR, odds ratio; CI, confidence intervals; MACE, composite outcome for the occurrence of either myocardial infarction, stroke or cardiovascular disease death; * ever; ** in the year prior to start; SES, socio-economic status; MI, myocardial infarction; BMI, body mass index; eGFR, estimated Glomerular Filtration Rate; SBP, cholesterol, systolic blood pressure; DBP, diastolic blood pressure.

#### Table S4b. Predictors of 1- and 2-year MI/Stroke models (risk factors selected by lasso regression)

| Predictor | OST OR (95%CI) | | IFX OR (95%CI) | | OBP OR (95%CI) | |
|---|---|---|---|---|---|---|
| | One year | Two years | One year | Two years | One year | Two years |
| Sex = Male (%) | 1.79 (1.43, 2.23) | 1.4 (1.17, 1.68) | x | x | 1.39 (1.22, 1.58) | 1.38 (1.24, 1.53) |
| SES (%) | x | | x | | x | |
| 1 | x | ref | x | ref | x | ref |
| 2 | x | 1.29 (1.06, 1.57) | x | 0.98 (0.84, 1.14) | x | 1.06 (0.94, 1.19) |
| 3 | x | 1.15 (0.94, 1.41) | x | 1 (0.86, 1.17) | x | 1.03 (0.91, 1.16) |
| 4 | x | 1.04 (0.84, 1.3) | x | 1.06 (0.91, 1.24) | x | 1.06 (0.93, 1.2) |
| 5 | x | 1.4 (1.11, 1.76) | x | 1.25 (1.05, 1.47) | x | 1.13 (0.98, 1.31) |
| Smoking** | x | x | | x | x | |
| Ex | x | x | ref | x | x | ref |
| No | x | x | 1.01 (0.84, 1.21) | x | x | 0.91 (0.76, 1.08) |
| Yes | x | x | 1.03 (0.79, 1.34) | x | x | 1.14 (0.92, 1.42) |
| Drinking** | | | | | | |
| Ex | ref | ref | x | ref | ref | ref |
| No | 1.07 (0.67, 1.7) | 1.06 (0.72, 1.55) | 0.97 (0.69, 1.37) | 0.88 (0.68, 1.15) | 1.18 (0.8, 1.74) | 0.95 (0.72, 1.26) |
| Yes | 0.83 (0.52, 1.31) | 0.91 (0.61, 1.37) | 0.96 (0.67, 1.36) | 0.83 (0.63, 1.09) | 0.96 (0.66, 1.39) | 0.86 (0.65, 1.13) |
| Diabetes type I* | x | x | x | x | x | x |
| Diabetes type II* | x | x | x | x | x | x |
| Chronic obstructive pulmonary disease* | x | x | x | x | x | x |
| Chronic kidney disease* | x | x | x | x | x | x |
| Rheumatoid arthritis* | x | x | x | x | x | x |
| Lupus* | x | x | x | x | x | x |
| Systemic heart disease** | x | x | x | x | x | x |
| Anti-osteoporosis use** | x | 1.02 (0.86, 1.21) | x | x | x | x |
| Heparin use** | x | x | x | x | x | x |
| Beta-blocker use** | x | 1.17 (0.99, 1.38) | 1.29 (1.1, 1.51) | 1.3 (1.14, 1.49) | 1.14 (0.99, 1.31) | 1.22 (1.1, 1.36) |
| Hypertension** | x | x | x | x | x | x |
| Deep vein thrombosis or pulmonary embolism** | x | x | x | x | x | x |
| Anticoagulant use** | x | x | x | x | x | x |
| Antidepressants TCA** | x | x | x | x | x | x |
| Antidepressants SSRI** | x | 1.27 (1.03, 1.57) | x | 0.94 (0.79, 1.12) | x | x |
| Hypercholesterolemia** | x | x | x | x | x | x |
| Statin use** | x | 0.97 (0.81, 1.15) | x | x | x | x |
| Osteoporosis history* | x | x | x | x | 0.8 (0.71, 0.91) | 0.81 (0.73, 0.89) |
| Family history of cardiovascular disease | x | x | x | x | x | x |
| Family history of cardiovascular disease before age 60 | x | x | x | x | x | x |
| Heart failure* | x | x | x | x | x | x |
| Migraine* | x | x | x | x | x | x |
| Severe mental illness* | x | x | x | x | x | x |
| Vascular Disease* | x | x | x | x | x | x |
| Atrial fibrillation* | x | 1.32 (1.07, 1.63) | x | x | 1.31 (1.1, 1.56) | x |
| On anti-hypertensive drug | 1.22 (0.94, 1.57) | 1.41 (1.16, 1.73) | 1.25 (1.05, 1.49) | 1.13 (0.96, 1.32) | 1.2 (1.02, 1.42) | 1.15 (1.02, 1.3) |
| Antipsychotic use** | x | x | x | x | x | x |

| Predictor | OST OR (95%CI) | | IFX OR (95%CI) | | OBP OR (95%CI) | |
|---|---|---|---|---|---|---|
| | One year | Two years | One year | Two years | One year | Two years |
| **Steroid use**** | x | | x | x | x | x |
| **Erectile dysfunction**** | x | x | x | x | x | x |
| **Age Group (%)** | | | | | | |
| **50-59** | ref | ref | ref | ref | ref | ref |
| **60-69** | 1.11 (0.65, 1.91) | 1.8 (1.17, 2.76) | 2.57 (1.37, 4.82) | 2.57 (1.59, 4.17) | 1.23 (0.87, 1.74) | 1.75 (1.34, 2.28) |
| **70-79** | 2.07 (1.26, 3.41) | 3.12 (2.06, 4.71) | 4.8 (2.66, 8.65) | 4.9 (3.07, 7.8) | 2.24 (1.62, 3.09) | 3.03 (2.35, 3.92) |
| **80-89** | 3.45 (2.09, 5.71) | 5.13 (3.37, 7.8) | 5.96 (3.31, 10.74) | 6.5 (4.03, 10.46) | 3.39 (2.43, 4.73) | 4.99 (3.83, 6.5) |
| **>89** | 5.27 (3.03, 9.16) | 6.9 (4.34, 10.97) | 6.3 (3.44, 11.54) | 6.45 (3.92, 10.62) | 3.58 (2.47, 5.2) | 5.2 (3.87, 6.99) |
| **Charlson score** | | | x | | | x |
| **0** | ref | ref | ref | ref | ref | x |
| **1** | 1.26 (1, 1.59) | 1.24 (1.04, 1.47) | 1.12 (0.94, 1.34) | 1.04 (0.89, 1.22) | 1.01 (0.87, 1.17) | x |
| **2** | 0.86 (0.64, 1.15) | 0.95 (0.76, 1.18) | 1.07 (0.86, 1.32) | 1.05 (0.88, 1.26) | 0.99 (0.82, 1.19) | x |
| **≥3** | 0.96 (0.71, 1.29) | 1.14 (0.91, 1.44) | 1.03 (0.82, 1.3) | 1.17 (0.96, 1.43) | 1.04 (0.83, 1.31) | x |
| **Cardiovascular disease** | x | x | x | | | |
| **No** | x | x | x | ref | ref | ref |
| **Ever >1 year before index date** | x | x | x | 1.18 (1.01, 1.38) | 1.2 (1.01, 1.42) | 1.22 (1.07, 1.38) |
| **1 year before index** | x | x | x | 1.29 (0.85, 1.96) | 1.29 (0.89, 1.85) | 1.09 (0.8, 1.48) |
| **6 months before index** | x | x | x | 1.38 (0.96, 2) | 2.2 (1.65, 2.93) | 1.51 (1.16, 1.96) |
| **1 month before index** | x | x | x | 1.95 (1.04, 3.65) | 2.62 (1.73, 3.98) | 2.15 (1.5, 3.08) |
| **MI or Stroke** | x | x | | | | |
| **No** | x | x | ref | ref | ref | ref |
| **Ever >1 year before index date** | x | x | 1.22 (0.95, 1.56) | 1.43 (1.15, 1.78) | 1.21 (0.96, 1.53) | 1.3 (1.09, 1.56) |
| **1 year before index** | x | x | 1.48 (1.09, 2.01) | 1.67 (1.27, 2.21) | 1.68 (1.29, 2.2) | 1.79 (1.45, 2.22) |
| **Established CVD *** | 2.19 (1.78, 2.7) | 2.14 (1.81, 2.53) | 1.68 (1.36, 2.06) | 1.18 (0.98, 1.43) | 1.36 (1.12, 1.66) | 1.37 (1.18, 1.59) |
| **Any fracture history** | x | x | x | x | x | x |
| **No** | x | x | x | x | x | x |
| **Ever >1 year before index date** | x | x | x | x | x | x |
| **1 year before index** | x | x | x | x | x | x |
| **Hip fracture history** | x | x | x | x | x | x |
| **No** | x | x | x | x | x | x |
| **Ever >1 year before index date** | x | x | x | x | x | x |
| **1 year before index** | x | x | x | x | x | x |
| **Shoulder fracture history** | x | x | x | x | x | x |
| **No** | x | x | x | x | x | x |
| **Ever >1 year before index date** | x | x | x | x | x | x |
| **1 year before index** | x | x | x | x | x | x |
| **Spine fracture history** | x | x | x | x | x | x |
| **No** | x | x | x | x | x | x |
| **Ever >1 year before index date** | x | x | x | x | x | x |
| **1 year before index** | x | x | x | x | x | x |
| **Wrist fracture history** | x | x | x | x | x | x |
| **No** | x | x | x | x | x | x |

| Predictor | OST OR (95%CI) | | IFX OR (95%CI) | | OBP OR (95%CI) | |
|---|---|---|---|---|---|---|
| | One year | Two years | One year | Two years | One year | Two years |
| Ever >1 year before index date | x | x | x | x | x | x |
| 1 year before index | x | x | x | x | x | x |
| **BMI**** | | | x | | | x |
| <18.5 | ref | ref | x | ref | ref | x |
| 18.6 - 24.9 | 0.86 (0.55, 1.34) | 0.79 (0.57, 1.11) | x | 1 (0.77, 1.31) | 0.82 (0.62, 1.09) | x |
| 25 - 29.9 | 0.77 (0.44, 1.35) | 0.68 (0.46, 1) | x | 1 (0.68, 1.47) | 0.69 (0.49, 0.96) | x |
| 30 - 39.9 | 0.64 (0.33, 1.22) | 0.63 (0.37, 1.05) | x | 1.1 (0.65, 1.87) | 0.7 (0.47, 1.04) | x |
| >=40 | 0.77 (0.22, 2.66) | 0.8 (0.31, 2.11) | x | 0.98 (0.36, 2.7) | 0.65 (0.3, 1.41) | x |
| **No. of GP visits**** | | | | | | |
| 0 | ref | ref | ref | ref | ref | ref |
| 1-5 | 0.99 (0.55, 1.78) | 0.7 (0.48, 1.02) | 1 (0.8, 1.25) | 1 (0.76, 1.31) | 0.87 (0.68, 1.1) | 0.94 (0.78, 1.13) |
| 6-10 | 1.03 (0.56, 1.91) | 0.68 (0.46, 1.02) | 0.89 (0.7, 1.12) | 0.92 (0.68, 1.24) | 0.85 (0.64, 1.13) | 0.94 (0.75, 1.16) |
| 11-15 | 1.05 (0.56, 1.96) | 0.67 (0.45, 1.02) | 0.83 (0.64, 1.08) | 0.99 (0.73, 1.35) | 0.76 (0.56, 1.02) | 0.93 (0.75, 1.17) |
| >=16 | 1.28 (0.69, 2.38) | 0.82 (0.55, 1.23) | 1.04 (0.82, 1.32) | 0.97 (0.71, 1.31) | 0.82 (0.61, 1.1) | 1.05 (0.84, 1.31) |
| **No. of GP emergency visits**** | x | x | x | x | | |
| 0 | x | x | x | x | ref | ref |
| 1 | x | x | x | x | 1.17 (0.99, 1.38) | 1.11 (0.97, 1.26) |
| 2 | x | x | x | x | 1.67 (1.35, 2.07) | 1.35 (1.13, 1.61) |
| 3-5 | x | x | x | x | 1.14 (0.89, 1.46) | 1.25 (1.04, 1.5) |
| >=6 | x | x | x | x | 2.21 (1.65, 2.98) | 1.61 (1.26, 2.06) |
| **eGFR**** | | | | x | | |
| <=29 | ref | ref | ref | ref | ref | ref |
| 30 – 44 | 0.79 (0.42, 1.49) | 0.96 (0.53, 1.74) | 0.95 (0.62, 1.45) | 1.04 (0.72, 1.52) | 1.04 (0.62, 1.76) | 0.96 (0.64, 1.44) |
| 45 – 59 | 0.64 (0.32, 1.27) | 0.74 (0.41, 1.35) | 0.77 (0.47, 1.25) | 0.89 (0.55, 1.45) | 0.91 (0.48, 1.7) | 0.81 (0.51, 1.3) |
| 60 – 89 | 0.44 (0.23, 0.86) | 0.57 (0.32, 1.02) | 0.68 (0.35, 1.3) | 0.91 (0.54, 1.54) | 0.65 (0.29, 1.5) | 0.65 (0.35, 1.21) |
| >=90 | 0.53 (0.2, 1.38) | 0.64 (0.28, 1.49) | 0.72 (0.32, 1.63) | 0.98 (0.53, 1.81) | 0.58 (0.22, 1.56) | 0.63 (0.31, 1.3) |
| **SBP**** | | | | | | |
| <120 | ref | ref | ref | ref | ref | ref |
| 120 - 139 | 0.94 (0.69, 1.27) | 0.87 (0.7, 1.1) | 1.06 (0.84, 1.34) | 1.14 (0.95, 1.37) | 1.01 (0.82, 1.25) | 1.08 (0.93, 1.25) |
| 140 - 159 | 1.16 (0.86, 1.58) | 0.96 (0.76, 1.22) | 1.27 (1, 1.62) | 1.29 (1.07, 1.57) | 1.18 (0.94, 1.48) | 1.19 (1.02, 1.39) |
| >=160 | 1.39 (0.97, 2) | 1.21 (0.91, 1.62) | 1.46 (1.1, 1.93) | 1.74 (1.38, 2.2) | 1.51 (1.14, 2.01) | 1.39 (1.15, 1.68) |
| **DBP**** | x | | x | x | | x |
| <80 | x | ref | x | x | ref | x |
| 80 - 89 | x | 1.07 (0.9, 1.27) | x | x | 1.02 (0.88, 1.18) | x |
| 90 - 99 | x | 1.07 (0.81, 1.42) | x | x | 1.03 (0.82, 1.3) | x |
| >=100 | x | 1.11 (0.66, 1.88) | x | x | 1.19 (0.76, 1.85) | x |
| **No. of concomitant medicines**** | | | x | | | |
| 0 | ref | ref | x | ref | ref | ref |
| 1 – 3 | 0.96 (0.55, 1.69) | 0.93 (0.63, 1.37) | x | 0.95 (0.71, 1.27) | 0.82 (0.64, 1.07) | 0.79 (0.66, 0.96) |
| 4 – 6 | 0.76 (0.42, 1.35) | 0.8 (0.54, 1.19) | x | 1 (0.74, 1.35) | 0.8 (0.6, 1.05) | 0.79 (0.64, 0.97) |
| 7 – 9 | 1.04 (0.58, 1.86) | 0.86 (0.57, 1.3) | x | 0.99 (0.72, 1.36) | 1.02 (0.76, 1.36) | 0.86 (0.69, 1.07) |
| 10 – 12 | 0.96 (0.52, 1.75) | 0.83 (0.54, 1.27) | x | 1.11 (0.79, 1.55) | 1.08 (0.79, 1.47) | 0.96 (0.76, 1.21) |
| >=13 | 1.35 (0.74, 2.47) | 0.88 (0.56, 1.36) | x | 1.06 (0.74, 1.52) | 1.09 (0.79, 1.49) | 0.9 (0.71, 1.15) |
| **Cholesterol measurement** (HDL/LDL)** | x | | x | | x | |
| <=3.5 | x | ref | x | ref | x | ref |
| 3.6 – 5 | x | 1.28 (0.95, 1.73) | x | 1.19 (0.48, 2.94) | x | 1.13 (0.96, 1.32) |
| >5 | x | 1.49 (1.04, 2.14) | x | 1.52 (0.27, 8.44) | x | 1.36 (1.01, 1.82) |
| **No. of previous fractures*** | x | | x | x | x | x |

| Predictor | OST OR (95%CI) | | IFX OR (95%CI) | | OBP OR (95%CI) | |
|---|---|---|---|---|---|---|
| | One year | Two years | One year | Two years | One year | Two years |
| **0** | x | ref | x | x | x | x |
| **1** | x | 1.15 (0.96, 1.36) | x | x | x | x |
| **>=2** | x | 1.05 (0.87, 1.26) | x | x | x | x |

**Abbreviations:** OST, patients with incident diagnosis of osteoporosis; IFX, patients with incident fragility fracture; OBP, incident users of oral bisphosphonates; OR, odds ratio; CI, confidence intervals; MI, myocardial infarction; * ever; ** in the year prior to start; SES, socio-economic status; BMI, body mass index; eGFR, estimated Glomerular Filtration Rate; SBP, cholesterol, systolic blood pressure; DBP, diastolic blood pressure.

#### Table S5a. Model equations for 2-year MACE models (risk factors selected by lasso regression)

| Predictor | OST Beta coefficients | IFX Beta coefficients | OBP Beta coefficients |
|---|---|---|---|
| **Intercept** | -3.711 | -4.652 | -3.863 |
| **Sex = Male (%)** | 0.425 | 0.307 | 0.342 |
| **SES (%)** | | x | x |
| **1** | ref | x | x |
| **2** | -0.043 | x | x |
| **3** | -0.010 | x | x |
| **4** | -0.119 | x | x |
| **5** | -0.030 | x | x |
| **Smoking**** | | x | |
| **Ex** | ref | x | ref |
| **No** | -0.001 | x | -0.058 |
| **Yes** | 0.223 | x | 0.139 |
| **Drinking**** | | | |
| **Ex** | ref | ref | ref |
| **No** | 0.031 | -0.117 | -0.131 |
| **Yes** | -0.119 | -0.285 | -0.280 |
| **Diabetes type I*** | x | x | x |
| **Diabetes type II*** | | x | 0.274 |
| **Chronic obstructive pulmonary disease*** | x | x | x |
| **Chronic kidney disease*** | x | x | x |
| **Rheumatoid arthritis*** | x | x | -0.152 |
| **Lupus*** | x | x | x |
| **Systemic heart disease**** | x | x | x |
| **Anti-osteoporosis use**** | x | x | x |
| **Heparin use**** | x | x | x |
| **Beta-blocker use**** | 0.039 | 0.160 | 0.137 |
| **Hypertension**** | x | x | x |
| **Deep vein thrombosis or pulmonary embolism**** | x | x | x |
| **Anticoagulant use**** | x | x | x |

| Predictor | OST<br>Beta coefficients | IFX<br>Beta coefficients | OBP<br>Beta coefficients |
|---|---|---|---|
| **Antidepressants TCA**** | x | x | x |
| **Antidepressants SSRI**** | 0.323 | x | x |
| **Hypercholesterolemia**** | x | x | x |
| **Statin use**** | x | -0.093 | x |
| **Osteoporosis history*** | x | x | -0.214 |
| **Family history of cardiovascular disease (%)** | x | x | x |
| **Family history of cardiovascular disease before age 60** | x | x | x |
| **Heart failure*** | x | x | x |
| **Migraine*** | x | x | x |
| **Severe mental illness*** | x | x | x |
| **Vascular Disease*** | x | x | x |
| **Atrial fibrillation*** | 0.141 | 0.066 | 0.299 |
| **On anti-hypertensive drug** | 0.259 | 0.200 | x |
| **Antipsychotic use**** | x | x | x |
| **Steroid use**** | x | | x |
| **Erectile dysfunction**** | x | x | x |
| **Age Group (%)** | | | |
| **50-59** | ref | ref | ref |
| **60-69** | 0.309 | 1.000 | 0.630 |
| **70-79** | 0.968 | 1.495 | 1.254 |
| **80-89** | 1.492 | 1.998 | 1.759 |
| **>89** | 1.878 | 2.044 | 2.076 |
| **Charlson score** | | x | |
| **0** | ref | x | ref |
| **1** | 0.190 | x | -0.016 |
| **2** | -0.092 | x | -0.134 |
| **≥3** | 0.064 | x | -0.115 |
| **Cardiovascular disease** | x | x | |
| **No** | x | x | ref |
| **Ever >1 year before index date** | x | x | 0.053 |
| **1 year before index** | x | x | 0.101 |
| **6 months before index** | x | x | 0.267 |
| **1 month before index** | x | x | 0.618 |
| **MI or Stroke** | | | |
| **No** | ref | ref | ref |
| **Ever >1 year before index date** | 0.148 | 0.230 | 0.263 |
| **1 year before index** | 0.711 | 0.506 | 0.628 |
| **Established CVD *** | 0.504 | 0.424 | 0.517 |
| **Any fracture history** | x | x | x |
| **No** | x | x | x |
| **Ever >1 year before index date** | x | x | x |
| **1 year before index** | x | x | x |
| **Hip fracture history** | x | x | x |
| **No** | x | x | x |

| Predictor | OST<br>Beta coefficients | IFX<br>Beta coefficients | OBP<br>Beta coefficients |
|---|---|---|---|
| Ever >1 year before index date | x | x | x |
| 1 year before index | x | x | x |
| Shoulder fracture history | x | x | x |
| No | x | x | x |
| Ever >1 year before index date | x | x | x |
| 1 year before index | x | x | x |
| Spine fracture history | x | x | x |
| No | x | x | x |
| Ever >1 year before index date | x | x | x |
| 1 year before index | x | x | x |
| Wrist fracture history | x | x | x |
| No | x | x | x |
| Ever >1 year before index date | x | x | x |
| 1 year before index | x | x | x |
| BMI** | | x | |
| <18.5 | ref | x | ref |
| 18.6 - 24.9 | -0.276 | x | -0.233 |
| 25 - 29.9 | -0.438 | x | -0.420 |
| 30 - 39.9 | -0.531 | x | -0.501 |
| >=40 | -0.273 | x | -0.472 |
| No. of GP visits** | | | |
| 0 | ref | ref | ref |
| 1-5 | -0.709 | -0.082 | -0.000 |
| 6-10 | -0.636 | -0.239 | -0.112 |
| 11-15 | -0.721 | -0.195 | -0.153 |
| >=16 | -0.606 | -0.214 | -0.072 |
| No. of GP emergency visits** | | | x |
| 0 | ref | ref | x |
| 1 | 0.173 | 0.147 | x |
| 2 | 0.278 | 0.334 | x |
| 3-5 | 0.158 | 0.182 | x |
| >=6 | 0.338 | 0.302 | x |
| eGFR** | | | |
| <=29 | ref | ref | ref |
| 30 – 44 | 0.020 | -0.003 | -0.082 |
| 45 – 59 | -0.186 | -0.123 | -0.249 |
| 60 – 89 | -0.427 | -0.147 | -0.459 |
| >=90 | -0.403 | -0.189 | -0.494 |
| SBP** | | | |
| <120 | ref | ref | ref |
| 120 - 139 | 0.044 | -0.008 | 0.025 |
| 140 - 159 | 0.221 | 0.079 | 0.127 |
| >=160 | 0.330 | 0.301 | 0.311 |
| DBP** | x | | x |

| Predictor | OST<br>Beta coefficients | IFX<br>Beta coefficients | OBP<br>Beta coefficients |
|---|---|---|---|
| **<80** | x | ref | x |
| **80 - 89** | x | -0.007 | x |
| **90 - 99** | x | 0.195 | x |
| **>=100** | x | 0.104 | x |
| **No. of concomitant medicines**** | | | |
| **0** | ref | ref | ref |
| **1 – 3** | -0.334 | 0.043 | -0.255 |
| **4 – 6** | -0.219 | 0.057 | -0.143 |
| **7 – 9** | 0.050 | 0.200 | 0.031 |
| **10 – 12** | 0.091 | 0.215 | 0.180 |
| **>=13** | 0.102 | 0.266 | 0.195 |
| **Cholesterol measurement** (HDL/LDL)** | | | |
| **<=3.5** | ref | ref | ref |
| **3.6 – 5** | 0.215 | 0.167 | 0.148 |
| **>5** | 0.423 | 0.294 | 0.310 |
| **No. of previous fractures*** | | x | x |
| **0** | ref | x | x |
| **1** | 0.060 | x | x |
| **>=2** | -0.205 | x | x |

**Abbreviations**: OST, patients with incident diagnosis of osteoporosis; IFX, patients with incident fragility fracture; OBP, incident users of oral bisphosphonates; OR, odds ratio; CI, confidence intervals; MACE, composite outcome for the occurrence of either myocardial infarction, stroke or cardiovascular disease death; * ever; ** in the year prior to start; SES, socio-economic status; MI, myocardial infarction; BMI, body mass index; eGFR, estimated Glomerular Filtration Rate; SBP, cholesterol, systolic blood pressure; DBP, diastolic blood pressure.

#### Table S5b. Model equations for 1- and 2-year MI/Stroke models (risk factors selected by lasso regression)

| Predictor | OST<br>Beta coefficients | | IFX<br>Beta coefficients | | OBP<br>Beta coefficients | |
|---|---|---|---|---|---|---|
| | **One year** | **Two years** | **One year** | **Two years** | **One year** | **Two years** |
| **Intercept** | -4.635 | -4.341 | -5.415 | -5.235 | -4.580 | -4.456 |
| **Sex = Male (%)** | 0.582 | 0.337 | x | x | 0.327 | 0.321 |
| **SES (%)** | x | | x | | x | |
| **1** | x | ref | x | ref | x | ref |
| **2** | x | 0.255 | x | -0.020 | x | 0.056 |
| **3** | x | 0.140 | x | 0.004 | x | 0.026 |
| **4** | x | 0.042 | x | 0.062 | x | 0.057 |
| **5** | x | 0.337 | x | 0.221 | x | 0.126 |
| **Smoking**** | x | x | x | x | x | |
| **Ex** | x | x | x | x | x | ref |
| **No** | x | x | 0.008 | x | x | -0.098 |
| **Yes** | x | x | 0.030 | x | x | 0.135 |

| Predictor | OST<br>Beta coefficients | | IFX<br>Beta coefficients | | OBP<br>Beta coefficients | |
|---|---|---|---|---|---|---|
| | One year | Two years | One year | Two years | One year | Two years |
| **Drinking**** | | | x | | | |
| **Ex** | ref | ref | x | ref | ref | ref |
| **No** | 0.065 | 0.055 | -0.031 | -0.123 | 0.168 | -0.049 |
| **Yes** | -0.192 | -0.091 | -0.043 | -0.184 | -0.045 | -0.151 |
| **Diabetes type I*** | x | x | x | x | x | x |
| **Diabetes type II*** | x | x | x | x | x | x |
| **Chronic obstructive pulmonary disease*** | x | x | x | x | x | x |
| **Chronic kidney disease*** | x | x | x | x | x | x |
| **Rheumatoid arthritis*** | x | x | x | x | x | x |
| **Lupus*** | x | x | x | x | x | x |
| **Systemic heart disease**** | x | x | x | x | x | x |
| **Anti-osteoporosis use**** | x | 0.019 | x | x | x | x |
| **Heparin use**** | x | x | x | x | x | x |
| **Beta-blocker use**** | x | 0.155 | 0.251 | 0.264 | 0.127 | 0.203 |
| **Hypertension**** | x | x | x | x | x | x |
| **Deep vein thrombosis or pulmonary embolism**** | x | x | x | x | x | x |
| **Anticoagulant use**** | x | x | x | x | x | x |
| **Antidepressants TCA**** | x | x | x | x | x | x |
| **Antidepressants SSRI**** | x | 0.240 | x | -0.066 | x | x |
| **Hypercholesterolemia**** | x | x | x | x | x | x |
| **Statin use**** | x | -0.035 | x | x | x | x |
| **Osteoporosis history*** | x | x | x | x | -0.221 | -0.212 |
| **Family history of cardiovascular disease (%)** | x | x | x | x | x | x |
| **Family history of cardiovascular disease before age 60** | x | x | x | x | x | x |
| **Heart failure*** | x | x | x | x | x | x |
| **Migraine*** | x | x | x | x | x | x |
| **Severe mental illness*** | x | x | x | x | x | x |
| **Vascular Disease*** | x | x | x | x | x | x |
| **Atrial fibrillation*** | x | 0.279 | x | x | 0.270 | x |
| **On anti-hypertensive drug** | 0.196 | 0.347 | 0.221 | 0.120 | 0.185 | 0.140 |
| **Antipsychotic use**** | x | x | x | x | x | x |
| **Steroid use**** | x | | x | x | x | x |
| **Erectile dysfunction**** | x | x | x | x | x | x |
| **Age Group (%)** | | | | | | |
| **50-59** | ref | ref | ref | ref | ref | ref |
| **60-69** | 0.108 | 0.586 | 0.945 | 0.945 | 0.208 | 0.560 |
| **70-79** | 0.730 | 1.136 | 1.568 | 1.588 | 0.805 | 1.109 |
| **80-89** | 1.239 | 1.634 | 1.786 | 1.871 | 1.222 | 1.608 |
| **>89** | 1.662 | 1.932 | 1.841 | 1.864 | 1.277 | 1.648 |

| Predictor | OST<br>Beta coefficients | | IFX<br>Beta coefficients | | OBP<br>Beta coefficients | |
|---|---|---|---|---|---|---|
| | One year | Two years | One year | Two years | One year | Two years |
| **Charlson score** | | | | | | x |
| **0** | ref | ref | ref | ref | ref | x |
| **1** | 0.233 | 0.212 | 0.115 | 0.044 | 0.009 | x |
| **2** | -0.150 | -0.056 | 0.065 | 0.053 | -0.010 | x |
| **≥3** | -0.042 | 0.134 | 0.030 | 0.159 | 0.040 | x |
| **Cardiovascular disease** | x | x | x | | | |
| **No** | x | x | x | ref | ref | ref |
| **Ever >1 year before index date** | x | x | x | 0.164 | 0.181 | 0.197 |
| **1 year before index** | x | x | x | 0.256 | 0.252 | 0.083 |
| **6 months before index** | x | x | x | 0.324 | 0.789 | 0.414 |
| **1 month before index** | x | x | x | 0.669 | 0.964 | 0.767 |
| **MI or Stroke** | x | x | | | | |
| **No** | x | x | ref | ref | ref | ref |
| **Ever >1 year before index date** | x | x | 0.199 | 0.357 | 0.190 | 0.264 |
| **1 year before index** | x | x | 0.390 | 0.515 | 0.519 | 0.583 |
| **Established CVD *** | 0.785 | 0.760 | 0.516 | 0.168 | 0.310 | 0.312 |
| **Any fracture history** | x | x | x | x | x | x |
| **No** | x | x | x | x | x | x |
| **Ever >1 year before index date** | x | x | x | x | x | x |
| **1 year before index** | x | x | x | x | x | x |
| **Hip fracture history** | x | x | x | x | x | x |
| **No** | x | x | x | x | x | x |
| **Ever >1 year before index date** | x | x | x | x | x | x |
| **1 year before index** | x | x | x | x | x | x |
| **Shoulder fracture history** | x | x | x | x | x | x |
| **No** | x | x | x | x | x | x |
| **Ever >1 year before index date** | x | x | x | x | x | x |
| **1 year before index** | x | x | x | x | x | x |
| **Spine fracture history** | x | x | x | x | x | x |
| **No** | x | x | x | x | x | x |
| **Ever >1 year before index date** | x | x | x | x | x | x |
| **1 year before index** | x | x | x | x | x | x |
| **Wrist fracture history** | x | x | x | x | x | x |
| **No** | x | x | x | x | x | x |
| **Ever >1 year before index date** | x | x | x | x | x | x |
| **1 year before index** | x | x | x | x | x | x |

| Predictor | OST Beta coefficients | | IFX Beta coefficients | | OBP Beta coefficients | |
|---|---|---|---|---|---|---|
| | One year | Two years | One year | Two years | One year | Two years |
| **BMI**** | | | x | x | | x |
| **<18.5** | ref | ref | x | x | ref | x |
| **18.6 - 24.9** | -0.149 | -0.232 | x | 0.004 | -0.201 | x |
| **25 - 29.9** | -0.261 | -0.391 | x | -0.002 | -0.371 | x |
| **30 - 39.9** | -0.453 | -0.469 | x | 0.097 | -0.361 | x |
| **>=40** | -0.265 | -0.218 | x | -0.019 | -0.434 | x |
| **No. of GP visits**** | | | | x | | |
| **0** | ref | ref | ref | ref | ref | ref |
| **1-5** | -0.009 | -0.356 | 0.002 | -0.004 | -0.142 | -0.061 |
| **6-10** | 0.032 | -0.380 | -0.120 | -0.084 | -0.164 | -0.065 |
| **11-15** | 0.045 | -0.394 | -0.182 | -0.009 | -0.279 | -0.068 |
| **>=16** | 0.246 | -0.200 | 0.039 | -0.035 | -0.197 | 0.051 |
| **No. of GP emergency visits**** | x | x | x | x | | |
| **0** | x | x | x | x | ref | ref |
| **1** | x | x | x | x | 0.156 | 0.100 |
| **2** | x | x | x | x | 0.513 | 0.299 |
| **3-5** | x | x | x | x | 0.129 | 0.222 |
| **>=6** | x | x | x | x | 0.795 | 0.476 |
| **eGFR**** | | | | | | |
| **<=29** | ref | ref | ref | ref | ref | ref |
| **30 – 44** | -0.237 | -0.039 | -0.050 | 0.043 | 0.040 | -0.037 |
| **45 – 59** | -0.448 | -0.301 | -0.266 | -0.112 | -0.098 | -0.208 |
| **60 – 89** | -0.812 | -0.565 | -0.392 | -0.096 | -0.424 | -0.426 |
| **>=90** | -0.642 | -0.442 | -0.326 | -0.020 | -0.540 | -0.458 |
| **SBP**** | | | | | | |
| **<120** | ref | ref | ref | ref | ref | ref |
| **120 - 139** | -0.062 | -0.136 | 0.058 | 0.134 | 0.014 | 0.077 |
| **140 - 159** | 0.151 | -0.039 | 0.239 | 0.257 | 0.164 | 0.175 |
| **>=160** | 0.331 | 0.193 | 0.375 | 0.555 | 0.413 | 0.329 |
| **DBP**** | x | | x | x | | x |
| **<80** | x | ref | x | x | ref | x |
| **80 - 89** | x | 0.063 | x | x | 0.018 | x |
| **90 - 99** | x | 0.071 | x | x | 0.034 | x |
| **>=100** | x | 0.108 | x | x | 0.171 | x |
| **No. of concomitant medicines**** | | | x | | | |
| **0** | ref | ref | x | ref | ref | ref |
| **1 – 3** | -0.040 | -0.076 | x | -0.055 | -0.193 | -0.231 |
| **4 – 6** | -0.279 | -0.223 | x | 0.002 | -0.229 | -0.237 |
| **7 – 9** | 0.038 | -0.153 | x | -0.008 | 0.015 | -0.151 |
| **10 – 12** | -0.045 | -0.191 | x | 0.100 | 0.077 | -0.044 |
| **>=13** | 0.298 | -0.133 | x | 0.059 | 0.085 | -0.104 |
| **Cholesterol measurement** (HDL/LDL)** | x | | x | | x | |

| Predictor | OST<br>Beta coefficients | | IFX<br>Beta coefficients | | OBP<br>Beta coefficients | |
|---|---|---|---|---|---|---|
| | One year | Two years | One year | Two years | One year | Two years |
| **<=3.5** | x | ref | x | ref | x | ref |
| **3.6 – 5** | x | 0.249 | x | 0.175 | x | 0.120 |
| **>5** | x | 0.399 | x | 0.417 | x | 0.306 |
| **No. of previous fractures*** | x | | x | x | | x |
| **0** | x | ref | x | x | ref | x |
| **1** | x | 0.136 | x | x | | x |
| **>=2** | x | 0.047 | x | x | | x |

**Abbreviations:** OST, patients with incident diagnosis of osteoporosis; IFX, patients with incident fragility fracture; OBP, incident users of oral bisphosphonates; MI, myocardial infarction; * ever; ** in the year prior to start; SES, socio-economic status; BMI, body mass index; eGFR, estimated Glomerular Filtration Rate; SBP, cholesterol, systolic blood pressure; DBP, diastolic blood pressure.

#### Table S6a. Risk factors selected by lasso for one- and two-year models in gender based models (OST cohort)

| OST cohort Predictor | Women OR (95%CI) | | | | Men OR (95%CI) | | | |
|---|---|---|---|---|---|---|---|---|
| | One year | | Two year | | One year | | Two year | |
| | MACE | Stroke/MI | MACE | Stroke/MI | MACE | Stroke/MI | MACE | Stroke/MI |
| **SES** | | x | | x | x | x | x | x |
| **1** | ref | x | ref | x | x | x | x | x |
| **2** | 0.92 (0.7, 1.21) | x | 1.01 (0.83, 1.24) | x | x | x | x | x |
| **3** | 1 (0.76, 1.32) | x | 1.15 (0.94, 1.42) | x | x | x | x | x |
| **4** | 1.03 (0.78, 1.37) | x | 0.99 (0.8, 1.24) | x | x | x | x | x |
| **5** | 0.96 (0.69, 1.32) | x | 1.35 (1.07, 1.69) | x | x | x | x | x |
| **Smoking**** | | x | | x | x | x | x | x |
| **Ex** | ref | x | ref | x | x | x | x | x |
| **No** | 1.01 (0.77, 1.32) | x | 1.03 (0.83, 1.29) | x | x | x | x | x |
| **Yes** | 1.51 (1.06, 2.16) | x | 1.32 (0.99, 1.76) | x | x | x | x | x |
| **Drinking**** | | | | | x | x | x | x |
| **Ex** | ref | ref | ref | ref | x | x | x | x |
| **No** | 1.03 (0.61, 1.75) | 1.02 (0.65, 1.61) | 0.97 (0.64, 1.45) | 0.99 (0.66, 1.46) | x | x | x | x |
| **Yes** | 0.8 (0.47, 1.35) | 0.75 (0.45, 1.27) | 0.83 (0.54, 1.26) | 0.85 (0.55, 1.3) | x | x | x | x |
| **Diabetes type I*** | x | x | x | x | x | x | x | x |
| **Diabetes type II*** | x | x | x | | x | x | x | x |
| **Chronic obstructive pulmonary disease*** | x | x | x | x | x | x | x | x |
| **Chronic kidney disease*** | x | x | 0.88 (0.66, 1.18) | x | x | x | x | x |
| **Rheumatoid arthritis*** | x | x | x | x | x | x | x | x |
| **Lupus*** | x | x | x | x | x | x | x | x |
| **Systemic heart disease**** | x | x | x | x | x | x | x | x |
| **Anti-osteoporosis use**** | | x | 1.12 (0.94, 1.32) | x | x | x | x | x |
| **Heparin use**** | x | x | x | x | x | x | x | x |
| **Beta-blocker use**** | x | 1.48 (1.17, 1.87) | x | 1.23 (1.03, 1.47) | x | x | x | x |
| **Hypertension**** | x | x | x | x | x | x | x | x |
| **Deep vein thrombosis or pulmonary embolism**** | x | x | x | x | x | x | x | x |
| **Anticoagulant use**** | x | x | x | x | x | x | x | x |
| **Antidepressants TCA**** | x | x | x | x | x | x | x | x |
| **Antidepressants SSRI**** | 1.42 (1.09, 1.86) | x | 1.27 (1.03, 1.57) | x | x | x | x | x |
| **Hypercholesterolemia**** | x | x | x | x | x | x | x | x |

| OST cohort<br>Predictor | Women<br>OR (95%CI) | | | | Men<br>OR (95%CI) | | | |
|---|---|---|---|---|---|---|---|---|
| | One year | | Two year | | One year | | Two year | |
| | MACE | Stroke/MI | MACE | Stroke/MI | MACE | Stroke/MI | MACE | Stroke/MI |
| **Statin use**** | x | x | x | x | x | x | x | x |
| **Osteoporosis history*** | x | x | x | x | x | x | x | x |
| **Family history of cardiovascular disease (%)** | x | x | x | x | x | x | x | x |
| **Family history of cardiovascular disease before age 60** | x | x | x | x | x | x | x | x |
| **Heart failure*** | x | x | x | x | x | x | x | x |
| **Migraine*** | x | x | x | | x | x | x | x |
| **Severe mental illness*** | x | x | x | x | x | x | x | x |
| **Vascular Disease*** | x | x | x | x | x | x | x | x |
| **Atrial fibrillation*** | 1.37 (1.03, 1.81) | x | 1.35 (1.08, 1.69) | x | x | x | x | x |
| **On anti-hypertensive drug** | 1.27 (0.97, 1.66) | 1.21 (0.89, 1.62) | 1.17 (0.97, 1.42) | 1.51 (1.21, 1.89) | x | x | | x |
| **Antipsychotic use**** | x | x | x | x | x | x | x | x |
| **Steroid use**** | 1.05 (0.81, 1.38) | x | 1.14 (0.94, 1.39) | | x | x | x | x |
| **Erectile dysfunction**** | x | x | x | x | x | x | x | x |
| **Age Group** | | | | | | x | | x |
| **50-59** | ref | ref | ref | ref | ref | x | ref | x |
| **60-69** | 1.03 (0.56, 1.89) | 1.04 (0.52, 2.06) | 2.23 (1.35, 3.68) | 1.63 (0.98, 2.72) | 1.73 (0.81, 3.71) | x | 2.98 (1.39, 6.41) | x |
| **70-79** | 2.57 (1.49, 4.42) | 2.87 (1.57, 5.27) | 4.57 (2.82, 7.4) | 3.5 (2.17, 5.64) | 1.52 (0.73, 3.2) | x | 3.46 (1.64, 7.3) | x |
| **80-89** | 4.66 (2.67, 8.15) | 5.21 (2.84, 9.56) | 7.87 (4.81, 12.86) | 5.25 (3.24, 8.52) | 2.2 (1.06, 4.56) | x | 4.66 (2.2, 9.88) | x |
| **>89** | 5.83 (3.13, 10.83) | 6.73 (3.48, 13.01) | 11.44 (6.74, 19.42) | 7.45 (4.42, 12.56) | 2.11 (0.8, 5.6) | x | 5.12 (2.1, 12.48) | x |
| **Charlson score** | | x | | | x | x | x | x |
| **0** | ref | x | ref | ref | x | x | x | x |
| **1** | 1.28 (1, 1.62) | x | 1.25 (1.05, 1.49) | 1.26 (1.05, 1.53) | x | x | x | x |
| **2** | 0.93 (0.69, 1.26) | x | 1.03 (0.81, 1.3) | 1 (0.79, 1.27) | x | x | x | x |
| **≥3** | 1.3 (0.96, 1.76) | x | 1.22 (0.92, 1.61) | 1.21 (0.95, 1.54) | x | x | x | x |
| **Cardiovascular disease** | x | x | x | x | x | x | x | x |
| **No** | x | x | x | x | x | x | x | x |
| **Ever >1 year before index date** | x | x | x | x | x | x | x | x |
| **1 year before index** | x | x | x | x | x | x | x | x |
| **6 months before index** | x | x | x | x | x | x | x | x |
| **1 month before index** | x | x | x | x | x | x | x | x |

| OST cohort<br>Predictor | Women<br>OR (95%CI) | | | | Men<br>OR (95%CI) | | | |
|---|---|---|---|---|---|---|---|---|
| | One year | | Two year | | One year | | Two year | |
| | MACE | Stroke/MI | MACE | Stroke/MI | MACE | Stroke/MI | MACE | Stroke/MI |
| **MI or Stroke** | | x | | x | x | x | x | x |
| **No** | ref | x | ref | x | x | x | x | x |
| **Ever >1 year before index date** | 1.33 (0.89, 1.99) | x | 1.3 (0.96, 1.77) | x | x | x | x | x |
| **1 year before index** | 2.45 (1.55, 3.86) | x | 1.68 (1.15, 2.47) | x | x | x | x | x |
| **Established CVD *** | 1.53 (1.11, 2.11) | 2.21 (1.75, 2.78) | 1.6 (1.26, 2.04) | 2.01 (1.68, 2.4) | 2.31 (1.58, 3.39) | 2.19 (1.42, 3.36) | 2.24 (1.67, 3.01) | 2.29 (1.64, 3.19) |
| **Any fracture history** | | x | x | x | x | x | x | x |
| **No** | ref | x | x | x | x | x | x | x |
| **Ever >1 year before index date** | 1 (0.64, 1.55) | x | x | x | x | x | x | x |
| **1 year before index** | 1.16 (0.78, 1.73) | x | x | x | x | x | x | x |
| **Hip fracture history** | x | x | x | x | x | x | x | x |
| **No** | x | x | x | x | x | x | x | x |
| **Ever >1 year before index date** | x | x | x | x | x | x | x | x |
| **1 year before index** | x | x | x | x | x | x | x | x |
| **Shoulder fracture history** | x | x | x | x | x | x | x | x |
| **No** | x | x | x | x | x | x | x | x |
| **Ever >1 year before index date** | x | x | x | x | x | x | x | x |
| **1 year before index** | x | x | x | x | x | x | x | x |
| **Spine fracture history** | x | x | x | x | x | x | x | x |
| **No** | x | x | x | x | x | x | x | x |
| **Ever >1 year before index date** | x | x | x | x | x | x | x | x |
| **1 year before index** | x | x | x | x | x | x | x | x |
| **Wrist fracture history** | x | x | x | x | x | x | x | x |
| **No** | x | x | x | x | x | x | x | x |
| **Ever >1 year before index date** | x | x | x | x | x | x | x | x |
| **1 year before index** | x | x | x | x | x | x | x | x |
| **BMI**** | | x | | | x | x | x | x |
| **<18.5** | ref | x | ref | ref | x | x | x | x |
| **18.6 - 24.9** | 0.65 (0.46, 0.91) | x | 0.71 (0.53, 0.95) | 0.83 (0.55, 1.25) | x | x | x | x |
| **25 - 29.9** | 0.52 (0.32, 0.85) | x | 0.6 (0.42, 0.86) | 0.71 (0.45, 1.13) | x | x | x | x |

| OST cohort<br>Predictor | Women<br>OR (95%CI) | | | | Men<br>OR (95%CI) | | | |
|---|---|---|---|---|---|---|---|---|
| | One year | | Two year | | One year | | Two year | |
| | MACE | Stroke/MI | MACE | Stroke/MI | MACE | Stroke/MI | MACE | Stroke/MI |
| **30 - 39.9** | 0.42 (0.23, 0.76) | x | 0.51 (0.29, 0.9) | 0.7 (0.37, 1.35) | x | x | x | x |
| **>=40** | 0.52 (0.14, 1.94) | x | 0.9 (0.37, 2.16) | 0.95 (0.37, 2.45) | x | x | x | x |
| **No. of GP visits**** | | | x | | x | x | x | x |
| **0** | ref | ref | x | ref | x | x | x | x |
| **1-5** | 0.73 (0.44, 1.22) | 0.88 (0.49, 1.56) | x | 0.66 (0.43, 1) | x | x | x | x |
| **6-10** | 0.57 (0.33, 1) | 0.76 (0.41, 1.41) | x | 0.63 (0.4, 0.97) | x | x | x | x |
| **11-15** | 0.6 (0.34, 1.06) | 0.78 (0.41, 1.46) | x | 0.65 (0.41, 1.02) | x | x | x | x |
| **>=16** | 0.64 (0.36, 1.13) | 0.9 (0.48, 1.68) | x | 0.73 (0.47, 1.14) | x | x | x | x |
| **No. of GP emergency visits**** | | x | | x | x | x | x | x |
| **0** | ref | x | ref | x | x | x | x | x |
| **1** | 1.59 (1.23, 2.06) | x | 1.01 (0.81, 1.25) | x | x | x | x | x |
| **2** | 1.67 (1.17, 2.37) | x | 1.15 (0.86, 1.53) | x | x | x | x | x |
| **3-5** | 1.46 (1.01, 2.1) | x | 1.37 (1.04, 1.8) | x | x | x | x | x |
| **>=6** | 2.09 (1.31, 3.34) | x | 1.91 (1.34, 2.71) | x | x | x | x | x |
| **eGFR**** | | | | | x | x | | |
| **<=29** | ref | ref | ref | ref | x | x | ref | ref |
| **30 – 44** | 0.6 (0.3, 1.22) | 0.71 (0.33, 1.55) | 0.98 (0.54, 1.76) | 0.72 (0.4, 1.29) | x | x | 1.96 (0.26, 14.93) | 1.47 (0.19, 11.23) |
| **45 – 59** | 0.56 (0.29, 1.1) | 0.59 (0.29, 1.19) | 0.77 (0.43, 1.38) | 0.61 (0.35, 1.08) | x | x | 1.44 (0.21, 9.73) | 0.96 (0.14, 6.55) |
| **60 – 89** | 0.44 (0.23, 0.84) | 0.46 (0.23, 0.95) | 0.62 (0.34, 1.15) | 0.49 (0.28, 0.84) | x | x | 1.17 (0.19, 7.38) | 0.74 (0.11, 4.75) |
| **>=90** | 0.47 (0.17, 1.28) | 0.55 (0.2, 1.56) | 0.67 (0.29, 1.56) | 0.52 (0.23, 1.16) | x | x | 1.19 (0.17, 8.26) | 0.59 (0.08, 4.5) |
| **SBP**** | | x | | x | x | x | x | x |
| **<120** | ref | x | ref | x | x | x | x | x |
| **120 - 139** | 1.09 (0.78, 1.52) | x | 0.96 (0.75, 1.22) | x | x | x | x | x |
| **140 - 159** | 1.21 (0.86, 1.7) | x | 1.21 (0.94, 1.57) | x | x | x | x | x |
| **>=160** | 1.3 (0.87, 1.94) | x | 1.32 (0.97, 1.81) | x | x | x | x | x |
| **DBP**** | x | x | | x | x | x | x | x |
| **<80** | x | x | ref | x | x | x | x | x |
| **80 – 89** | x | x | 1.08 (0.9, 1.29) | x | x | x | x | x |
| **90 – 99** | x | x | 1.02 (0.76, 1.36) | x | x | x | x | x |
| **>=100** | x | x | 1.26 (0.76, 2.07) | x | x | x | x | x |
| **No. of concomitant medicines**** | | | | | x | | x | x |
| **0** | ref | ref | ref | ref | x | ref | x | x |
| **1 – 3** | 0.54 (0.32, 0.94) | 0.72 (0.4, 1.28) | 0.64 (0.46, 0.89) | 0.84 (0.54, 1.31) | x | 0.35 (0.09, 1.42) | x | x |
| **4 – 6** | 0.72 (0.43, 1.22) | 0.68 (0.38, 1.22) | 0.66 (0.48, 0.91) | 0.84 (0.54, 1.32) | x | 0.9 (0.29, 2.75) | x | x |

| OST cohort<br>Predictor | Women<br>OR (95%CI) | | | | Men<br>OR (95%CI) | | | |
|---|---|---|---|---|---|---|---|---|
| | One year | | Two year | | One year | | Two year | |
| | MACE | Stroke/MI | MACE | Stroke/MI | MACE | Stroke/MI | MACE | Stroke/MI |
| **7 – 9** | 0.81 (0.47, 1.4) | 0.81 (0.44, 1.47) | 0.74 (0.53, 1.02) | 0.91 (0.57, 1.44) | x | 1.66 (0.57, 4.83) | x | x |
| **10 – 12** | 0.95 (0.54, 1.67) | 0.82 (0.44, 1.54) | 0.86 (0.61, 1.21) | 1.05 (0.65, 1.69) | x | 0.95 (0.3, 2.96) | x | x |
| **>=13** | 0.95 (0.53, 1.69) | 1.03 (0.55, 1.91) | 0.82 (0.57, 1.16) | 1.1 (0.68, 1.78) | x | 1.9 (0.67, 5.42) | x | x |
| **Cholesterol measurement** (HDL/LDL)** | | x | | | x | x | x | x |
| **<=3.5** | ref | x | ref | ref | x | x | x | x |
| **3.6 – 5** | 1.37 (0.92, 2.03) | x | 1.21 (0.93, 1.58) | | x | x | x | x |
| **>5** | 1.8 (1.1, 2.94) | x | 1.4 (0.95, 2.08) | | x | x | x | x |
| **No. of previous fractures*** | | | x | | x | x | x | x |
| **0** | ref | ref | x | ref | x | x | x | x |
| **1** | 1.15 (0.78, 1.69) | 1.12 (0.87, 1.46) | x | | x | x | x | x |
| **>=2** | 0.89 (0.6, 1.32) | 0.92 (0.69, 1.21) | x | | x | x | x | x |

**Abbreviations**: OST, patients with incident diagnosis of osteoporosis; IFX, patients with incident fragility fracture; OBP, incident users of oral bisphosphonates; OR, odds ratio; CI, confidence intervals; MACE, composite outcome for the occurrence of either myocardial infarction, stroke or cardiovascular disease death; MI, myocardial infarction; * ever; ** in the year prior to start; SES, socio-economic status; BMI, body mass index; eGFR, estimated Glomerular Filtration Rate; SBP, cholesterol, systolic blood pressure; DBP, diastolic blood pressure.

#### Table S6b. Risk factors selected by lasso for one- and two-year models in gender based models (IFX cohort)

| IFX cohort<br>Predictor | Women<br>OR (95%CI) | | | | Men<br>OR (95%CI) | | | |
|---|---|---|---|---|---|---|---|---|
| | One year | | Two year | | One year | | Two year | |
| | MACE | Stroke/MI | MACE | Stroke/MI | MACE | Stroke/MI | MACE | Stroke/MI |
| **SES** | x | x | x | x | x | x | x | x |
| **1** | x | x | x | x | x | x | x | x |
| **2** | x | x | x | x | x | x | x | x |
| **3** | x | x | x | x | x | x | x | x |
| **4** | x | x | x | x | x | x | x | x |
| **5** | x | x | x | x | x | x | x | x |
| **Smoking**** | x | x | x | x | | x | x | x |
| **Ex** | x | x | x | x | ref | x | x | x |
| **No** | x | x | x | x | 1.08 (0.84, 1.4) | x | x | x |
| **Yes** | x | x | x | x | 1.02 (0.74, 1.4) | x | x | x |
| **Drinking**** | | x | | | | x | | x |
| **Ex** | ref | x | ref | ref | ref | x | ref | x |

| IFX cohort Predictor | Women OR (95%CI) | | | | Men OR (95%CI) | | | |
|---|---|---|---|---|---|---|---|---|
| | One year | | Two year | | One year | | Two year | |
| | MACE | Stroke/MI | MACE | Stroke/MI | MACE | Stroke/MI | MACE | Stroke/MI |
| No | 0.93 (0.67, 1.29) | x | 0.88 (0.69, 1.13) | 0.89 (0.66, 1.21) | 1.15 (0.69, 1.9) | x | 1.15 (0.78, 1.7) | x |
| Yes | 0.83 (0.59, 1.18) | x | 0.75 (0.58, 0.97) | 0.8 (0.59, 1.09) | 1.01 (0.65, 1.58) | x | 0.89 (0.61, 1.3) | x |
| Diabetes type I* | x | x | x | x | x | x | x | x |
| Diabetes type II* | x | x | x | 1.37 (1.1, 1.69) | x | x | x | x |
| Chronic obstructive pulmonary disease* | x | x | x | x | x | x | x | x |
| Chronic kidney disease* | x | x | x | x | x | x | x | x |
| Rheumatoid arthritis* | x | x | x | x | x | x | x | x |
| Lupus* | x | x | x | x | x | x | x | x |
| Systemic heart disease** | x | x | x | x | x | x | x | x |
| Anti-osteoporosis use** | x | x | x | x | x | x | x | x |
| Heparin use** | x | x | x | x | x | x | x | x |
| Beta-blocker use** | 1.02 (0.88, 1.2) | 1.03 (0.85, 1.25) | x | 1.11 (0.95, 1.29) | x | x | x | x |
| Hypertension** | x | x | x | x | x | x | x | x |
| Deep vein thrombosis or pulmonary embolism** | x | x | x | x | x | x | x | x |
| Anticoagulant use** | x | x | x | x | x | x | x | x |
| Antidepressants TCA** | x | x | x | x | x | x | x | x |
| Antidepressants SSRI** | x | x | x | 0.73 (0.6, 0.89) | x | x | x | x |
| Hypercholesterolemia** | x | x | x | x | x | x | x | x |
| Statin use** | x | x | x | x | x | x | x | x |
| Osteoporosis history* | x | x | x | x | x | x | x | x |
| Family history of cardiovascular disease (%) | x | x | x | x | x | x | x | x |
| Family history of cardiovascular disease before age 60 | x | x | x | x | x | x | x | x |
| Heart failure* | 1.18 (0.94, 1.47) | x | x | x | x | x | x | x |
| Migraine* | x | x | x | x | x | x | x | x |
| Severe mental illness* | x | x | x | x | x | x | x | x |
| Vascular Disease* | X | x | x | x | x | x | x | x |
| Atrial fibrillation* | 1.22 (1, 1.47) | 1.48 (1.18, 1.85) | x | 1.27 (1.06, 1.54) | x | x | x | x |
| On anti-hypertensive drug | 1.06 (0.9, 1.25) | 1.03 (0.84, 1.26) | 1.09 (0.95, 1.25) | 1.19 (1, 1.42) | 1.31 (1.04, 1.64) | 1.46 (1.08, 1.96) | 1.5 (1.24, 1.8) | 1.51 (1.2, 1.91) |
| Antipsychotic use** | x | x | x | x | x | x | x | x |
| Steroid use** | x | x | x | x | x | x | x | x |

| IFX cohort Predictor | Women OR (95%CI) | | | | Men OR (95%CI) | | | |
|---|---|---|---|---|---|---|---|---|
| | One year | | Two year | | One year | | Two year | |
| | MACE | Stroke/MI | MACE | Stroke/MI | MACE | Stroke/MI | MACE | Stroke/MI |
| **Erectile dysfunction**** | x | x | x | x | x | x | x | x |
| **Age Group (%)** | | | | | | | | |
| **50-59** | ref | ref | ref | ref | ref | ref | ref | ref |
| **60-69** | 5.82 (2.09, 16.18) | 6.29 (1.94, 20.39) | 2.15 (1.21, 3.81) | 2.32 (1.2, 4.47) | 2.75 (1.27, 5.99) | 2.64 (1.15, 6.04) | 1.81 (1.12, 2.92) | 2.77 (1.5, 5.13) |
| **70-79** | 10.94 (4.05, 29.55) | 10.76 (3.42, 33.79) | 5.3 (3.1, 9.05) | 5.05 (2.74, 9.32) | 5.3 (2.55, 11.01) | 3.55 (1.61, 7.83) | 2.44 (1.56, 3.81) | 3.18 (1.77, 5.72) |
| **80-89** | 16.19 (6.02, 43.53) | 15.01 (4.8, 46.97) | 8.57 (4.98, 14.76) | 7.25 (3.94, 13.33) | 6.76 (3.27, 13.96) | 4.07 (1.88, 8.83) | 3.62 (2.35, 5.55) | 3.62 (2.04, 6.43) |
| **>89** | 23.97 (8.88, 64.74) | 15.62 (4.95, 49.32) | 9.87 (5.64, 17.25) | 6.76 (3.63, 12.61) | 7.31 (3.45, 15.49) | 3.52 (1.54, 8.04) | 3.56 (2.26, 5.62) | 2.89 (1.55, 5.39) |
| **Charlson score** | x | x | x | x | x | x | x | x |
| **0** | x | x | x | x | x | x | x | x |
| **1** | x | x | x | x | x | x | x | x |
| **2** | x | x | x | x | x | x | x | x |
| **≥3** | x | x | x | x | x | x | x | x |
| **Cardiovascular disease** | x | x | x | | x | x | x | x |
| **No** | x | x | x | ref | x | x | x | x |
| **Ever >1 year before index date** | x | x | x | 1.13 (0.94, 1.35) | x | x | x | x |
| **1 year before index** | x | x | x | 1.2 (0.75, 1.9) | x | x | x | x |
| **6 months before index** | x | x | x | 1.2 (0.78, 1.84) | x | x | x | x |
| **1 month before index** | x | x | x | 2.44 (1.26, 4.74) | x | x | x | x |
| **MI or Stroke** | | x | x | x | x | x | x | x |
| **No** | ref | x | x | x | x | x | x | x |
| **Ever >1 year before index date** | 1.13 (0.89, 1.45) | x | x | x | x | x | x | x |
| **1 year before index** | 1.47 (1.09, 1.98) | x | x | x | x | x | x | x |
| **Established CVD *** | 1.53 (1.26, 1.86) | 1.67 (1.41, 1.98) | 1.93 (1.71, 2.18) | 1.62 (1.4, 1.88) | 1.9 (1.56, 2.32) | 1.93 (1.49, 2.51) | 1.86 (1.57, 2.2) | 1.77 (1.43, 2.19) |
| **Any fracture history** | x | x | x | x | x | x | x | x |
| **No** | x | x | x | x | x | x | x | x |
| **Ever >1 year before index date** | x | x | x | x | x | x | x | x |
| **1 year before index** | x | x | x | x | x | x | x | x |
| **Hip fracture history** | x | x | x | x | x | x | x | x |

| IFX cohort Predictor | Women OR (95%CI) | | | | Men OR (95%CI) | | | |
|---|---|---|---|---|---|---|---|---|
| | One year | | Two year | | One year | | Two year | |
| | MACE | Stroke/MI | MACE | Stroke/MI | MACE | Stroke/MI | MACE | Stroke/MI |
| **No** | x | x | x | x | x | x | x | x |
| **Ever >1 year before index date** | x | x | x | x | x | x | x | x |
| **1 year before index** | x | x | x | x | x | x | x | x |
| **Shoulder fracture history** | x | x | x | x | x | x | x | x |
| **No** | x | x | x | x | x | x | x | x |
| **Ever >1 year before index date** | x | x | x | x | x | x | x | x |
| **1 year before index** | x | x | x | x | x | x | x | x |
| **Spine fracture history** | x | x | x | x | x | x | x | x |
| **No** | x | x | x | x | x | x | x | x |
| **Ever >1 year before index date** | x | x | x | x | x | x | x | x |
| **1 year before index** | x | x | x | x | x | x | x | x |
| **Wrist fracture history** | x | x | x | x | x | x | x | x |
| **No** | x | x | x | x | x | x | x | x |
| **Ever >1 year before index date** | x | x | x | x | x | x | x | x |
| **1 year before index** | x | x | x | x | x | x | x | x |
| **BMI**** | x | x | x | | x | x | x | x |
| **<18.5** | x | x | x | ref | x | x | x | x |
| **18.6 - 24.9** | x | x | x | 1.03 (0.8, 1.32) | x | x | x | x |
| **25 - 29.9** | x | x | x | 1.01 (0.73, 1.4) | x | x | x | x |
| **30 - 39.9** | x | x | x | 1.17 (0.77, 1.8) | x | x | x | x |
| **>=40** | x | x | x | 1.43 (0.57, 3.61) | x | x | x | x |
| **No. of GP visits**** | | x | x | x | x | x | x | x |
| **0** | ref | x | x | x | x | x | x | x |
| **1-5** | 0.85 (0.62, 1.18) | x | x | x | x | x | x | x |
| **6-10** | 0.76 (0.54, 1.07) | x | x | x | x | x | x | x |
| **11-15** | 0.77 (0.54, 1.1) | x | x | x | x | x | x | x |
| **>=16** | 0.85 (0.6, 1.22) | x | x | x | x | x | x | x |
| **No. of GP emergency visits**** | x | x | x | x | x | x | x | x |
| **0** | x | x | x | x | x | x | x | x |
| **1** | x | x | x | x | x | x | x | x |

| IFX cohort<br>Predictor | Women<br>OR (95%CI) | | | | Men<br>OR (95%CI) | | | |
|---|---|---|---|---|---|---|---|---|
| | One year | | Two year | | One year | | Two year | |
| | MACE | Stroke/MI | MACE | Stroke/MI | MACE | Stroke/MI | MACE | Stroke/MI |
| **2** | x | x | x | x | x | x | x | x |
| **3-5** | x | x | x | x | x | x | x | x |
| **>=6** | x | x | | x | x | x | x | x |
| **eGFR**** | | | | | | x | x | x |
| **<=29** | ref | ref | ref | ref | ref | x | x | x |
| **30 – 44** | 0.88 (0.56, 1.39) | 0.89 (0.53, 1.49) | 1.1 (0.71, 1.69) | 1 (0.64, 1.56) | 0.91 (0.46, 1.78) | x | x | x |
| **45 – 59** | 0.8 (0.49, 1.32) | 0.78 (0.44, 1.38) | 0.92 (0.61, 1.39) | 0.92 (0.57, 1.47) | 0.74 (0.42, 1.32) | x | x | x |
| **60 – 89** | 0.72 (0.41, 1.25) | 0.69 (0.32, 1.47) | 0.91 (0.6, 1.38) | 0.91 (0.54, 1.54) | 0.69 (0.37, 1.29) | x | x | x |
| **>=90** | 0.7 (0.31, 1.59) | 0.82 (0.32, 2.09) | 1.03 (0.51, 2.08) | 1 (0.48, 2.1) | 0.49 (0.16, 1.47) | x | x | x |
| **SBP**** | | | | | x | | x | |
| **<120** | ref | ref | ref | ref | x | ref | x | ref |
| **120 - 139** | 1.17 (0.94, 1.45) | 1.27 (0.96, 1.7) | 1.03 (0.87, 1.23) | 1.01 (0.82, 1.25) | x | 0.79 (0.53, 1.18) | x | |
| **140 - 159** | 1.37 (1.09, 1.72) | 1.4 (1.04, 1.88) | 1.08 (0.89, 1.3) | 1.13 (0.91, 1.41) | x | 1.42 (0.96, 2.09) | x | |
| **>=160** | 1.57 (1.21, 2.03) | 1.71 (1.23, 2.38) | 1.37 (1.11, 1.7) | 1.52 (1.18, 1.97) | x | 1.82 (1.11, 2.99) | x | |
| **DBP**** | x | x | x | x | x | x | x | x |
| **<80** | x | x | x | x | x | x | x | x |
| **80 – 89** | x | x | x | x | x | x | x | x |
| **90 – 99** | x | x | x | x | x | x | x | x |
| **>=100** | x | x | x | x | x | x | x | x |
| **No. of concomitant medicines**** | | | | | x | x | x | x |
| **0** | ref | ref | ref | ref | x | x | x | x |
| **1 – 3** | 1.17 (0.82, 1.66) | 0.9 (0.66, 1.21) | 1.04 (0.86, 1.26) | 0.8 (0.64, 1.01) | x | x | x | x |
| **4 – 6** | 1.41 (0.99, 2.02) | 1.19 (0.91, 1.56) | 1.29 (1.08, 1.54) | 0.86 (0.69, 1.06) | x | x | x | x |
| **7 – 9** | 1.51 (1.05, 2.18) | 1.15 (0.86, 1.52) | 1.33 (1.11, 1.59) | 1.02 (0.82, 1.27) | x | x | x | x |
| **10 – 12** | 1.56 (1.06, 2.28) | 1.37 (1.01, 1.84) | 1.39 (1.14, 1.69) | 1.06 (0.84, 1.34) | x | x | x | x |
| **>=13** | 1.75 (1.19, 2.57) | 1.49 (1.1, 2.02) | 1.39 (1.14, 1.69) | 1 (0.79, 1.27) | x | x | x | x |
| **Cholesterol measurement**** **(HDL/LDL)** | x | x | | x | x | x | x | x |
| **<=3.5** | x | x | ref | x | x | x | x | x |
| **3.6 – 5** | x | x | 1.24 (0.76, 2) | x | x | x | x | x |
| **>5** | x | x | 1.39 (0.44, 4.4) | x | x | x | x | x |
| **No. of previous fractures*** | x | x | x | | x | x | x | x |
| **0** | x | x | x | ref | x | x | x | x |
| **1** | x | x | x | | x | x | x | x |

| IFX cohort Predictor | Women OR (95%CI) | | | | Men OR (95%CI) | | | |
|---|---|---|---|---|---|---|---|---|
| | One year | | Two year | | One year | | Two year | |
| | MACE | Stroke/MI | MACE | Stroke/MI | MACE | Stroke/MI | MACE | Stroke/MI |
| >=2 | x | x | x | | x | x | x | x |

**Abbreviations**: OST, patients with incident diagnosis of osteoporosis; IFX, patients with incident fragility fracture; OBP, incident users of oral bisphosphonates; OR, odds ratio; CI, confidence intervals; MACE, composite outcome for the occurrence of either myocardial infarction, stroke or cardiovascular disease death; MI, myocardial infarction; * ever; ** in the year prior to start; SES, socio-economic status; BMI, body mass index; eGFR, estimated Glomerular Filtration Rate; SBP, cholesterol, systolic blood pressure; DBP, diastolic blood pressure.

#### Table S6c. Risk factors selected by lasso for one- and two-year models in gender based models (OBP cohort)

| OBP cohort Predictor | Women OR (95%CI) | | | | Men OR (95%CI) | | | |
|---|---|---|---|---|---|---|---|---|
| | One year | | Two year | | One year | | Two year | |
| | MACE | Stroke/MI | MACE | Stroke/MI | MACE | Stroke/MI | MACE | Stroke/MI |
| **SES** | | | | | x | x | | x |
| **1** | ref | ref | ref | x | x | x | ref | x |
| **2** | 0.99 (0.84, 1.17) | 1.14 (0.94, 1.38) | 1.07 (0.95, 1.22) | x | x | x | 1.11 (0.9, 1.37) | x |
| **3** | 1.18 (1, 1.39) | 1.34 (1.11, 1.61) | 1.25 (1.1, 1.42) | x | x | x | 1.09 (0.88, 1.35) | x |
| **4** | 1.14 (0.95, 1.36) | 1.26 (1.03, 1.54) | 1.19 (1.04, 1.37) | x | x | x | 1.26 (1.01, 1.56) | x |
| **5** | 1.27 (1.05, 1.54) | 1.24 (0.99, 1.56) | 1.2 (1.03, 1.4) | x | x | x | 1.1 (0.85, 1.43) | x |
| **Smoking**** | x | | | | x | | x | x |
| **Ex** | x | ref | ref | ref | x | ref | x | x |
| **No** | x | 0.83 (0.61, 1.13) | 0.88 (0.74, 1.04) | 0.91 (0.74, 1.11) | x | 0.97 (0.68, 1.41) | x | x |
| **Yes** | x | 0.91 (0.67, 1.24) | 1.09 (0.88, 1.35) | 1.23 (0.96, 1.58) | x | 1.17 (0.78, 1.75) | x | x |
| **Drinking**** | | | | | x | x | | x |
| **Ex** | ref | ref | ref | ref | x | x | ref | x |
| **No** | 0.95 (0.64, 1.42) | 1.05 (0.69, 1.6) | 0.88 (0.65, 1.2) | 0.9 (0.67, 1.2) | x | x | 0.84 (0.58, 1.22) | x |
| **Yes** | 0.82 (0.57, 1.19) | 0.87 (0.59, 1.28) | 0.78 (0.57, 1.05) | 0.78 (0.59, 1.04) | x | x | 0.81 (0.58, 1.14) | x |

| OBP cohort<br>Predictor | Women<br>OR (95%CI) | | | | Men<br>OR (95%CI) | | | |
|---|---|---|---|---|---|---|---|---|
| | One year | | Two year | | One year | | Two year | |
| | MACE | Stroke/MI | MACE | Stroke/MI | MACE | Stroke/MI | MACE | Stroke/MI |
| **Diabetes type I*** | x | x | x | x | x | x | x | x |
| **Diabetes type II*** | 1.45 (1.15, 1.84) | 1.49 (1.18, 1.9) | 1.36 (1.14, 1.63) | x | x | x | x | x |
| **Chronic obstructive pulmonary disease*** | x | 1.26 (0.97, 1.64) | x | x | x | x | x | x |
| **Chronic kidney disease*** | 0.81 (0.61, 1.08) | x | 0.86 (0.7, 1.07) | x | x | x | x | x |
| **Rheumatoid arthritis*** | x | x | 1 (0.86, 1.16) | x | 0.57 (0.42, 0.76) | 0.73 (0.52, 1.02) | x | x |
| **Lupus*** | x | x | x | x | x | x | x | x |
| **Systemic heart disease**** | x | x | x | x | x | x | x | x |
| **Anti-osteoporosis use**** | x | x | x | x | x | x | x | x |
| **Heparin use**** | x | x | x | x | x | x | x | x |
| **Beta-blocker use**** | 1.08 (0.93, 1.25) | 1.25 (1.06, 1.48) | 1.19 (1.06, 1.34) | 1.17 (1.03, 1.32) | x | 1.04 (0.8, 1.37) | x | x |
| **Hypertension**** | x | x | x | x | x | x | x | x |
| **Deep vein thrombosis or pulmonary embolism**** | x | x | x | x | x | x | x | x |
| **Anticoagulant use**** | x | x | x | x | x | x | x | x |
| **Antidepressants TCA**** | x | x | x | x | x | x | x | x |
| **Antidepressants SSRI**** | x | 1.29 (1.06, 1.56) | 1.16 (1.01, 1.34) | x | x | x | x | x |
| **Hypercholesterolemia**** | x | x | x | x | x | x | x | x |
| **Statin use**** | x | 0.84 (0.69, 1.01) | x | x | x | x | x | x |
| **Osteoporosis history*** | 0.76 (0.66, 0.86) | 0.77 (0.66, 0.89) | 0.77 (0.7, 0.85) | 0.85 (0.76, 0.95) | 0.84 (0.66, 1.06) | x | 0.83 (0.69, 1.01) | 0.82 (0.66, 1.01) |
| **Family history of cardiovascular disease** | x | x | 0.99 (0.84, 1.16) | x | x | x | x | x |
| **Family history of cardiovascular disease before age 60** | x | x | x | x | x | x | x | x |
| **Heart failure*** | x | x | x | x | x | 1.43 (1.01, 2.05) | x | x |
| **Migraine*** | x | x | x | x | x | x | | x |
| **Severe mental illness*** | x | x | x | x | x | x | x | x |
| **Vascular Disease*** | x | x | x | x | x | x | x | x |

| OBP cohort<br>Predictor | Women<br>OR (95%CI) | | | | Men<br>OR (95%CI) | | | |
|---|---|---|---|---|---|---|---|---|
| | One year | | Two year | | One year | | Two year | |
| | MACE | Stroke/MI | MACE | Stroke/MI | MACE | Stroke/MI | MACE | Stroke/MI |
| **Atrial fibrillation*** | 1.57 (1.3, 1.88) | 1.45 (1.17, 1.78) | 1.52 (1.31, 1.76) | 1.4 (1.19, 1.64) | x | x | x | x |
| **On anti-hypertensive drug** | x | 1.14 (0.94, 1.39) | 1.09 (0.96, 1.25) | 1.17 (1.01, 1.36) | x | x | 1.14 (0.93, 1.4) | 1.11 (0.88, 1.39) |
| **Antipsychotic use**** | x | x | x | x | x | x | x | x |
| **Steroid use**** | x | 1.05 (0.89, 1.25) | x | x | x | 0.99 (0.77, 1.26) | 0.89 (0.75, 1.06) | x |
| **Erectile dysfunction**** | x | x | x | x | x | x | x | x |
| **Age Group** | | | | | | | | |
| **50-59** | ref | ref | ref | ref | ref | ref | ref | ref |
| **60-69** | 2.31 (1.42, 3.76) | 1.86 (1.19, 2.9) | 1.62 (1.17, 2.23) | 1.8 (1.27, 2.55) | 1.58 (0.95, 2.62) | 1.26 (0.77, 2.06) | 1.39 (0.95, 2.03) | 1.2 (0.81, 1.78) |
| **70-79** | 4.33 (2.72, 6.88) | 2.68 (1.75, 4.12) | 3.3 (2.44, 4.47) | 3.4 (2.44, 4.75) | 2.39 (1.49, 3.85) | 1.48 (0.92, 2.38) | 1.95 (1.37, 2.8) | 1.96 (1.36, 2.84) |
| **80-89** | 8.6 (5.38, 13.74) | 4.91 (3.18, 7.6) | 6.33 (4.65, 8.62) | 5.76 (4.09, 8.1) | 2.8 (1.73, 4.53) | 1.81 (1.11, 2.95) | 2.53 (1.76, 3.65) | 2.01 (1.38, 2.95) |
| **>89** | 9.96 (6.07, 16.35) | 5.22 (3.23, 8.43) | 8.2 (5.88, 11.45) | 6.35 (4.37, 9.23) | 5.41 (3.2, 9.17) | 2.27 (1.25, 4.12) | 3.3 (2.17, 5.02) | 2.35 (1.49, 3.72) |
| **Charlson score** | | x | | x | x | | | x |
| **0** | ref | x | ref | x | x | ref | ref | x |
| **1** | 0.85 (0.72, 1.01) | x | 0.94 (0.82, 1.06) | x | x | 1.02 (0.75, 1.38) | 0.89 (0.73, 1.08) | x |
| **2** | 0.9 (0.73, 1.1) | x | 0.97 (0.83, 1.13) | x | x | 1.09 (0.77, 1.53) | 0.78 (0.62, 0.99) | x |
| **≥3** | 0.98 (0.77, 1.25) | x | 0.97 (0.79, 1.18) | x | x | 1.08 (0.75, 1.56) | 0.93 (0.74, 1.18) | x |
| **Cardiovascular disease** | | | x | | x | | x | |
| **No** | ref | ref | x | ref | x | ref | x | ref |
| **Ever >1 year before index date** | 1.15 (0.95, 1.38) | 1.31 (1.06, 1.61) | x | 1.38 (1.19, 1.61) | x | 1.42 (1.05, 1.93) | x | 1.26 (1, 1.58) |
| **1 year before index** | 1.11 (0.73, 1.69) | 0.98 (0.6, 1.61) | x | 1.09 (0.74, 1.59) | x | 1.83 (1.03, 3.27) | x | 1.17 (0.69, 2) |
| **6 months before index** | 1.24 (0.85, 1.82) | 1.5 (1, 2.24) | x | 1.5 (1.09, 2.08) | x | 3.03 (1.9, 4.85) | x | 2.69 (1.86, 3.88) |

| OBP cohort<br>Predictor | Women<br>OR (95%CI) | | | | Men<br>OR (95%CI) | | | |
|---|---|---|---|---|---|---|---|---|
| | One year | | Two year | | One year | | Two year | |
| | MACE | Stroke/MI | MACE | Stroke/MI | MACE | Stroke/MI | MACE | Stroke/MI |
| 1 month before index | 2.15 (1.32, 3.49) | 2.84 (1.75, 4.61) | x | 2.14 (1.35, 3.38) | x | 3.66 (1.91, 7.03) | x | 2.51 (1.39, 4.53) |
| **MI or Stroke** | | | x | | x | | | x |
| **No** | ref | ref | ref | ref | x | ref | ref | x |
| **Ever >1 year before index date** | 1.31 (1.02, 1.68) | 1.03 (0.77, 1.38) | 1.4 (1.15, 1.69) | 1.28 (1.03, 1.58) | x | 0.76 (0.5, 1.17) | 1.35 (1.02, 1.78) | x |
| **1 year before index** | 2.09 (1.59, 2.77) | 2.15 (1.57, 2.95) | 1.98 (1.59, 2.47) | 1.64 (1.27, 2.12) | x | 1.88 (1.19, 2.98) | 2.33 (1.68, 3.23) | x |
| **Established CVD *** | 1.58 (1.28, 1.96) | 1.41 (1.11, 1.79) | 1.64 (1.4, 1.91) | 1.37 (1.15, 1.64) | 2.3 (1.89, 2.8) | 1.38 (0.98, 1.94) | 1.71 (1.35, 2.16) | 1.49 (1.22, 1.82) |
| **Any fracture history** | x | x | x | x | x | x | x | x |
| **No** | x | x | x | x | x | x | x | x |
| **Ever >1 year before index date** | x | x | x | x | x | x | x | x |
| **1 year before index** | x | x | x | x | x | x | x | x |
| **Hip fracture history** | x | x | x | x | x | x | | x |
| **No** | x | x | x | x | x | x | ref | x |
| **Ever >1 year before index date** | x | x | x | x | x | x | 0.9 (0.43, 1.87) | x |
| **1 year before index** | x | x | x | x | x | x | 1.18 (0.91, 1.54) | x |
| **Shoulder fracture history** | x | x | x | x | x | x | x | x |
| **No** | x | x | x | x | x | x | x | x |
| **Ever >1 year before index date** | x | x | x | x | x | x | x | x |
| **1 year before index** | x | x | x | x | x | x | x | x |
| **Spine fracture history** | x | x | x | x | x | x | x | x |
| **No** | x | x | x | x | x | x | x | x |
| **Ever >1 year before index date** | x | x | x | x | x | x | x | x |
| **1 year before index** | x | x | x | x | x | x | x | x |
| **Wrist fracture history** | x | x | x | x | x | x | x | x |
| **No** | x | x | x | x | x | x | x | x |
| **Ever >1 year before index date** | x | x | x | x | x | x | x | x |

| OBP cohort Predictor | Women OR (95%CI) | | | | Men OR (95%CI) | | | |
|---|---|---|---|---|---|---|---|---|
| | One year | | Two year | | One year | | Two year | |
| | MACE | Stroke/MI | MACE | Stroke/MI | MACE | Stroke/MI | MACE | Stroke/MI |
| **1 year before index** | x | x | x | x | x | x | x | x |
| **BMI** | | | | | x | | | |
| **<18.5** | ref | ref | ref | ref | x | ref | ref | ref |
| **18.6 - 24.9** | 0.72 (0.54, 0.96) | 0.78 (0.58, 1.04) | 0.75 (0.61, 0.91) | 0.79 (0.64, 0.97) | x | 1.11 (0.59, 2.07) | 0.83 (0.58, 1.2) | 0.92 (0.6, 1.39) |
| **25 - 29.9** | 0.58 (0.4, 0.84) | 0.68 (0.47, 0.98) | 0.58 (0.44, 0.75) | 0.67 (0.51, 0.88) | x | 0.87 (0.44, 1.74) | 0.72 (0.48, 1.08) | 0.82 (0.53, 1.27) |
| **30 - 39.9** | 0.54 (0.34, 0.86) | 0.59 (0.37, 0.93) | 0.58 (0.43, 0.78) | 0.68 (0.49, 0.93) | x | 0.78 (0.39, 1.57) | 0.57 (0.37, 0.9) | 0.76 (0.47, 1.23) |
| **>=40** | 0.42 (0.17, 1.04) | 0.59 (0.25, 1.42) | 0.61 (0.32, 1.15) | 0.73 (0.37, 1.44) | x | 0.89 (0.24, 3.39) | 0.81 (0.31, 2.14) | 0.83 (0.29, 2.34) |
| **No. of GP visits** | | | x | | x | | | |
| **0** | ref | ref | x | ref | x | ref | ref | ref |
| **1-5** | 0.95 (0.76, 1.2) | 0.85 (0.65, 1.12) | x | 0.91 (0.74, 1.12) | x | 0.52 (0.3, 0.9) | 1.06 (0.76, 1.5) | 0.97 (0.65, 1.44) |
| **6-10** | 0.84 (0.64, 1.11) | 0.87 (0.63, 1.2) | x | 0.84 (0.66, 1.06) | x | 0.56 (0.31, 1.04) | 0.88 (0.59, 1.31) | 0.69 (0.44, 1.08) |
| **11-15** | 0.7 (0.52, 0.94) | 0.79 (0.56, 1.11) | x | 0.77 (0.6, 0.99) | x | 0.55 (0.29, 1.03) | 0.89 (0.59, 1.34) | 0.75 (0.47, 1.19) |
| **>=16** | 0.79 (0.59, 1.06) | 0.86 (0.62, 1.2) | x | 0.88 (0.68, 1.12) | x | 0.69 (0.37, 1.27) | 1.14 (0.76, 1.7) | 0.87 (0.55, 1.36) |
| **No. of GP emergency visits** | | | | x | x | x | x | x |
| **0** | ref | ref | ref | x | x | x | x | x |
| **1** | 1.09 (0.91, 1.31) | 1.09 (0.89, 1.33) | 1.17 (1.02, 1.34) | x | x | x | x | x |
| **2** | 1.32 (1.03, 1.7) | 1.36 (1.05, 1.78) | 1.33 (1.11, 1.61) | x | x | x | x | x |
| **3-5** | 1.39 (1.09, 1.78) | 1.09 (0.82, 1.45) | 1.19 (0.99, 1.44) | x | x | x | x | x |
| **>=6** | 2.41 (1.8, 3.24) | 1.7 (1.2, 2.43) | 1.64 (1.26, 2.12) | x | x | x | x | x |
| **eGFR** | | | | | | | | |
| **<=29** | ref | ref | ref | ref | ref | ref | ref | ref |
| **30 – 44** | 0.89 (0.51, 1.54) | 0.94 (0.49, 1.82) | 0.93 (0.6, 1.44) | 0.88 (0.54, 1.42) | 1.43 (0.5, 4.1) | 1.21 (0.35, 4.21) | 1.38 (0.6, 3.17) | 1.57 (0.53, 4.68) |

| OBP cohort<br>Predictor | Women<br>OR (95%CI) | | | | Men<br>OR (95%CI) | | | |
|---|---|---|---|---|---|---|---|---|
| | One year | | Two year | | One year | | Two year | |
| | MACE | Stroke/MI | MACE | Stroke/MI | MACE | Stroke/MI | MACE | Stroke/MI |
| **45 – 59** | 0.8 (0.43, 1.48) | 0.8 (0.38, 1.68) | 0.82 (0.5, 1.35) | 0.76 (0.46, 1.26) | 1.29 (0.46, 3.63) | 1.29 (0.35, 4.73) | 1.17 (0.51, 2.65) | 1.31 (0.47, 3.67) |
| **60 – 89** | 0.56 (0.25, 1.23) | 0.58 (0.22, 1.52) | 0.64 (0.34, 1.2) | 0.6 (0.32, 1.13) | 1.05 (0.34, 3.24) | 1.05 (0.24, 4.59) | 0.99 (0.42, 2.34) | 1.1 (0.38, 3.17) |
| **>=90** | 0.58 (0.2, 1.64) | 0.53 (0.19, 1.5) | 0.63 (0.29, 1.37) | 0.54 (0.28, 1.06) | 0.79 (0.21, 3.06) | 0.78 (0.15, 4.15) | 0.82 (0.28, 2.35) | 0.81 (0.2, 3.23) |
| **SBP**** | | | | | | | | |
| **<120** | ref | ref | ref | ref | ref | ref | ref | ref |
| **120 - 139** | 1.02 (0.81, 1.27) | 0.94 (0.74, 1.19) | 1.14 (0.97, 1.33) | 1.06 (0.88, 1.29) | 0.94 (0.7, 1.27) | 1.34 (0.94, 1.9) | 1.03 (0.82, 1.31) | 1.3 (0.99, 1.69) |
| **140 - 159** | 1.06 (0.82, 1.38) | 1.05 (0.82, 1.34) | 1.18 (0.99, 1.4) | 1.14 (0.93, 1.41) | 1.19 (0.87, 1.62) | 1.76 (1.21, 2.56) | 1.34 (1.05, 1.71) | 1.55 (1.17, 2.05) |
| **>=160** | 1.2 (0.88, 1.66) | 1.29 (0.95, 1.74) | 1.42 (1.16, 1.75) | 1.43 (1.15, 1.79) | 1.46 (1, 2.13) | 1.7 (1.05, 2.73) | 1.51 (1.11, 2.06) | 1.94 (1.37, 2.74) |
| **DBP**** | | | | x | x | x | x | x |
| **<80** | ref | ref | ref | x | x | x | x | x |
| **80 – 89** | 0.99 (0.84, 1.16) | 0.93 (0.77, 1.12) | 0.96 (0.86, 1.08) | x | x | x | x | x |
| **90 – 99** | 1.15 (0.89, 1.5) | 1.16 (0.89, 1.51) | 0.96 (0.78, 1.19) | x | x | x | x | x |
| **>=100** | 1.54 (1.04, 2.27) | 1 (0.6, 1.65) | 1.36 (1, 1.84) | x | x | x | x | x |
| **No. of concomitant medicines**** | | | | | x | | | |
| **0** | ref | ref | ref | ref | x | ref | ref | ref |
| **1 – 3** | 0.94 (0.74, 1.19) | 0.86 (0.64, 1.15) | 0.71 (0.6, 0.84) | 0.82 (0.66, 1.02) | x | 1.1 (0.61, 1.97) | 0.79 (0.56, 1.13) | 0.69 (0.45, 1.05) |
| **4 – 6** | 0.87 (0.67, 1.12) | 0.83 (0.61, 1.15) | 0.67 (0.56, 0.8) | 0.88 (0.69, 1.11) | x | 0.93 (0.5, 1.73) | 0.78 (0.54, 1.15) | 0.94 (0.61, 1.45) |
| **7 – 9** | 1.07 (0.81, 1.4) | 0.96 (0.68, 1.35) | 0.81 (0.67, 0.97) | 0.98 (0.76, 1.26) | x | 1.27 (0.68, 2.38) | 0.91 (0.61, 1.35) | 1.08 (0.69, 1.69) |
| **10 – 12** | 1.24 (0.93, 1.66) | 0.98 (0.68, 1.42) | 0.91 (0.74, 1.1) | 1.16 (0.89, 1.51) | x | 1.7 (0.9, 3.23) | 0.96 (0.64, 1.46) | 1.19 (0.75, 1.9) |
| **>=13** | 1.28 (0.95, 1.73) | 1.11 (0.76, 1.62) | 0.88 (0.72, 1.08) | 1.21 (0.92, 1.6) | x | 1.66 (0.87, 3.2) | 0.98 (0.64, 1.5) | 1.33 (0.83, 2.12) |
| **Cholesterol measurement** (HDL/LDL)** | | | | x | x | | x | x |
| **<=3.5** | ref | ref | ref | x | x | ref | x | x |

| OBP cohort<br>Predictor | Women<br>OR (95%CI) | | | | Men<br>OR (95%CI) | | | |
|---|---|---|---|---|---|---|---|---|
| | One year | | Two year | | One year | | Two year | |
| | MACE | Stroke/MI | MACE | Stroke/MI | MACE | Stroke/MI | MACE | Stroke/MI |
| **3.6 – 5** | 1.18 (0.95, 1.48) | 1.15 (0.91, 1.46) | 1.13 (0.95, 1.34) | x | x | 1.21 (0.82, 1.78) | x | x |
| **>5** | 1.44 (0.96, 2.18) | 1.28 (0.82, 2) | 1.39 (1.02, 1.9) | x | x | 1.56 (0.85, 2.86) | | x |
| **No. of previous fractures*** | x | | x | x | x | x | x | x |
| **0** | x | ref | x | x | x | x | x | x |
| **1** | x | 1.09 (0.91, 1.31) | x | x | x | x | x | x |
| **>=2** | x | 1.01 (0.83, 1.23) | x | x | x | x | x | x |

**Abbreviations**: OST, patients with incident diagnosis of osteoporosis; IFX, patients with incident fragility fracture; OBP, incident users of oral bisphosphonates; OR, odds ratio; CI, confidence intervals; MACE, composite outcome for the occurrence of either myocardial infarction, stroke or cardiovascular disease death; MI, myocardial infarction; * ever; ** in the year prior to start; SES, socio-economic status; BMI, body mass index; eGFR, estimated Glomerular Filtration Rate; SBP, cholesterol, systolic blood pressure; DBP, diastolic blood pressure.

#### Table S7a. Model equations from lasso selection for women and men based models (OST cohort)

| OST cohort<br>Predictor | Women<br>Beta coefficients | | | | Men<br>Beta coefficients | | | |
|---|---|---|---|---|---|---|---|---|
| | One year | | Two year | | One year | | Two year | |
| | MACE | Stroke/MI | MACE | Stroke/MI | MACE | Stroke/MI | MACE | Stroke/MI |
| **Intercept** | -4.047 | -4.538 | -4.560 | -3.974 | -4.267 | -4.134 | -4.577 | -3.165 |
| **SES** | | x | | x | x | x | x | x |
| **1** | ref | x | ref | x | x | x | x | x |
| **2** | -0.083 | x | 0.014 | x | x | x | x | x |
| **3** | 0.003 | x | 0.144 | x | x | x | x | x |
| **4** | 0.030 | x | -0.006 | x | x | x | x | x |
| **5** | -0.046 | x | 0.298 | x | x | x | x | x |
| **Smoking**** | | x | | x | x | x | x | x |
| **Ex** | ref | x | ref | x | x | x | x | x |
| **No** | 0.009 | x | 0.032 | x | x | x | x | x |
| **Yes** | 0.414 | x | 0.280 | x | x | x | x | x |
| **Drinking**** | | | | | x | x | x | x |
| **Ex** | ref | ref | ref | ref | x | x | x | x |
| **No** | 0.031 | 0.022 | -0.036 | -0.014 | x | x | x | x |

| OST cohort<br>Predictor | Women<br>Beta coefficients | | | | Men<br>Beta coefficients | | | |
|---|---|---|---|---|---|---|---|---|
| | One year | | Two year | | One year | | Two year | |
| | MACE | Stroke/MI | MACE | Stroke/MI | MACE | Stroke/MI | MACE | Stroke/MI |
| Yes | -0.226 | -0.283 | -0.186 | -0.168 | x | x | x | x |
| Diabetes type I* | x | x | x | x | x | x | x | x |
| Diabetes type II* | 0.343 | x | x | x | x | x | x | x |
| Chronic obstructive pulmonary disease* | x | x | x | x | x | x | x | x |
| Chronic kidney disease* | x | x | -0.124 | x | x | x | x | x |
| Rheumatoid arthritis* | x | x | x | x | x | x | x | x |
| Lupus* | x | x | x | x | x | x | x | x |
| Systemic heart disease** | x | x | x | x | x | x | x | x |
| Anti-osteoporosis use** | | x | 0.111 | x | x | x | x | x |
| Heparin use** | x | x | x | x | x | x | x | x |
| Beta-blocker use** | x | 0.391 | x | 0.210 | x | x | x | x |
| Hypertension** | x | x | x | x | x | x | x | x |
| Deep vein thrombosis or pulmonary embolism** | x | x | x | x | x | x | x | x |
| Anticoagulant use** | x | x | x | x | x | x | x | x |
| Antidepressants TCA** | x | x | x | x | x | x | x | x |
| Antidepressants SSRI** | 0.353 | x | 0.242 | | x | x | x | x |
| Hypercholesterolemia** | x | x | x | x | x | x | x | x |
| Statin use** | x | x | x | x | x | x | x | x |
| Osteoporosis history* | x | x | x | x | x | x | x | x |
| Family history of cardiovascular disease (%) | x | x | x | x | x | x | x | x |
| Family history of cardiovascular disease before age 60 | x | x | x | x | x | x | x | x |
| Heart failure* | x | x | x | x | x | x | x | x |
| Migraine* | x | x | x | | x | x | x | x |
| Severe mental illness* | x | x | x | x | x | x | x | x |
| Vascular Disease* | x | x | x | x | x | x | x | x |
| Atrial fibrillation* | 0.312 | x | 0.302 | x | x | x | x | x |
| On anti-hypertensive drug | 0.236 | 0.187 | 0.158 | 0.413 | x | x | | x |
| Antipsychotic use** | x | x | x | x | x | x | x | x |
| Steroid use** | 0.052 | x | 0.134 | x | x | x | x | x |
| Erectile dysfunction** | x | x | x | x | x | x | x | x |

| OST cohort Predictor | Women Beta coefficients | | | | Men Beta coefficients | | | |
|---|---|---|---|---|---|---|---|---|
| | One year | | Two year | | One year | | Two year | |
| | MACE | Stroke/MI | MACE | Stroke/MI | MACE | Stroke/MI | MACE | Stroke/MI |
| **Age Group** | | | | | | x | | x |
| **50-59** | ref | ref | ref | ref | ref | x | ref | x |
| **60-69** | 0.034 | 0.039 | 0.801 | 0.488 | 0.548 | x | 1.093 | x |
| **70-79** | 0.942 | 1.055 | 1.520 | 1.252 | 0.420 | x | 1.240 | x |
| **80-89** | 1.539 | 1.650 | 2.063 | 1.659 | 0.789 | x | 1.539 | x |
| **>89** | 1.762 | 1.906 | 2.437 | 2.008 | 0.749 | x | 1.633 | x |
| **Charlson score** | | x | | | x | x | x | x |
| **0** | ref | x | ref | ref | x | x | x | x |
| **1** | 0.244 | x | 0.224 | 0.235 | x | x | x | x |
| **2** | -0.071 | x | 0.027 | 0.001 | x | x | x | x |
| **≥3** | 0.262 | x | 0.197 | 0.187 | x | x | x | x |
| **Cardiovascular disease** | x | x | x | x | x | x | x | x |
| **No** | x | x | x | x | x | x | x | x |
| **Ever >1 year before index date** | x | x | x | x | x | x | x | x |
| **1 year before index** | x | x | x | x | x | x | x | x |
| **6 months before index** | x | x | x | x | x | x | x | x |
| **1 month before index** | x | x | x | x | x | x | x | x |
| **MI or Stroke** | | x | | x | x | x | x | x |
| **No** | ref | x | ref | x | x | x | x | x |
| **Ever >1 year before index date** | 0.284 | x | 0.263 | x | x | x | x | x |
| **1 year before index** | 0.895 | x | 0.522 | x | x | x | x | x |
| **Established CVD *** | 0.427 | 0.791 | 0.472 | 0.699 | 0.839 | 0.783 | 0.808 | 0.828 |
| **Any fracture history** | | x | x | x | x | x | x | x |
| **No** | ref | x | x | x | x | x | x | x |
| **Ever >1 year before index date** | -0.000 | x | x | x | x | x | x | x |
| **1 year before index** | 0.149 | x | x | x | x | x | x | x |
| **Hip fracture history** | x | x | x | x | x | x | x | x |
| **No** | x | x | x | x | x | x | x | x |
| **Ever >1 year before index date** | x | x | x | x | x | x | x | x |
| **1 year before index** | x | x | x | x | x | x | x | x |

| OST cohort<br>Predictor | Women<br>Beta coefficients | | | | Men<br>Beta coefficients | | | |
|---|---|---|---|---|---|---|---|---|
| | One year | | Two year | | One year | | Two year | |
| | MACE | Stroke/MI | MACE | Stroke/MI | MACE | Stroke/MI | MACE | Stroke/MI |
| **Shoulder fracture history** | x | x | x | x | x | x | x | x |
| **No** | x | x | x | x | x | x | x | x |
| **Ever >1 year before index date** | x | x | x | x | x | x | x | x |
| **1 year before index** | x | x | x | x | x | x | x | x |
| **Spine fracture history** | x | x | x | x | x | x | x | x |
| **No** | x | x | x | x | x | x | x | x |
| **Ever >1 year before index date** | x | x | x | x | x | x | x | x |
| **1 year before index** | x | x | x | x | x | x | x | x |
| **Wrist fracture history** | x | x | x | x | x | x | x | x |
| **No** | x | x | x | x | x | x | x | x |
| **Ever >1 year before index date** | x | x | x | x | x | x | x | x |
| **1 year before index** | x | x | x | x | x | x | x | x |
| **BMI**** | | x | | | x | x | x | x |
| **<18.5** | ref | x | ref | ref | x | x | x | x |
| **18.6 - 24.9** | -0.438 | x | -0.347 | -0.186 | x | x | x | x |
| **25 - 29.9** | -0.646 | x | -0.507 | -0.344 | x | x | x | x |
| **30 - 39.9** | -0.879 | x | -0.666 | -0.352 | x | x | x | x |
| **>=40** | -0.647 | x | -0.107 | -0.046 | x | x | x | x |
| **No. of GP visits**** | | x | x | | x | x | x | x |
| **0** | ref | x | x | ref | x | x | x | x |
| **1-5** | -0.312 | x | x | -0.416 | x | x | x | x |
| **6-10** | -0.559 | x | x | -0.469 | x | x | x | x |
| **11-15** | -0.517 | x | x | -0.432 | x | x | x | x |
| **>=16** | -0.447 | x | x | -0.315 | x | x | x | x |
| **No. of GP emergency visits**** | | | | x | x | x | x | x |
| **0** | ref | ref | ref | x | x | x | x | x |
| **1** | 0.463 | -0.133 | 0.010 | x | x | x | x | x |
| **2** | 0.511 | -0.271 | 0.136 | x | x | x | x | x |
| **3-5** | 0.378 | -0.253 | 0.314 | x | x | x | x | x |
| **>=6** | 0.739 | -0.103 | 0.646 | x | x | x | x | x |
| **eGFR**** | | | | | x | x | | |

| OST cohort<br>Predictor | Women<br>Beta coefficients | | | | Men<br>Beta coefficients | | | |
|---|---|---|---|---|---|---|---|---|
| | One year | | Two year | | One year | | Two year | |
| | MACE | Stroke/MI | MACE | Stroke/MI | MACE | Stroke/MI | MACE | Stroke/MI |
| **<=29** | ref | ref | ref | ref | x | x | ref | ref |
| **30 – 44** | -0.508 | -0.338 | -0.022 | -0.335 | x | x | 0.673 | 0.385 |
| **45 – 59** | -0.573 | -0.533 | -0.257 | -0.490 | x | x | 0.368 | -0.042 |
| **60 – 89** | -0.820 | -0.770 | -0.473 | -0.723 | x | x | 0.159 | -0.307 |
| **>=90** | -0.763 | -0.591 | -0.400 | -0.663 | x | x | 0.171 | -0.531 |
| **SBP**** | | x | | x | x | x | x | x |
| **<120** | ref | x | ref | x | x | x | x | x |
| **120 - 139** | 0.085 | x | -0.046 | x | x | x | x | x |
| **140 - 159** | 0.189 | x | 0.193 | x | x | x | x | x |
| **>=160** | 0.262 | x | 0.279 | x | x | x | x | x |
| **DBP**** | x | x | | x | x | x | x | x |
| **<80** | x | x | ref | x | x | x | x | x |
| **80 – 89** | x | x | 0.073 | x | x | x | x | x |
| **90 – 99** | x | x | 0.021 | x | x | x | x | x |
| **>=100** | x | x | 0.229 | x | x | x | x | x |
| **No. of concomitant medicines**** | | | | | x | | x | x |
| **0** | ref | ref | ref | ref | x | ref | x | x |
| **1 – 3** | -0.608 | -0.329 | -0.441 | -0.177 | x | -1.044 | x | x |
| **4 – 6** | -0.328 | -0.387 | -0.409 | -0.173 | x | -0.111 | x | x |
| **7 – 9** | -0.209 | -0.215 | -0.308 | -0.099 | x | 0.506 | x | x |
| **10 – 12** | -0.052 | -0.193 | -0.153 | 0.048 | x | -0.052 | x | x |
| **>=13** | -0.055 | 0.025 | -0.204 | 0.096 | x | 0.642 | x | x |
| **Cholesterol measurement** (HDL/LDL)** | | x | | x | x | x | x | x |
| **<=3.5** | ref | x | ref | x | x | x | x | x |
| **3.6 – 5** | 0.315 | x | 0.193 | x | x | x | x | x |
| **>5** | 0.586 | x | 0.339 | x | x | x | x | x |
| **No. of previous fractures*** | | | x | x | x | x | x | x |
| **0** | ref | ref | x | x | x | x | x | x |
| **1** | 0.138 | 0.117 | x | x | x | x | x | x |
| **>=2** | -0.118 | -0.087 | x | x | x | x | x | x |

**Abbreviations**: OST, patients with incident diagnosis of osteoporosis; IFX, patients with incident fragility fracture; OBP, incident users of oral bisphosphonates; MACE, composite outcome for the occurrence of either myocardial infarction, stroke or cardiovascular disease death; MI, myocardial infarction; * ever; ** in the year prior to start; SES, socio-economic status; BMI, body mass index; eGFR, estimated Glomerular Filtration Rate; SBP, cholesterol, systolic blood pressure; DBP, diastolic blood pressure.

#### Table S7b. Model equations from lasso selection for women and men based models (IFX cohort)

| IFX cohort Predictor | Women Beta coefficients | | | | Men Beta coefficients | | | |
|---|---|---|---|---|---|---|---|---|
| | One year | | Two year | | One year | | Two year | |
| | MACE | Stroke/MI | MACE | Stroke/MI | MACE | Stroke/MI | MACE | Stroke/MI |
| **Intercept** | -6.015 | -6.485 | -4.958 | -5.035 | -4.720 | -5.321 | -3.951 | -4.628 |
| **SES** | x | x | x | x | x | x | x | x |
| **1** | x | x | x | x | x | x | x | x |
| **2** | x | x | x | x | x | x | x | x |
| **3** | x | x | x | x | x | x | x | x |
| **4** | x | x | x | x | x | x | x | x |
| **5** | x | x | x | x | x | x | x | x |
| **Smoking**** | x | x | x | x | | x | x | x |
| **Ex** | x | x | x | x | ref | x | x | x |
| **No** | x | x | x | x | 0.079 | x | x | x |
| **Yes** | x | x | x | x | 0.016 | x | x | x |
| **Drinking**** | | x | | | | x | | x |
| **Ex** | ref | x | ref | ref | ref | x | ref | x |
| **No** | -0.072 | x | -0.128 | -0.115 | 0.138 | x | 0.140 | x |
| **Yes** | -0.182 | x | -0.285 | -0.224 | 0.012 | x | -0.113 | x |
| **Diabetes type I*** | x | x | x | x | x | x | x | x |
| **Diabetes type II*** | x | x | x | 0.312 | x | x | x | x |
| **Chronic obstructive pulmonary disease*** | x | x | x | x | x | x | x | x |
| **Chronic kidney disease*** | x | x | x | x | x | x | x | x |
| **Rheumatoid arthritis*** | x | x | x | x | x | x | x | x |
| **Lupus*** | x | x | x | x | x | x | x | x |
| **Systemic heart disease**** | x | x | x | x | x | x | x | x |
| **Anti-osteoporosis use**** | x | x | x | x | x | x | x | x |
| **Heparin use**** | x | x | x | x | x | x | x | x |
| **Beta-blocker use**** | 0.025 | 0.028 | x | 0.105 | x | x | x | x |
| **Hypertension**** | x | x | x | x | x | x | x | x |
| **Deep vein thrombosis or pulmonary embolism**** | x | x | x | x | x | x | x | x |
| **Anticoagulant use**** | x | x | x | x | x | x | x | x |
| **Antidepressants TCA**** | x | x | x | x | x | x | x | x |
| **Antidepressants SSRI**** | x | x | x | -0.312 | x | x | x | x |

| IFX cohort Predictor | Women Beta coefficients | | | | Men Beta coefficients | | | |
|---|---|---|---|---|---|---|---|---|
| | One year | | Two year | | One year | | Two year | |
| | MACE | Stroke/MI | MACE | Stroke/MI | MACE | Stroke/MI | MACE | Stroke/MI |
| **Hypercholesterolemia**** | x | x | x | x | x | x | x | x |
| **Statin use**** | x | x | x | x | x | x | x | x |
| **Osteoporosis history*** | x | x | x | x | x | x | x | x |
| **Family history of cardiovascular disease (%)** | x | x | x | x | x | x | x | x |
| **Family history of cardiovascular disease before age 60** | x | x | x | x | x | x | x | x |
| **Heart failure*** | 0.162 | x | x | x | x | x | x | x |
| **Migraine*** | x | x | x | x | x | x | x | x |
| **Severe mental illness*** | x | x | x | x | x | x | x | x |
| **Vascular Disease*** | X | x | x | x | x | x | x | x |
| **Atrial fibrillation*** | 0.195 | 0.391 | x | 0.243 | x | x | x | x |
| **On anti-hypertensive drug** | 0.060 | 0.027 | 0.086 | 0.178 | 0.269 | 0.377 | 0.403 | 0.413 |
| **Antipsychotic use**** | x | x | x | x | x | x | x | x |
| **Steroid use**** | x | x | x | x | x | x | x | x |
| **Erectile dysfunction**** | x | x | x | x | x | x | x | x |
| **Age Group (%)** | | | | | | | | |
| **50-59** | ref | ref | ref | ref | ref | ref | ref | ref |
| **60-69** | 1.761 | 1.839 | 0.766 | 0.841 | 1.013 | 0.969 | 0.593 | 1.020 |
| **70-79** | 2.393 | 2.376 | 1.668 | 1.620 | 1.668 | 1.268 | 0.892 | 1.157 |
| **80-89** | 2.784 | 2.709 | 2.149 | 1.980 | 1.911 | 1.404 | 1.285 | 1.287 |
| **>89** | 3.177 | 2.748 | 2.289 | 1.911 | 1.989 | 1.259 | 1.270 | 1.062 |
| **Charlson score** | x | x | x | x | x | x | x | x |
| **0** | x | x | x | x | x | x | x | x |
| **1** | x | x | x | x | x | x | x | x |
| **2** | x | x | x | x | x | x | x | x |
| **≥3** | x | x | x | x | x | x | x | x |
| **Cardiovascular disease** | x | x | x | | x | x | x | x |
| **No** | x | x | x | ref | x | x | x | x |
| **Ever >1 year before index date** | x | x | x | 0.122 | x | x | x | x |
| **1 year before index** | x | x | x | 0.179 | x | x | x | x |
| **6 months before index** | x | x | x | 0.183 | x | x | x | x |
| **1 month before index** | x | x | x | 0.893 | x | x | x | x |

| IFX cohort<br>Predictor | Women<br>Beta coefficients | | | | Men<br>Beta coefficients | | | |
|---|---|---|---|---|---|---|---|---|
| | One year | | Two year | | One year | | Two year | |
| | MACE | Stroke/MI | MACE | Stroke/MI | MACE | Stroke/MI | MACE | Stroke/MI |
| **MI or Stroke** | | x | x | x | x | x | x | x |
| **No** | ref | x | x | x | x | x | x | x |
| **Ever >1 year before index date** | 0.125 | x | x | x | x | x | x | x |
| **1 year before index** | 0.384 | x | x | x | x | x | x | x |
| **Established CVD *** | 0.423 | 0.513 | 0.656 | 0.483 | 0.641 | 0.658 | 0.619 | 0.569 |
| **Any fracture history** | x | x | x | x | x | x | x | x |
| **No** | x | x | x | x | x | x | x | x |
| **Ever >1 year before index date** | x | x | x | x | x | x | x | x |
| **1 year before index** | x | x | x | x | x | x | x | x |
| **Hip fracture history** | x | x | x | x | x | x | x | x |
| **No** | x | x | x | x | x | x | x | x |
| **Ever >1 year before index date** | x | x | x | x | x | x | x | x |
| **1 year before index** | x | x | x | x | x | x | x | x |
| **Shoulder fracture history** | x | x | x | x | x | x | x | x |
| **No** | x | x | x | x | x | x | x | x |
| **Ever >1 year before index date** | x | x | x | x | x | x | x | x |
| **1 year before index** | x | x | x | x | x | x | x | x |
| **Spine fracture history** | x | x | x | x | x | x | x | x |
| **No** | x | x | x | x | x | x | x | x |
| **Ever >1 year before index date** | x | x | x | x | x | x | x | x |
| **1 year before index** | x | x | x | x | x | x | x | x |
| **Wrist fracture history** | x | x | x | x | x | x | x | x |
| **No** | x | x | x | x | x | x | x | x |
| **Ever >1 year before index date** | x | x | x | x | x | x | x | x |
| **1 year before index** | x | x | x | x | x | x | x | x |
| **BMI**** | x | x | x | | x | x | x | x |
| **<18.5** | x | x | x | ref | x | x | x | x |
| **18.6 - 24.9** | x | x | x | 0.028 | x | x | x | x |
| **25 - 29.9** | x | x | x | 0.011 | x | x | x | x |

| IFX cohort<br>Predictor | Women<br>Beta coefficients | | | | Men<br>Beta coefficients | | | |
|---|---|---|---|---|---|---|---|---|
| | One year | | Two year | | One year | | Two year | |
| | MACE | Stroke/MI | MACE | Stroke/MI | MACE | Stroke/MI | MACE | Stroke/MI |
| **30 - 39.9** | x | x | x | 0.160 | x | x | x | x |
| **>=40** | x | x | x | 0.357 | x | x | x | x |
| **No. of GP visits**** | | x | x | x | x | x | x | x |
| **0** | ref | x | x | x | x | x | x | x |
| **1-5** | -0.158 | x | x | x | x | x | x | x |
| **6-10** | -0.278 | x | x | x | x | x | x | x |
| **11-15** | -0.264 | x | x | x | x | x | x | x |
| **>=16** | -0.159 | x | x | x | x | x | x | x |
| **No. of GP emergency visits**** | x | x | x | x | x | x | x | x |
| **0** | x | x | x | x | x | x | x | x |
| **1** | x | x | x | x | x | x | x | x |
| **2** | x | x | x | x | x | x | x | x |
| **3-5** | x | x | x | x | x | x | x | x |
| **>=6** | x | x | x | x | x | x | x | x |
| **eGFR**** | | | | | | x | | x |
| **<=29** | ref | ref | ref | ref | ref | x | ref | x |
| **30 – 44** | -0.128 | -0.117 | 0.092 | -0.003 | -0.095 | x | | x |
| **45 – 59** | -0.221 | -0.254 | -0.080 | -0.086 | -0.296 | x | | x |
| **60 – 89** | -0.329 | -0.372 | -0.091 | -0.092 | -0.374 | x | | x |
| **>=90** | -0.359 | -0.197 | 0.028 | 0.003 | -0.721 | x | | x |
| **SBP**** | | | | | x | | | |
| **<120** | ref | ref | ref | ref | x | ref | ref | ref |
| **120 - 139** | 0.154 | 0.242 | 0.032 | 0.008 | x | -0.234 | | |
| **140 - 159** | 0.315 | 0.335 | 0.075 | 0.125 | x | 0.348 | | |
| **>=160** | 0.449 | 0.536 | 0.317 | 0.421 | x | 0.600 | | |
| **DBP**** | x | x | x | x | x | x | | x |
| **<80** | x | x | x | x | x | x | ref | x |
| **80 – 89** | x | x | x | x | x | x | | x |
| **90 – 99** | x | x | x | x | x | x | | x |
| **>=100** | x | x | x | x | x | x | | x |
| **No. of concomitant medicines**** | | | | | x | x | x | x |
| **0** | ref | ref | ref | ref | x | x | x | x |
| **1 – 3** | 0.157 | -0.110 | 0.040 | -0.218 | x | x | x | x |
| **4 – 6** | 0.347 | 0.175 | 0.254 | -0.153 | x | x | x | x |

| IFX cohort Predictor | Women Beta coefficients | | | | Men Beta coefficients | | | |
|---|---|---|---|---|---|---|---|---|
| | One year | | Two year | | One year | | Two year | |
| | MACE | Stroke/MI | MACE | Stroke/MI | MACE | Stroke/MI | MACE | Stroke/MI |
| **7 – 9** | 0.415 | 0.136 | 0.283 | 0.018 | x | x | x | x |
| **10 – 12** | 0.442 | 0.311 | 0.327 | 0.056 | x | x | x | x |
| **>=13** | 0.559 | 0.398 | 0.327 | 0.002 | x | x | x | x |
| **Cholesterol measurement** (HDL/LDL)** | x | x | | x | x | x | x | x |
| **<=3.5** | x | x | ref | x | x | x | x | x |
| **3.6 – 5** | x | x | 0.211 | x | x | x | x | x |
| **>5** | x | x | 0.327 | x | x | x | x | x |
| **No. of previous fractures*** | x | x | x | x | x | x | x | x |
| **0** | x | x | x | x | x | x | x | x |
| **1** | x | x | x | x | x | x | x | x |
| **>=2** | x | x | x | x | x | x | x | x |

**Abbreviations**: OST, patients with incident diagnosis of osteoporosis; IFX, patients with incident fragility fracture; OBP, incident users of oral bisphosphonates; MACE, composite outcome for the occurrence of either myocardial infarction, stroke or cardiovascular disease death; MI, myocardial infarction; * ever; ** in the year prior to start; SES, socio-economic status; BMI, body mass index; eGFR, estimated Glomerular Filtration Rate; SBP, cholesterol, systolic blood pressure; DBP, diastolic blood pressure.

#### Table S7c. Model equations from lasso selection for women and men based models (OBP cohort)

| OBP cohort Predictor | Women Beta coefficients | | | | Men Beta coefficients | | | |
|---|---|---|---|---|---|---|---|---|
| | One year | | Two year | | One year | | Two year | |
| | MACE | Stroke/MI | MACE | Stroke/MI | MACE | Stroke/MI | MACE | Stroke/MI |
| **Intercept** | -4.769 | -4.669 | -3.946 | -4.056 | -4.521 | -4.673 | -3.412 | -4.022 |
| **SES** | | | | x | x | x | | x |
| **1** | ref | ref | ref | x | x | x | ref | x |
| **2** | -0.006 | 0.131 | 0.070 | x | x | x | 0.106 | x |
| **3** | 0.164 | 0.291 | 0.223 | x | x | x | 0.087 | x |
| **4** | 0.129 | 0.230 | 0.177 | x | x | x | 0.228 | x |
| **5** | 0.240 | 0.216 | 0.181 | x | x | x | 0.099 | x |
| **Smoking**** | x | | | | x | | x | x |
| **Ex** | x | ref | ref | ref | x | ref | x | x |
| **No** | x | -0.184 | -0.128 | -0.099 | x | -0.025 | x | x |
| **Yes** | x | -0.090 | 0.085 | 0.206 | x | 0.154 | x | x |
| **Drinking**** | | | | | x | x | | x |
| **Ex** | ref | ref | ref | ref | x | x | ref | x |

| OBP cohort Predictor | Women Beta coefficients | | | | Men Beta coefficients | | | |
|---|---|---|---|---|---|---|---|---|
| | One year | | Two year | | One year | | Two year | |
| | MACE | Stroke/MI | MACE | Stroke/MI | MACE | Stroke/MI | MACE | Stroke/MI |
| No | -0.046 | 0.049 | -0.126 | -0.107 | x | x | -0.172 | x |
| Yes | -0.194 | -0.139 | -0.253 | -0.249 | x | x | -0.206 | x |
| Diabetes type I* | x | x | x | x | x | x | x | x |
| Diabetes type II* | 0.374 | 0.401 | 0.308 | x | x | x | x | x |
| Chronic obstructive pulmonary disease* | x | 0.234 | x | x | x | x | x | x |
| Chronic kidney disease* | -0.208 | x | -0.148 | x | x | x | x | x |
| Rheumatoid arthritis* | x | x | -0.004 | x | -0.569 | -0.317 | x | x |
| Lupus* | x | x | x | x | x | x | x | x |
| Systemic heart disease** | x | x | x | x | x | x | x | x |
| Anti-osteoporosis use** | x | x | x | x | x | x | x | x |
| Heparin use** | x | x | x | x | x | x | x | x |
| Beta-blocker use** | 0.073 | 0.225 | 0.175 | 0.154 | x | 0.044 | x | x |
| Hypertension** | x | x | x | x | x | x | x | x |
| Deep vein thrombosis or pulmonary embolism** | x | x | x | x | x | x | x | x |
| Anticoagulant use** | x | x | x | x | x | x | x | x |
| Antidepressants TCA** | x | x | x | x | x | x | x | x |
| Antidepressants SSRI** | x | 0.252 | 0.152 | x | x | x | x | x |
| Hypercholesterolemia** | x | x | x | x | x | x | x | x |
| Statin use** | x | -0.179 | x | x | x | x | x | x |
| Osteoporosis history* | -0.280 | -0.261 | -0.263 | -0.164 | -0.180 | x | -0.185 | -0.201 |
| Family history of cardiovascular disease | x | x | -0.013 | x | x | x | x | x |
| Family history of cardiovascular disease before age 60 | x | x | x | x | x | x | x | x |
| Heart failure* | x | x | x | x | x | 0.361 | x | x |
| Migraine* | x | x | x | x | x | x | | x |
| Severe mental illness* | x | x | x | x | x | x | x | x |
| Vascular Disease* | x | x | x | x | x | x | x | x |
| Atrial fibrillation* | 0.448 | 0.369 | 0.418 | 0.336 | x | x | x | x |
| On anti-hypertensive drug | x | 0.130 | 0.089 | 0.160 | x | x | 0.131 | 0.101 |
| Antipsychotic use** | x | x | x | x | x | x | x | x |
| Steroid use** | x | 0.052 | x | x | x | -0.013 | x | x |

| OBP cohort<br>Predictor | Women<br>Beta coefficients | | | | Men<br>Beta coefficients | | | |
|---|---|---|---|---|---|---|---|---|
| | One year | | Two year | | One year | | Two year | |
| | MACE | Stroke/MI | MACE | Stroke/MI | MACE | Stroke/MI | MACE | Stroke/MI |
| **Erectile dysfunction**** | x | x | x | x | x | x | -0.112 | x |
| **Age Group** | | | | | | | | |
| **50-59** | ref | ref | ref | ref | ref | ref | ref | ref |
| **60-69** | 0.836 | 0.621 | 0.481 | 0.587 | 0.455 | 0.231 | 0.328 | 0.182 |
| **70-79** | 1.465 | 0.987 | 1.195 | 1.225 | 0.872 | 0.395 | 0.670 | 0.675 |
| **80-89** | 2.152 | 1.592 | 1.845 | 1.751 | 1.030 | 0.592 | 0.930 | 0.700 |
| **>89** | 2.299 | 1.653 | 2.105 | 1.849 | 1.689 | 0.818 | 1.193 | 0.855 |
| **Charlson score** | | x | | x | x | | | x |
| **0** | ref | x | ref | x | x | ref | ref | x |
| **1** | -0.159 | x | -0.066 | x | x | 0.018 | -0.121 | x |
| **2** | -0.108 | x | -0.032 | x | x | 0.085 | -0.246 | x |
| **≥3** | -0.020 | x | -0.032 | x | x | 0.078 | -0.070 | x |
| **Cardiovascular disease** | | | x | | x | | x | |
| **No** | ref | ref | x | ref | x | ref | x | ref |
| **Ever >1 year before index date** | 0.136 | 0.267 | x | 0.322 | x | 0.351 | x | 0.229 |
| **1 year before index** | 0.106 | -0.019 | x | 0.083 | x | 0.605 | x | 0.159 |
| **6 months before index** | 0.218 | 0.403 | x | 0.409 | x | 1.109 | x | 0.988 |
| **1 month before index** | 0.764 | 1.042 | x | 0.759 | x | 1.298 | x | 0.920 |
| **MI or Stroke** | | | | | x | | | x |
| **No** | ref | ref | ref | ref | x | ref | ref | x |
| **Ever >1 year before index date** | 0.267 | 0.030 | 0.334 | 0.244 | x | -0.268 | 0.299 | x |
| **1 year before index** | 0.739 | 0.765 | 0.684 | 0.494 | x | 0.634 | 0.846 | x |
| **Established CVD *** | 0.459 | 0.344 | 0.493 | 0.317 | 0.834 | 0.319 | 0.535 | 0.399 |
| **Any fracture history** | x | x | x | x | x | x | x | x |
| **No** | x | x | x | x | x | x | x | x |
| **Ever >1 year before index date** | x | x | x | x | x | x | x | x |
| **1 year before index** | x | x | x | x | x | x | x | x |
| **Hip fracture history** | x | x | x | x | x | x | | x |
| **No** | x | x | x | x | x | x | ref | x |
| **Ever >1 year before index date** | x | x | x | x | x | x | -0.107 | x |

| OBP cohort<br>Predictor | Women<br>Beta coefficients | | | | Men<br>Beta coefficients | | | |
|---|---|---|---|---|---|---|---|---|
| | One year | | Two year | | One year | | Two year | |
| | MACE | Stroke/MI | MACE | Stroke/MI | MACE | Stroke/MI | MACE | Stroke/MI |
| 1 year before index | x | x | x | x | x | x | 0.166 | x |
| **Shoulder fracture history** | x | x | x | x | x | x | x | x |
| No | x | x | x | x | x | x | x | x |
| Ever >1 year before index date | x | x | x | x | x | x | x | x |
| 1 year before index | x | x | x | x | x | x | x | x |
| **Spine fracture history** | x | x | x | x | x | x | x | x |
| No | x | x | x | x | x | x | x | x |
| Ever >1 year before index date | x | x | x | x | x | x | x | x |
| 1 year before index | x | x | x | x | x | x | x | x |
| **Wrist fracture history** | x | x | x | x | x | x | x | x |
| No | x | x | x | x | x | x | x | x |
| Ever >1 year before index date | x | x | x | x | x | x | x | x |
| 1 year before index | x | x | x | x | x | x | x | x |
| **BMI**** | | | | | x | | | |
| <18.5 | ref | ref | ref | ref | x | ref | ref | ref |
| 18.6 - 24.9 | -0.328 | -0.249 | -0.290 | -0.234 | x | 0.100 | -0.185 | -0.087 |
| 25 - 29.9 | -0.553 | -0.389 | -0.549 | -0.404 | x | -0.136 | -0.330 | -0.198 |
| 30 - 39.9 | -0.609 | -0.529 | -0.540 | -0.389 | x | -0.252 | -0.558 | -0.272 |
| >=40 | -0.867 | -0.525 | -0.493 | -0.313 | x | -0.112 | -0.206 | -0.187 |
| **No. of GP visits**** | | | x | | x | | | |
| 0 | ref | ref | x | ref | x | ref | ref | ref |
| 1-5 | -0.046 | -0.159 | x | -0.093 | x | -0.658 | 0.061 | -0.029 |
| 6-10 | -0.175 | -0.137 | x | -0.180 | x | -0.575 | -0.126 | -0.376 |
| 11-15 | -0.364 | -0.233 | x | -0.266 | x | -0.594 | -0.118 | -0.287 |
| >=16 | -0.233 | -0.150 | x | -0.132 | x | -0.377 | 0.130 | -0.141 |
| **No. of GP emergency visits**** | | | | x | x | x | x | x |
| 0 | ref | ref | ref | x | x | x | x | x |
| 1 | 0.086 | 0.085 | 0.157 | x | x | x | x | x |
| 2 | 0.281 | 0.310 | 0.288 | x | x | x | x | x |
| 3-5 | 0.332 | 0.087 | 0.177 | x | x | x | x | x |
| >=6 | 0.881 | 0.533 | 0.492 | x | x | x | x | x |

| OBP cohort Predictor | Women Beta coefficients | | | | Men Beta coefficients | | | |
|---|---|---|---|---|---|---|---|---|
| | One year | | Two year | | One year | | Two year | |
| | MACE | Stroke/MI | MACE | Stroke/MI | MACE | Stroke/MI | MACE | Stroke/MI |
| **eGFR**** | | | | | | | | |
| **<=29** | ref | ref | ref | ref | ref | ref | ref | ref |
| **30 – 44** | -0.118 | -0.060 | -0.069 | -0.132 | 0.355 | 0.190 | 0.320 | 0.451 |
| **45 – 59** | -0.221 | -0.224 | -0.195 | -0.276 | 0.253 | 0.255 | 0.154 | 0.270 |
| **60 – 89** | -0.585 | -0.550 | -0.445 | -0.508 | 0.049 | 0.050 | -0.008 | 0.092 |
| **>=90** | -0.553 | -0.630 | -0.455 | -0.609 | -0.233 | -0.247 | -0.202 | -0.212 |
| **SBP**** | | | | | | | | |
| **<120** | ref | ref | ref | ref | ref | ref | ref | ref |
| **120 - 139** | 0.015 | -0.063 | 0.129 | 0.061 | -0.058 | 0.290 | 0.033 | 0.260 |
| **140 - 159** | 0.062 | 0.047 | 0.166 | 0.134 | 0.174 | 0.566 | 0.291 | 0.440 |
| **>=160** | 0.186 | 0.253 | 0.353 | 0.360 | 0.379 | 0.528 | 0.413 | 0.663 |
| **DBP**** | | | | x | x | x | x | x |
| **<80** | ref | ref | ref | x | x | x | x | x |
| **80 – 89** | -0.014 | -0.072 | -0.041 | x | x | x | x | x |
| **90 – 99** | 0.144 | 0.148 | -0.040 | x | x | x | x | x |
| **>=100** | 0.431 | -0.003 | 0.304 | x | x | x | x | x |
| **No. of concomitant medicines**** | | | | | x | | | |
| **0** | ref | ref | ref | ref | x | ref | ref | ref |
| **1 – 3** | -0.063 | -0.152 | -0.339 | -0.197 | x | 0.091 | -0.231 | -0.372 |
| **4 – 6** | -0.141 | -0.182 | -0.400 | -0.131 | x | -0.075 | -0.243 | -0.061 |
| **7 – 9** | 0.063 | -0.041 | -0.214 | -0.021 | x | 0.242 | -0.095 | 0.077 |
| **10 – 12** | 0.217 | -0.016 | -0.098 | 0.147 | x | 0.532 | -0.037 | 0.177 |
| **>=13** | 0.247 | 0.106 | -0.126 | 0.193 | x | 0.510 | -0.018 | 0.285 |
| **Cholesterol measurement** (HDL/LDL)** | | | | x | x | | x | x |
| **<=3.5** | ref | ref | ref | x | x | ref | x | x |
| **3.6 – 5** | 0.169 | 0.141 | 0.119 | x | x | 0.190 | x | x |
| **>5** | 0.368 | 0.247 | 0.331 | x | x | 0.442 | x | x |
| **No. of previous fractures*** | x | | x | x | x | x | x | x |
| **0** | x | ref | x | x | x | x | x | x |
| **1** | x | 0.089 | x | x | x | x | x | x |
| **>=2** | x | 0.010 | x | x | x | x | x | x |

**Abbreviations**: OST, patients with incident diagnosis of osteoporosis; IFX, patients with incident fragility fracture; OBP, incident users of oral bisphosphonates; MACE, composite outcome for the occurrence of either myocardial infarction, stroke or cardiovascular disease death; MI, myocardial infarction; * ever; ** in the

| OBP cohort<br>Predictor | Women<br>Beta coefficients | | | | Men<br>Beta coefficients | | | |
|---|---|---|---|---|---|---|---|---|
| | One year | | Two year | | One year | | Two year | |
| | MACE | Stroke/MI | MACE | Stroke/MI | MACE | Stroke/MI | MACE | Stroke/MI |
| year prior to start; SES, socio-economic status; BMI, body mass index; eGFR, estimated Glomerular Filtration Rate; SBP, cholesterol, systolic blood pressure; DBP, diastolic blood pressure. | | | | | | | | |

*Supplementary figures*

#### Figure S1. Incidence rates by age groups

##### a. After one year of follow up (MACE)

| Age group | Cohort | Number of events (%) | Incidence Rate (cases/1000py) |
|---|---|---|---|
| 50-59 | OST | 4.73 (0.06%) | 4.73 |
| | IFX | 4.43 (0.03%) | 4.43 |
| | OBP | 5.58 (0.06%) | 5.58 |
| 60-69 | OST | 7.17 (0.17%) | 7.17 |
| | IFX | 16.98 (0.20%) | 16.98 |
| | OBP | 10.02 (0.21%) | 10.02 |
| 70-79 | OST | 16.95 (0.53%) | 16.95 |
| | IFX | 36.68 (0.76%) | 36.68 |
| | OBP | 20.67 (0.62%) | 20.67 |
| 80-89 | OST | 36.08 (0.86%) | 36.08 |
| | IFX | 65.64 (2.13%) | 65.64 |
| | OBP | 44.30 (1.17%) | 44.3 |
| 90+ | OST | 60.92 (0.27%) | 60.92 |
| | IFX | 101.22 (1.17%) | 101.22 |
| | OBP | 71.30 (0.42%) | 71.3 |

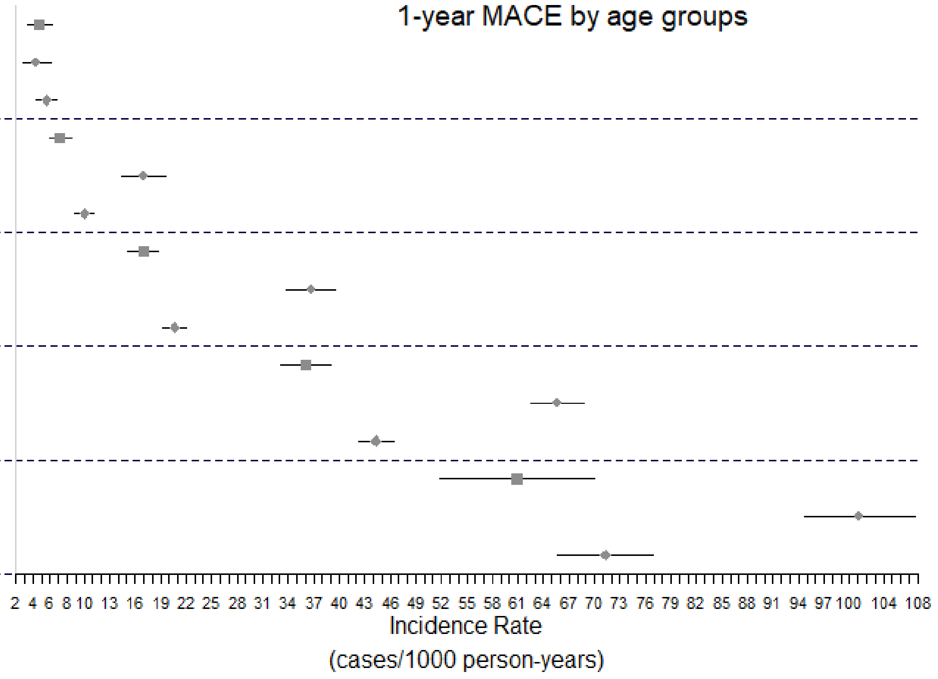

#### b. After two years of follow up (MACE)

| Age group | Cohort | Number of events (%) | Incidence Rate (cases/1000py) |
|---|---|---|---|
| | OST | 3.91 (0.10%) | 3.91 |
| 50-59 | IFX | 5.42 (0.08%) | 5.42 |
| | OBP | 4.91 (0.11%) | 4.91 |
| | OST | 7.27 (0.35%) | 7.27 |
| 60-69 | IFX | 14.28 (0.32%) | 14.28 |
| | OBP | 9.44 (0.39%) | 9.44 |
| | OST | 17.06 (1.05%) | 17.06 |
| 70-79 | IFX | 30.80 (1.21%) | 30.8 |
| | OBP | 20.10 (1.16%) | 20.1 |
| | OST | 34.71 (1.57%) | 34.71 |
| 80-89 | IFX | 55.70 (3.29%) | 55.7 |
| | OBP | 42.16 (2.07%) | 42.16 |
| | OST | 60.88 (0.48%) | 60.88 |
| 90+ | IFX | 79.54 (1.58%) | 79.54 |
| | OBP | 68.03 (0.70%) | 68.03 |

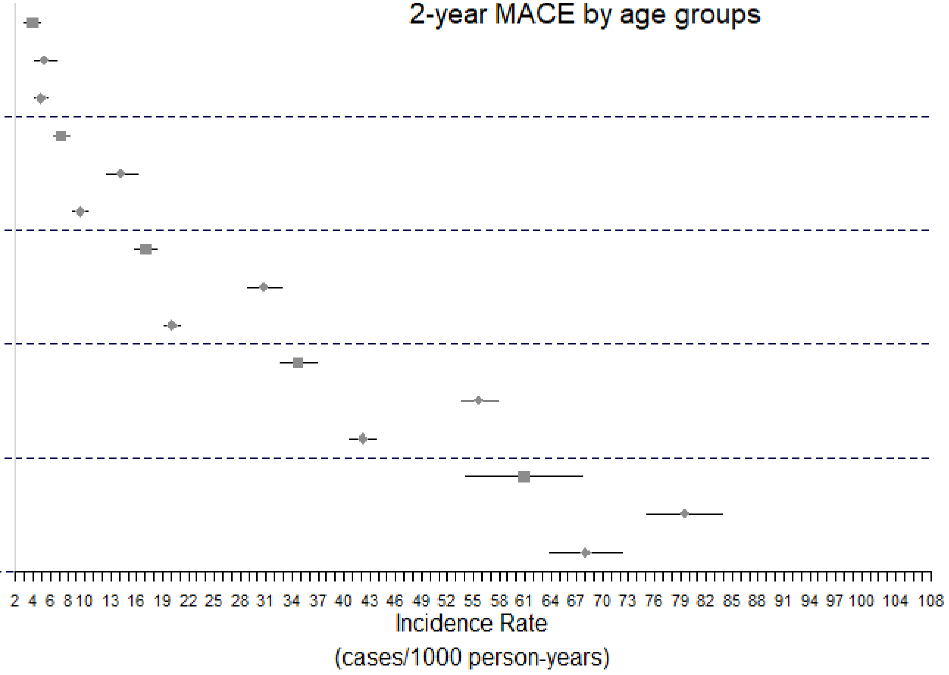

#### c. After one year of follow up (Stroke/MI)

| Age group | Cohort | Number of events (%) | Incidence Rate (cases/1000py) |
|---|---|---|---|
| 50-59 | OST | 4.37 (0.06%) | 4.37 |
| | IFX | 3.94 (0.03%) | 3.94 |
| | OBP | 5.09 (0.06%) | 5.09 |
| 60-69 | OST | 6.60 (0.16%) | 6.6 |
| | IFX | 12.82 (0.15%) | 12.82 |
| | OBP | 8.73 (0.18%) | 8.73 |
| 70-79 | OST | 14.47 (0.46%) | 14.47 |
| | IFX | 26.06 (0.54%) | 26.06 |
| | OBP | 17.84 (0.54%) | 17.84 |
| 80-89 | OST | 29.18 (0.70%) | 29.18 |
| | IFX | 40.36 (1.31%) | 40.36 |
| | OBP | 34.60 (0.91%) | 34.6 |
| 90+ | OST | 46.61 (0.20%) | 46.61 |
| | IFX | 50.33 (0.58%) | 50.33 |
| | OBP | 46.84 (0.27%) | 46.84 |

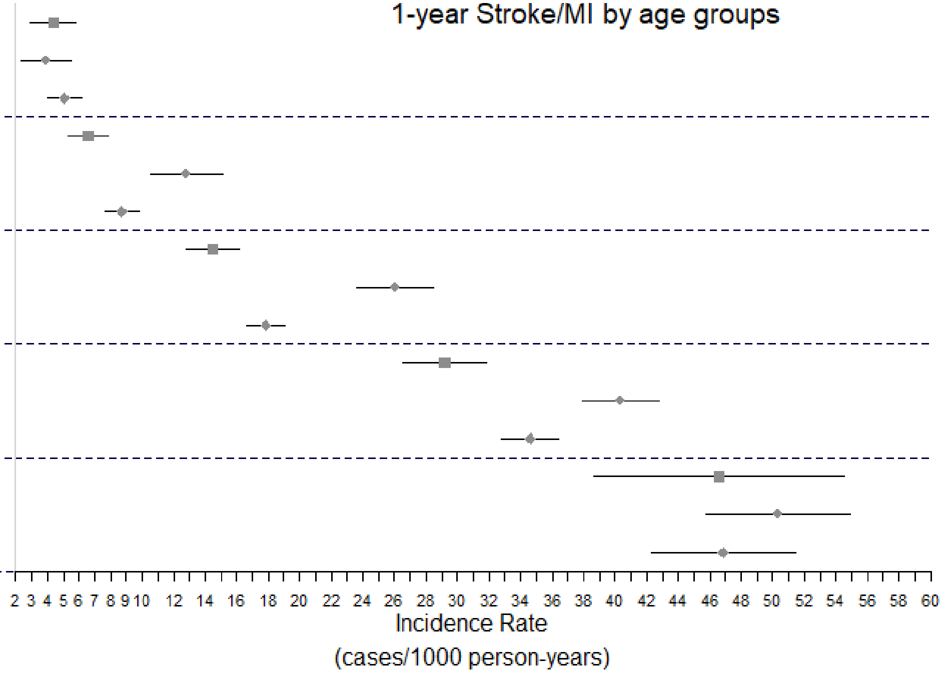

#### d. After two years of follow up (Stroke/MI)

| Age group | Cohort | Number of events (%) | Incidence Rate (cases/1000py) |
|---|---|---|---|
| 50-59 | OST | 3.61 (0.09%) | 3.61 |
| | IFX | 4.42 (0.07%) | 4.42 |
| | OBP | 4.43 (0.10%) | 4.43 |
| 60-69 | OST | 6.63 (0.32%) | 6.63 |
| | IFX | 10.71 (0.24%) | 10.71 |
| | OBP | 8.20 (0.34%) | 8.2 |
| 70-79 | OST | 14.65 (0.90%) | 14.65 |
| | IFX | 22.61 (0.89%) | 22.61 |
| | OBP | 17.25 (1.00%) | 17.25 |
| 80-89 | OST | 28.06 (1.27%) | 28.06 |
| | IFX | 36.17 (2.13%) | 36.17 |
| | OBP | 32.87 (1.61%) | 32.87 |
| 90+ | OST | 41.85 (0.33%) | 41.85 |
| | IFX | 41.97 (0.83%) | 41.97 |
| | OBP | 43.76 (0.45%) | 43.76 |

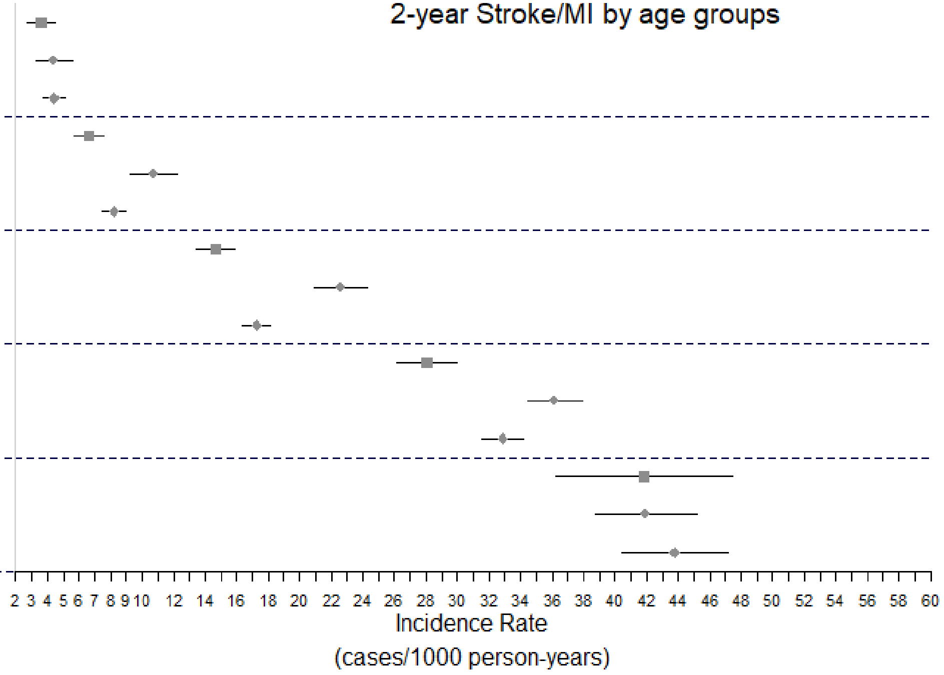

**Figure S2. Incidence rates of two-year MACE, one- and two-year stroke/MI, and MACE and stroke/MI by gender**

a. **After two years of follow up (MACE)**

| Cohort | Number of events (%) | Incidence Rate (cases/1000py) |
|---|---|---|
| OST n=65295 | 2311 (3.54%) | 18.91 |
| IFX n=80587 | 5227 (6.49%) | 41.63 |
| OBP n=145959 | 6470 (4.25%) | 24.54 |

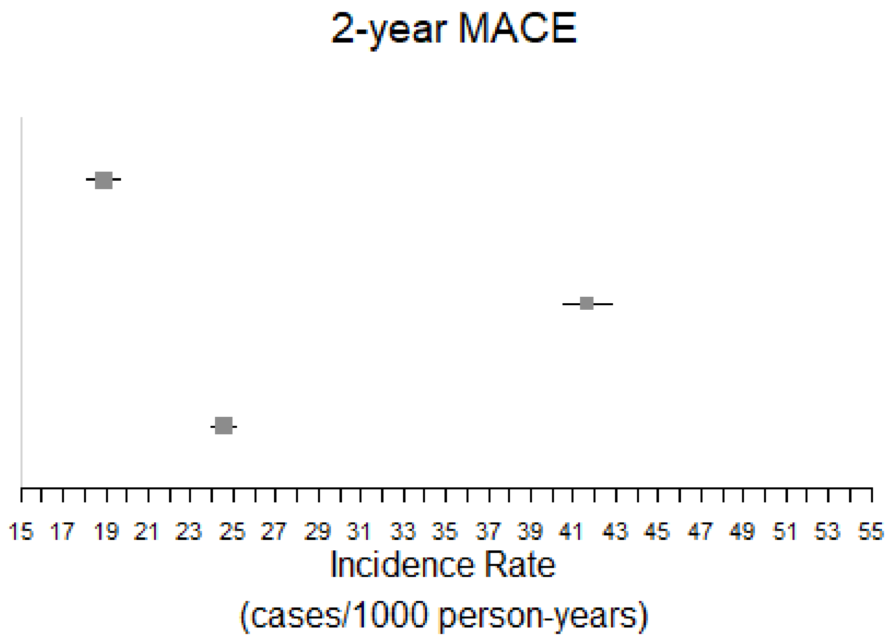

**b.    After one and two years of follow up (Stroke/MI)**

| Cohort | Time | Number of events (%) | Incidence Rate (cases/1000py) |
|---|---|---|---|
| OST | 1-year | 1026 (1.57%) | 16.28 |
| n=65295 | 2-year | 1894 (2.90%) | 15.50 |
| IFX | 1-year | 2099 (2.60%) | 31.08 |
| n=80587 | 2-year | 3355 (4.16%) | 26.74 |
| OBP | 1-year | 2866 (1.88%) | 20.78 |
| n=145959 | 2-year | 5105 (3.35%) | 19.38 |

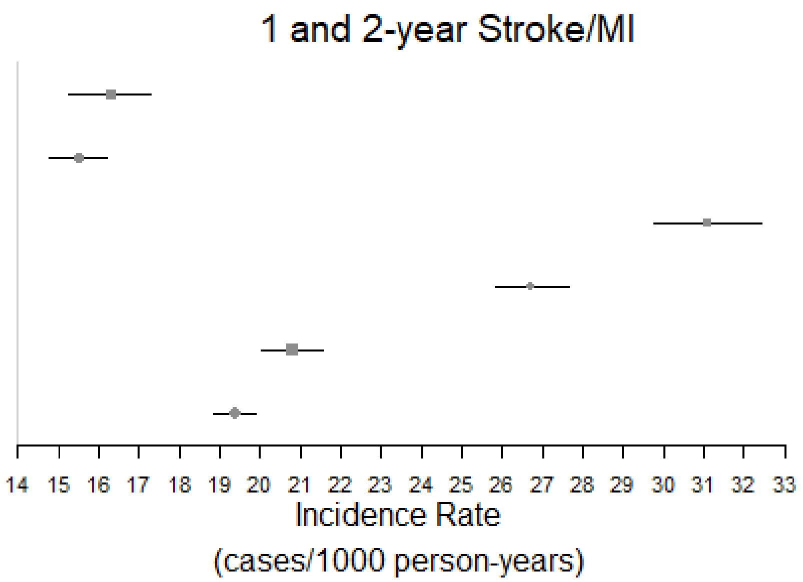

**c. After one and two years of follow up (Females)**

| Outcome | Cohort | Time | Number of events (%) | Incidence Rate (cases/1000py) |
|---|---|---|---|---|
| MACE | OST | 1-year | 987 (1.7%) | 17.95 |
| | n=56679 | 2-year | 1862 (3.3%) | 17.41 |
| | IFX | 1-year | 2499 (4.0%) | 47.29 |
| | n=61793 | 2-year | 3846 (6.2%) | 38.92 |
| | OBP | 1-year | 2659 (2.3%) | 23.96 |
| | n=116412 | 2-year | 4776 (4.1%) | 22.39 |
| Stroke/MI | OST | 1-year | 825 (1.5%) | 15.00 |
| | n=56679 | 2-year | 1550 (2.7%) | 14.50 |
| | IFX | 1-year | 1570 (2.5%) | 29.73 |
| | n=61793 | 2-year | 2536 (4.1%) | 25.69 |
| | OBP | 1-year | 2093 (1.8%) | 18.87 |
| | n=116412 | 2-year | 3760 (3.2%) | 17.64 |

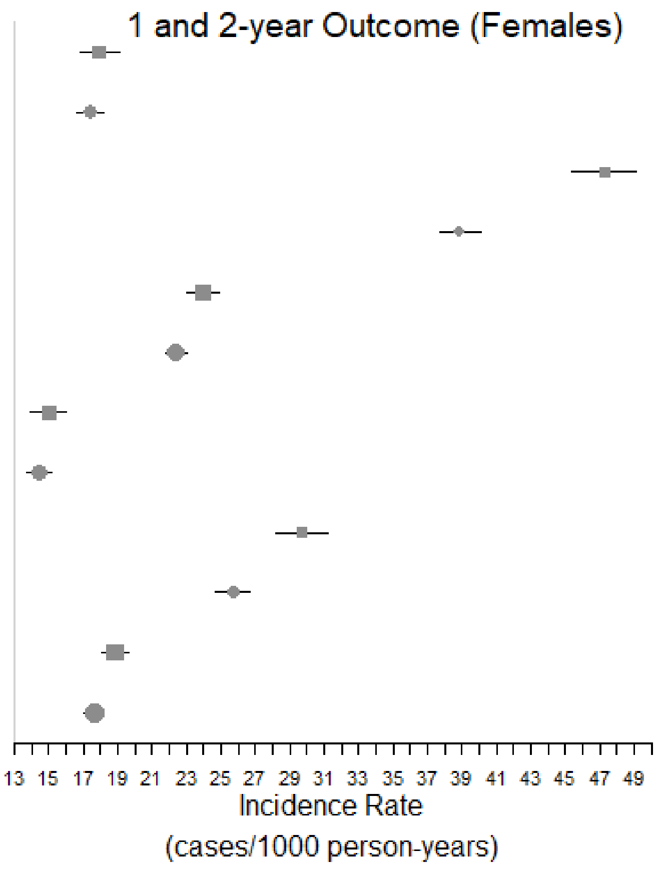

**d. After one and two years of follow up (Males)**

| Outcome | Cohort | Time | Number of events (%) | Incidence Rate (cases/1000py) |
|---|---|---|---|---|
| MACE | OST n=8616 | 1-year | 251 (2.9%) | 31.07 |
| | | 2-year | 449 (5.2%) | 29.40 |
| | IFX n=18794 | 1-year | 954 (5.1%) | 64.74 |
| | | 2-year | 1381 (7.3%) | 51.63 |
| | OBP n=29547 | 1-year | 964 (3.3%) | 35.72 |
| | | 2-year | 1694 (5.7%) | 33.65 |
| Stroke/MI | OST n=8616 | 1-year | 201 (2.3%) | 24.89 |
| | | 2-year | 344 (4.0%) | 22.54 |
| | IFX n=18794 | 1-year | 529 (2.8%) | 35.93 |
| | | 2-year | 819 (4.4%) | 30.65 |
| | OBP n=29547 | 1-year | 773 (2.6%) | 28.66 |
| | | 2-year | 1345 (4.6%) | 26.74 |

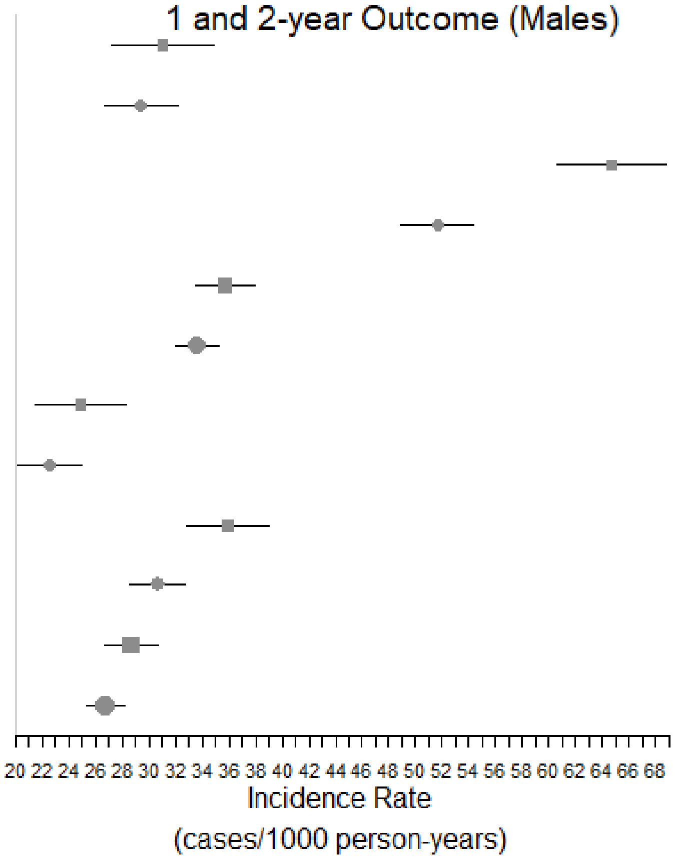

**Figure S3. Area under ROC curve for internal validation**

a. **Two-year MACE**

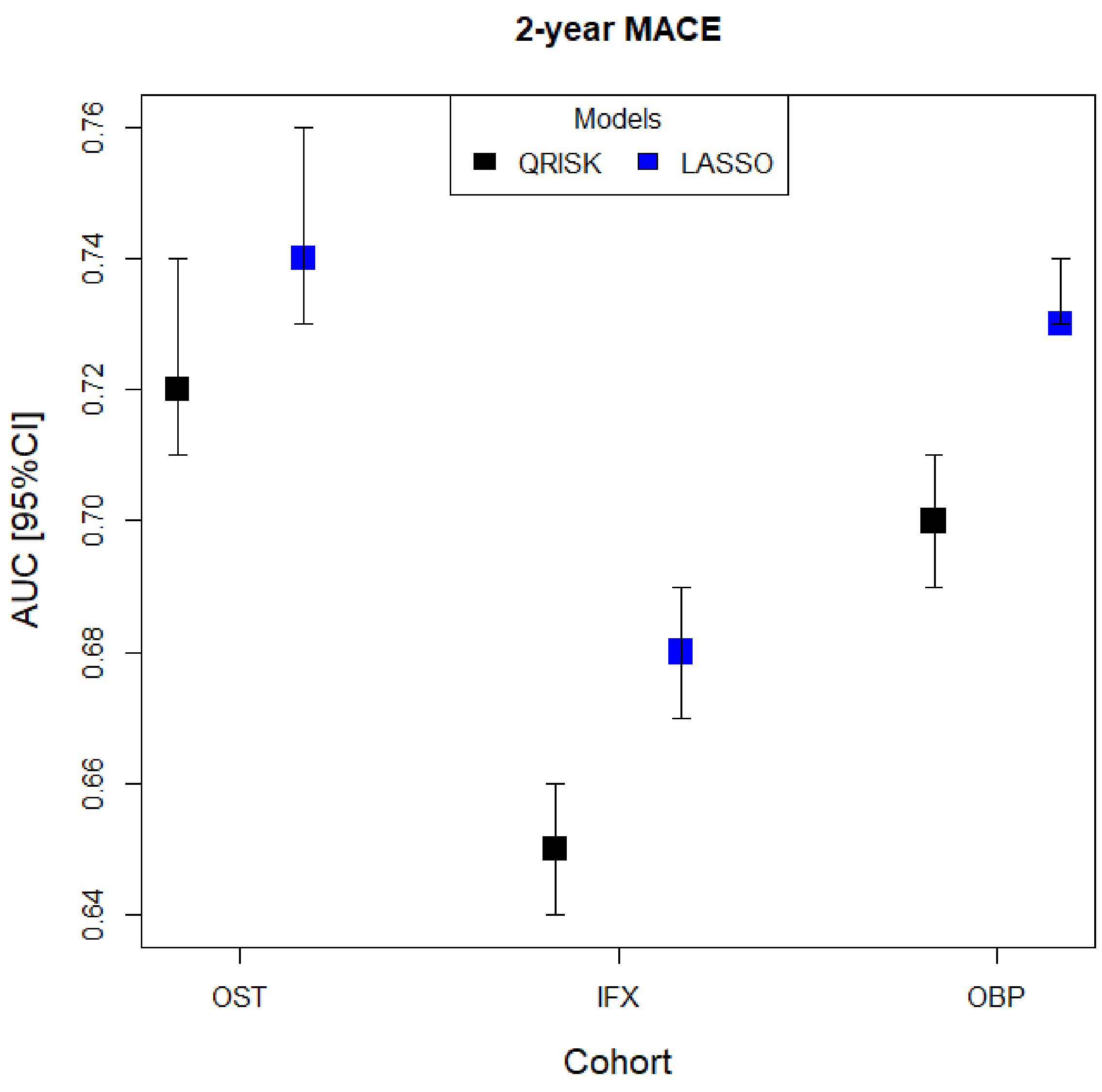

2-year MACE
Models
QRISK
LASSO
AUC [95%CI]
0.64
0.66
0.68
0.70
0.72
0.74
0.76
OST
IFX
OBP
Cohort

#### b. One- and two-year Stroke/MI

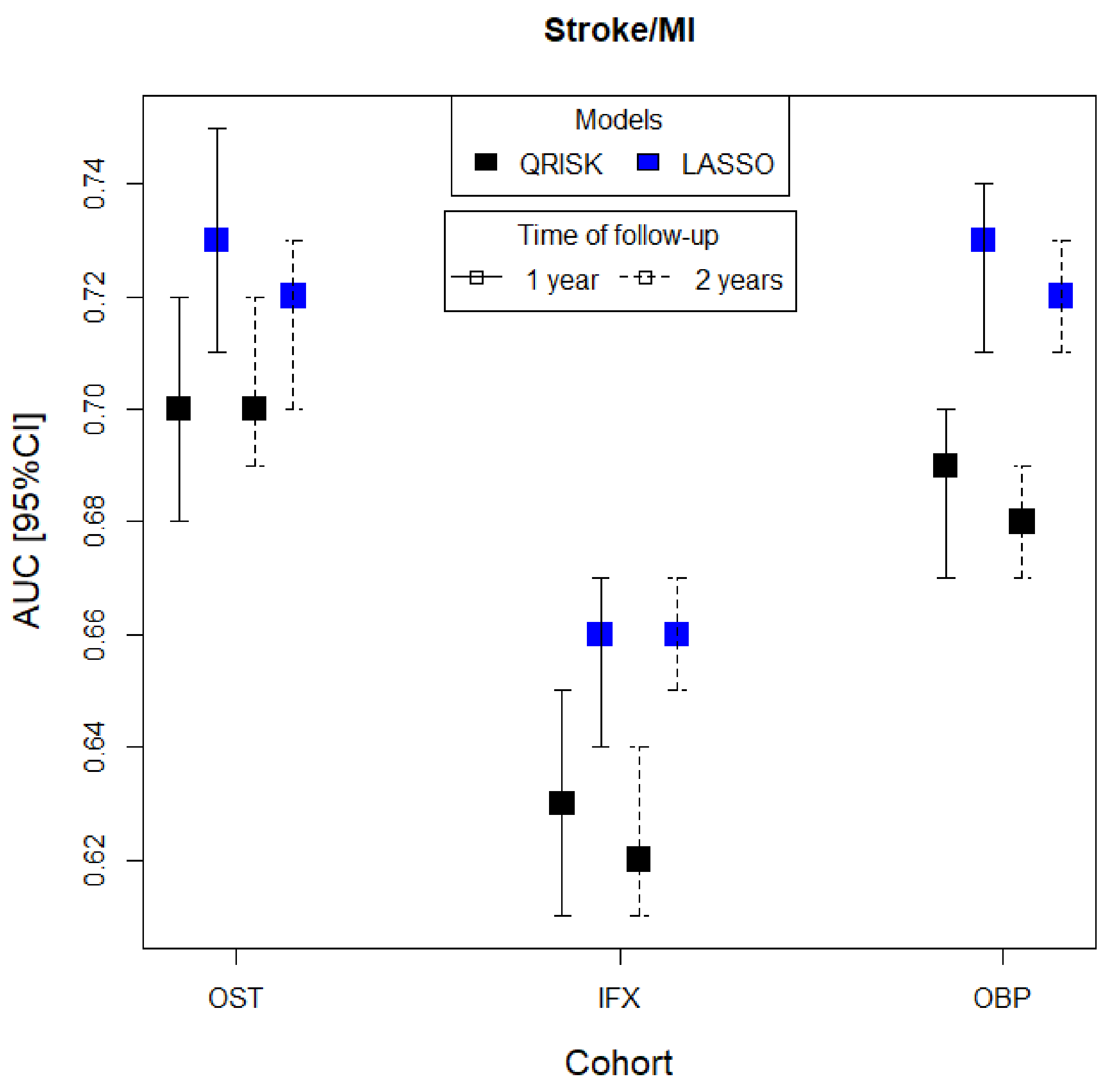
Stroke/MI
Models
QRISK
LASSO
Time of follow-up
1 year
2 years
0.74
0.72
0.70
0.68
0.66
0.64
0.62
AUC [95%CI]
OST
IFX
OBP
Cohort

#### c. One- and two-year MACE (gender-based models)

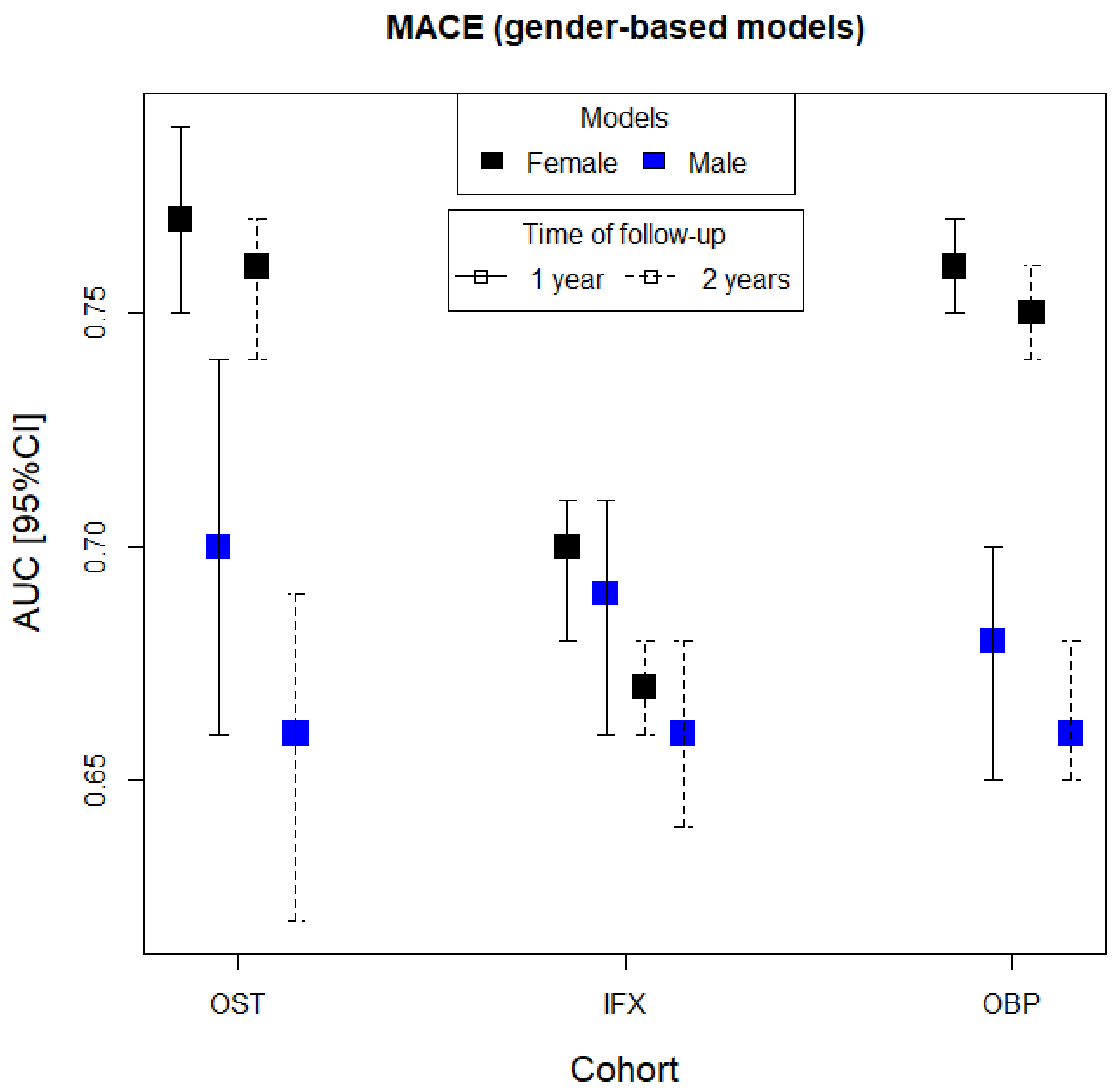

MACE (gender-based models)
Models
Female
Male
Time of follow-up
1 year
2 years
AUC [95%CI]
0.65
0.70
0.75
OST
IFX
OBP
Cohort

#### d. One- and two-year Stroke/MI (gender-based models)

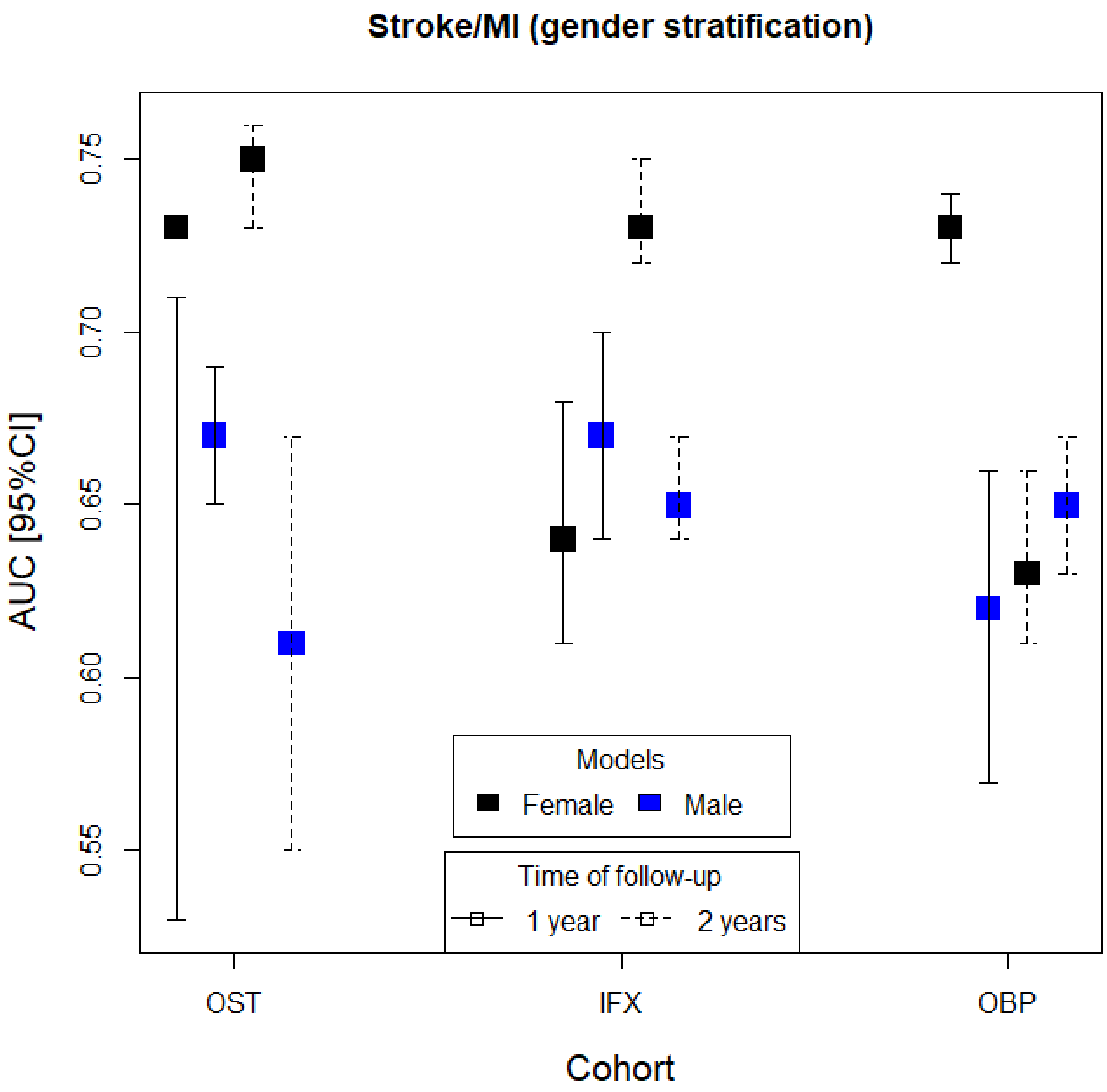

Stroke/MI (gender stratification)
AUC [95%CI]
0.55
0.60
0.65
0.70
0.75
Models
Female
Male
Time of follow-up
1 year
2 years
OST
IFX
OBP
Cohort

**Figure S4. Calibration curves for internal validation of two-year MACE prediction in cohorts OST, IFX and OBP from left to right**

a. **Two-year MACE1 prediction by age deciles**

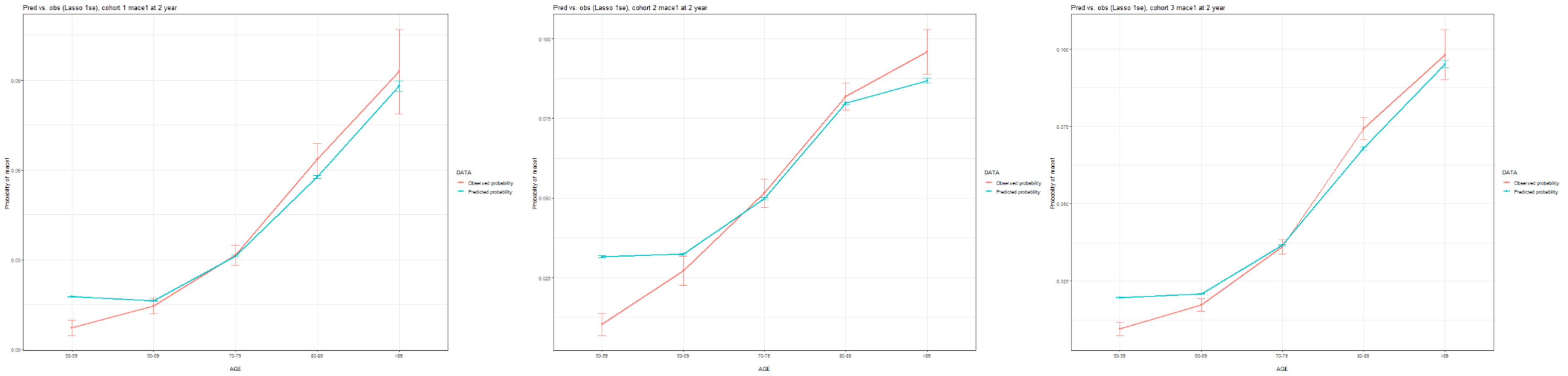

#### b. Two-year MACE1 prediction by age and gender

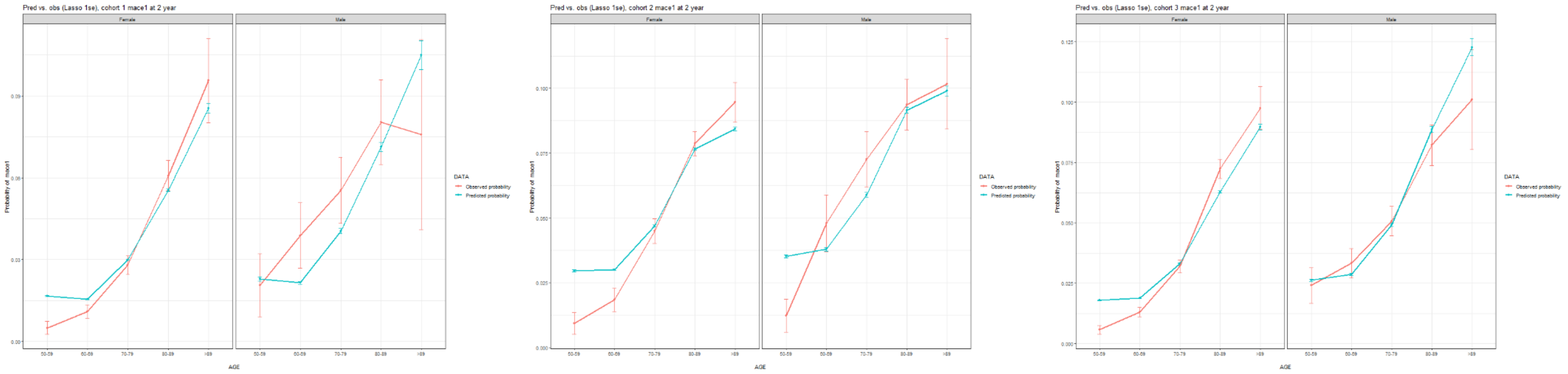


### Figure S5. Calibration curves for internal validation of MI/stroke prediction by age deciles

#### a. One-year MI/stroke prediction cohorts OST, IFX and OBP from left to right

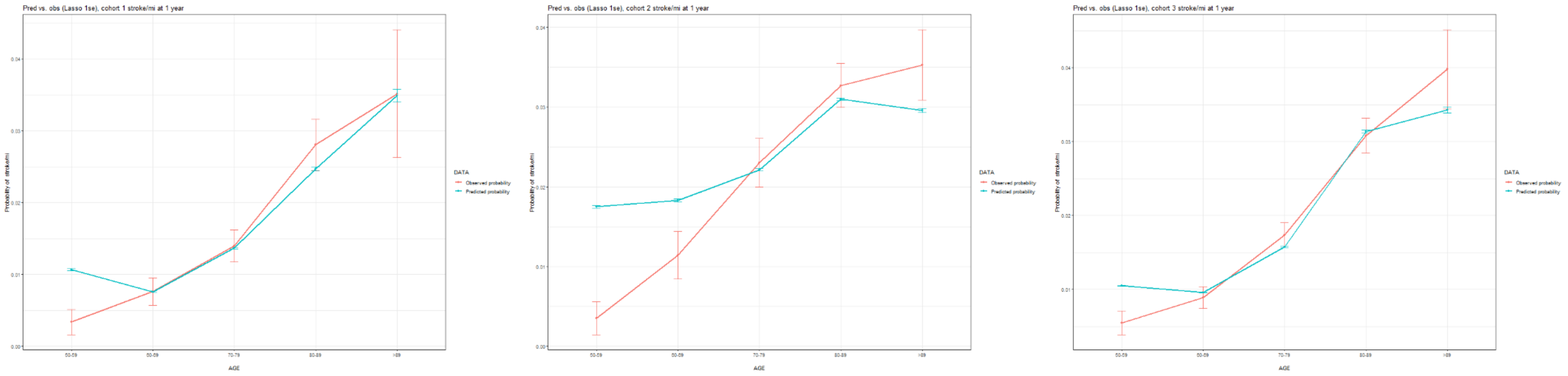

**b. Two-year MI/stroke prediction in cohorts OST, IFX and OBP from left to right**

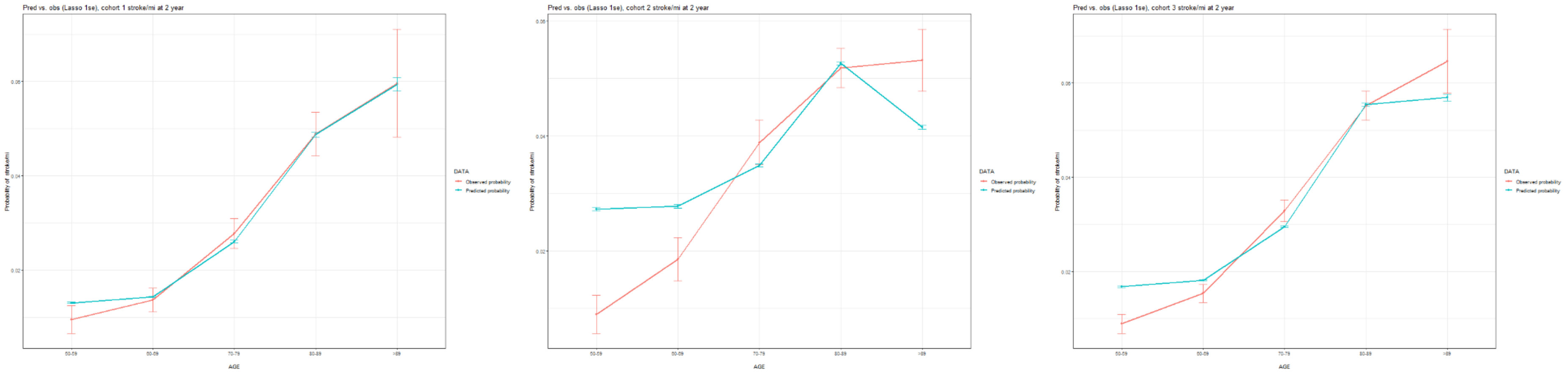


**Figure S6. Calibration curves of MI/stroke prediction stratified by age and gender**

**a. One-year MI/stroke prediction in cohorts OST, IFX and OBP from left to right**

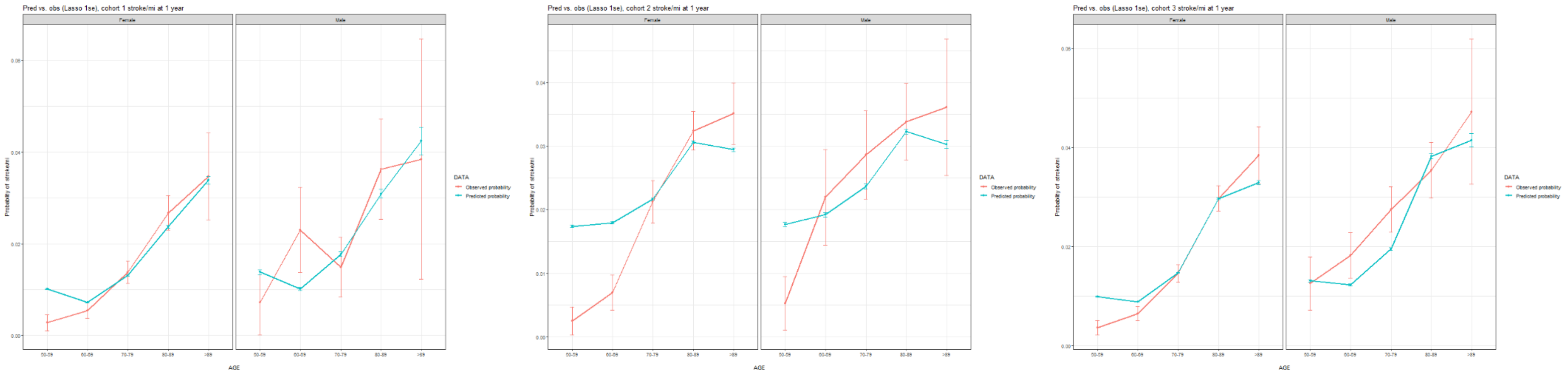

#### b. Two-year MI/stroke prediction in cohorts OST, IFX and OBP from left to right

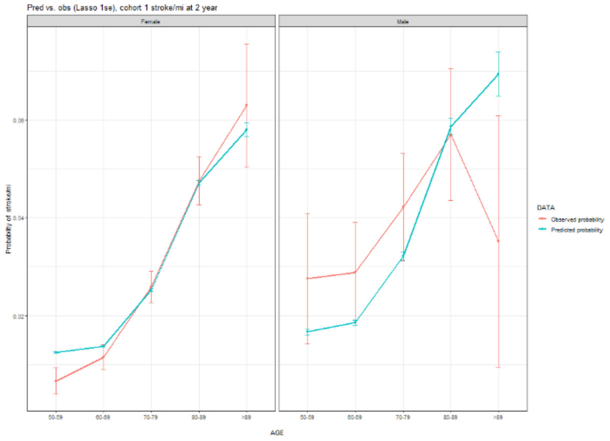


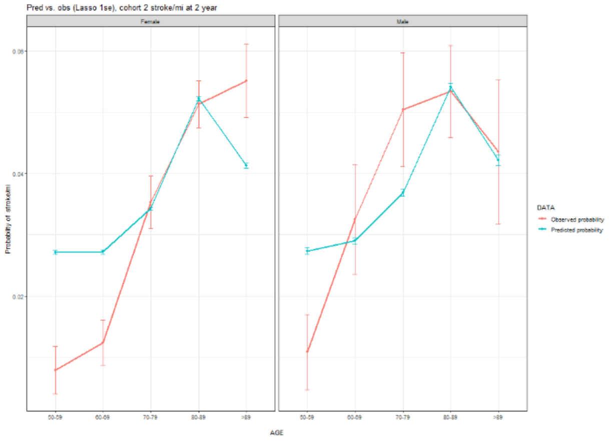


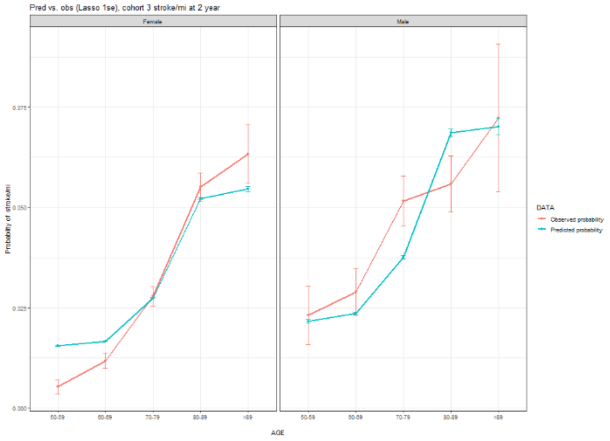
